# Latent Genetic Factors and Causal Pathways Linking Substance Use to Psychiatric and Personality Phenotypes: a Genomic SEM and Mendelian Randomization Study

**DOI:** 10.64898/2026.09.28.26364160

**Authors:** Kaya C. van Grondelle, Adrià Túnez, Joëlle A. Pasman, Merel Wouda, Laura W. Wesseldijk, Bochao D. Lin, Anastasia A. Goula, Karin J. H. Verweij, Jurjen J. Luykx

**Author notes:** Corresponding authors 1. Kaya C. van Grondelle, Meibergdreef 9, 1105AZ, Amsterdam, the Netherlands, 2. Jurjen J. Luykx, Amstelveenseweg 589, 1081 JC, Amsterdam, the Netherlands. Shared supervision (Karin J. H. Verweij, Jurjen J. Luykx).

## Abstract

Substance use and psychiatric disorders frequently co-occur, but establishing causality of their relationships is difficult given the multifactorial nature of these traits. To assess these relationships, we applied two genetically informed methods using the largest available European-ancestry genome-wide association studies of six substance use behaviors and twelve psychiatric and personality phenotypes. Using Genomic Structural Equation Modelling, we modelled the latent genetic architecture of the psychiatric and personality traits and examined these factors’ correlations with substance use traits. A structure with three factors fit the data best: F1 (PTSD, MDD, GAD, ADHD, agreeableness), F2 (BD, SCZ, OCD, openness), and F3 (OCD, GAD, extraversion, neuroticism), which showed differential correlations with substance use. Smoking correlated significantly which each factor, however, strongest with F1, alcohol use showed weak but significant correlations with F1 and F2, and cannabis use showed moderate correlations with F1 and F2, but strongest with F2. Using Mendelian Randomization, we examined bidirectional causal relationships. We found strong evidence that smoking continuation was causally associated with increased bipolar disorder risk, cigarettes per day with increased schizophrenia, and smoking initiation with increased extraversion and decreased agreeableness. Genetic liability to schizophrenia and PTSD were causally associated with increased risk of cannabis use, and schizophrenia with increased alcohol use. Liability to neuroticism, openness and agreeableness was causally linked to smoking behaviors, and extraversion to cannabis use. Most remaining associations showed weak causal evidence, largely due to heterogeneity and pleiotropy. Together, these findings provide evidence for both shared genetic liability and, for a subset of relationships, direct causal effects between substance use behaviors, psychiatric disorders, and personality traits, which can help inform future intervention and prevention research.

## INTRODUCTION

Use of harmful substances like tobacco, cannabis and alcohol is highly prevalent in the general population, with an estimated one in four adults smoking and one in three consuming alcohol globally [1, 2]. Individuals with mental health problems are even more likely to use these substances, showing two to three times higher dependency rates [3–6]. However, the nature and direction of the relationship between substance use and psychiatric symptomatology remains poorly understood [7]. Personality traits show substantial overlap with both psychiatric disorders and substance use behaviors [8–10], and examining these alongside psychiatric outcomes may therefore contribute to a more comprehensive understanding of the etiology underlying their complex relationship with substance use behaviors.

Several theoretical frameworks have been proposed to explain the high co-occurrence of substance use with psychiatric and personality traits, broadly falling into two categories: unidirectional causal models and shared liability models [7, 11]. Unidirectional models propose causal pathways in either direction: the self-medication and vulnerability hypotheses propose that individuals with mental health problems or certain personality traits, such as high neuroticism, use substances more frequently to alleviate distressing symptoms or unpleasant emotions, thereby functioning as a coping mechanism [12–14], while the causal risk factor and scar models propose the reverse, where substance use can increase the likelihood of developing a psychiatric disorder or lead to lasting personality changes by inducing neurobiological changes [11, 15–17]. In contrast, the second framework, including common liability and common cause models, instead proposes that substance use and personality/psychiatric traits arise from shared underlying biological, psychological or environmental risk factors [18, 19]. Moreover, observational evidence consistently shows associations between personality and psychiatric traits. High neuroticism, and low agreeableness and conscientiousness are linked to both internalizing and externalizing psychopathology, while higher extraversion and openness are associated with greater risk-taking in general, including substance use [9, 17, 20]. These high rates of comorbidity suggest that psychiatric and personality traits may cluster into broader groups sharing similar underlying relationships with substance use [21]. Considering personality alongside psychiatric traits therefore allows us to test whether substance use relates to such broader, overlapping clusters of traits, rather than to individual diagnostic categories in isolation.

In psychiatry, proving causation is a challenging task. Randomized controlled trials (RCTs) can be costly, time-consuming, and ethically questionable, whereas observational studies are easier to perform but prone to confounding and reverse causation [22, 23]. Genome-wide association studies (GWAS) and post-GWAS methods such as Genomic Structural Equation Modelling (gSEM) and Mendelian Randomization (MR) offer powerful tools to dissect the nature of genetic overlap between traits [24]. gSEM uses GWAS summary statistics to model shared genetic architecture across traits and identify latent genetic factors underlying their correlations, without requiring individual-level data [25]. MR uses genetic variants robustly associated with an exposure as instrumental variables to estimate its potential causal effect on an outcome [26]. In a two-sample design, exposure and outcome associations are drawn from two independent GWAS samples, allowing the use of large, publicly available GWAS summary statistics [27]. Because genetic variants, primarily single-nucleotide polymorphisms (SNPs), are randomly assigned at conception and precede symptom onset, they are unaffected by environmental confounders or by the outcome itself. This can be used to address reverse causality, where it is often unclear whether a risk factor precedes or follows the onset of a disorder [28]. Moreover, MR addresses pleiotropy, where a single genetic variant influences multiple traits [25, 29]. Vertical pleiotropy, where one SNP influences one trait which in turn influences another, is consistent with unidirectional causal models and is the form of pleiotropy MR is designed to accommodate when estimating causal effects [30, 31]. In contrast, horizontal pleiotropy occurs when a single genetic variant independently affects two or more distinct traits or pathways, pointing towards shared underlying biological processes, and can be assessed using sensitivity analyses [32, 33]. Ruling out horizontal pleiotropic effects allows MR estimates to be interpreted as causal in a way similar to RCTs.

Previous MR studies investigating associations between substance use and psychiatric traits support a self-medication model by demonstrating that genetic predispositions to psychiatric disorders are causally associated with increased substance use, most consistently internalizing disorders to alcohol and cannabis use [34–36]. Moreover, MR studies mostly provide support for personality traits having a causal effect on substance use behaviors, and weaker support for the alternative direction [24, 36–38]. In contrast, other MR evidence is consistent with causal risk factor models, by causally linking genetic liability for substance use to increased psychiatric risk, with smoking and cannabis often showing the strongest evidence for causal effects [35, 36, 39–41]. Further studies have indicated that overlapping risk factors, including shared genetic influences, jointly increase risk for both substance use and psychopathology, thus providing support for common liability models [42, 43]. Previous gSEM studies have distinguished broad internalizing and externalizing dimensions, with substance-use traits loading particularly strongly on an externalizing factor, while also revealing genetic overlap across these dimensions [19].

Overall, recent MR studies examining substance use and psychiatric disorders yielded inconsistent findings across studies, and many used methodologically limited designs, and many have relied on a limited set of sensitivity analyses to assess pleiotropy, despite recommendations that IVW estimates be triangulated against multiple robust methods (e.g., MR-Egger, median- and mode-based estimators, MR-PRESSO) to strengthen causal inference [24, 28, 44]. Existing studies have largely examined individual substance use and psychiatric phenotypes in isolation, few have systematically modelled shared genetic architecture across broader trait domains, and many rely on older or smaller GWASs that limit statistical power. Causal relationships between substance use and personality traits remain understudied, likely due to the limited power of earlier personality GWASs, and to our knowledge these traits have not yet been examined alongside psychiatric disorders using a comparable bidirectional MR approach, despite their well-established overlap. Recently, a new GWAS of the Big Five personality traits was published by the revived Genomics of Personality Consortium (ReGPC), combining data across 46 cohorts which resulted in 611,037 to 1.14 million participants per trait [45]. As GWAS sample sizes continue to grow, there is now an opportunity to revisit these relationships with more powerful instruments and a broader, more systematic scope. gSEM studies are less common, although they have been applied to psychiatric traits and their associations with substance behaviors before [7, 25, 38, 46].

The present study therefore aims to (1) model the shared genetic architecture across the Big Five personality traits (openness, conscientiousness, extraversion, agreeableness and neuroticism) and seven psychiatric disorder phenotypes (Attention-Deficit Hyperactivity Disorder, Post-Traumatic Stress Disorder, Major Depressive Disorder, Schizophrenia, Obsessive-Compulsive Disorder, Bipolar Disorder, General Anxiety Disorder), identify latent genetic factors underlying their genetic associations, and examine how these correlate genetically with substance use (2) examine bidirectional causal relationships of substance use behaviors – including alcohol use, smoking, and cannabis use – with a broad range of psychiatric and personality phenotypes, using the most recent GWASs of European ancestry individuals. This will then allow us to construct a comparative overview of causal effects, highlighting which exposures show the strongest evidence for affecting specific disorder domains and trait clusters, and vice versa.

## METHODS

### Data sources

This study used publicly available summary-level data from large-scale GWAS. Where multiple GWAS were available for a given phenotype, the largest dataset was selected to maximize statistical power and instrument strength. All analyses were restricted to individuals of European ancestry, as both MR and gSEM rely on linkage disequilibrium (LD) patterns that differ substantially across ancestries, requiring ancestry-matched reference panels and genetically homogeneous input data to produce valid estimates. Combining GWAS across ancestries without appropriate methods would therefore risk biased or uninterpretable results. Furthermore, for most of the phenotypes included in this study, GWAS in non-European populations are not yet sufficiently powered or are not available yet. Details of all GWAS summary statistics used are reported in Supplementary Table S1. Traits were included only if their SNP-based heritability was at least 3% and had a Z-score of at least 10. Sample overlap between exposure and outcome GWAS was minimized by drawing on non-overlapping samples where possible, see Supplementary Methods (S2) for full details.

#### Substance use behaviors

Six substance use phenotypes were included, derived from two large-scale GWAS meta-analyses. Smoking and alcohol phenotypes were obtained from the GWAS & Sequencing Consortium of Alcohol and Nicotine use (GSCAN) [47]. Four smoking phenotypes representing distinct stages of tobacco use were included: Smoking Initiation and Smoking Cessation, indicating whether an individual ever smoked at least 100 cigarettes or had successfully quit, respectively, and Age of Smoking Initiation and Cigarettes per Day, reflecting onset age and consumption level among current and former smokers, respectively. Alcohol use was operationalized as Drinks per Week. Cannabis Ever Use was derived from the International Cannabis Consortium [48]. Smoking Initiation, Smoking Cessation, and Cannabis Ever Use were operationalized as binary case-control phenotypes, all remaining substance use phenotypes were continuous.

#### Psychiatric and personality phenotypes

Seven psychiatric disorders were assessed: Major Depressive Disorder (MDD) [49], Schizophrenia (SCZ) [50], Bipolar Disorder (BD) [51], Post-Traumatic Stress Disorder (PTSD) [52], Generalized Anxiety Disorder (GAD) [53], Obsessive-Compulsive Disorder (OCD) [54], and Attention-Deficit/Hyperactivity Disorder (ADHD) [55]. Summary statistics were obtained from the Psychiatric Genomics Consortium (PGC), except for ADHD, which were obtained from the meta-analyses performed by van der Laan et al. (2025) [55]. The five OCEAN personality traits (Openness, Conscientiousness, Extraversion, Agreeableness, and Neuroticism) were included, with summary statistics obtained from a meta-analysis performed by the Revived Genomics of Personality Consortium [56]. BD, OCD, and MDD were case-control phenotypes, all remaining phenotypes were continuous measures. Further details on individual phenotype definitions are provided in the Supplementary Methods (S3).

### Statistical analyses

#### Genetic architecture of substance use, personality traits and psychiatric disorders

We conducted 2 sets of analyses: (1) Latent factor architecture mapping, to capture genetic signal overlapping between psychiatric disorders and personality traits, and (2) Genetic correlations between the latent factors with substance use traits, to compare how substance use associates with different domains of psychiatric disorders and personality traits. All summary statistics were pre-processed using the tidyGWAS R package to scan for missing values, duplications, indels, and to match the RSID of the summary statistics with a dbSNP reference file.

First, we used LDSC [57] to assess SNP-based heritability and genetic correlations between all traits included in the model. Genetic correlations were plotted as a heatmap using the corrplot package in R. Next, we used gSEM [25] to model the joint genetic architecture of the psychiatric disorders and personality traits, in order to identify latent genetic factors across the different personality and psychiatric traits. We first conducted exploratory factor analyses (EFA) to identify the underlying factor structure. We used Promax rotation and established a minimum cutoff of 0.3 for factor loadings in the EFA model. We prompted gSEM to identify the structure with 1, 2, 3, 4, and 5 latent factors separately (Supplementary Figure 1). Then, we performed confirmatory factor analyses (CFA) to formally test and estimate these structures. Latent factors were allowed to correlate with each other, cross-loadings were also allowed, and all models were estimated using diagonally weighted least squares (WLS). Model fit was evaluated using the Comparative Fit Index (CFI) and Standardized Root Mean Square Residual (SRMR), with commonly used thresholds of CFI >0.90 and SRMR <0.10 taken as reference. The best fitting model was chosen to feed forward to the second set of analyses.

To characterize the association between the identified latent personality traits/psychiatric disorders factors with substance use, we then estimated the genetic correlations between the latent factors from the best fitting model and traits related to the use of the three different substances.. Genetic correlations were calculated within gSEM by modelling each substance use trait as a latent factor and allowing this factor to correlate to each latent factor from the model. The substance use traits are first loaded to latent factors to avoid scaling problems derived from calculating correlations between indicators in different hierarchical layers. The four tobacco use traits were loaded onto the same smoking latent factor.

#### Causal relationships between substance use, and personality and psychiatric phenotypes

The overall analytic workflow, from exposure/outcome selection through primary and sensitivity analyses, is summarized in Supplementary Figure 2. MR analysis relies on three core assumptions, relevance, independence, and exclusion restriction (Supplementary Figure 3), which were addressed and evaluated throughout the analytical pipeline described below. ADHD was excluded as an outcome in the forward analyses, as its typical onset in early childhood precedes the initiation of substance use behaviors, violating the temporal precedence.

##### Instrument selection

The process for instrumental variable (IV) selection from the summary statistics was performed following common practice. First, genome-wide significant SNPs (P < 5 x 10^-8^) from each exposure trait were identified. Where fewer than 10 SNPs were identified at this threshold, the significance criterion was relaxed to p < 5 × 10 ^7^. To account for linkage disequilibrium (LD) and ensure independence of the final IVs used in the assessment, significant SNPs were clumped using the *clump_data* function in the TwoSampleMR package in R (10,000 kb window, r^2^ threshold = 0.001). The 1000 Genomes European sample data was used as the reference panel.

##### Data harmonization and quality control

Exposure and outcome data were harmonized to ensure consistency of effect allele coding and directionality across datasets, retaining only SNPs present in both the exposure and outcome GWAS. Palindromic SNPs with an effect allele frequency exceeding 0.42 were removed to prevent misalignment of ambiguous variants. Instrument strength was evaluated using the F-statistic, calculated per SNP as β²/SE²; SNPs with F < 10 were excluded to minimize weak instrument bias. Instrument strength was summarized per exposure-outcome pair using mean and median F-statistics, alongside R², indicating the proportion of variance in the exposure explained by the instruments.

To investigate the causality of the relationships between substance use and psychiatric disorders and personality, bidirectional two-sample MR analyses were conducted. The random-effects inverse variance weighted (IVW) method was used as the primary estimator, as it provides precise causal estimates under the assumption that pleiotropy is balanced across instruments [58]. However, IVW estimates may be biased in the presence of horizontal pleiotropy, and thus four additional MR methods were applied to assess robustness: the weighted median (WM) estimator, the weighted mode (WMo) estimator, the simple mode estimator, and MR-Egger regression slope (see Supplementary Methods (S4) for a full description of each method and their assumptions). To account for multiple testing across primary IVW analyses, a false discovery rate (FDR) correction was applied separately within each direction of analysis using the Benjamini-Hochberg procedure, with a significance threshold of p_FDR_ < 0.05. As sensitivity analyses serve to assess directional consistency of significant IVW findings rather than independent hypothesis tests, no multiple testing correction was applied to the results of the complementary effect estimators. All results are reported as beta coefficients with standard errors. For binary outcomes, effect estimates are additionally expressed as odds ratios (OR = exp(β)) with 95% confidence intervals for interpretability.

The robustness of the IVW estimates was further evaluated using Steiger filtering to verify assumed causal direction, Cochran’s Q to assess heterogeneity, the MR-Egger intercept test to detect directional horizontal pleiotropy, and MR-PRESSO to test for instruments with outlier effects (see Supplementary Methods S4 – S5 for full descriptions). To systematically classify the robustness of each association, evidence was categorized as strong, weak, or absent. Strong evidence required a statistically significant IVW estimate after FDR correction, directional consistency across all complementary estimators (weighted median, weighted mode, simple mode and MR-Egger), a Steiger-filtered instrument set consistent with the IVW’s causal direction, and no evidence of directional horizontal pleiotropy (MR-Egger intercept and MR-PRESSO). Weak evidence was assigned when the IVW estimate was FDR-significant but showed some inconsistency across complementary estimators, sensitivity tests, or evidence of pleiotropy. Associations for which the IVW estimate was not FDR-significant were classified as showing no evidence and were not evaluated further. Convergence of results across the primary IVW analysis, complementary effect estimators, and sensitivity analyses was the main criterion for evidence for causality.

## RESULTS

### Genetic architecture of substance use, personality traits and psychiatric disorders

To assess the genetic similarity of the traits, we calculated SNP-based heritability estimates and genetic correlations between all eighteen traits (Figure 1). As expected, correlations among personality traits were low, while correlations among the psychiatric outcomes were in the expected direction and ranged widely (absolute correlations ranging from 0.18 to 0.90; all p_FDR_<.05). Correlations between personality traits and psychiatric disorders were generally low, except for neuroticism showing a high genetic correlation with PTSD (rg=0.68), GAD (0.85), and MDD (0.70). Agreeableness showed negative genetic correlations (ranging from rg= -0.10 with OCD to rg= -0.40 with PTSD) with all psychiatric disorders. Substance use traits were more heterogeneous: correlations were generally high between different traits indexing the same substance and more modest to high across different substances (ranging from rg= 0.08 between drinks per week and age of smoking initiation, to rg=0.55 between cannabis use and smoking initiation. Correlations of substance use traits with both personality and psychiatric disorders traits were also heterogeneous, with openness showing the highest correlation with ever use of cannabis (rg=0.46), and ADHD the highest with smoking initiation (rg= 0.58).

**FIGURE 1.**
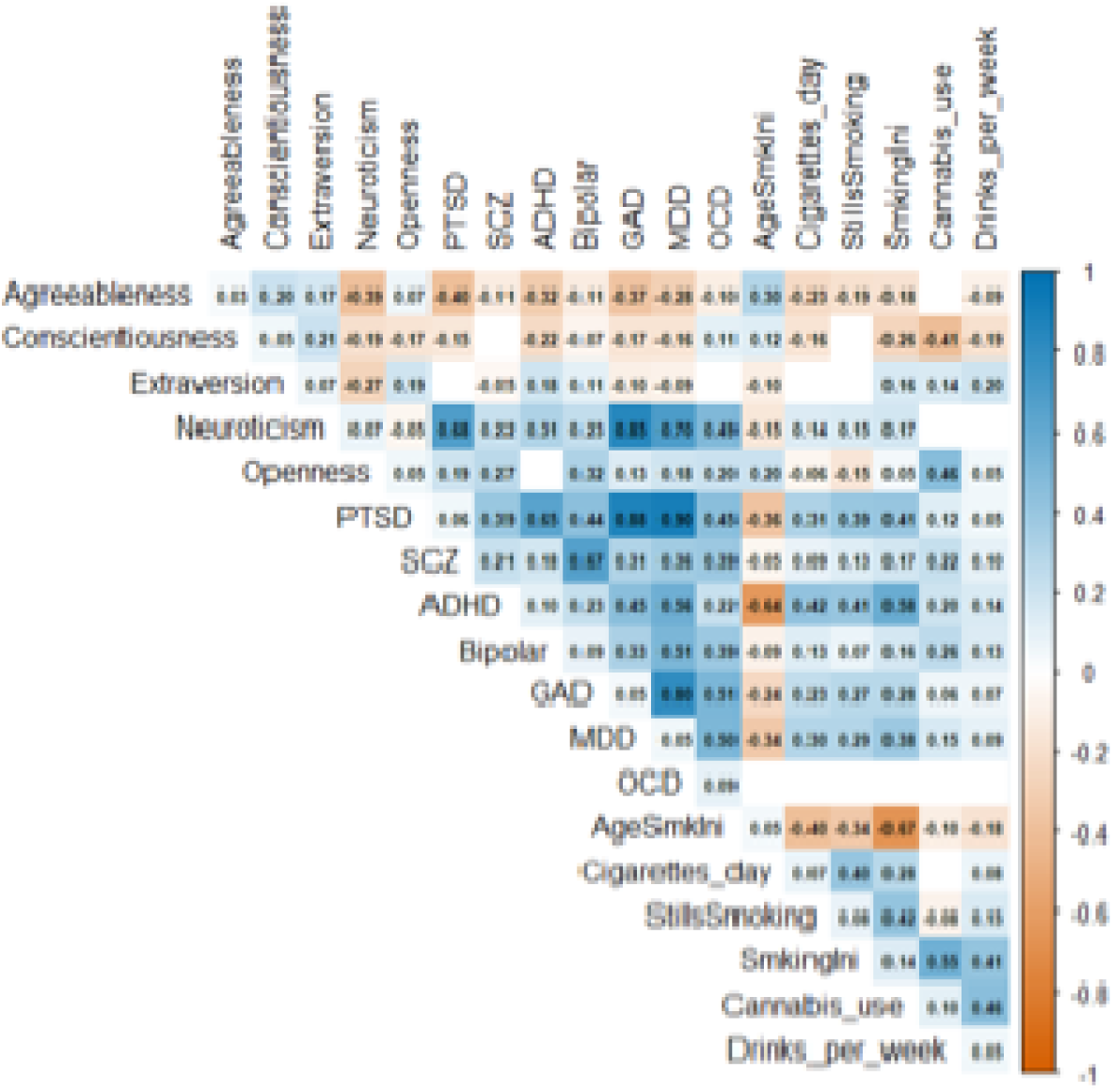
Genetic correlations between all substance use/psychiatric and personality traits. Positive correlations are depicted in blue, negative in orange. Darker colours indicate stronger correlations. Diagonal cells show SNP−based heritability (h^2^) on the liability scale. Blank off−diagonal cells indicate genetic correlations not significant after FDR multiple testing correction (p_FDR_ < .05). Acronyms for trait names are detailed in the summary statistics table (Table S1). <u>Abbreviations</u>: PTSD = Post-Traumatic Stress Disorder, MDD = Major Depressive Disorder, SCZ = schizophrenia, Bipolar = Bipolar Disorder, GAD = General Anxiety Disorder, OCD = Obsessive-Compulsive Disorder, SmkingIni = smoking initiation, StillsSmoking = smoking cessation, AgeSmkIni = age of smoking initiation

Our next objective was to identify the latent factor model that best captured the relationships among the personality/psychiatric traits by grouping traits with similar genetic architectures. The model with three latent factors performed best with statistics showing excellent fit (CFI=0.96, SRMR=0.065). Nearly all traits loaded significantly (p<0.05) onto at least one of the latent factors, and GAD and OCD loaded on multiple latent factors. Conscientiousness did not load onto any of the three factors with the 0.3 cutoff that was used in EFA and was therefore excluded from the model. Factor loadings are presented in Figure 2 with full results in Supplementary Table T17. Although the three latent factors were correlated, they differed in their composition. F1 predominantly captured internalizing psychopathology, with loadings from PTSD, MDD, and GAD, as well as from ADHD and agreeableness. F2 captured the remaining psychiatric traits - BD, SCZ, and OCD - together with openness. F3 was defined by loadings from extraversion, neuroticism, OCD, and GAD. Due to these loadings not corresponding to traditional psychiatric/ personality grouping, we decided to refer to the factors as F1, F2 and F3.

**FIGURE 2.**
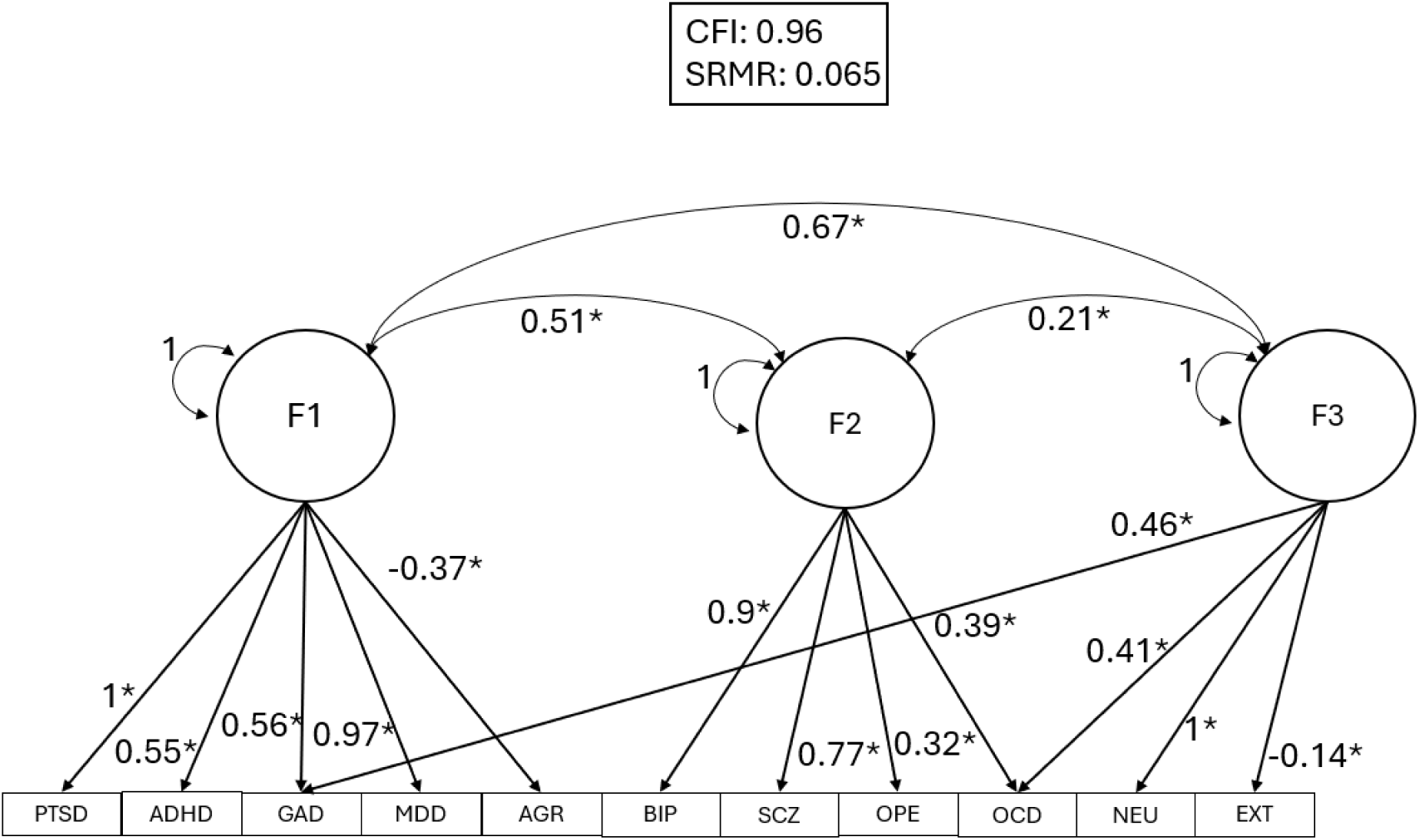
gSEM model results. Circles represent latent factors, and rectangles are “observed” variables (Summary-level GWAS data). Asterisks represent significance at nominal level p<0.05. Statistical fit is shown in the box above the model. <u>Abbreviations</u>: PTSD = Post-Traumatic Stress Disorder, MDD = Major Depressive Disorder, SCZ = schizophrenia, BIP = Bipolar Disorder, GAD = General Anxiety Disorder, OCD = Obsessive-Compulsive Disorder, Ope = openness, Ext = extraversion, Agr = agreeableness, Neu = neuroticism

### Genetic correlations of F1, F2, and F3 with substance use

To better understand the association between personality, psychiatric disorders, and substance use, we evaluated the genetic association of the three latent factors with the three substance use domains (Figure 3). The tobacco smoking factor showed significant (p_FDR_<.05) and positive genetic correlations with all three latent factors. The correlation with F1 (rg=0.68) was higher than with F2 (rg=0.17) and F3 (rg=0.19). Alcohol use showed small but significant positive genetic correlations with F1 and F2 (rg = 0.10 and rg=0.14, respectively), whereas its genetic correlation with F3 was not significant. Cannabis use showed a moderate positive genetic correlation with F2 (rg = 0.34), a small positive genetic correlation with F1, and no significant genetic correlation with F3.

**FIGURE 3.**
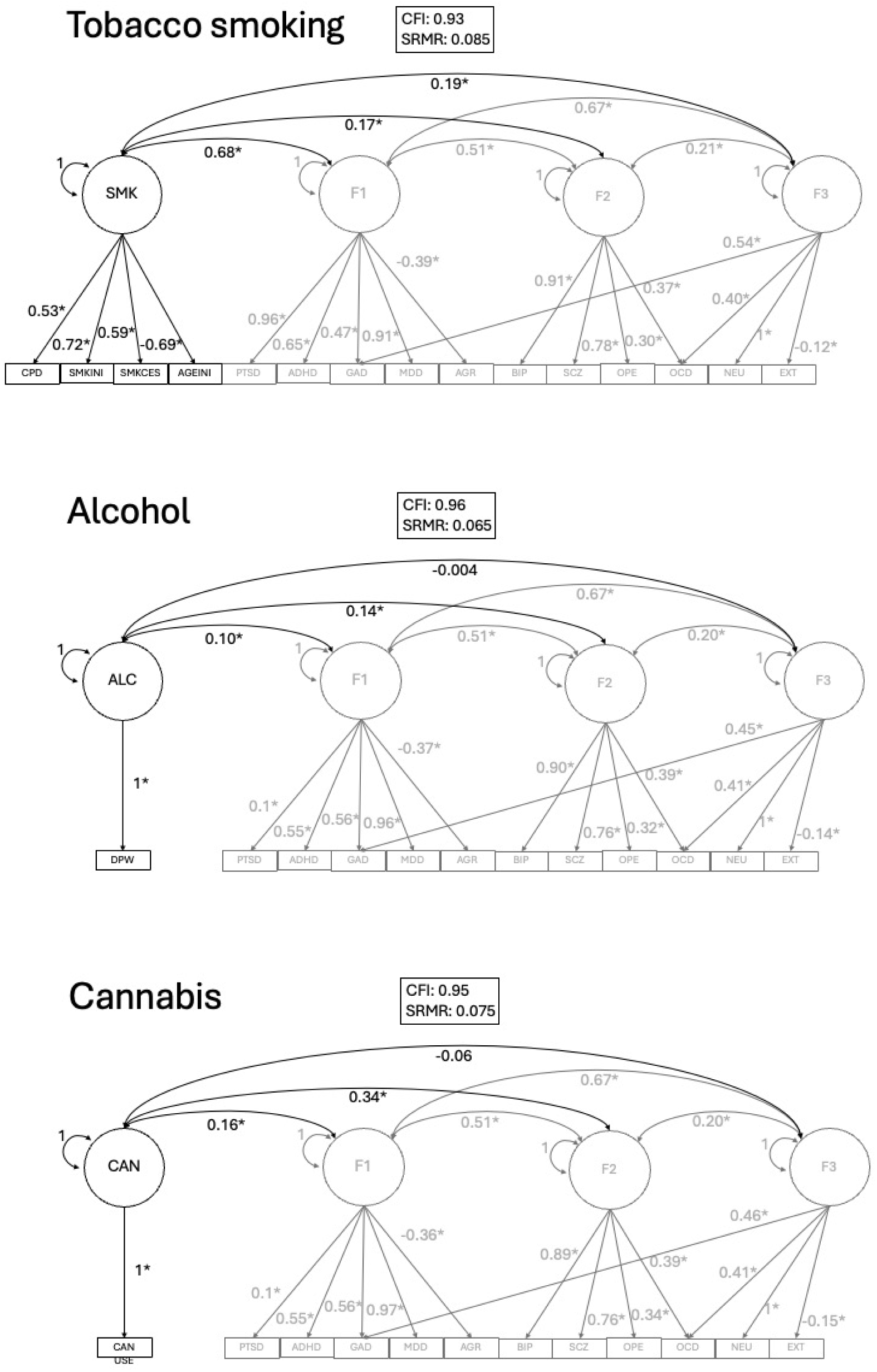
Genetic correlations between latent factors and substance use domains within gSEM model. Standardized factor loadings from the 3-factor model are represented in grey text and grey arrows. Substance use traits and factors are represented in black. Correlations between substance use factors and latent factors are represented in black. Circles represent latent factors, and rectangles are “observed” variables (Summary-level GWAS data). Asterisks represent significance at p<0.05. Statistical fit is shown in the box above the model. <u>Abbreviations</u>: PTSD = Post-Traumatic Stress Disorder, MDD = Major Depressive Disorder, SCZ = schizophrenia, BIP = Bipolar Disorder, GAD = General Anxiety Disorder, OCD = Obsessive-Compulsive Disorder, Ope = openness, Ext = extraversion, Agr = agreeableness, Neu = neuroticism, CPD = cigarettes per day, Smkini = smoking initiation, Smkces = smoking cessation, Ageini = age of smoking initiation, DPW = drinks per week, Can use = cannabis ever use

### Causal effects of substance use on personality and psychiatric phenotypes

Genetic instruments were selected at genome-wide significance (p < 5×10 ), except for Cannabis, for which <10 SNPs met this threshold. The significance criterion was therefore relaxed to p < 5×10 for this phenotype, warranting cautious interpretation of these results given the reduced instrument strength. The number of instruments per exposure ranged from 14 (age of smoking initiation) to 92 (smoking initiation and cigarettes per day) SNPs, with mean F ranging from 32.31 (Cannabis) to 165.08 (smoking initiation) (full instrument counts, F-statistics, and R² in Supplementary Table S1). We identified 32 significant causal effects for the primary IVW estimator after FDR correction (Figure 4; Supplementary Table T2); the remaining 34 pairs showed no evidence of a causal effect and were not evaluated further.

**FIGURE 4.**
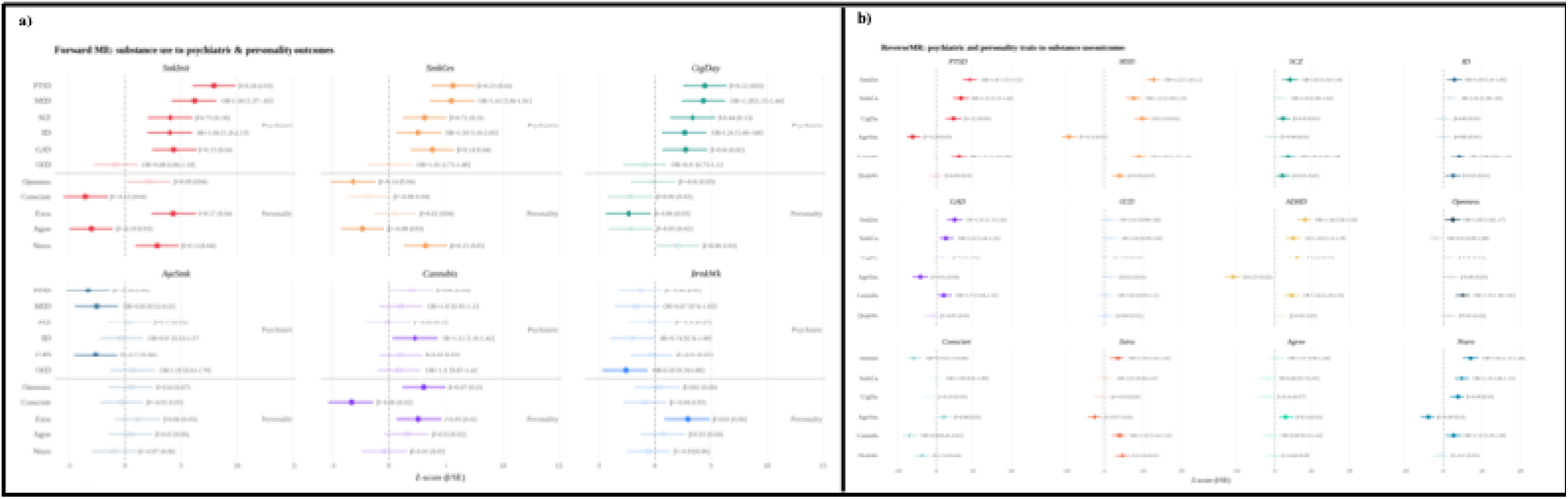
**a)** Putative causal effects of substance use behaviors on psychiatric and personality phenotypes. The forest plots show the IVW estimate, presented as Z-scores (b/SE) for comparability across outcome types, with error bars displaying the 95% CI. Filled points show estimates that were significantly different from zero (P_FDR_ <0.05). Effect sizes are presented for each association, as b(SE) for all continuous outcomes (PTSD, SCZ, GAD, Openness, Conscient, Extra, Agree, Neuro) and as OR (95% CI) for binary outcomes (MDD, BD, OCD). **b)** Putative causal effects of psychiatric and personality phenotypes on substance use behaviors. <u>Abbreviations</u>: PTSD = Post-Traumatic Stress Disorder, MDD = Major Depressive Disorder, SCZ = schizophrenia, BD = Bipolar Disorder, GAD = General Anxiety Disorder, OCD = Obsessive-Compulsive Disorder, Conscient = conscientiousness, Extra = extraversion, Agree = agreeableness, Neuro = neuroticism, SmkInit = smoking initiation, SmkCes = smoking cessation, CigDay = cigarettes per day, AgeSmk = age of smoking initiation, DrnkWk = drinks per week

Integrating evidence from the sensitivity analyses, five IVW-significant associations showed strong evidence of robust causal effects: smoking initiation → extraversion, smoking initiation → agreeableness, smoking cessation → BD, cigarettes per day → SCZ, and Cannabis → extraversion. Due to the lowered threshold that was applied for Cannabis, the effect of Cannabis on extraversion was not considered strong evidence. The remaining 28 significant associations (excluding age of smoking initiation → GAD, which was categorized as non-significant) showed weak evidence, most commonly reflecting inconsistency across complementary estimators, insufficient I²GX to support a reliable MR-Egger estimate, or evidence of directional pleiotropy. This categorization of evidence into strong, weak and no evidence is visualized in Figure 5, and explained more extensively in Supplementary Materials S6.

**FIGURE 5.**
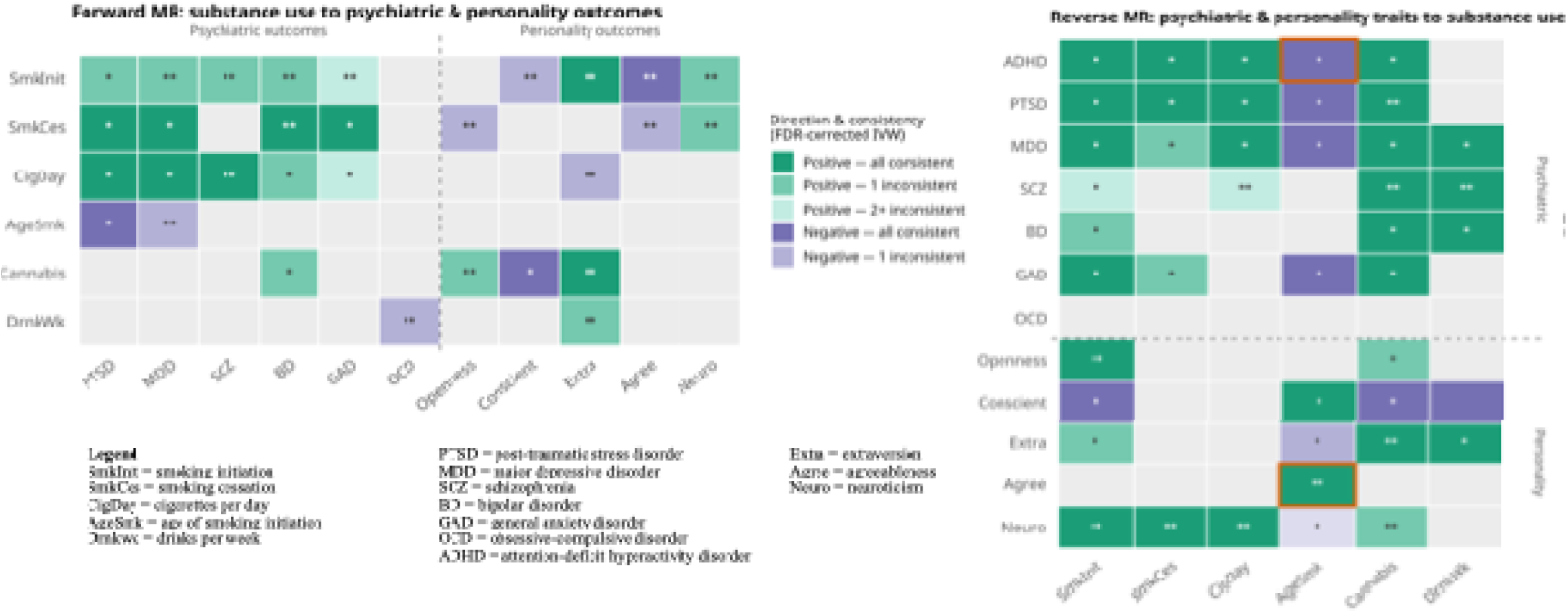
Heatmaps representing robustness of significant forward and reverse IVW results across the sensitivity analyses. Color intensity reflects directional consistency across complementary estimators (WM, WMo, SM, MR-Egger). Associations not remaining significant after Steiger filtering or MR-PRESSO outlier correction are displayed as non-significant. ** = pleiotropy-consistent; Egger intercept non-significant and MR-PRESSO distortion test non-significant. * = one pleiotropy check passed. Orange border = non-significant Cochran’s Q (heterogeneity) Grey = IVW-effect not FDR-significant <u>Abbreviations</u>: PTSD = Post-Traumatic Stress Disorder, MDD = Major Depressive Disorder, SCZ = schizophrenia, BD = Bipolar Disorder, GAD = General Anxiety Disorder, OCD = Obsessive-Compulsive Disorder, Conscient = conscientiousness, Extra = extraversion, Agree = agreeableness, Neuro = neuroticism, SmkInit = smoking initiation, SmkCes = smoking cessation, CigDay = cigarettes per day, AgeSmk = age of smoking initiation, DrnkWk = drinks per week

### Causal effects of personality and psychiatric phenotypes on substance use

Genetic instruments were selected at genome-wide significance (p < 5×10 ), except for OCD, for which only a single SNP met this threshold. The significance criterion was therefore relaxed to p < 1×10 ^6^ for this phenotype, warranting cautious interpretation of these results given the reduced instrument strength. Following clumping and exclusion of weak instruments, instrument counts ranged from 14 (OCD) to 305(Neuroticism), with mean F ranging from 26.94 (OCD) to 46.48 (Neuroticism) (Supplementary Table T9). Of the 72 tested pairs, 43 showed a statistically significant causal effect for the primary IVW estimator after FDR correction (Figure 8; Supplementary Table T10).

Across sensitivity analyses, nine IVW-significant associations showed strong evidence of causality: Neuroticism → smoking initiation, Neuroticism → smoking cessation, Neuroticism → cigarettes per day, PTSD → Cannabis, SCZ → Cannabis, SCZ → drinks per week, Extraversion → Cannabis, Openness → smoking initiation and Agreeableness → age of smoking initiation. Of these, Agreeableness → age of smoking initiation showed the most consistent robustness overall, as it was the only strong association without significant heterogeneity. The remaining 34 significant associations exhibited weak evidence of causality (Figure 5).

## DISCUSSION

We investigated the genetic overlap and potential causal relationships between common substance use behaviors and a broad range of psychiatric and personality phenotypes, to provide a clearer, more comprehensive overview of how substance use relates to common psychiatric disorders and personality traits. The gSEM analyses revealed a three-factor genetic architecture, with substance use domains showing distinct genetic correlations across the three latent factors. The MR analyses identified many causal relationships in the IVW analyses, though only a minority withstood all sensitivity analyses and were classified as showing strong evidence. Most significant associations showed heterogeneity, and many also showed evidence of horizontal pleiotropy, partly explaining why most findings were classified as of weak rather than strong evidence.

Our data-driven gSEM approach showed that a three-factor solution fit the data best. F1 combined internalizing disorders (PTSD, MDD, GAD) and ADHD with agreeableness (negative loading), F2 grouped psychotic-spectrum disorders (BD, SCZ) with OCD and openness, and F3 combined anxiety-related traits (neuroticism, OCD and GAD) with extraversion (negative loading), while conscientiousness did not load onto any factor. This factor structure suggests that shared genetic liability among psychiatric disorders, when combined with personality traits, may be organized differently than the often-hypothesized internalizing/externalizing distinction. Examining the relationships between substance use traits and these three factors showed smoking-related traits correlated most strongly with F1, cannabis showed a moderate genetic correlation specifically with F2, and alcohol showed uniformly weak correlations with all three factors. These results partly align with Pasman et al. (2025) [59], who reported weak genetic associations between alcohol use and psychiatric outcomes. They also reported an association between their latent cannabis factor, bipolar disorder and schizophrenia, broadly consistent with our finding that cannabis correlated with F2, onto which these two disorders loaded most strongly. This selectivity indicates that different substances may relate differently to the shared genetic architecture underlying psychiatric and personality traits, rather than showing a uniform pattern of association across all three factors. This contrasts with earlier gSEM studies which have identified broader genetic overlap between substance use and psychiatric disorders at the transdiagnostic level [60].

Overall, the MR findings provided stronger and more consistent evidence for causal effects of psychiatric and personality traits on substance use behaviors than in the other direction. This possibly reflects the more robust instruments and higher heritability of these traits. In the direction with substance use as the exposure, the strongest evidence for psychiatric traits involved smoking cessation and cigarettes smoked per day, exhibiting positive causal effects on bipolar disorder and schizophrenia, respectively, consistent with a causal risk factor model [61]. One plausible neurobiological pathway underlying the smoking-psychopathology associations involves nicotinic acetylcholine receptor (nAChR) signaling, whereby chronic exposure to nicotine dysregulates dopaminergic and glutamatergic transmission – pathways implicated in both psychotic-spectrum vulnerability and the reinforcing properties of smoking – alongside broader neuroinflammatory and oxidative stress mechanisms triggered by tobacco combustion [39, 62].

In the other direction, the most robust findings for psychiatric traits were found for schizophrenia and PTSD, with genetic liability for both predicting increased risk of cannabis use, and liability for schizophrenia additionally predicting greater alcohol consumption, consistent with a self-medication model. The effect of schizophrenia on cannabis use adds to a literature in which this association has been consistently reported but its causal direction has remained debated [24, 28, 63]. No strong evidence emerged for causal effects of any psychiatric disorder on smoking behaviors, in line with recent MR literature showing that this relationship predominantly occurs in the opposite direction [39, 64, 65]. IVW estimates for MDD, GAD, and PTSD on smoking outcomes are consistent with a self-medication model but proved inconsistent across sensitivity analyses, making it difficult to draw firm causal conclusions. The absence of robust evidence in this direction does not rule out a self-medication mechanism, but suggests that if present, it may be more modest or indirect than is sometimes assumed [24, 66]. Weak evidence was similarly found for ADHD, though here any underlying mechanism would more plausibly reflect externalizing traits such as impulsivity and behavioral disinhibition rather than the affect-regulating processes associated with internalizing disorders [67]. This similarity of effects is, however, in line with the latent grouping of ADHD with internalizing factors in our gSEM results.

Among personality traits, the strongest evidence of causal effects occurred in the direction of genetic liability for personality traits causally influencing risk of substance use behaviors, with six associations classified as strong evidence. The strongest evidence was found for neuroticism, where higher scores predicted increased smoking initiation, smoking cessation, and cigarettes smoked per day. Neuroticism is the personality trait most strongly correlated to psychopathology, in particular with MDD, and may reflect broader predispositions that increase susceptibility to maladaptive coping strategies, including substance use [38, 68]. In addition, we found strong evidence that higher agreeableness predicted increased age of smoking initiation, openness increased smoking initiation, and extraversion increased cannabis use, in line with earlier observational evidence [9, 69, 70]. The other way around, smoking initiation predicted increased extraversion and decreased agreeableness. Previous evidence for personality change following substance use is less common: some observational studies link sustained heavy alcohol use to personality shifts, but comparable effects for tobacco use remain weak and inconclusive [71]. This causal effect of smoking initiation on personality traits is therefore a novel finding and requires further investigation using longitudinal designs to better characterize the mechanisms and stability of such personality shifts.

The heterogeneity and pleiotropy observed across the MR analyses of these highly polygenic traits likely reflect multiple, biologically distinct pathways linking substance use and psychiatric disorders, lending support for a shared risk factor framework [31, 33]. This is corroborated by the gSEM analyses, which showed that a latent three factor model fits the genetic correlations among psychiatric phenotypes and personality traits well, with different substance use behaviors showing distinct genetic relationships with these factors. Critically, the variation in the robustness of MR associations across traits was not uniform. Psychotic-spectrum disorders BD and SCZ exhibited the strongest evidence for causal effects, whereas internalizing disorders showed strong associations under the IVW approach but substantial evidence of pleiotropy. Personality traits emerged primarily as risk factors for substance use traits, though the finding that substance use also exhibited causal effects on specific traits suggests that personality cannot be treated as a uniform liability category either, a distinction the latent factor structure also captured. This pattern of results is consistent with recent shifts in psychiatric nosology away from DSM-based categorical classification toward dimensional frameworks such as the Research Domain Criteria, the Hierarchical Taxonomy of Psychopathology, and the Network Theory of psychopathology, which conceptualize psychiatric phenotypes as quantitative spectra of shared and specific liability rather than discrete disease entities [19, 72–75]. Within such a framework, substance use behaviors may be differentially embedded across dimensions of psychopathology.

This study draws on some of the largest and most recent GWAS datasets available, enabling well-powered conclusions, reflecting patterns that single-exposure studies do not reveal. Several findings can help in resolving longstanding inconsistencies in the literature: the cannabis-schizophrenia finding supports a self-medicating effect rather than cannabis being a causal risk factor for schizophrenia; the consistent absence of causal effects of internalizing disorders on smoking nuances the assumed self-medication pathway; and the identification of robust causal effects across several personality traits – including both extensively studied traits such as neuroticism and extraversion, and less-studied traits such as openness and agreeableness. Notably, our finding of causal effects of smoking initiation on personality traits is one rarely tested in existing literature. Together with the correlations found between various personality traits and psychiatric phenotypes, and their grouping into latent factors in the gSEM analyses, this underscores that personality is broadly and consistently implicated in this relationship.

The findings of the present study should be interpreted in light of several limitations. For OCD, the exclusion of 23andMe data left only a single genome-wide significant instrument, necessitating a relaxed significance threshold to obtain a sufficient number of instruments, likely resulting in insufficient power to detect any effect. The significance threshold for cannabis use was relaxed as well, and while a robust signal was identified, associations involving cannabis as an exposure should be considered preliminary pending replication with a stricter threshold. The growing sample sizes of large psychiatric GWASs present the opposite challenge: as a fixed significance threshold of 5×10 is met by an increasing number of variants with small effect sizes and pleiotropic influences, heterogeneity and low I² values accumulate across analyses. Phenotype operationalization poses additional concerns; cannabis ever use as a binary variable captures heterogeneous patterns of use, due to one-time use being measured as equivalent to chronic use [48]. Another limitation related to the phenotypical design of this study is the unequal balance in substance use phenotypes: four smoking-related phenotypes were included, representing a range of consumption behaviors, but only single phenotypes for cannabis and alcohol were considered, as other candidate measures either lacked sufficient power to yield genome-wide significant instruments or captured problematic/dependence-related use rather than the habitual use patterns this study aimed to examine. Finally, analyses were restricted to individuals of European ancestry, reflecting the limited availability of well-powered GWAS in other ancestries for the examined traits, which reduces the generalizability of these findings to non-European populations. Moreover, even after excluding 23andMe and UKB samples to minimize cohort overlap across GWASs, there was still some sample overlap between exposure and outcome GWASs, which may bias MR estimates.

Future research should prioritize triangulating these genetic findings with observational and longitudinal study designs, allowing validation of the specific pathways identified and help clarify their underlying mechanisms, particularly for unexpected findings such as the causal effect of smoking initiation on personality traits [76]. Incorporating biologically informed instrument selection criteria could further improve the precision of causal estimates by prioritizing genetic variants with established functional relevance, an increasingly valuable approach as GWAS sample sizes continue to grow [77]. Clinically, these robust causal signals could inform future research aimed at identifying at-risk individuals and refining targeted intervention strategies, and more broadly, may help inform public health policy and prevention strategies.

In conclusion, this study provides insight into the complex genetic relationships of substance use behaviors with psychiatric disorders and personality traits. Our findings suggest that while substance use behaviors and psychiatric disorders and personality traits are frequently linked through shared genetic architecture, only a subset of these relationships reflect direct causal pathways. Additionally, substance use relates to broader trait dimensions rather than to isolated diagnostic categories alone, though many causal associations were affected by heterogeneity and pleiotropy and should be interpreted with caution. Together, these results deepen our understanding of the nature of substance use–psychopathology comorbidity, reinforcing that causality, shared liability, and diagnostic overlap all contribute to this relationship in ways that differ across specific traits and substances.

## Data Availability

All GWAS summary statistics analysed in this study are from previously published studies and are available online from the sources listed in the Data Availability Links section. All results generated in this study are contained in the manuscript and its Supplementary Materials.

## Acknowledgements

A.T and K.J.H.V are supported by the Dutch Research Council (NWO), Open Competition grant (project number 406.22.GO.043). K.J.H.V is supported by the Foundation Volksbond Rotterdam. J.A.P. was supported by the Amsterdam University Medical Center Postdoc Career Bridging grant (27527).

## Conflict of interest

The authors declare no conflict of interests.

## Data availability statement

All GWAS summary statistics analysed in this study are from previously published studies. Summary statistics are available at the following locations:

- Age of smoking initiation, cigarettes per day, drinks per week, smoking cessation, and smoking initiation (Saunders et al., 2022): https://conservancy.umn.edu/items/91f6a003-6af2-4809-9785-53dc579dc788
- Cannabis ever use (Pasman et al., 2018): https://www.ru.nl/bsi/research/group-pages-0/substance-use-addiction-food-saf/vm-saf/genetics/international-cannabis-consortium-icc/
- Agreeableness, extraversion, neuroticism, conscientiousness and openness (Schwaba et al., 2026): https://doi.org/10.17605/OSF.IO/HGNSM
- ADHD (van der Laan et al., 2025): https://www.ebi.ac.uk/gwas/studies/GCST90568441
- Bipolar disorder (O’Connell et al., 2025): https://figshare.com/ndownloader/files/49760772
- Generalized anxiety symptoms (Skelton et al., 2026): https://figshare.com/ndownloader/files/62613733
- Major depressive disorder (Adams et al., 2025): https://figshare.com/ndownloader/files/51487019
- Obsessive-compulsive disorder (Strom et al., 2025): https://figshare.com/ndownloader/files/53357735
- Post-traumatic stress disorder (Nievergelt et al., 2024): https://figshare.com/ndownloader/files/47850151
- Schizophrenia (Trubetskoy et al., 2022): https://figshare.com/ndownloader/files/34517828

## Code availability statement

The code used for the analyses in this study will be made publicly available online upon acceptance of the paper.

## Notes

### Competing Interest Statement

The authors have declared no competing interest.

### Author Declarations

All GWAS summary statistics analysed in this study are from previously published studies. Summary statistics are available at the following locations: Age of smoking initiation, cigarettes per day, drinks per week, smoking cessation, and smoking initiation (Saunders et al., 2022): https://conservancy.umn.edu/items/91f6a003-6af2-4809-9785-53dc579dc788 Cannabis ever use (Pasman et al., 2018): https://www.ru.nl/bsi/research/group-pages-0/substance-use-addiction-food-saf/vm-saf/genetics/international-cannabis-consortium-icc/ Agreeableness, extraversion, neuroticism, conscientiousness and openness (Schwaba et al., 2026): https://doi.org/10.17605/OSF.IO/HGNSM ADHD (van der Laan et al., 2025): https://www.ebi.ac.uk/gwas/studies/GCST90568441 Bipolar disorder (O'Connell et al., 2025): https://figshare.com/ndownloader/files/49760772 Generalized anxiety symptoms (Skelton et al., 2026): https://figshare.com/ndownloader/files/62613733 Major depressive disorder (Adams et al., 2025): https://figshare.com/ndownloader/files/51487019 Obsessive-compulsive disorder (Strom et al., 2025): https://figshare.com/ndownloader/files/53357735 Post-traumatic stress disorder (Nievergelt et al., 2024): https://figshare.com/ndownloader/files/47850151 Schizophrenia (Trubetskoy et al., 2022): https://figshare.com/ndownloader/files/34517828

