## Supplementary materials for "Latent Genetic Factors and Causal Pathways Linking Substance Use to Psychiatric and Personality Phenotypes: a Genomic SEM and Mendelian Randomization Study"

**SUPPLEMENTARY METHODS**

**INDEX**

**S1. Overview of GWAS studies**	………………………………………………….	2

**S2. Sample overlap**	………………………………………………………………..	4

**S3. Phenotype descriptions**	………………………………………………………..	4

Substance use phenotypes	………………………………………………….	4

Psychiatric phenotypes	……………………………………………………..	6

Personality phenotypes	……………………………………………………..	8

**S4. Main methods**	………………………………………………………………….	8

**S5. Sensitivity analyses**	…………………………………………………………….	9

**S6. Results**	………………………………………………………………………..	10

**References**	………………………………………………………………………………16

**Plots**	……………………………………………..………………………………..	18

P1. Forwards funnel plots	……………………………………………………..	18

P2. Forwards scatter plots	……………………………………………………..	52

P3. Forwards leave-one-out plots	……………………………………………	86

P4. Reverse funnel plots	……………………………………………………..	120

P5. Reverse scatter plots	……………………………………………………..	157

P6. Reverse leave-one-out plots ……………………………………………………193

S1. Overview of GWAS studies

Characteristics of the GWAS from which the summary statistics were obtained.

| Trait | Reference | Cases | Controls | Total N | MR #SNPs | SNP h^2^ | SNP h^2^ SE | Link to download summary statistics |
| --- | --- | --- | --- | --- | --- | --- | --- | --- |
| Age of smoking initiation | Saunders et al., 2022 | - | - | 618,541 | 14 | 0.047 | 0.0027 | <https://conservancy.umn.edu/items/91f6a003-6af2-4809-9785-53dc579dc788> |
| Agreeableness | Schwaba et al., 2026 | - | - | 567.556 | 17 | 0.0339 | 0.0018 | <https://doi-org.vu-nl.idm.oclc.org/10.17605/OSF.IO/HGNSM> |
| Attention-Deficit Hyperactivity Disorder (ADHD) | van der Laan et al., 2025 | - | - | 290,134 | 39 | 0.102 | 0.0044 | <https://www.ebi.ac.uk/gwas/studies/GCST90568441> |
| Bipolar Disorder (BD) | O'Connell et al., 2025 (PGC) | 59,287 | 781,022 | BIN | 68 | 0.1137 | 0.0054 | https://figshare.com/ndownloader/files/49760772 |
| Cannabis Ever Use | Pasman et al., 2018 | - | - | 184,765 | 17 | 0.0671 | 0.004 | <https://www.ru.nl/bsi/research/group-pages-0/substance-use-addiction-food-saf/vm-saf/genetics/international-cannabis-consortium-icc/> |
| Cigarettes per day | Saunders et al., 2022 | - | - | 618,489 | 92 | 0.0737 | 0.0038 | <https://conservancy.umn.edu/items/91f6a003-6af2-4809-9785-53dc579dc788> |
| Conscientiousness | Schwaba et al., 2026 | - | - | 598.047 | 68 | 0.0505 | 0.0019 | <https://doi-org.vu-nl.idm.oclc.org/10.17605/OSF.IO/HGNSM> |
| Drinks per week | Saunders et al., 2022 | - | - | 2,428,851 | 79 | 0.0543 | 0.0024 | <https://conservancy.umn.edu/items/91f6a003-6af2-4809-9785-53dc579dc788> |
| Extraversion | Schwaba et al., 2026 | - | - | 619.416 | 105 | 0.0651 | 0.0021 | <https://doi-org.vu-nl.idm.oclc.org/10.17605/OSF.IO/HGNSM> |
| Generalized anxiety symptoms (GAD) | Skelton et al., 2026 (PGC) | - | - | 693,869 | 66 | 0.0495 | 0.0019 | https://figshare.com/ndownloader/files/62613733 |
| Major Depressive Disorder (MDD) | Adams et al., 2025 | 412,305 | 1588397 | BIN | 198 | 0.0452 | 0.0014 | https://figshare.com/ndownloader/files/51487019 |
| Neuroticism | Schwaba et al., 2026 | - | - | 1,088,604 | 305 | 0.0701 | 0.0026 | <https://doi-org.vu-nl.idm.oclc.org/10.17605/OSF.IO/HGNSM> |
| Obsessive-Compulsive Disorder (OCD) | Strom et al., 2025 (PGC) | 23,493 | 1,114,613 | BIN | 14 | 0.1247 | 0.0078 | https://figshare.com/ndownloader/files/53357735 |
| Openness | Schwaba et al., 2026 | - | - | 568.489 | 64 | 0.0525 | 0.0021 | <https://doi-org.vu-nl.idm.oclc.org/10.17605/OSF.IO/HGNSM> |
| Post-Traumatic Stress Disorder (PTSD) | Nievergelt et al., 2024 (PGC) | 137,136 | 1,085,746 | BIN | 67 | 0.0708 | 0.0027 | <https://figshare.com/ndownloader/files/47850151> |
| Schizophrenia (SCZ) | Trubetskoy et al., 2022 (PGC) | 53,386 | 77,258 | BIN | 155 | 0.3823 | 0.0129 | <https://figshare.com/ndownloader/files/34517828> |
| Smoking cessation | Saunders et al., 2022 | 270,310 | 677,350 | 947,660 | 84 | 0.0559 | 003 | <https://conservancy.umn.edu/items/91f6a003-6af2-4809-9785-53dc579dc788> |
| Smoking initiation | Saunders et al., 2022 | 973,419 | 1,247,411 | 2,220,830 | 92 | 0.0875 | 0.0029 | <https://conservancy.umn.edu/items/91f6a003-6af2-4809-9785-53dc579dc788> |

**Table S1.** Characteristics of the GWAS from which the summary statistics were obtained.

CONT = continuous variable, BIN = binary variable.

S2. Sample overlap

To minimize bias from sample overlap between exposure and outcome GWAS - a known source of inflation in MR effect estimates [1] - exposures and outcomes were drawn from non-overlapping samples where possible. Specifically, 23andMe summary statistics were included in the substance use GWAS but excluded from all psychiatric and personality samples, while UK Biobank data were excluded from the substance use data and retained for the psychiatric and personality traits. As 23andMe summary statistics are not publicly available in full, only the top 10,000 SNPs were available for use here.

S3. Phenotype descriptions

**Substance use phenotypes.**

The smoking and alcohol phenotypes summary statistics were obtained from the publicly available summary statistics by the GSCAN consortium [2] and represent different stages of substance use. Whenever possible, measures of smoking include cigarette smoking only, as use of other nicotine delivery types were rarely reported across studies. The one exception was for the Amish cohort in the Trans-Omics for Precision Medicine (TOPMed) project, as nearly all individuals in that study smoked small cigars.

Smoking initiation

1. Binary phenotype with any participant reporting ever being a regular smoker in their life (current or former) coded “2”, while any participant who reported never being a regular smoker in their life coded “1”.
2. Does not include information about pipes/cigar/chew, or other non-cigarette forms of tobacco use.
3. This phenotype was measured in a variety of ways.

*a. Have you smoked over 100 cigarettes over the course of your life?*

*b. Have you ever smoked every day for at least a month?*

*c. Have you ever smoked regularly?*

Smoking Cessation

1. Binary phenotype with current smokers coded as “2”, former smokers coded as “1”, and never smokers coded as missing.
2. Does not include information about pipes/cigars/chew, or other non-cigarette forms of tobacco use.
3. Usually measured through a combination of questions, including:

a. *Do you currently smoke?* and *Have you ever smoked regularly?*

b. *Do you smoke?* and *Have you smoked over 100 cigarettes in your entire life?*

Cigarettes per day

1. Defined as the average number of cigarettes smoked per day, either as a current smoker or former smoker. Individuals who either never smoked, or for whom there is no available data (e.g., someone was a former smoker, but for whom former smoking was never assessed) were set to missing.
2. For studies that collected a quantitative measure of cigarettes per day, where the respondent is free to provide any integer (e.g., 13 cigarettes per day) responses were binned as follows.

a. 1 = 1–5

b. 2 = 6–15

c. 3 = 16–25

d. 4 = 26–35

e. 5 = 36+

3. For studies with pre-defined bins, the pre-defined bins were used.

4. Does not include information about pipes/cigars/chew, or other non-cigarette forms of tobacco use.

5. Cigarettes per day was measured with a single question for most contributing studies using, for example:

*a. How many cigarettes do you smoke per day?*

*b. How many cigarettes did you smoke per day?*

Age of smoking initiation

1. Age (in years) at which an individual started smoking cigarettes regularly
2. Does not include information about pipes/cigars/chew, or other non-cigarette forms of tobacco use.
3. Measured in a variety of ways:

*a. At what age did you begin smoking regularly?*

b. *How long have you smoked?* Combined with *What is your current age?*

Drinks per week

1. Defined as the average number of drinks a participant reported drinking each week, aggregated across all types of alcohol. If a study recorded binned response ranges (e.g., 1–4 drinks per week, 5–10 drinks per week) the midpoint of the range was used. For example, if an individual reported 1–5 drinks per week, it was assumed they drank 2.5 drinks per week on average.

2. This was measured in a variety of ways.

*a. In the past week, how many alcoholic beverages did you have?*

*b. Thinking about the past year, on the average how many drinks did you have each week?*

3. This phenotype was left-anchored at 1 and log-transformed prior to analysis, in order to prevent outliers from having undue leverage on analyses.

Cannabis ever use

Cannabis ever use was defined as having used cannabis at least once in one's lifetime (cases) versus never having used cannabis (controls). GWAS summary statistics were derived from the most recent GWAS meta-analysis by the International Cannabis Consortium [3], which combined data across 34 cohorts with the largest contributing cohorts derived from 23andMe Research Institute, All of Us, and UK Biobank (N=556,258 with 273,372 cases) using effect sizes weighted by inverse standard errors in METAL. The version used for the present analyses included data from 23andMe but excluded UK Biobank.

**Psychiatric phenotypes**

Attention-Deficit Hyperactivity Disorder (ADHD)

ADHD symptom GWAS data were derived from a large collaborative meta-analysis coordinated within the ACTION and EAGLE consortia, drawing on 28 cohorts (154 individual GWASs, 290,134 observations from 70,953 unique individuals) spanning population-based and clinical samples across Europe, the US, Australia, and New Zealand, including twin registries and birth/pregnancy cohorts [4]. ADHD symptoms were assessed in children and adolescents (ages 1.5–18 years), with additional adult retrospective assessments in two cohorts, using one of 20 standardized instruments, most commonly the Achenbach System of Empirically Based Assessment (e.g., CBCL, TRF, YSR) and the Strengths and Difficulties Questionnaire (SDQ), rated by mothers, fathers, teachers, or the individuals themselves. DNA for genotyping was collected via blood, saliva, or buccal swabs, generally during childhood or adolescence depending on cohort protocol. Cohort-specific GWAS analyses were conducted locally by each cohort's researchers following a standardized protocol, with separate analyses performed for every unique combination of rater, instrument, and age (maximum one measure per individual per GWAS, minimum 450 observations per GWAS). All data collections were approved by local ethics committees, with overall study approval from the Central Ethics Committee on Research Involving Human Subjects of the VU University Medical Center, Amsterdam.

Bipolar Disorder (BD)

Bipolar disorder summary statistics came from the latest release of the PGC Bipolar Disorder Working group [5]. Cases and controls were drawn from clinical (55 cohorts), community (20 cohorts), and self-reported (four 23andMe cohorts) samples, with cases ascertained via semi-structured/structured interviews, medical records and registries, or self-reported diagnosis/treatment via web-based surveys, respectively. Controls were generally screened for absence of lifetime psychiatric disorders, though some unscreened community controls were included given BD's low population prevalence. As 23andMe data were excluded from our analyses, we used the non-23andMe European-ancestry subset: 59,287 cases and 781,022 controls (derived from the reported totals of 131,969 cases/2,322,416 controls including 23andMe, minus the 72,682 cases/1,541,394 controls contributed by 23andMe).

Generalized Anxiety Disorder (GAD)

Generalized anxiety symptom data came from a PGC Anxiety Disorders Working Group meta-analysis [6] of 693,869 European-ancestry individuals across 14 cohorts (e.g., ALSPAC, UK Biobank, Estonian Biobank, Generation Scotland, MoBa, TEDS, TRAILS, Million Veteran Program), combining 13 PGC-ANX cohorts with pre-existing GWAS summary statistics by the Million Veteran Program (2020) [7]. The majority (70%) of participants completed the GAD-7 or closely related brief self-report measures of recent anxiety symptoms, with the remainder assessed using other validated brief anxiety scales; total symptom sum scores were analyzed, with missing items (<25%) imputed from each participant's mean score. No 23andMe data were included in this GWAS.

Major Depressive Disorder (MDD)

Major depression case-control data came from the third PGC MDD GWAS, combining a core European-ancestry sample (MDD49, building on Wray et al. [8]) with 24 additional European-ancestry cohorts and further multi-ancestry cohorts, for a total discovery sample of 688,808 cases and 4,364,225 controls across ancestries [9]. Case status was based on clinical diagnosis, medical records, or (in some cohorts) self-report, with cases and controls as defined by original cohort investigators. As 23andMe contributed a substantial share of the European-ancestry sample (112,892 cases/1,773,938 controls out of a total 525,197 cases/3,362,335 controls), we used only the non-23andMe European-ancestry subset: 412,305 cases and 1,588,397 controls.

Obsessive-Compulsive Disorder (OCD)

OCD case-control data came from the PGC OCD-3 GWAS, comprising 28 cohorts and a total of 53,660 cases and 2,044,417 controls across ancestries [10]. Roughly 40% of cases met DSM-5 or ICD-10 criteria as assessed by a healthcare professional or derived from (electronic) health records, while the remainder were based on self-reported diagnosis (including 23andMe, AGDS, and part of UK Biobank). Since 23andMe data were excluded from our analyses, we used the non-23andMe European-ancestry summary statistics: 23,493 cases and 1,114,613 controls.

Post-Traumatic Stress Disorder (PTSD)

The PTSD data came from the GWAS meta-analysis by the PGC PTSD Working Group [11]. It combined studies that assessed PTSD as either a case/control diagnosis or as continuous symptom scores; because these measures are on different scales (log odds vs. symptom score change), beta estimates were harmonized using the transformation formulae described by Nievergelt et al. (PMID: 27019110), which preserve relative effect size and direction (positive = risk increasing, negative = risk decreasing) but do not provide a directly interpretable causal effect magnitude in MR – significant MR estimates for this exposure/outcome can therefore only be interpreted directionally as risk-increasing or risk-decreasing. Data came from ten EHR-based cohorts plus the Million Veteran Program (MVP), including four US sites (BioVU, Mass General Brigham Biobank, BioMe, MayoGC) and six non-US sites (iPSYCH, FinnGen, HUNT, STR-STAGE, UK Biobank, Estonian Biobank), totaling 817,181 participants (78,687 cases, 9.6%). Cases were defined broadly as individuals with at least one ICD-9/10 code for PTSD or another stress disorder, with all remaining participants classified as controls. PTSD assessment and DNA collection followed each study's own protocol, with all studies approved by relevant institutional review boards (including UCSD IRB protocol 16097x) and written informed consent obtained from all participants.

Schizophrenia (SCZ)

The schizophrenia data was derived from the GWAS by the PGC Schizophrenia Working Group [12]. This was a dataset of 90 cohorts with individual-level genotype data processed under a uniform pipeline, comprising 161,405 unrelated individuals (67,390 SCZ/schizoaffective cases, 94,015 controls; ~80% of European ancestry), with a parent–proband trio counted as one case and one control. This was meta-analysed with an additional 1,979 cases and 142,626 controls of European ancestry from deCODE genetics, yielding a final analysis of 320,404 diploid genomes; neither UK Biobank nor 23andMe data were included. Across cohorts, cases were predominantly ascertained from psychiatric inpatient and outpatient services, hospital registers, or clinical trial populations, with diagnoses established via structured or semi-structured clinical interviews (e.g., SCID, SCAN, DIGS) combined with medical record review and confirmed according to DSM-III/IV or ICD-10 criteria following a best-estimate consensus procedure. Controls were generally population-based (e.g., blood donors, biobank participants, or registry samples) and, where possible, screened to exclude a personal or family history of psychiatric illness, though some cohorts used unscreened general-population controls matched on ancestry and genotyping platform. All studies obtained written informed consent and institutional/ethics committee approval, with genotyping performed at centralized facilities (e.g., Broad Institute, deCODE, Affymetrix) to harmonize data across the consortium, and samples recruited in mainland China were genotyped and analysed locally in compliance with Chinese Human Genetic Resources Administrative Regulations.

**Personality phenotypes (Extraversion, Agreeableness, Conscientiousness, Neuroticism, Openness)**

GWAS summary statistics were obtained from the Revived Genomics of Personality Consortium (ReGPC), which meta-analyzed data across 46 cohorts [13]. Each Big Five trait was assessed via multi-item self-report questionnaires, predominantly validated Big Five instruments such as the Big Five Inventory or NEO Personality Inventory-Revised (k = 39 cohorts), with a subset of cohorts contributing theoretically congruent scales from the Eysenck Personality Questionnaire (extraversion and neuroticism only; k = 4) or the HEXACO inventory (extraversion, conscientiousness, and openness; k = 1). Item responses were averaged into continuous composite scores and standardized (M = 0, SD = 1) prior to GWAS. As effect allele frequency (EAF) was not reported in the original summary statistics, EAF was derived by merging with a MAF/EAF crosswalk file using a unique variant identifier (chromosome, position, and alphabetically first allele), with EAF assigned as MAF or 1 − MAF depending on whether the minor allele corresponded to the effect allele. Full cohort-level phenotyping, instrument congruence analyses, and quality control are described in the original publication's supplementary materials. Sample sizes used for the individual personality traits are reported in table S1.

S4. gSEM methods

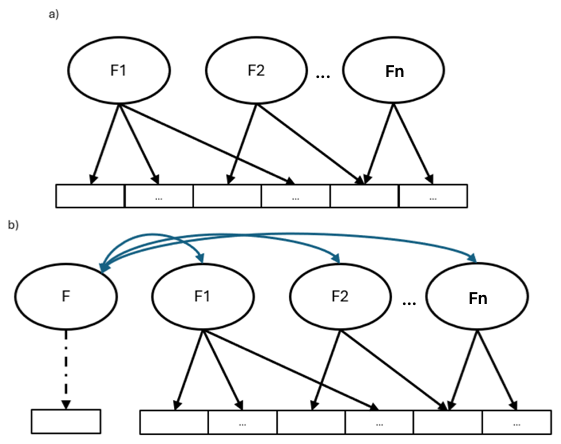

**SUPPLEMENTARY FIGURE 1. a)** Illustrative representation of a multi-factor gSEM model. Observed traits (which correspond to GWAS summary statistics) are represented as squares, latent factors as circles. Observed traits load onto one or multiple factors (arrows) in this example. **b)** The same model with additional genetic correlations with another factor. An observed trait is first modelled as a latent factor (F). F is allowed to correlate with the latent factors.

S5. MR methods

IVW estimates may be biased in the presence of horizontal pleiotropy, and thus four additional MR methods were applied to assess robustness: the weighted median (WM) estimator, which yields consistent estimates when up to 50% of IVs are invalid [14]; the weighted mode (WMo) estimator, which provides unbiased estimates when the largest cluster of IVs shares the same causal effect; the simple mode estimator, which assumes that the most common IV estimate approximates the true causal effect [15]; and MR-Egger regression slope, which relaxes the exclusion restriction assumption by allowing all instruments to exhibit pleiotropy, provided their pleiotropic effects are uncorrelated with instrument strength (the InSIDE assumption) [16].

The I²_GX_ statistic was used to evaluate the degree to which the NO Measurement Error (NOME) assumption was violated in MR-Egger analyses. The NOME assumption requires that genetic variant–exposure association estimates are measured without error; in practice this is never perfectly satisfied, and when violated it introduces regression dilution bias that attenuates the MR-Egger estimate toward the null [17]. The I²_GX_ quantifies the severity of this violation, with values approaching 1 indicating minimal measurement error. Where I²_GX_ > 0.9, instruments were included in MR-Egger analyses directly. Where 0.6 ≤ I²_GX_ ≤ 0.9, SIMEX (Simulation Extrapolation) correction was applied to adjust for the resulting attenuation bias [18]. MR-Egger results of instruments with I²_GX_ < 0.6 were not interpreted, as correction in this range is considered unreliable.

The robustness of the IVW estimates was evaluated through a series of sensitivity analyses. To verify the assumed causal direction for each exposure-outcome pair, Steiger filtering was applied, testing whether the genetic variants explain more variance in the exposure than in the outcome [19]. MR analyses were run both with and without Steiger filtering, and results were compared to assess whether directionality or instrument validity influenced the findings. Consistency of effect estimates across both analyses was taken as evidence of robustness; discrepancies were flagged for closer inspection. Heterogeneity among IV estimates was assessed using Cochran's Q statistic, with a significant Q (p < 0.05) taken as indicative of potential violations of the exclusion restriction assumption. The MR-Egger intercept test was used to detect directional horizontal pleiotropy, with a non-zero intercept suggesting that genetic variants systematically affect the outcome through pathways independent of the exposure [16]; a non-significant intercept indicates an absence of detectable directional pleiotropy, supporting the validity of the IVW estimate. Where the I²_G_X criterion for SIMEX correction was met, the corrected MR-Egger estimate was interpreted instead. MR-PRESSO was applied to test for the presence of outlying instruments and to provide corrected causal estimates following outlier removal [20]. Results were visualized using scatter plots, forest plots, funnel plots and leave-one-out plots to facilitate interpretation of individual instrument contributions and overall effect consistency. Convergence of results across the primary IVW analysis, complementary effect estimators, and sensitivity analyses was used as the main criterion for concluding robustness of causal estimates. Significant effects that remained consistent across all these analyses, were classified as ‘strong evidence’ of causality; significant effects that did not remain consistent across all the analyses as ‘weak evidence’.

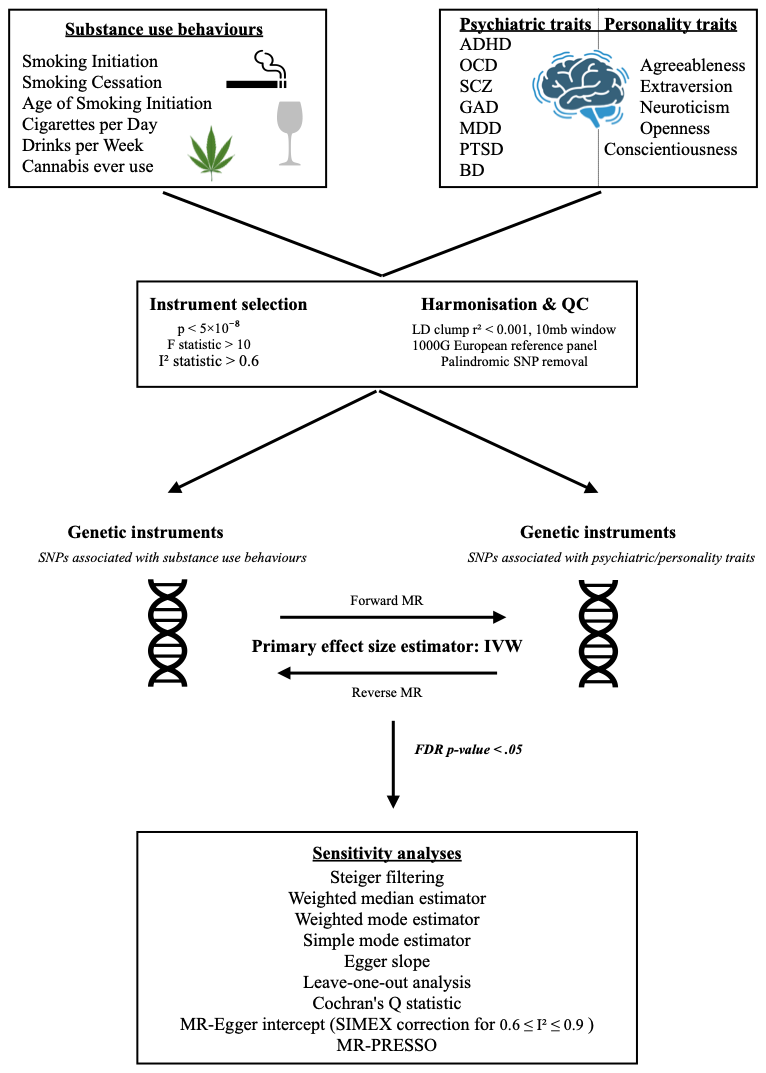

**SUPPLEMENTARY FIGURE 2.** Overview of the bidirectional two-sample Mendelian randomization workflow. Six substance use phenotypes and twelve psychiatric and personality phenotypes served as exposures and outcomes in the forward and reverse directions, respectively (ADHD excluded as an outcome in the forward direction).

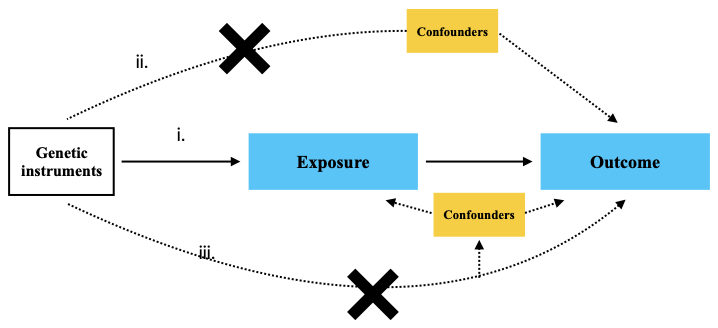

**SUPPLEMENTARY FIGURE 3**. Diagram of instrumental variable assumptions for Mendelian Randomization. The three assumptions (i, ii, iii) are illustrated by the presence of an arrow, indicating (i.) a strong association between genetic variants and the exposure (relevance assumption); (ii.) genetic variants are independent of potential confounders that could influence the relationship between the exposure and the outcome (independence assumption); and (iii.) genetic variants do not affect the outcome, except through their association with the exposure (exclusion restriction assumption).

#### S6. Results

**Causal effects of substance use on personality and psychiatric phenotypes**

Sensitivity analyses were used to evaluate the robustness of the 32 significant causal effects found in the IVW analyses. Steiger filtering confirmed the expected causal direction for all interpretable associations. SNPs were removed for 16 associations (1.1–12.3% of instruments), and all IVW estimates remained significant (p < 0.05) after filtering. The largest reduction was for smoking cessation → SCZ (10 SNPs removed), for which the estimate attenuated from 0.72 to 0.38 (p = 0.041). I²GX values indicated that MR-Egger could be interpreted directly (I² > 0.9) for 10 pairs and required SIMEX correction (0.6 ≤ I²GX ≤ 0.9) for 19 pairs. The remaining three associations, age of smoking initiation on PTSD (I²GX = 0.599), age of smoking initiation on GAD (I²GX = 0.599), and cannabis use on BD (I²GX = 0), fell below this threshold and were consequently classified as showing, at most, weak evidence. Effect size estimators for the WM, WMo, simple mode, and MR-Egger analyses were directionally consistent with IVW for 13 associations; the remaining 19 showed inconsistency in at least one estimator (up to three for cigarettes per day → GAD and smoking cessation → SCZ), warranting cautious interpretation. Significant heterogeneity (Cochran's Q, p < 0.05) and global pleiotropy (MR-PRESSO) were detected across all FDR-significant associations. The MR-PRESSO distortion test indicated that outlier removal did not meaningfully alter causal estimates for 29 of 32 pairs. The three exceptions, cannabis → conscientiousness, smoking cessation → SCZ, and age of smoking initiation → GAD, showed significant distortion (p = 0.006, 0.008, and 0.019, respectively). Age of smoking initiation → GAD lost significance following outlier correction (pPRESSO = 0.130) and was excluded from further interpretation on this basis. The other two remained significant after correction (p = 0.006 and 0.0095) but with attenuated estimates (−0.060 → −0.038 and 0.72 → 0.44), so they should be interpreted with caution. Directional pleiotropy, indicated by a significant MR-Egger intercept, was detected for 8 of the 31 remaining associations (Supplementary Table T7). Visual inspection of the diagnostic plots (Scatter, Leave-One-Out, and Funnel plots; presented in Supplementary Materials P1 – P3) revealed no further inconsistencies.

**Causal effects of personality and psychiatric phenotypes on substance use traits**

Steiger filtering confirmed the expected causal direction for all 43 reverse-direction associations, with instrument removal rates generally low (0.5–1.5%). I²GX values were below 0.6 for the majority of associations - including all PTSD (except for age of smoking initiation and Cannabis), GAD, BD, MDD, ADHD, conscientiousness and extraversion (except for Cannabis) pairs - meaning MR-Egger could not be interpreted for these traits. These associations were consequently classified as showing, at most, weak evidence for causality. For neuroticism, SCZ, openness, and agreeableness associations, and for extraversion → Cannabis, PTSD → age of smoking initiation, and PTSD → Cannabis, SIMEX correction was applied (corrected estimates in Supplementary Table T12). Effect size estimators were directionally consistent with IVW for 33 of the 43 significant associations (Supplementary Table T10). Significant Cochran's Q heterogeneity and MR-PRESSO pleiotropy were detected for the majority of associations, with the exceptions of ADHD → age of smoking initiation and agreeableness → age of smoking initiation, for which neither was detected. The distortion test was non-significant for all pairs except SCZ → smoking initiation (p_DISTORTION_ = 0.013) and conscientiousness → drinks per week (p_DISTORTION_ = 0.022), but both pairs remained significant after outlier correction, indicating that outlier removal did not meaningfully alter the causal estimates. Directional pleiotropy, indicated by a significant MR-Egger intercept, was observed for openness → Cannabis (p = 0.023) and Neuroticism → age of smoking initiation (p = 0.004); intercepts were not interpreted for the 28 associations with I²GX < 0.6. Visual inspection of the diagnostic plots (Scatter, Leave-One-Out, and Funnel plots; presented in Supplementary Material P4 – P6) revealed no further inconsistencies.

**PLOTS**

### **Forwards analyses**

P1. Funnel plots

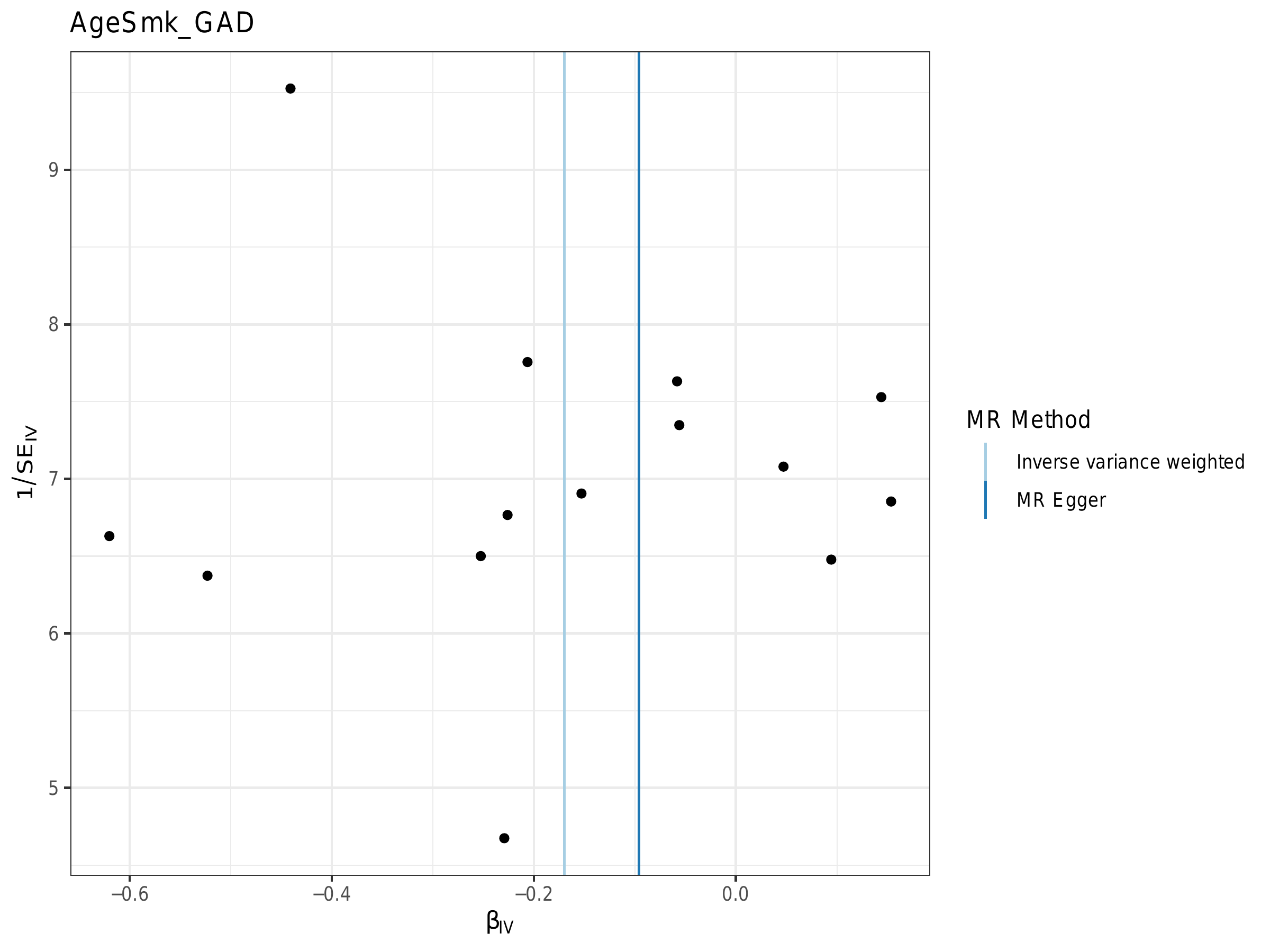

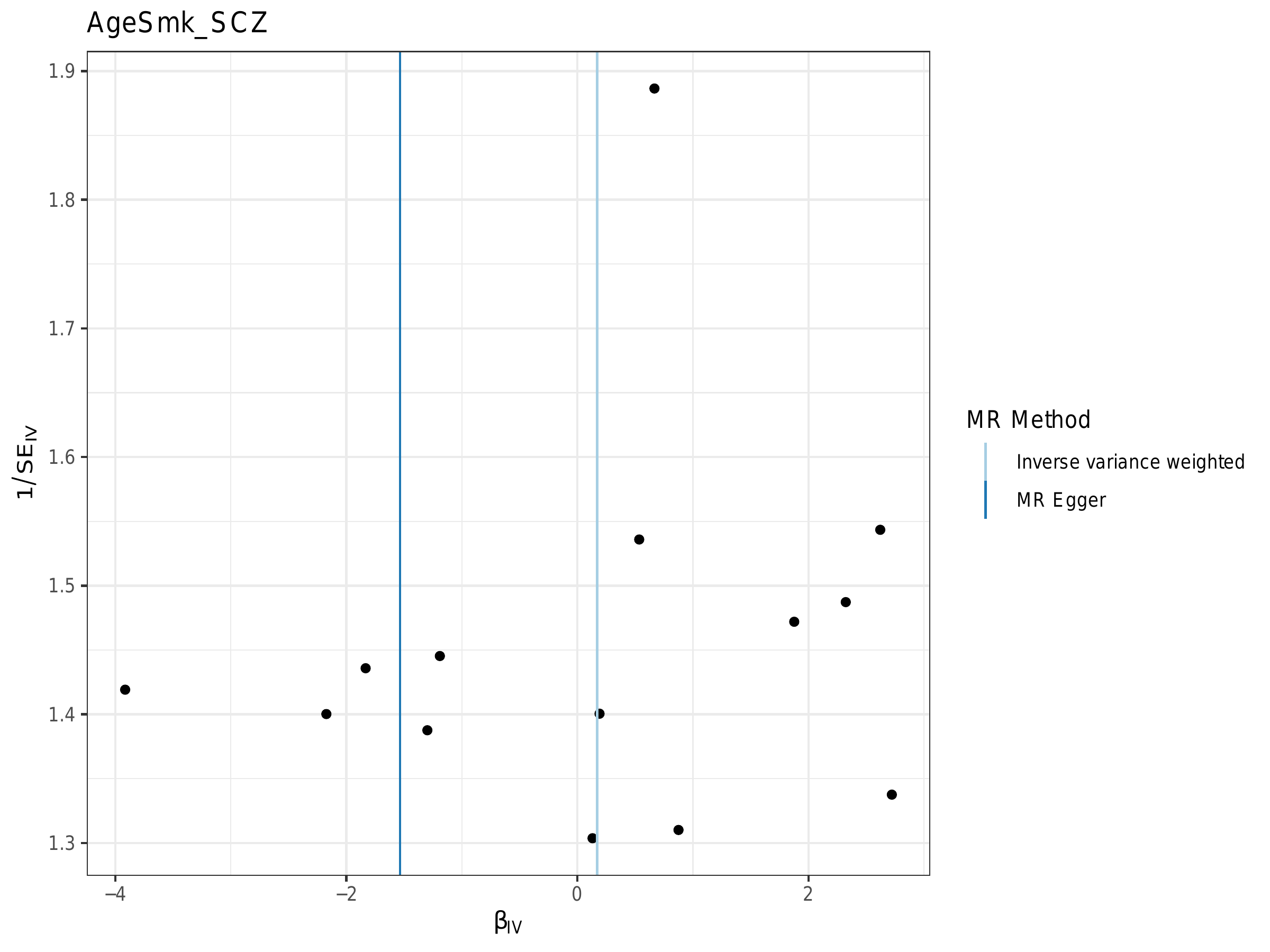

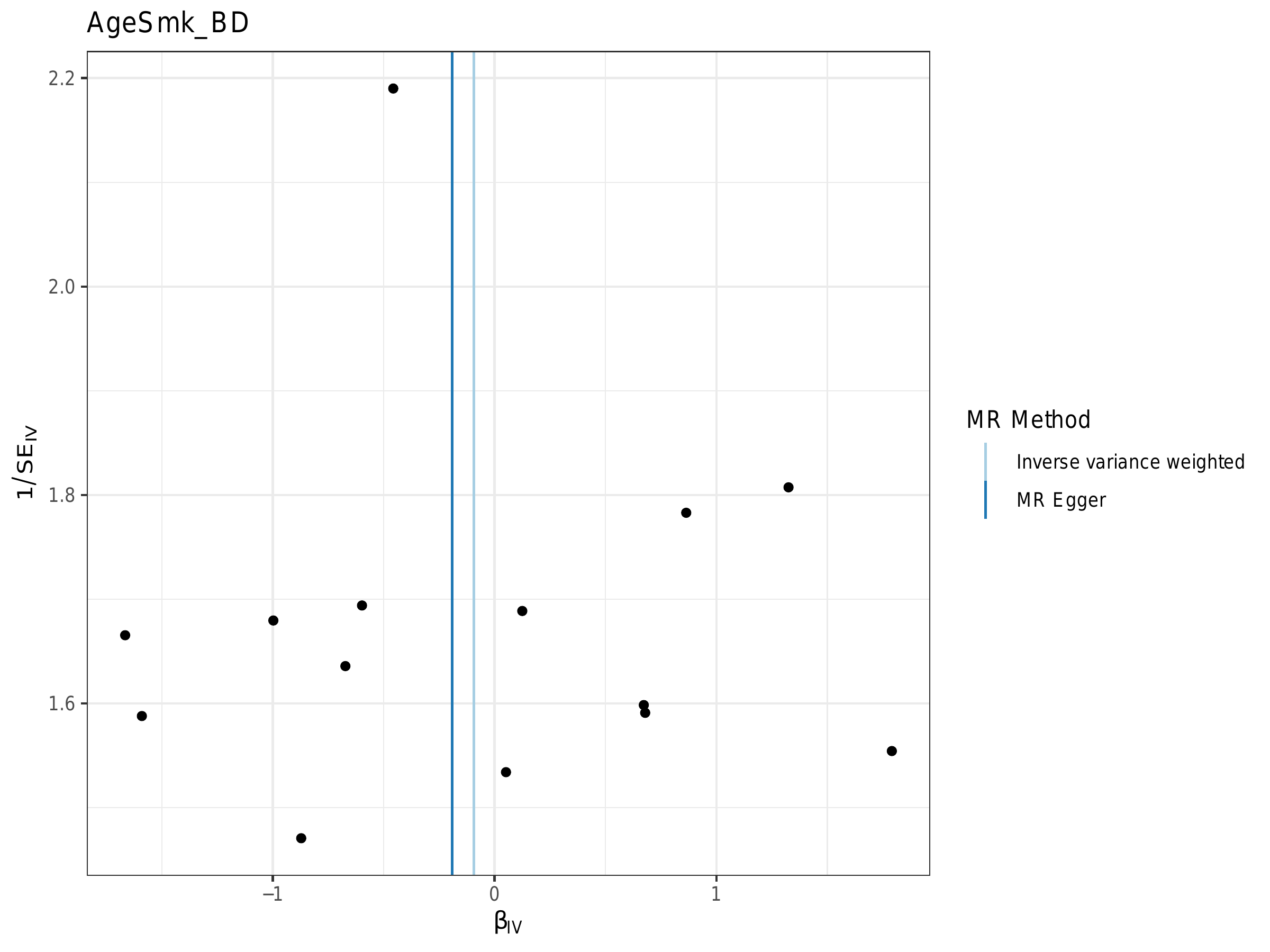

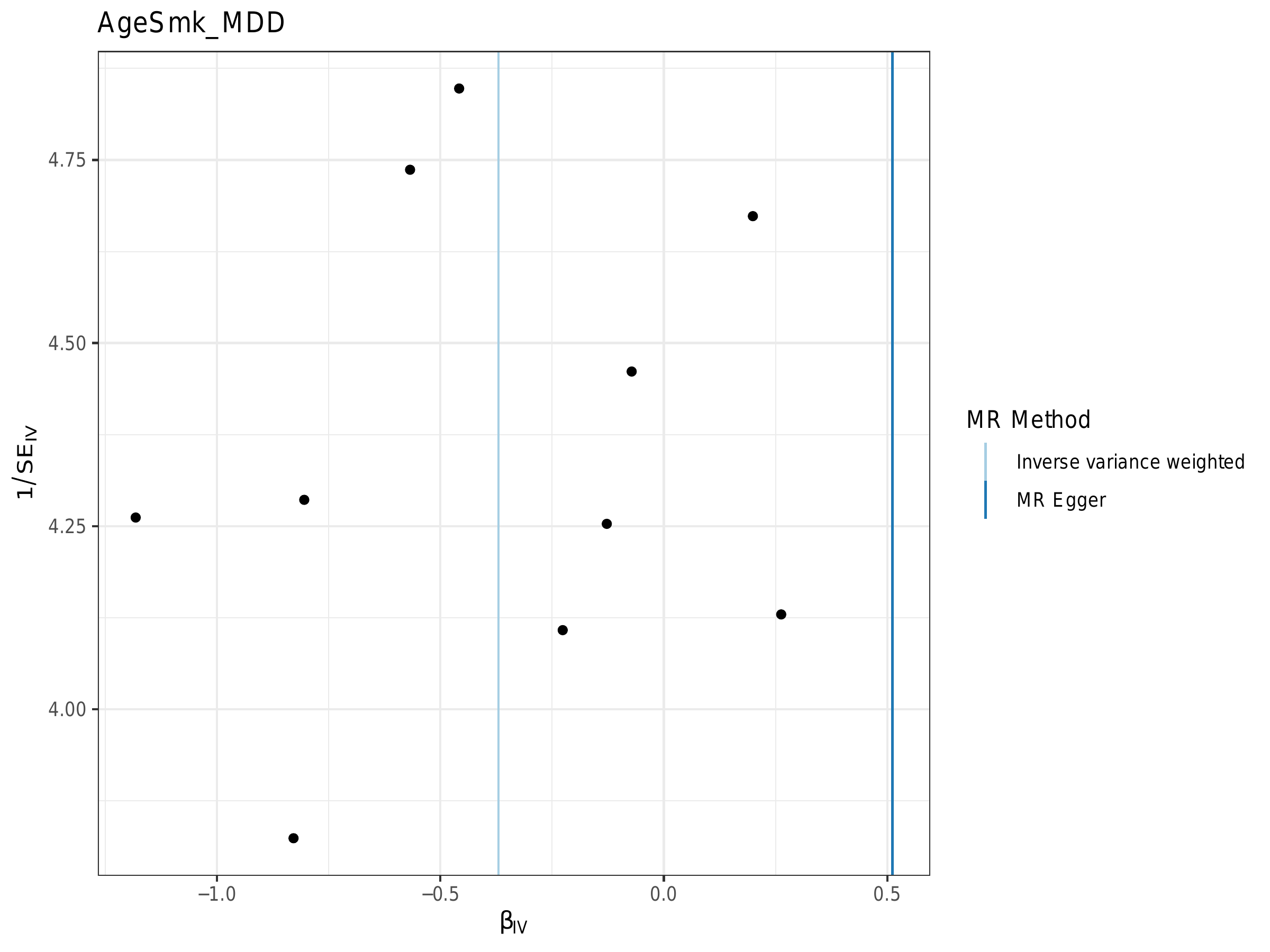

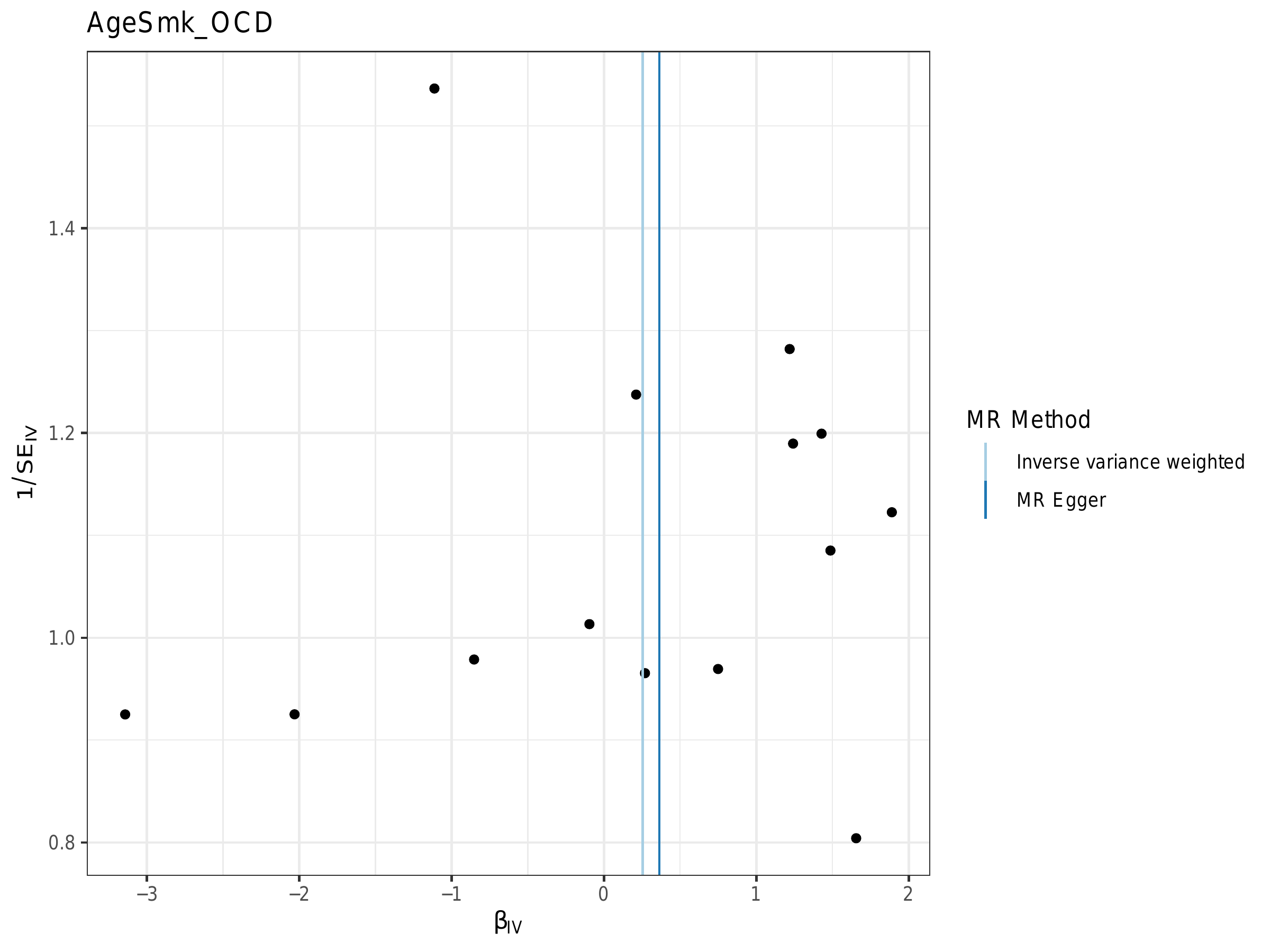

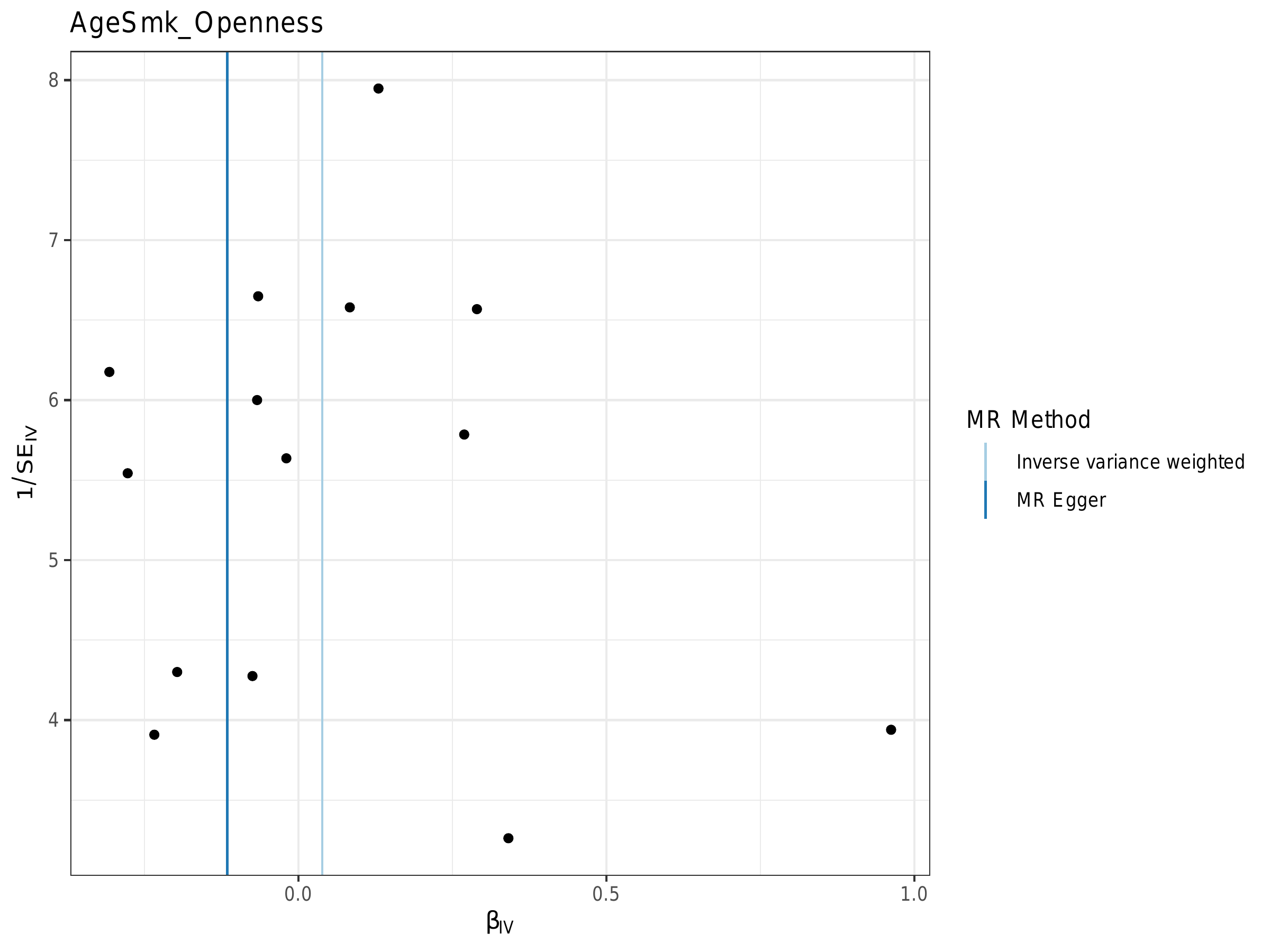

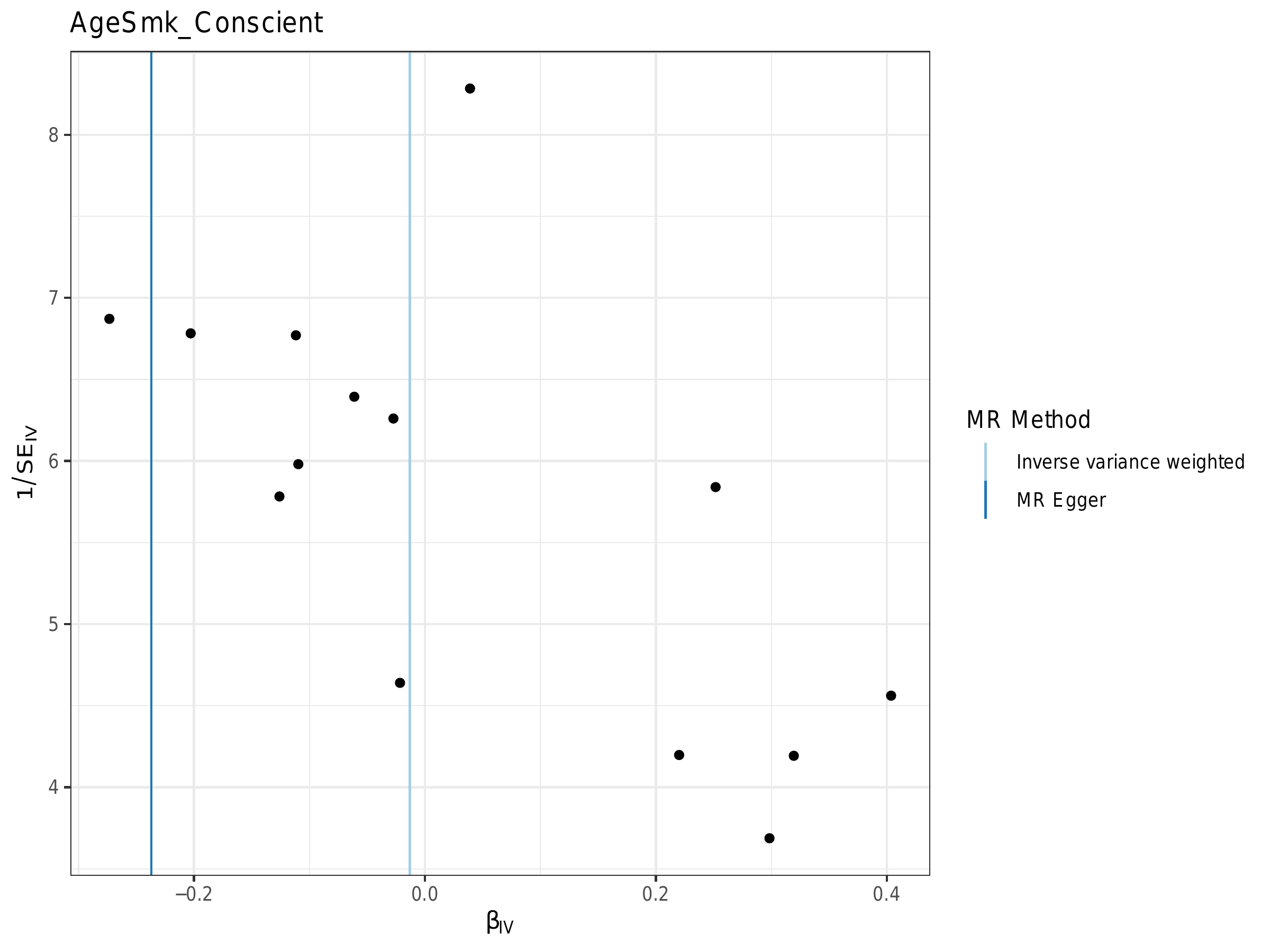

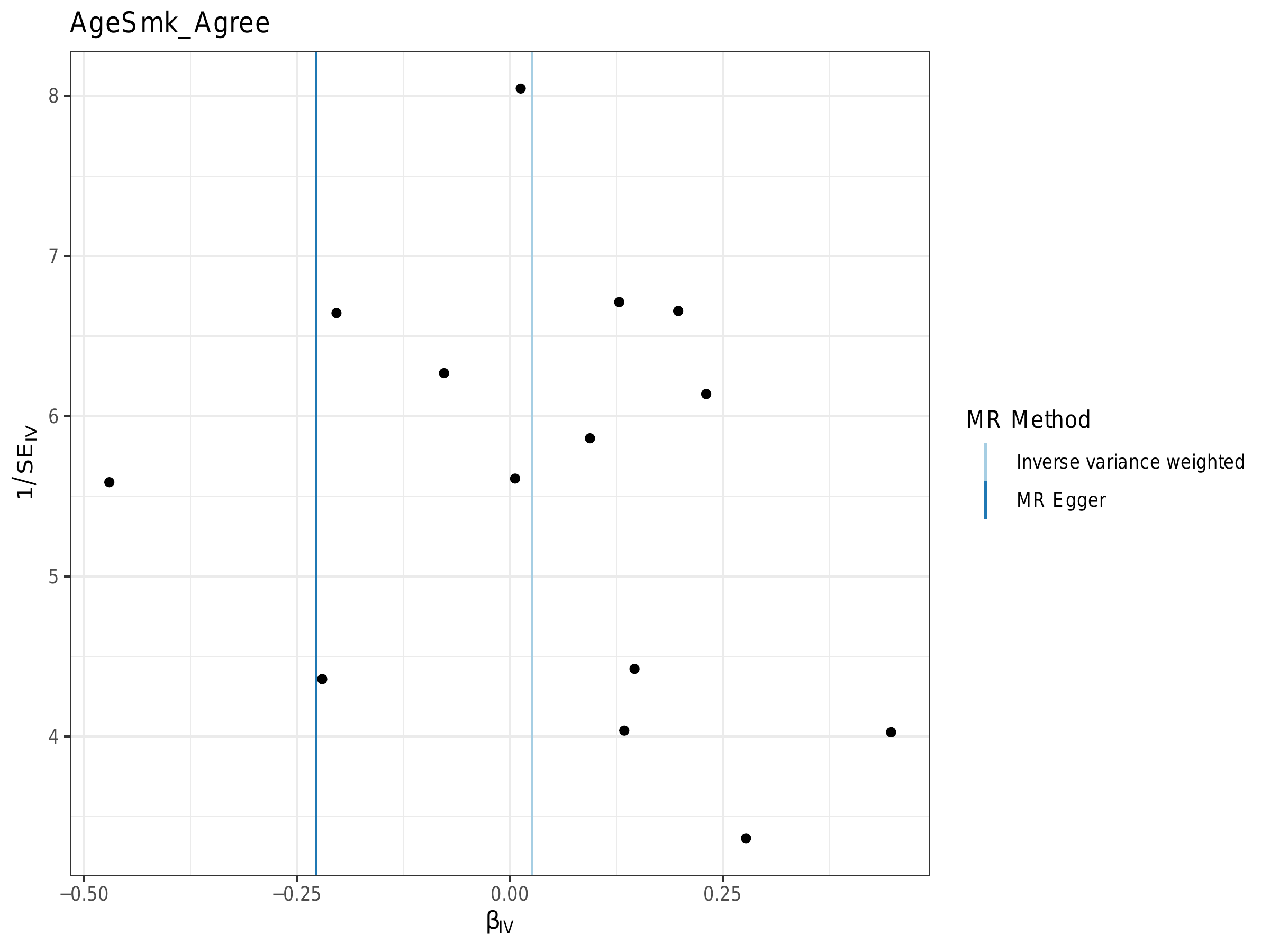

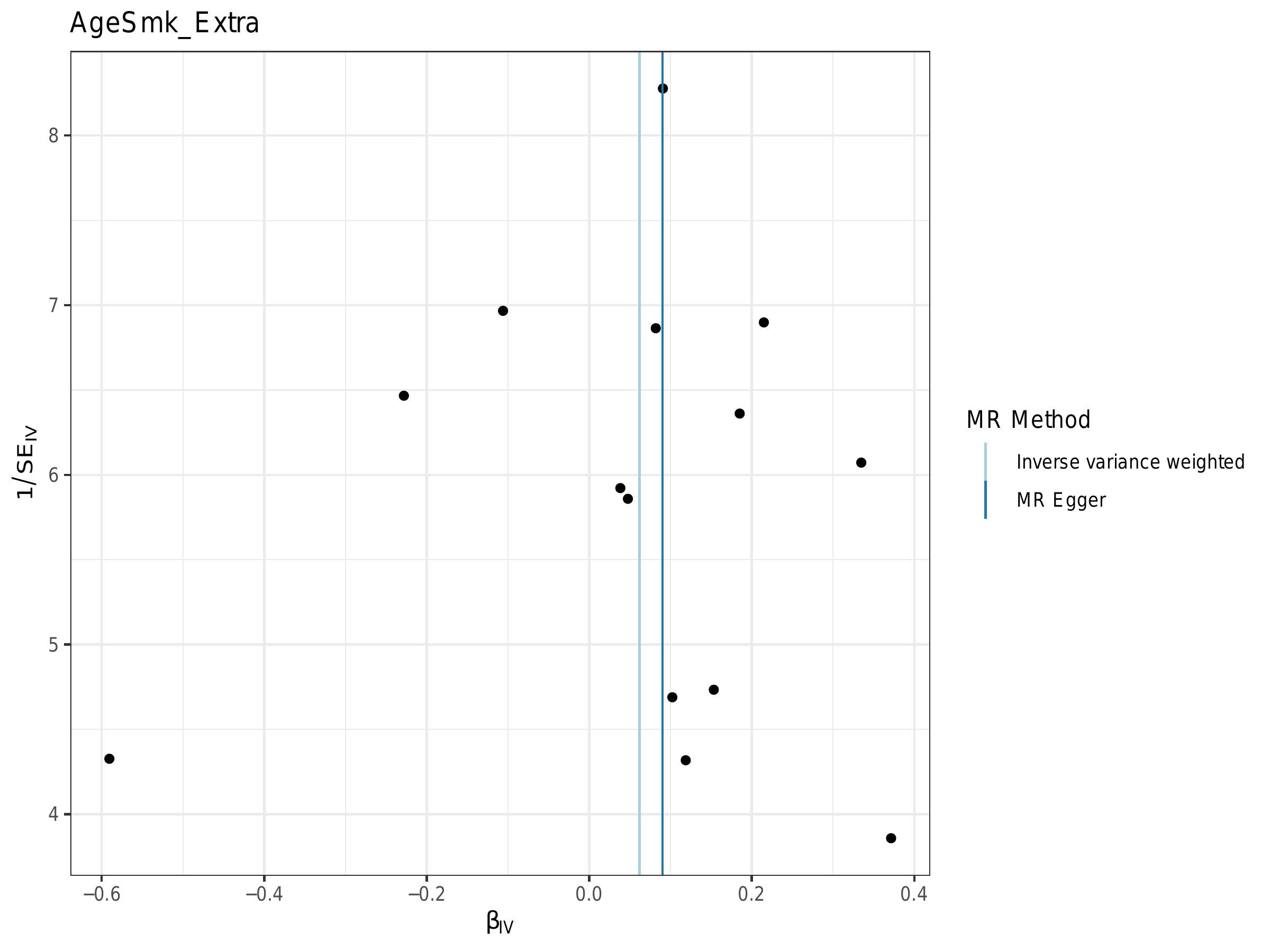

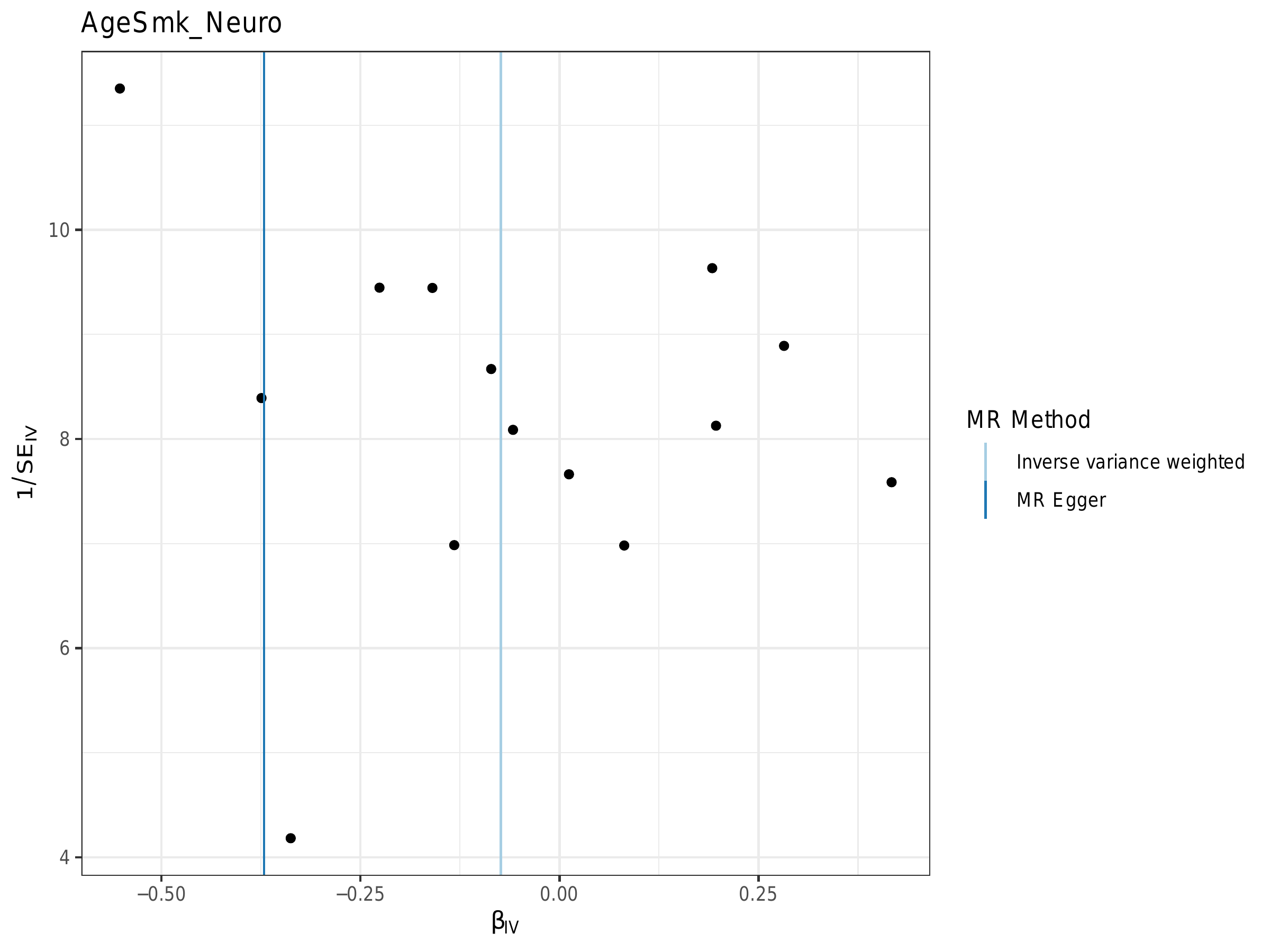

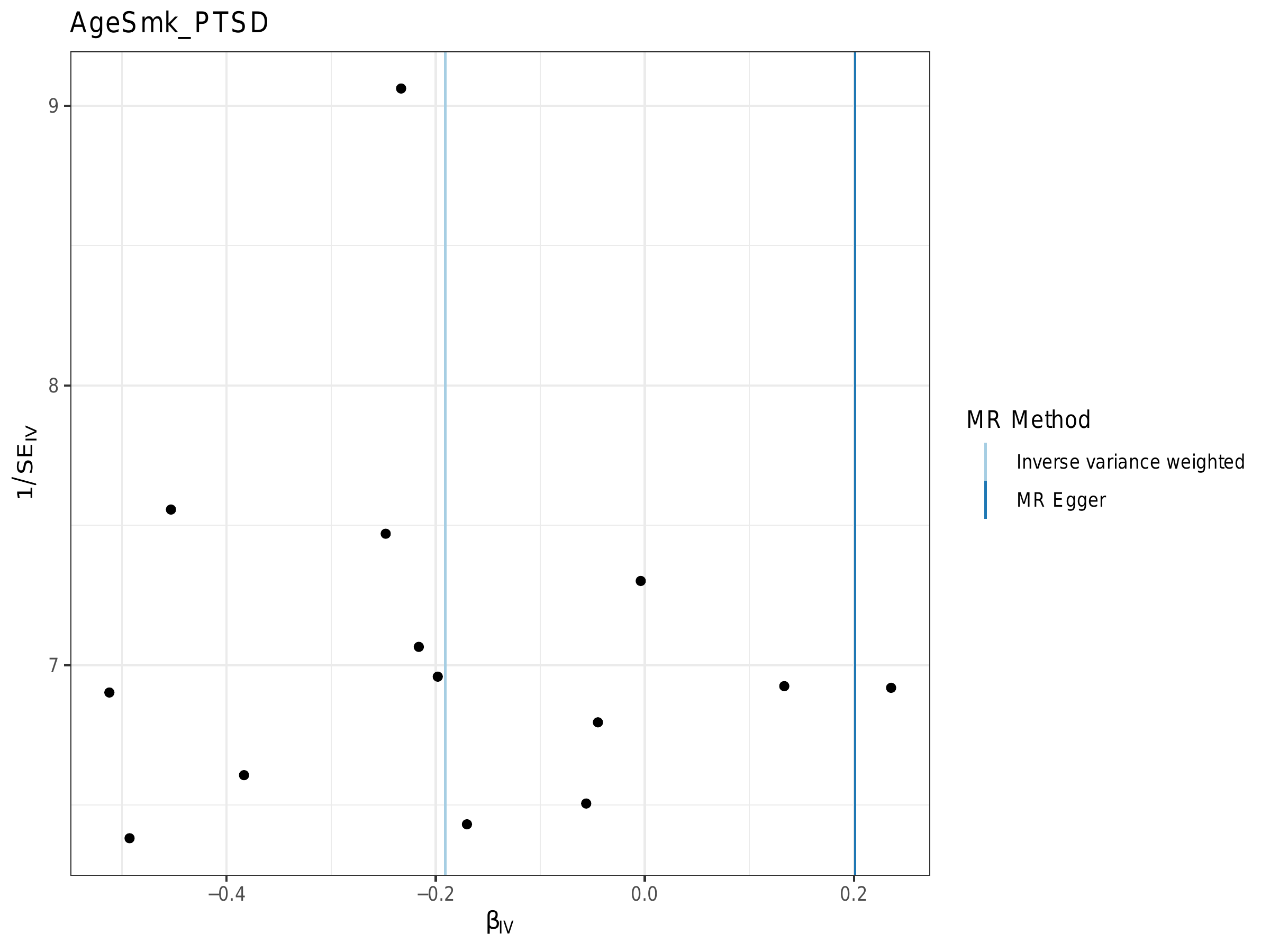

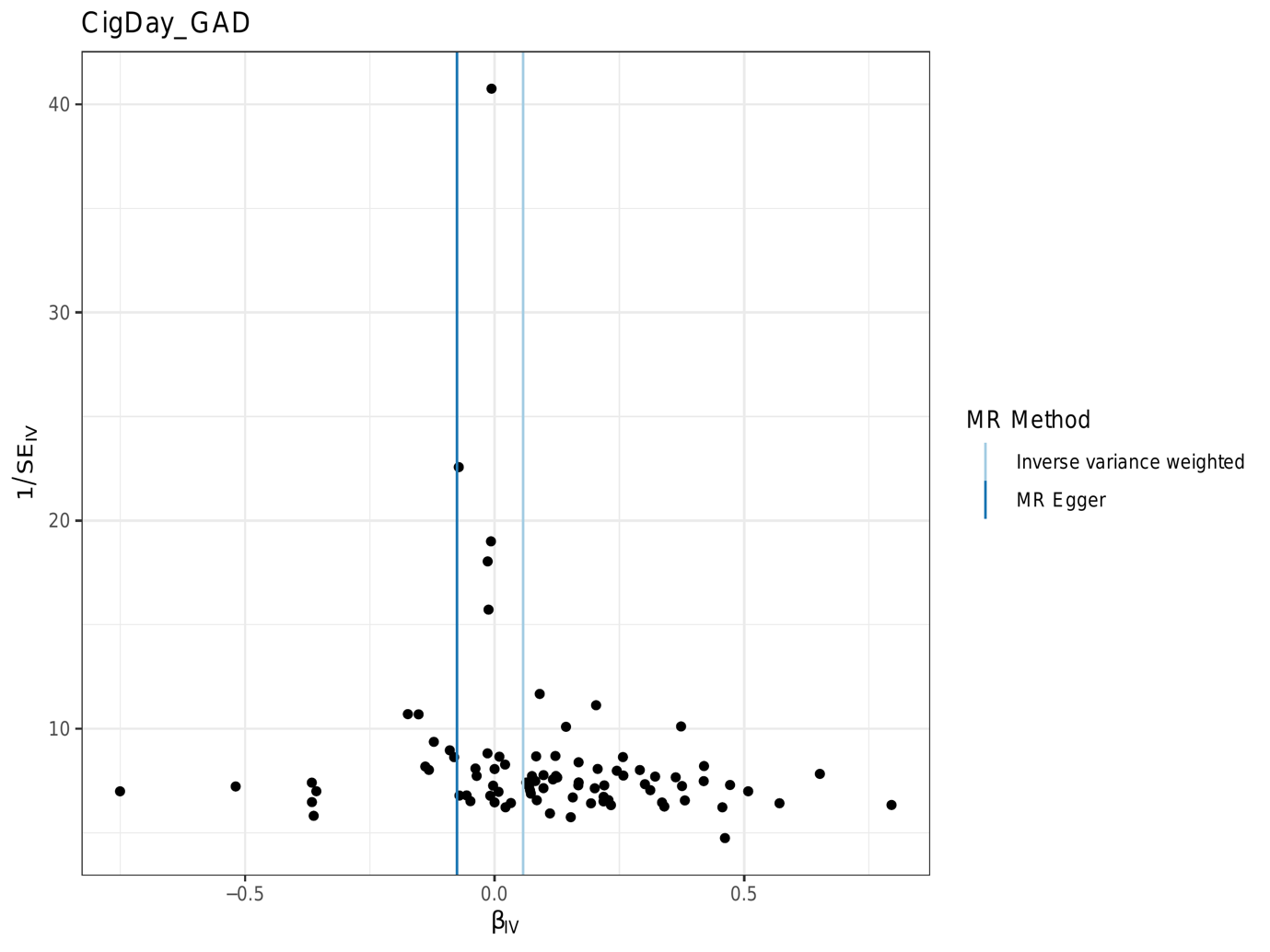

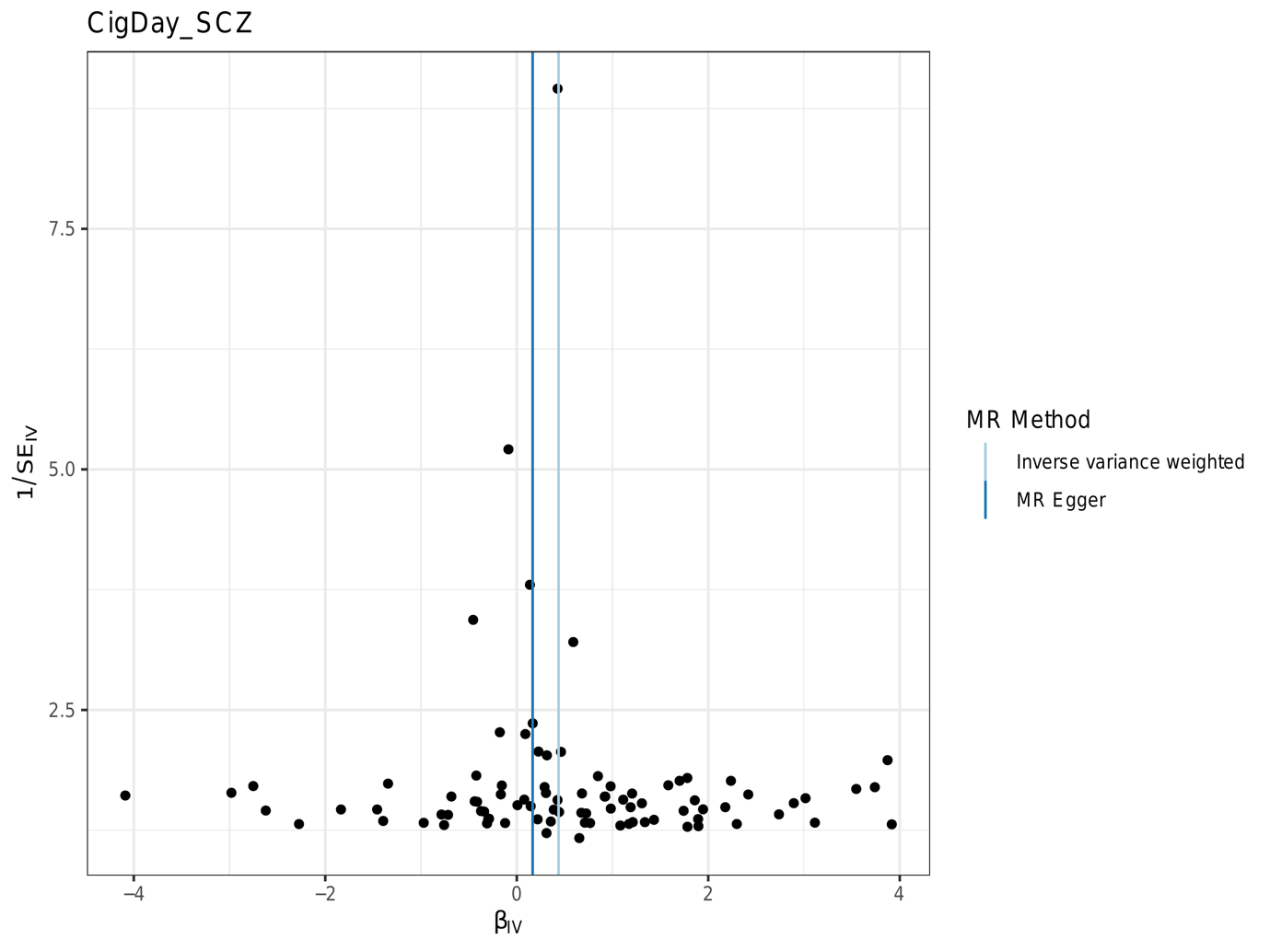

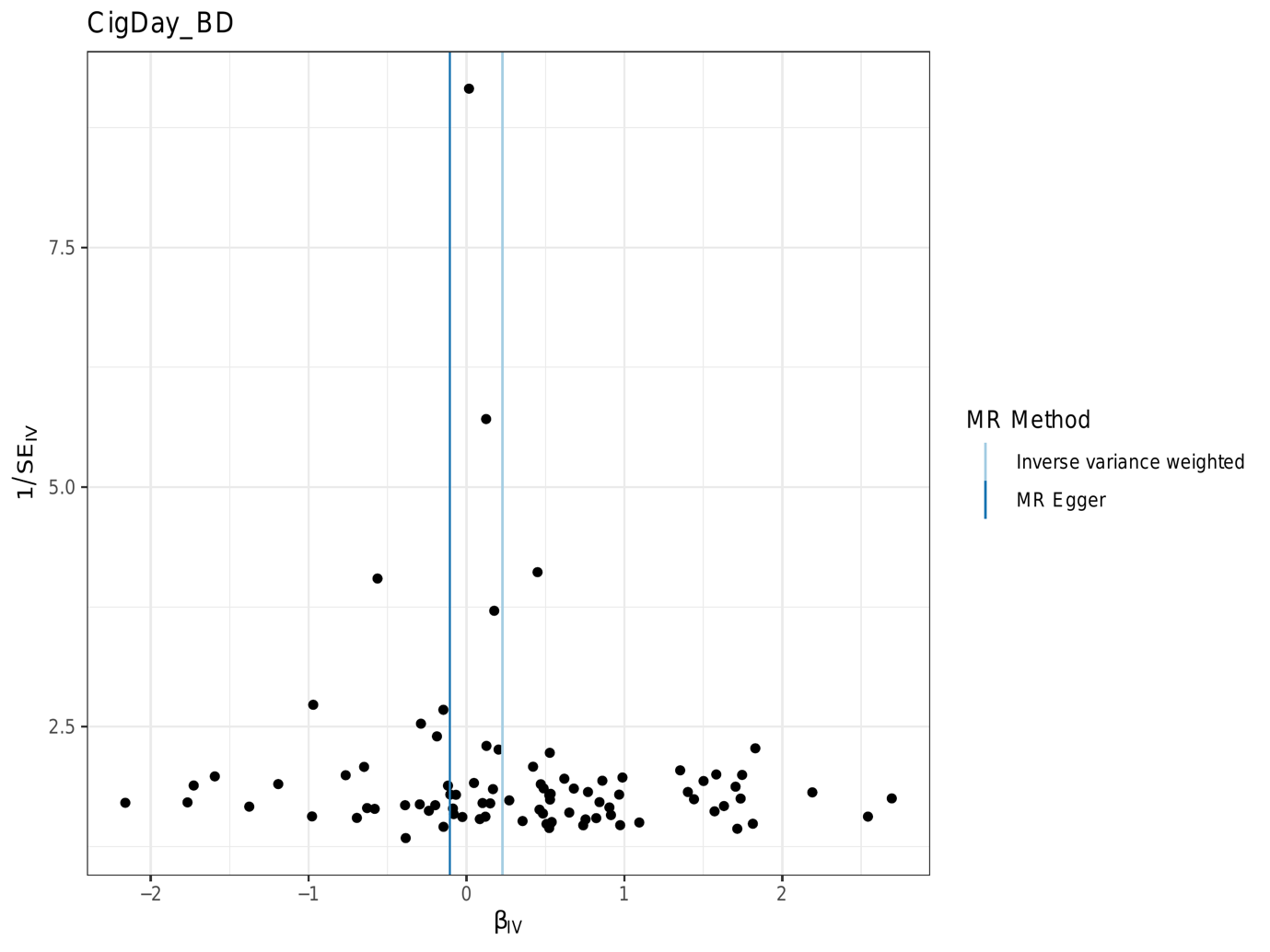

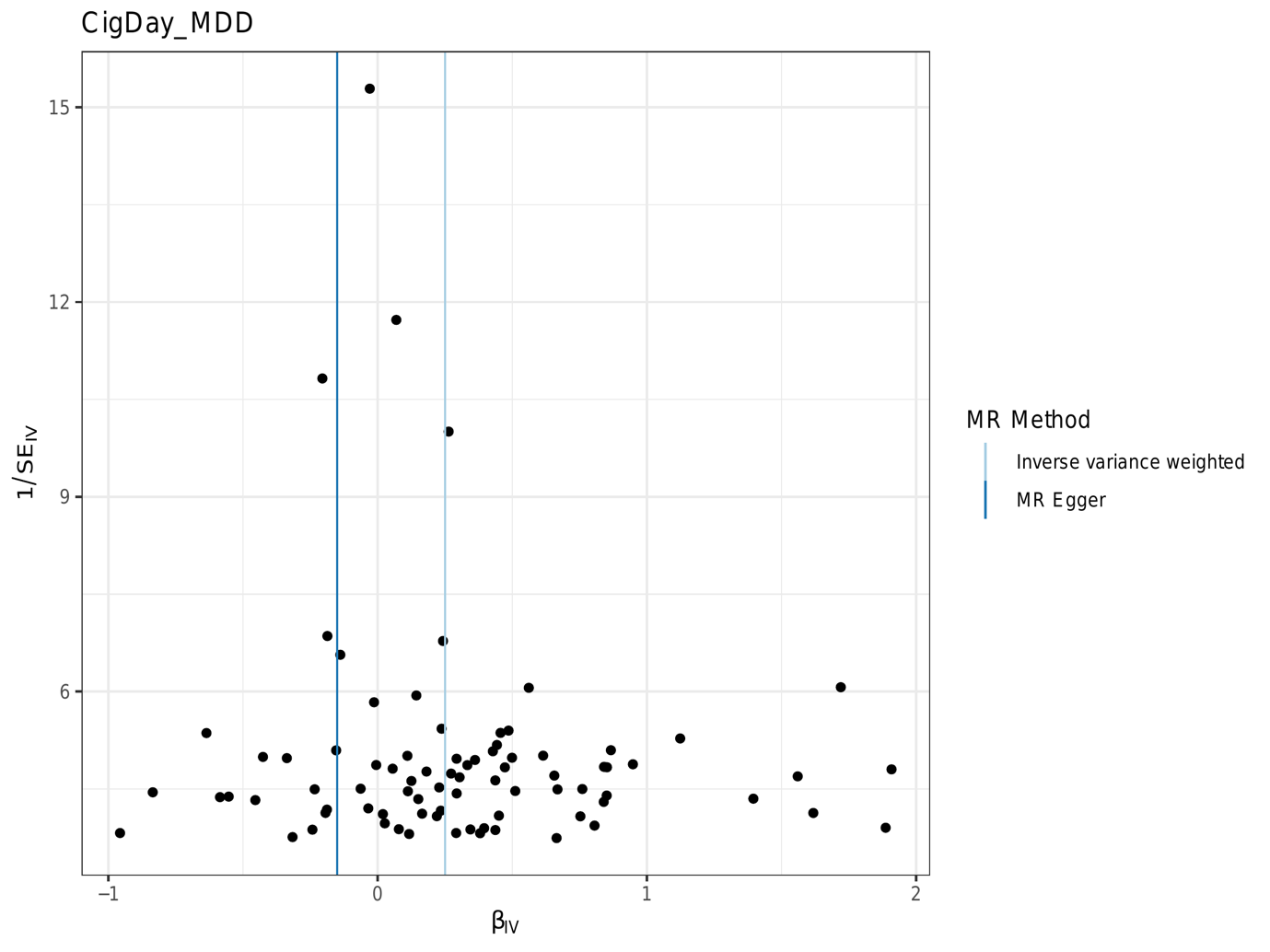

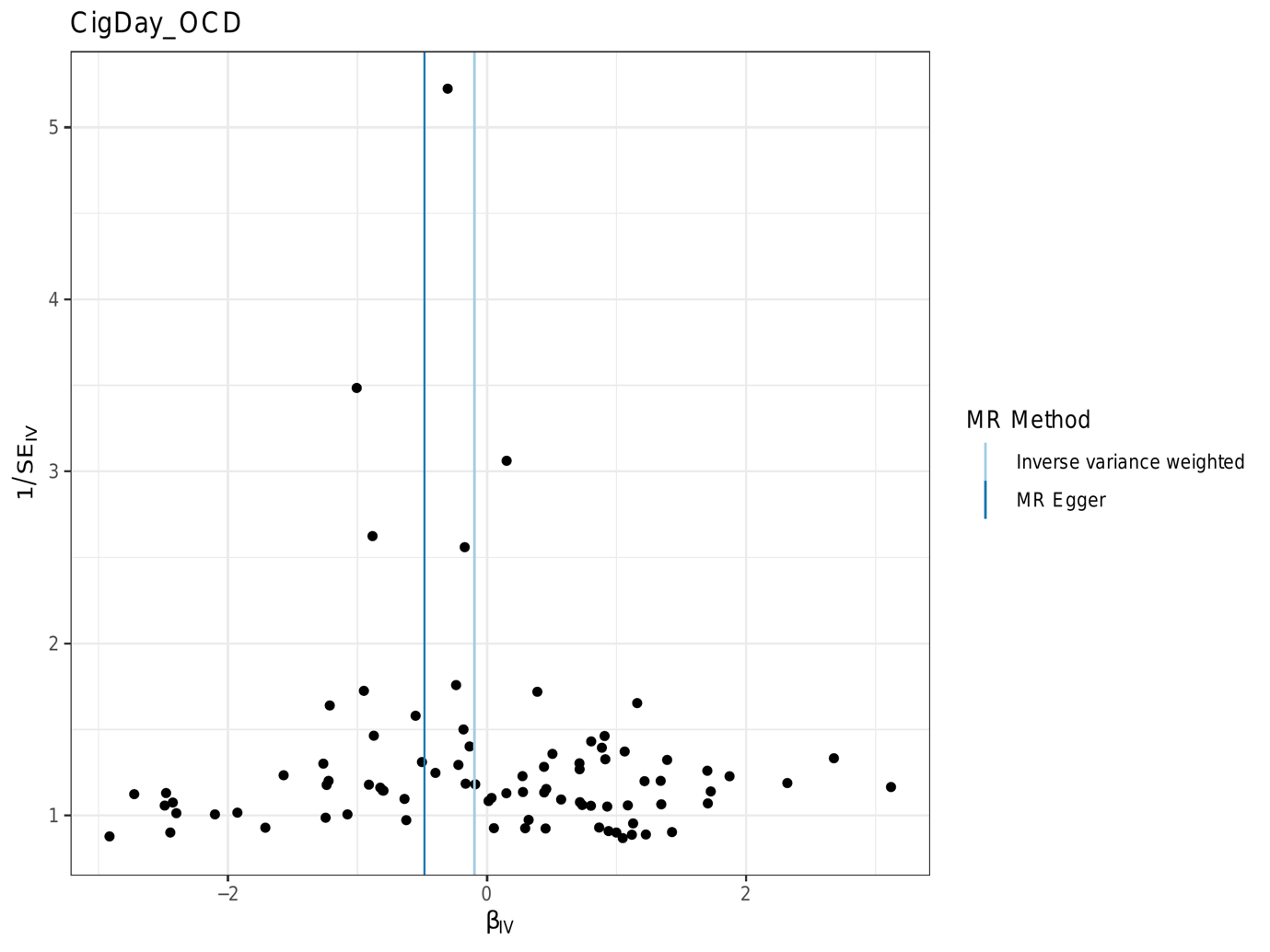

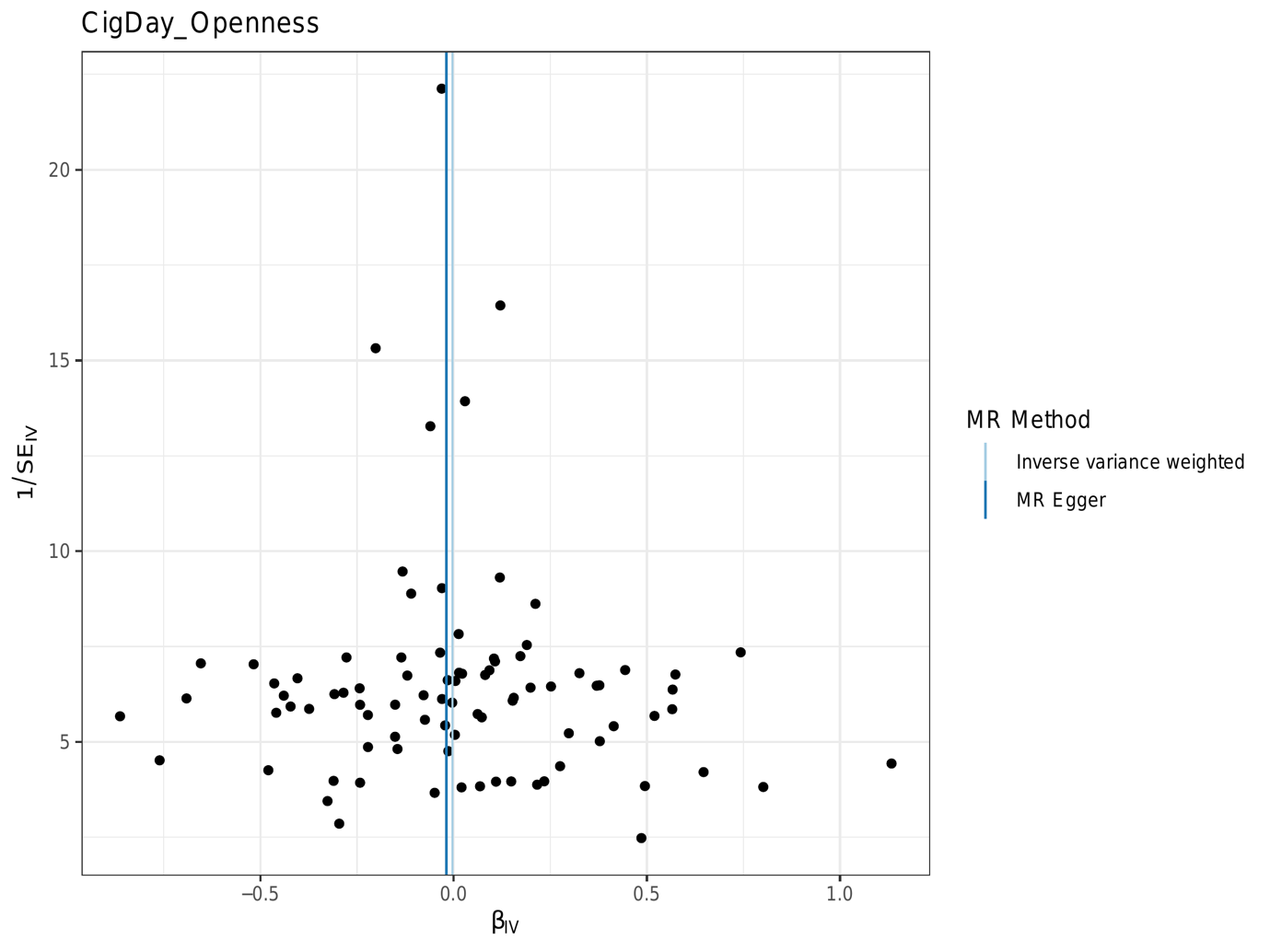

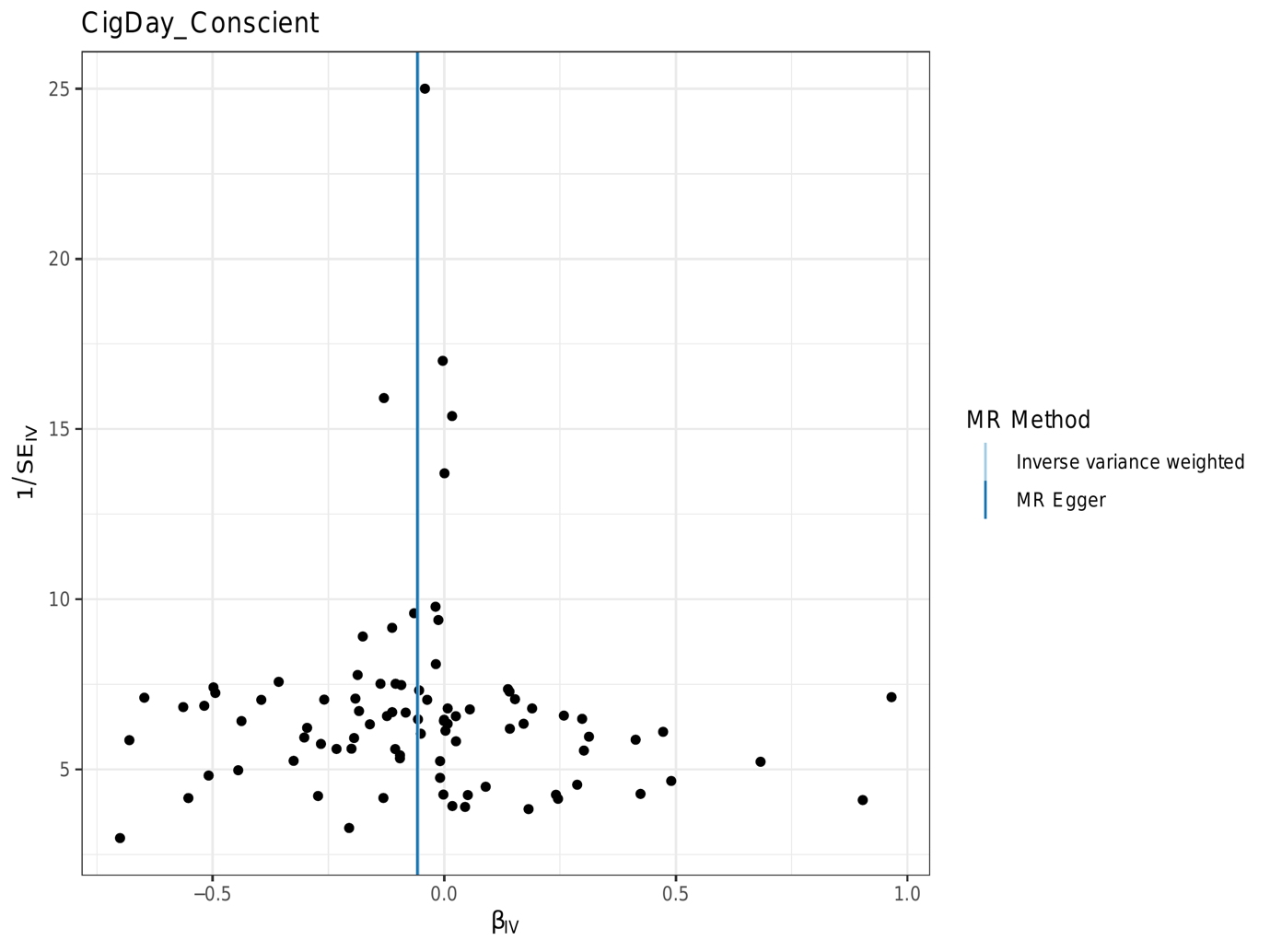

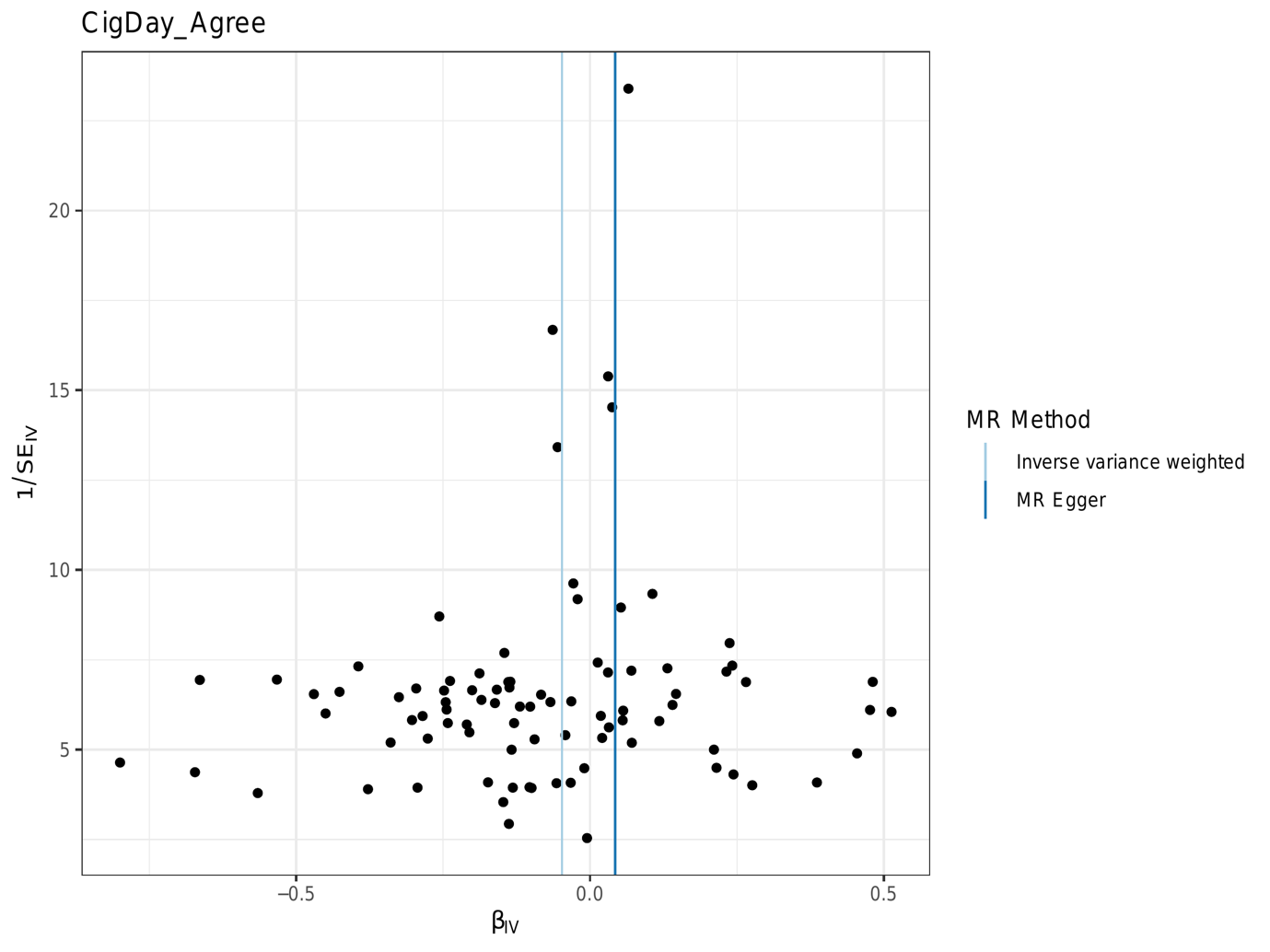

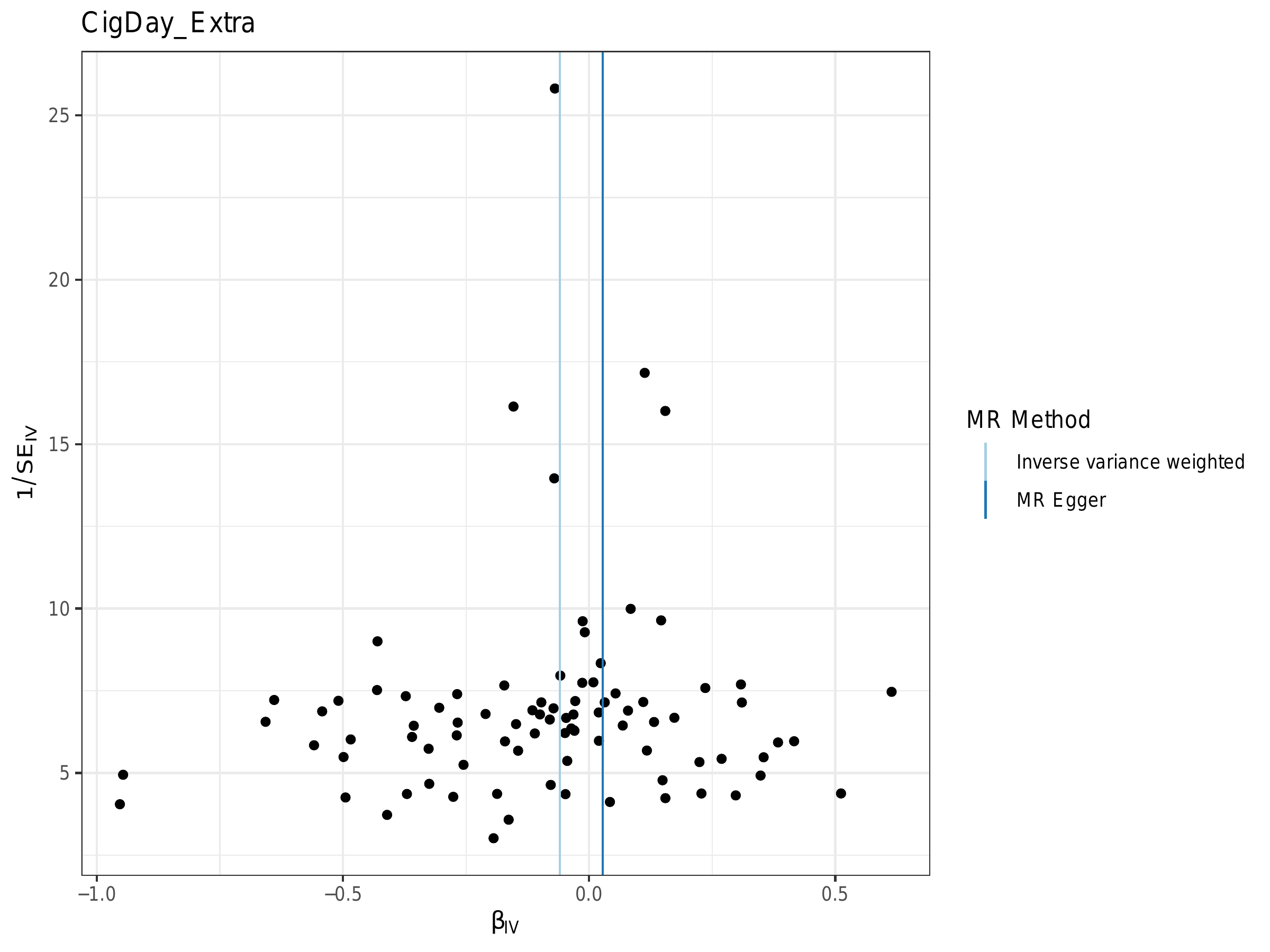

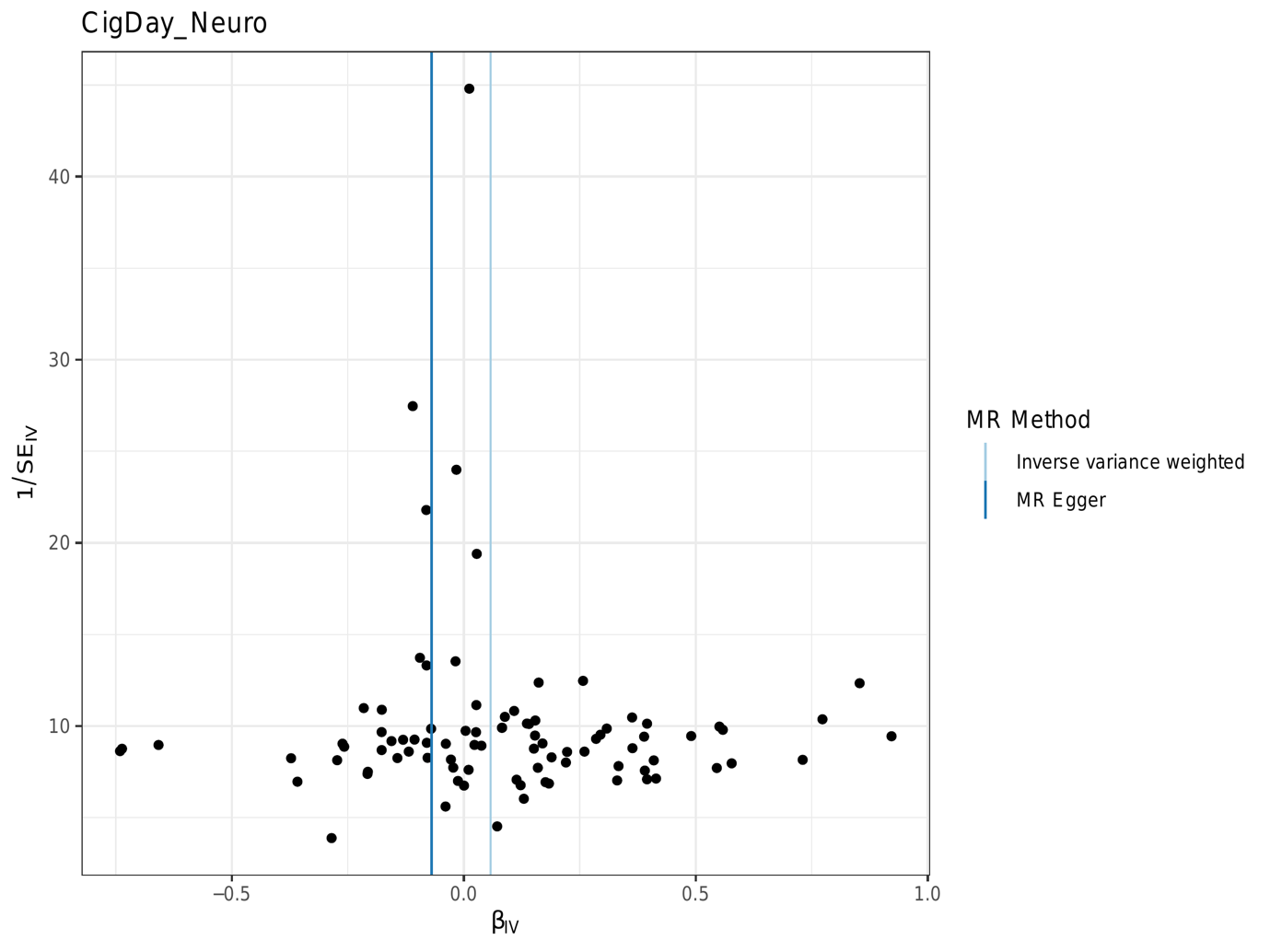

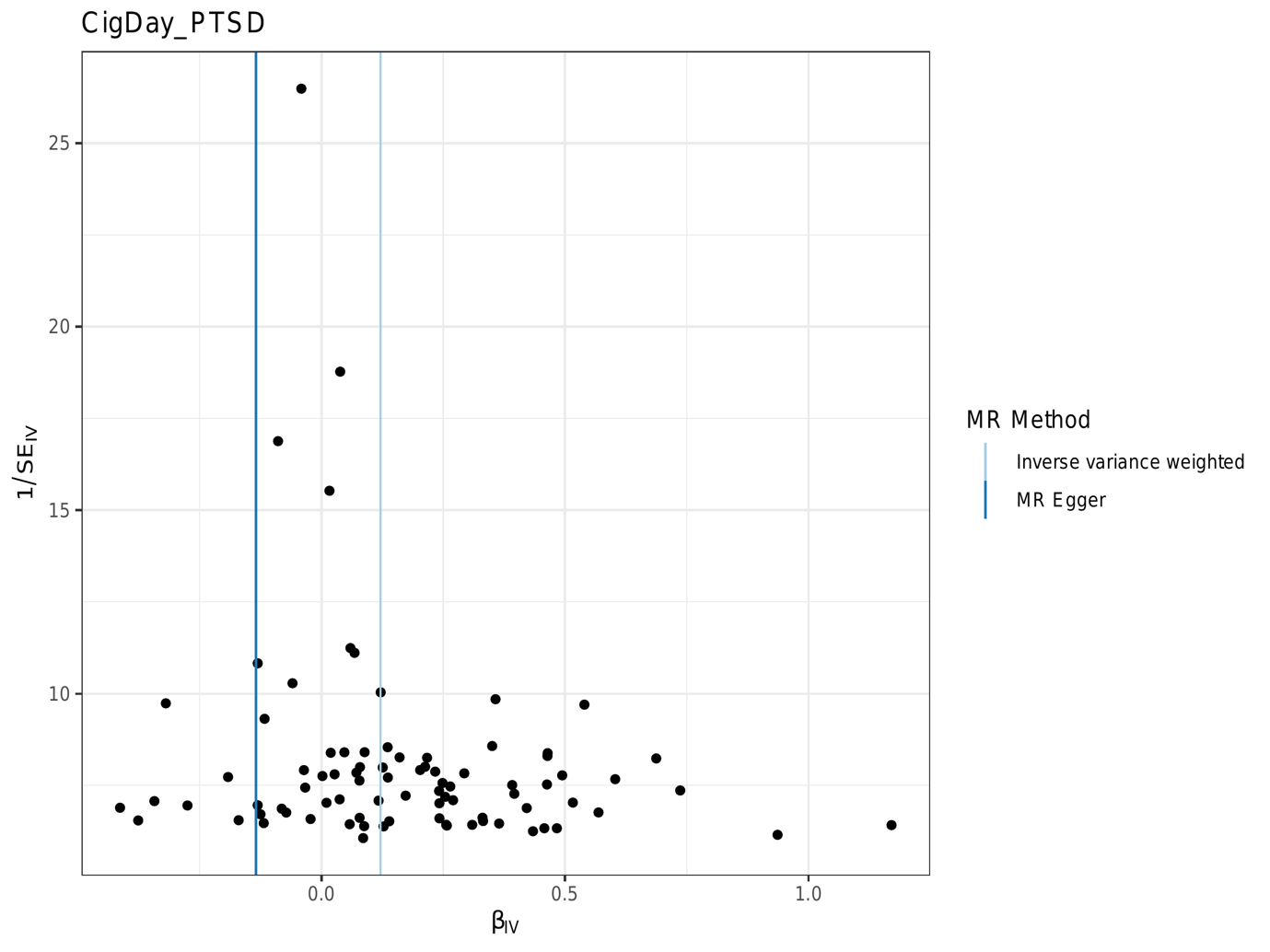

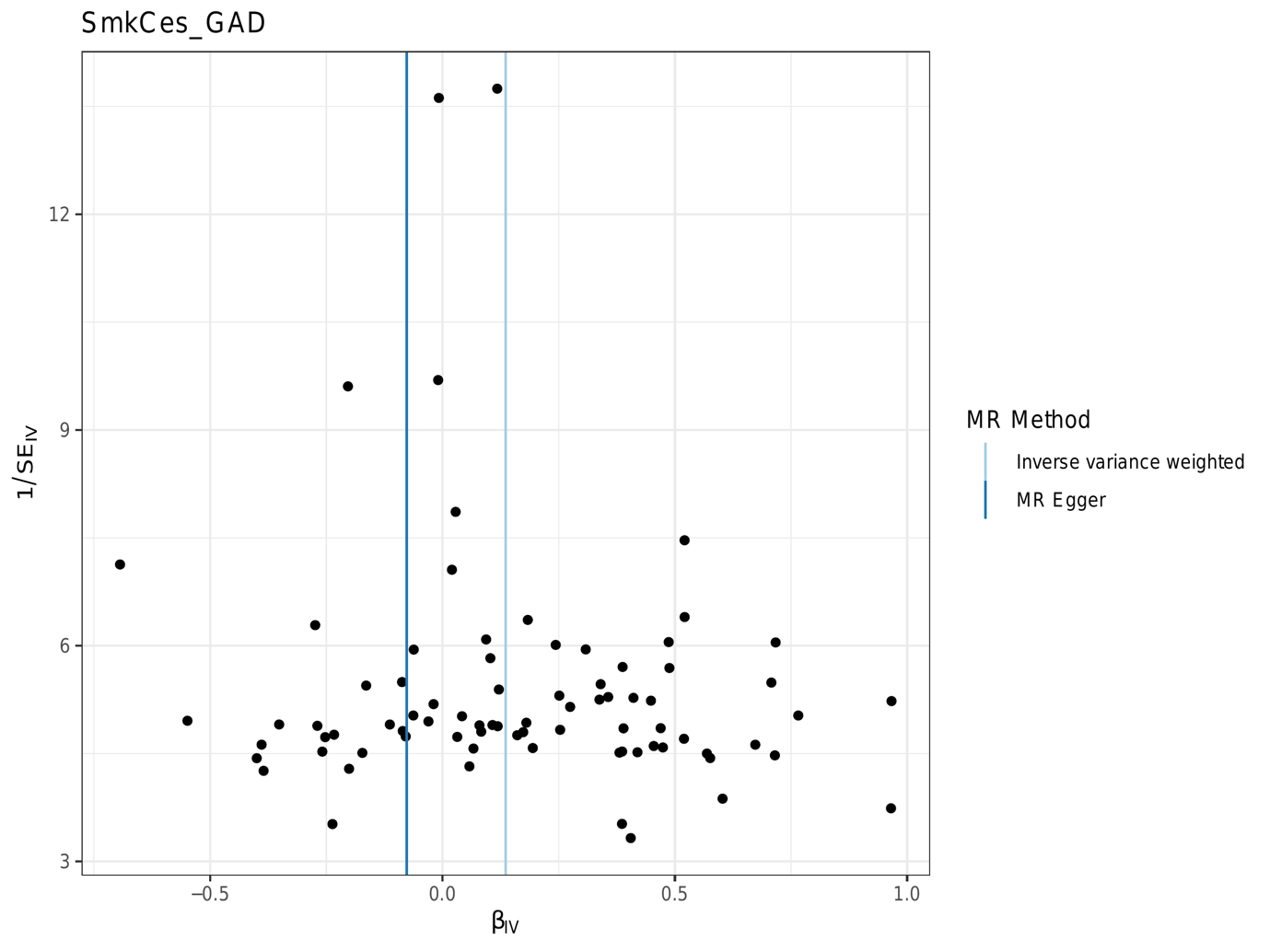

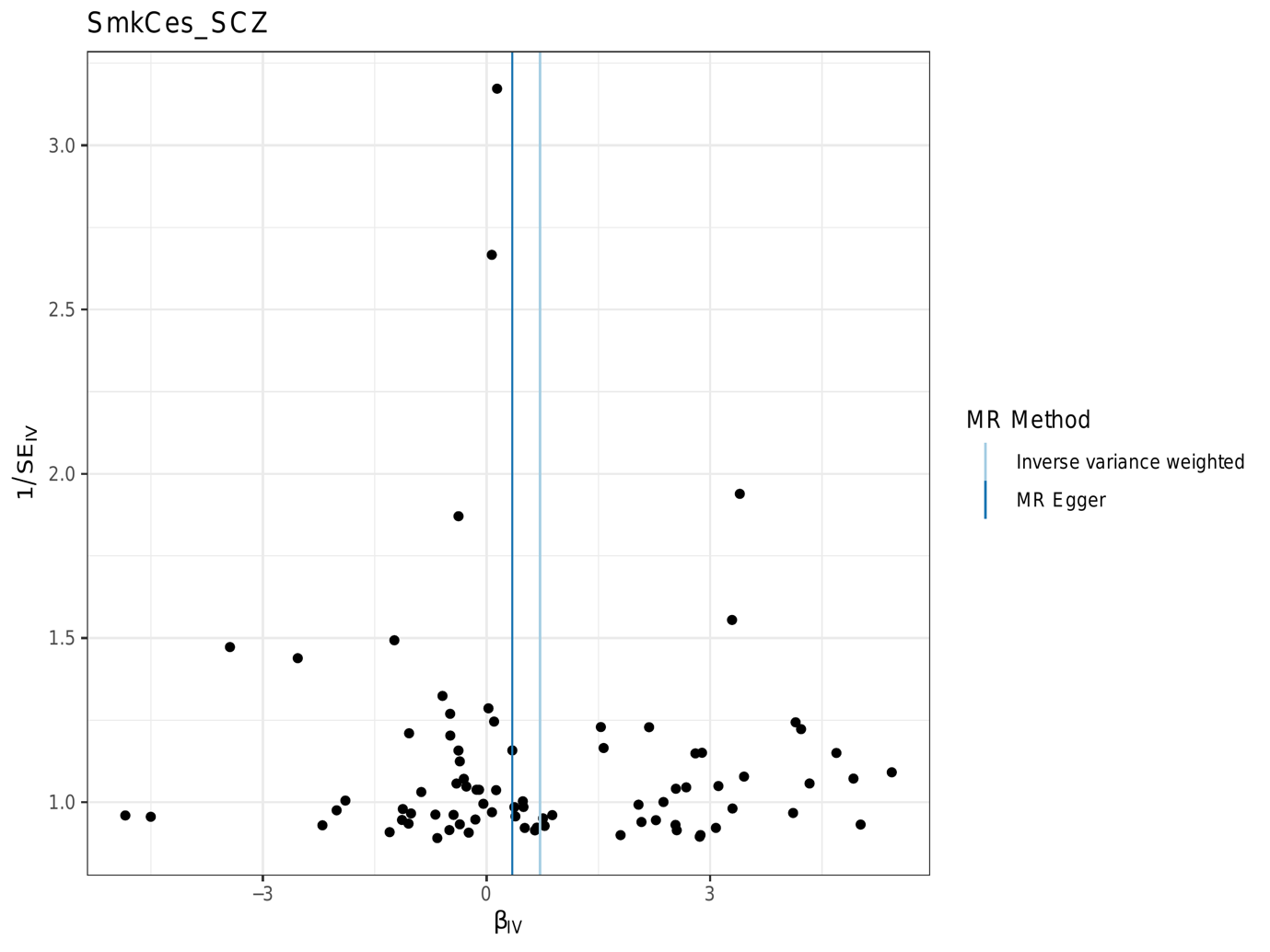

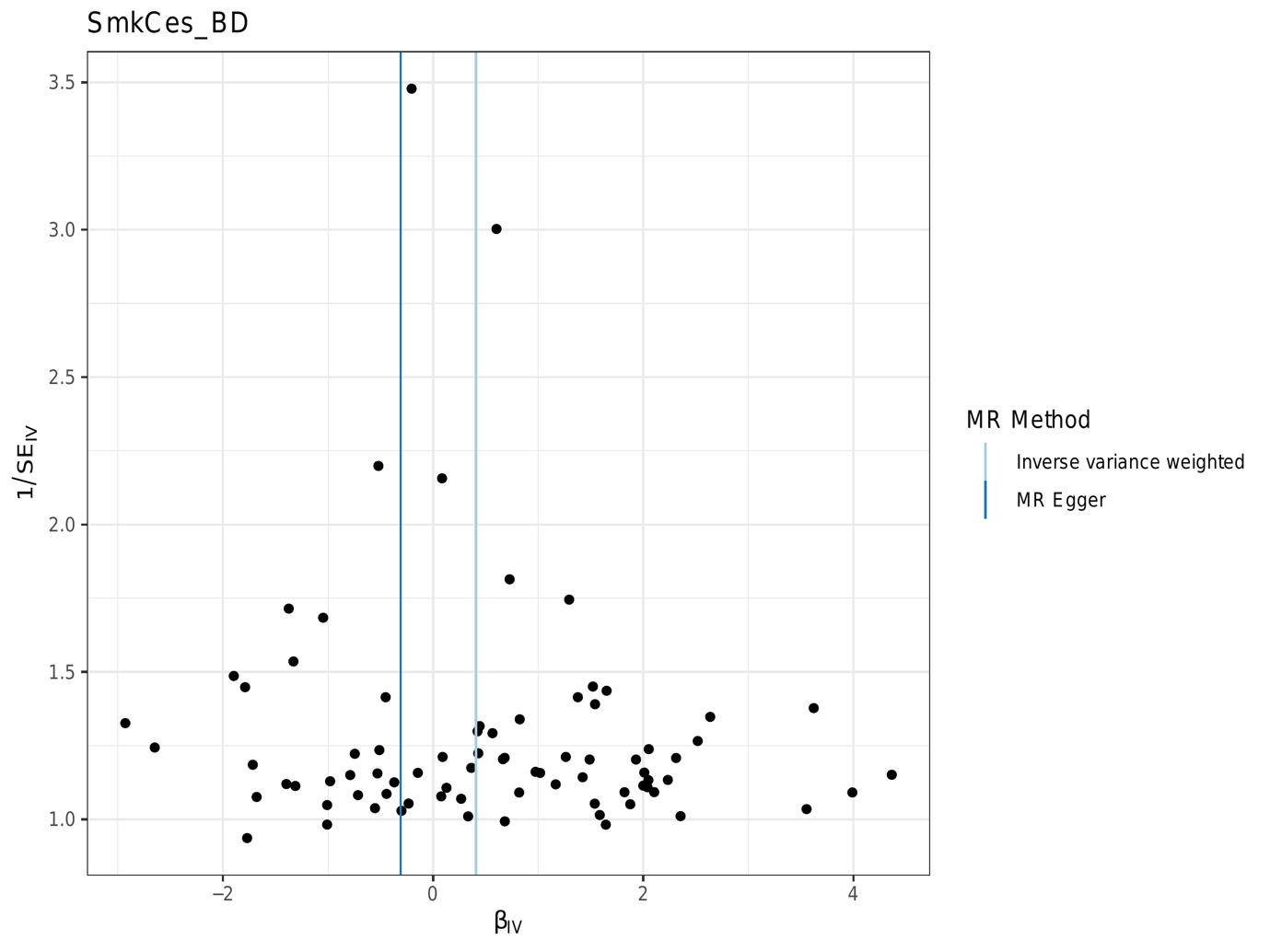

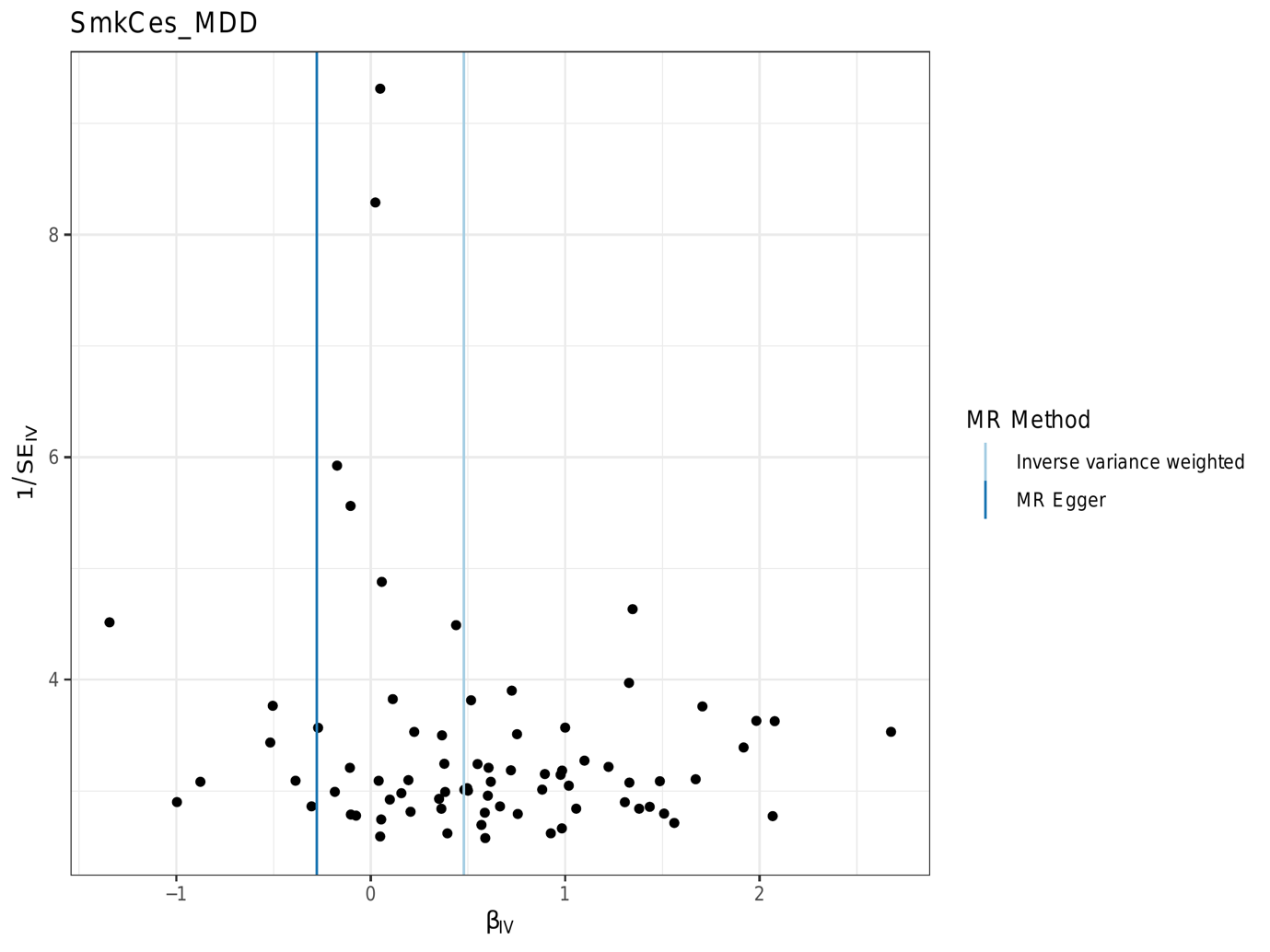

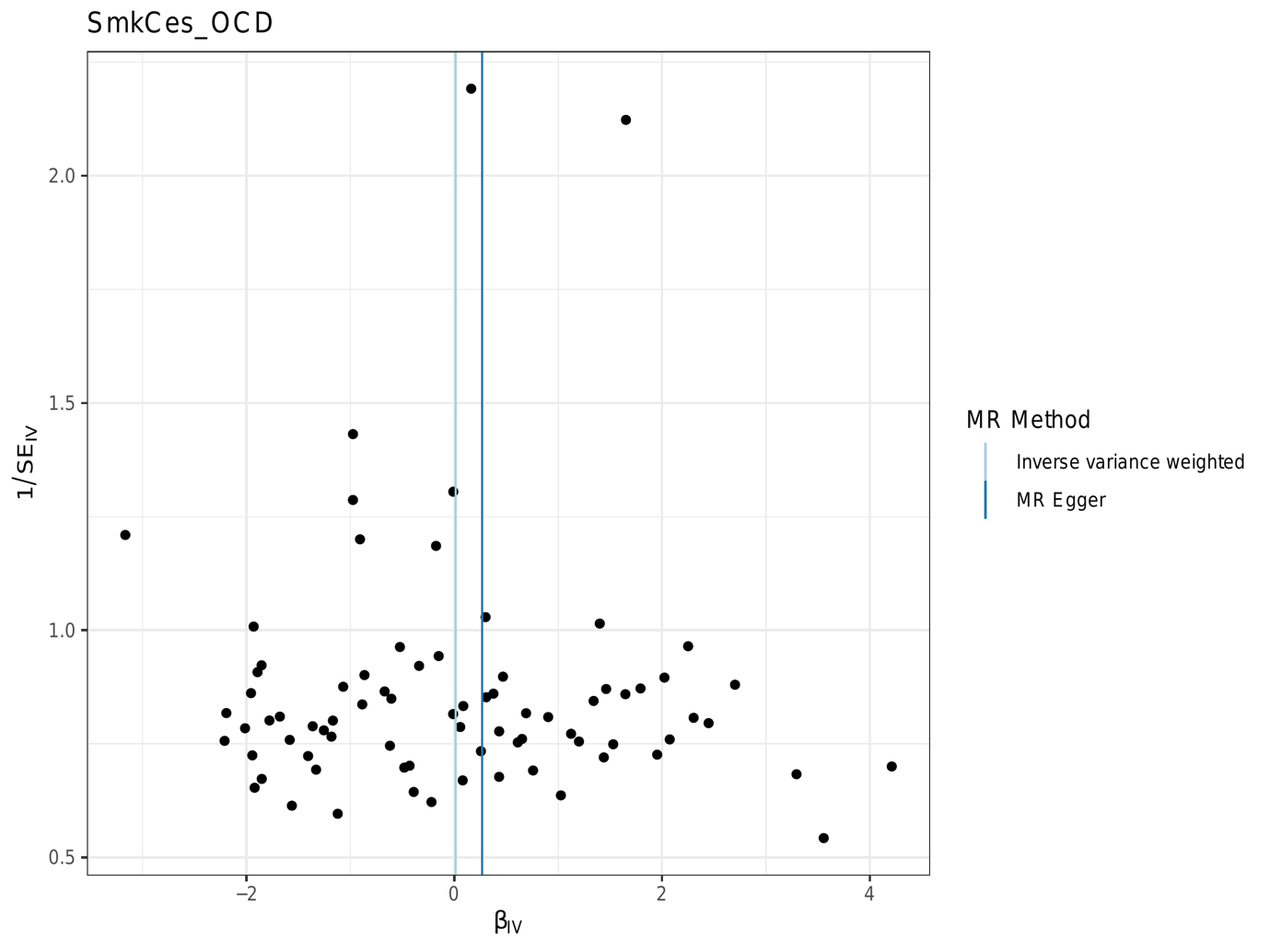

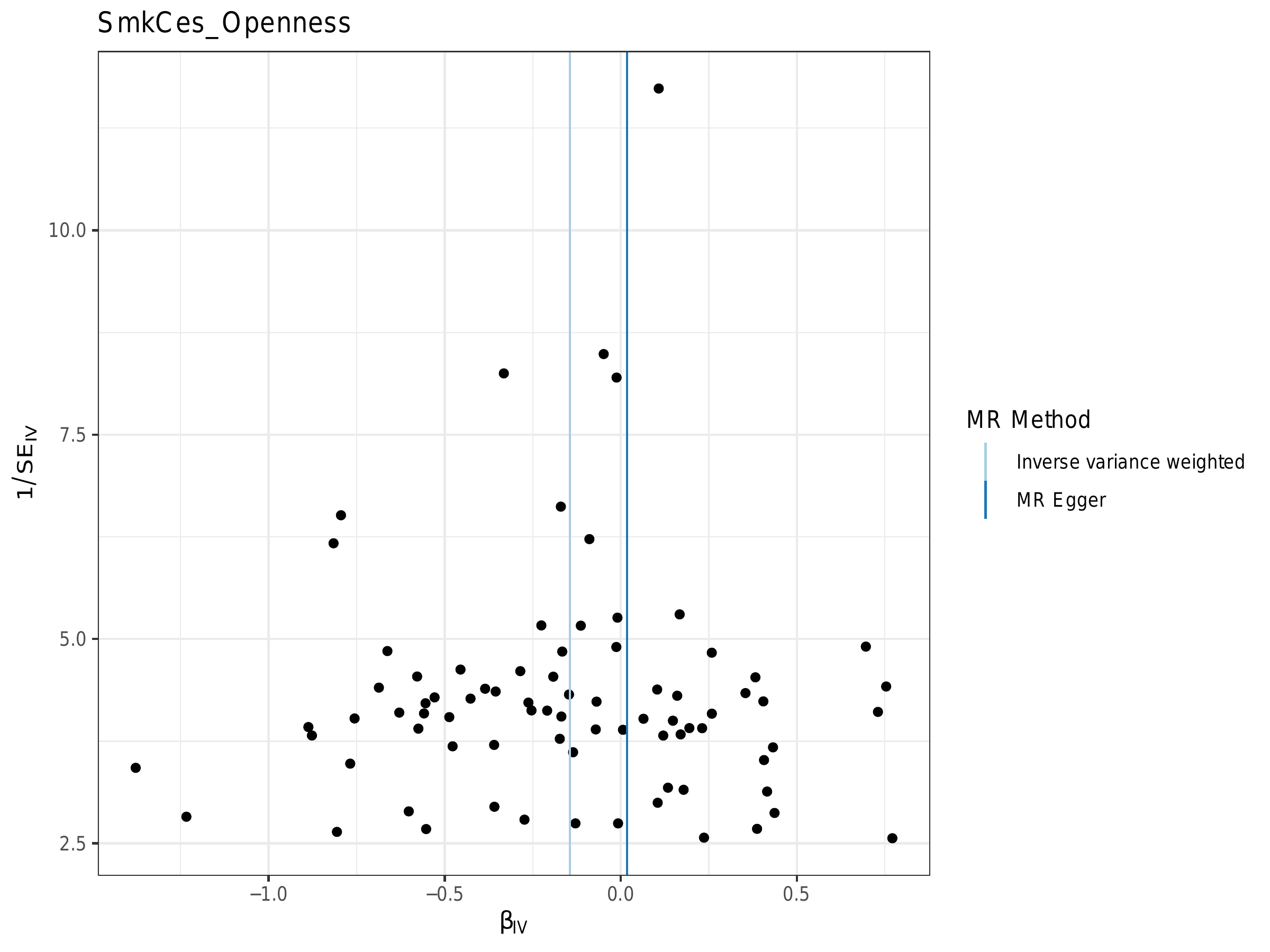

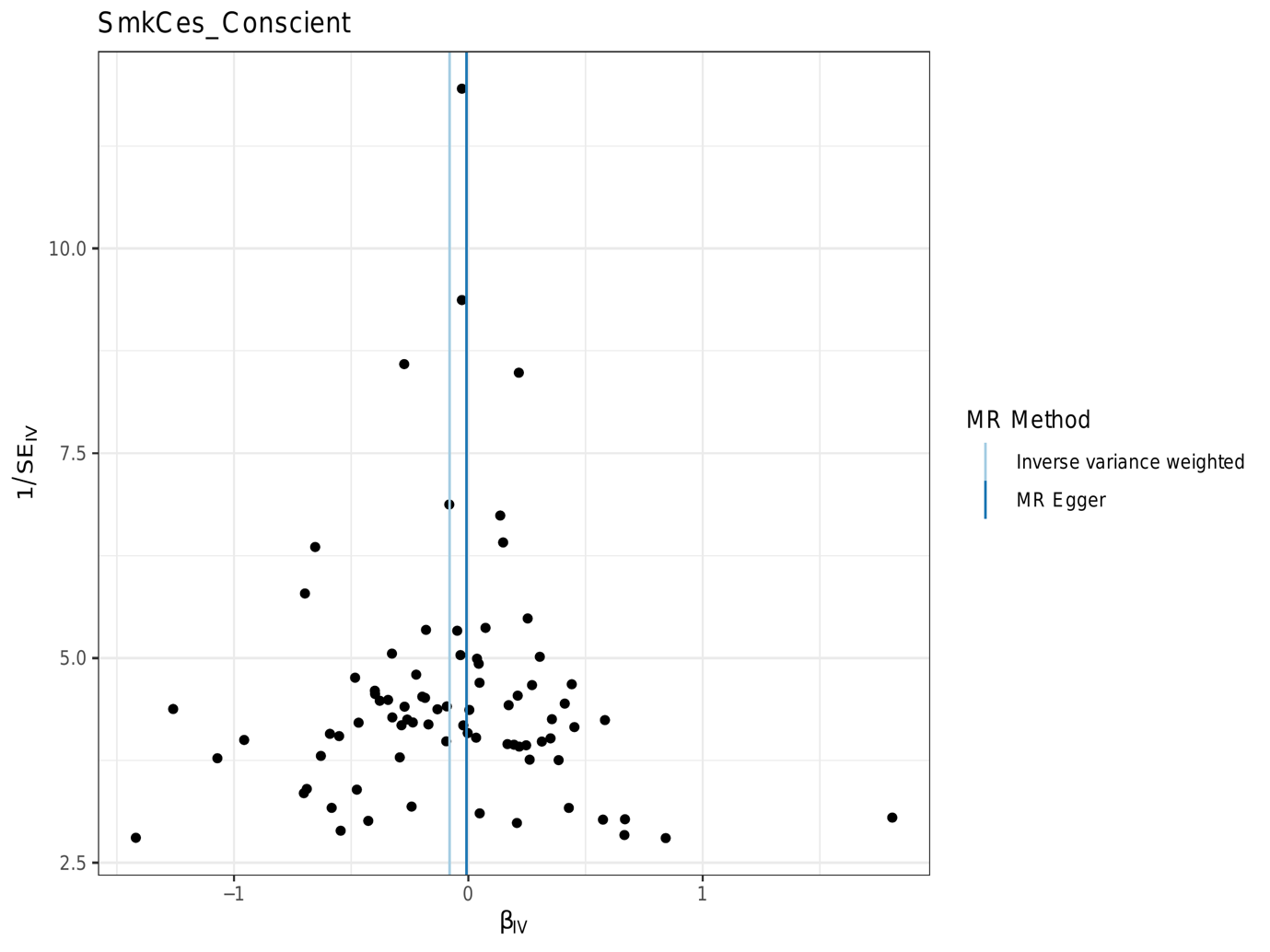

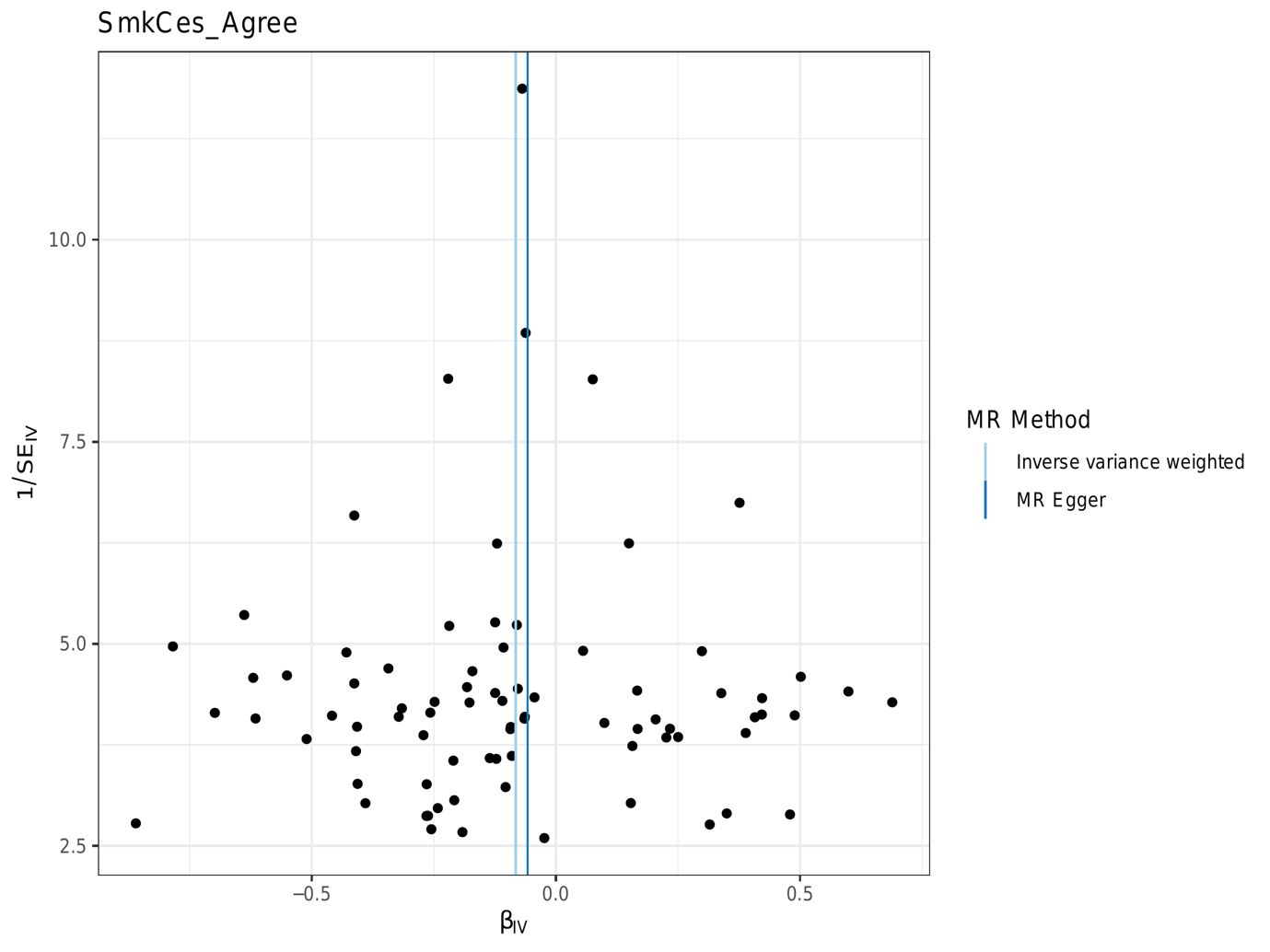

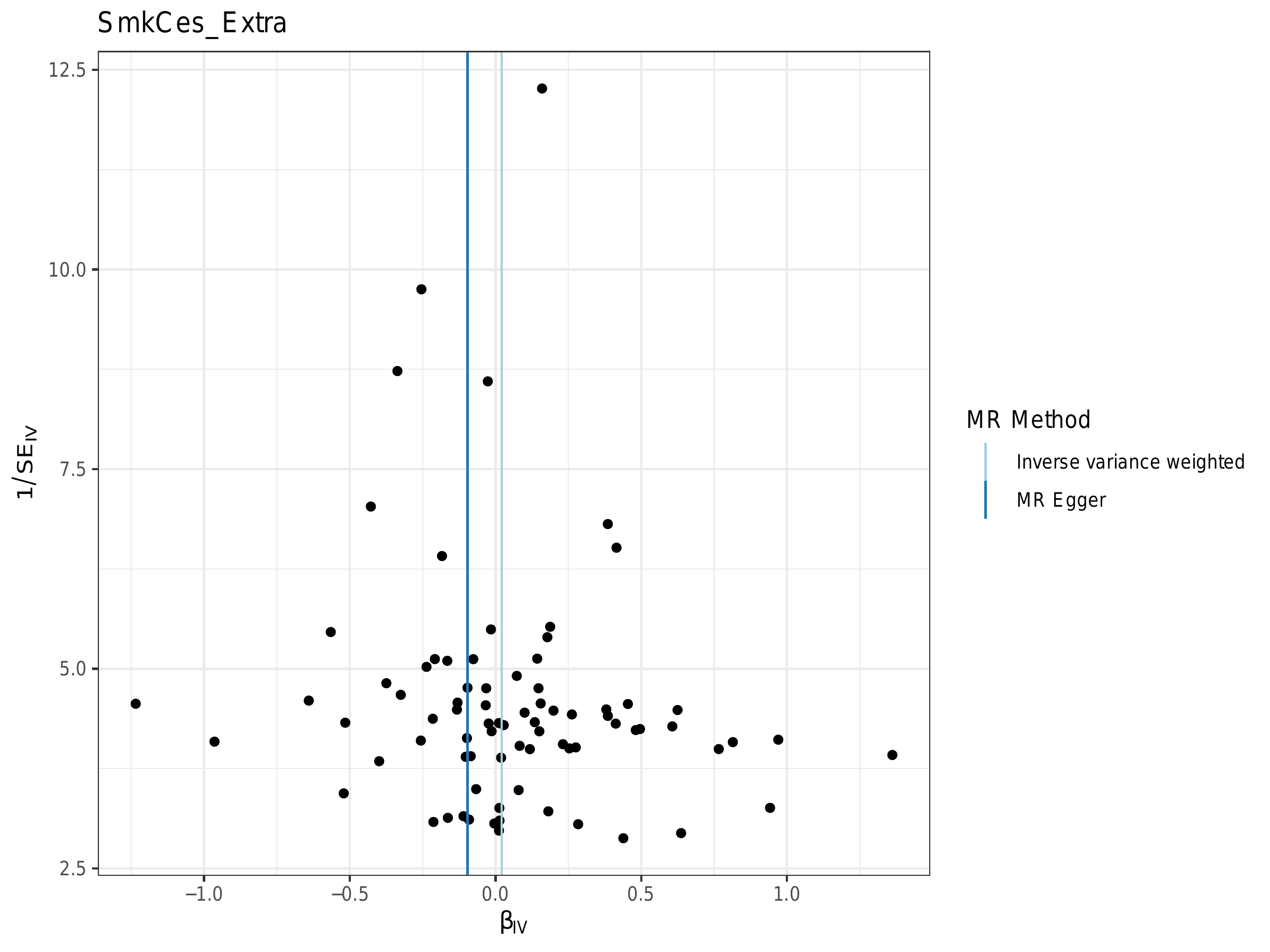

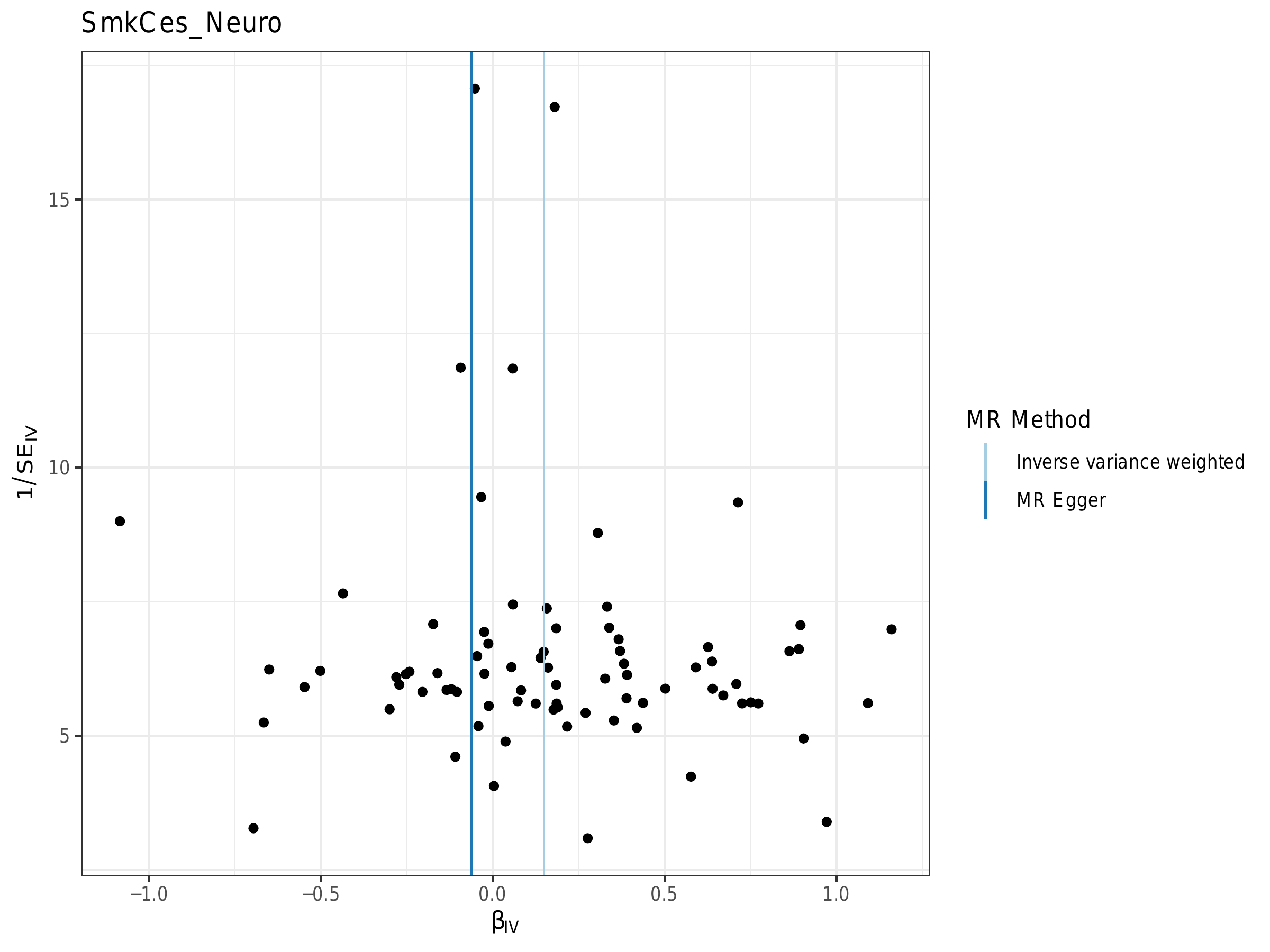

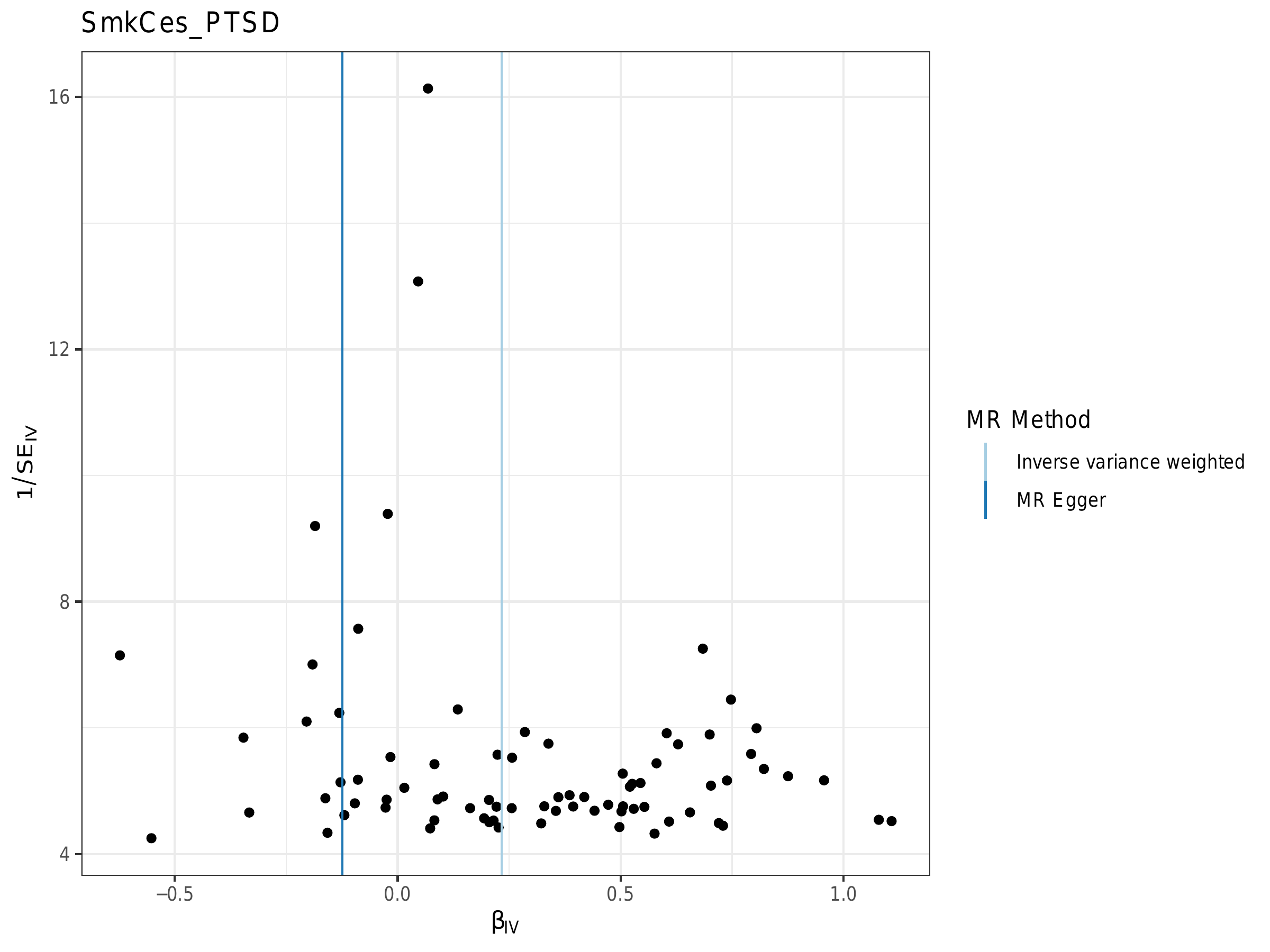

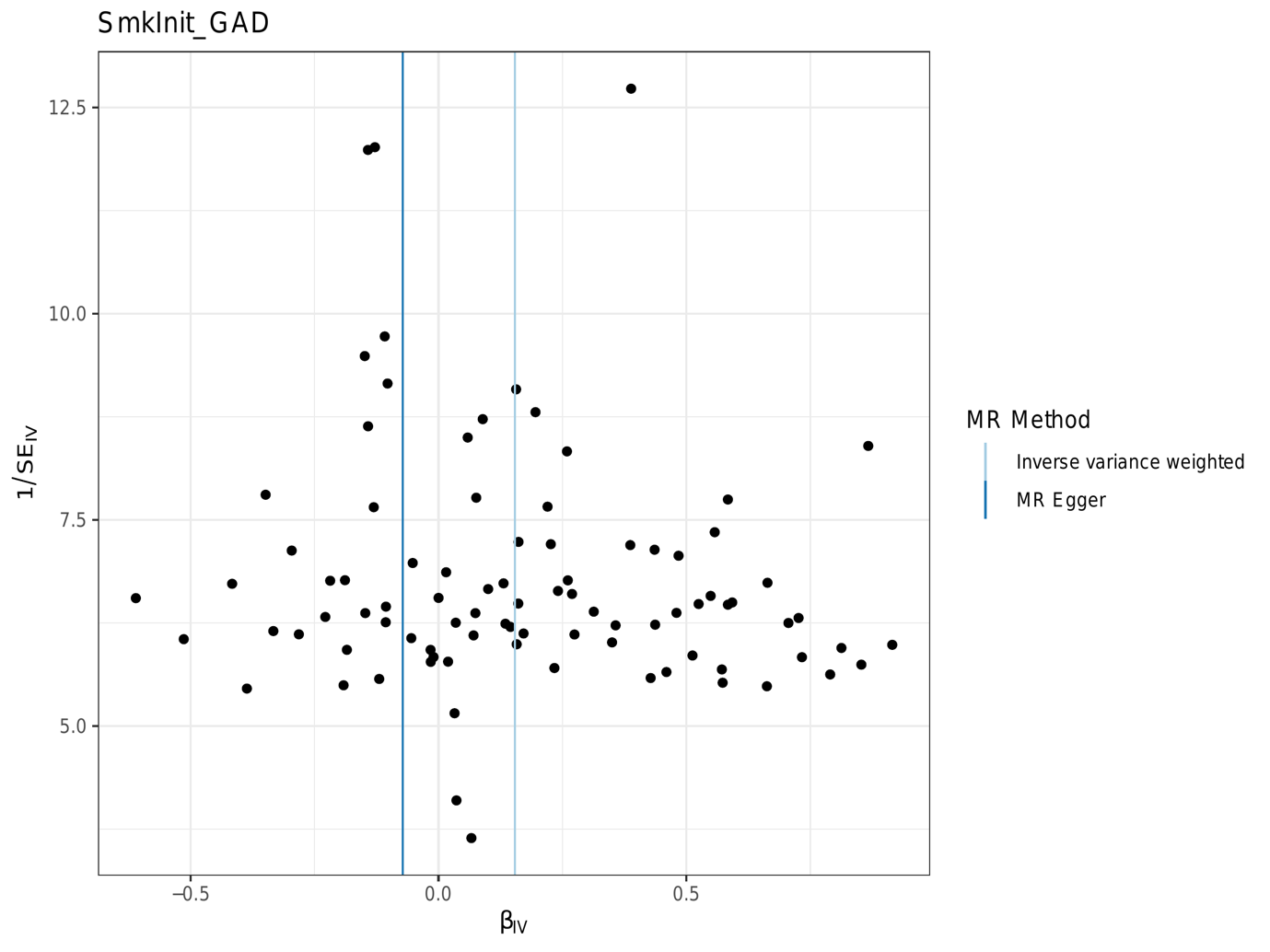

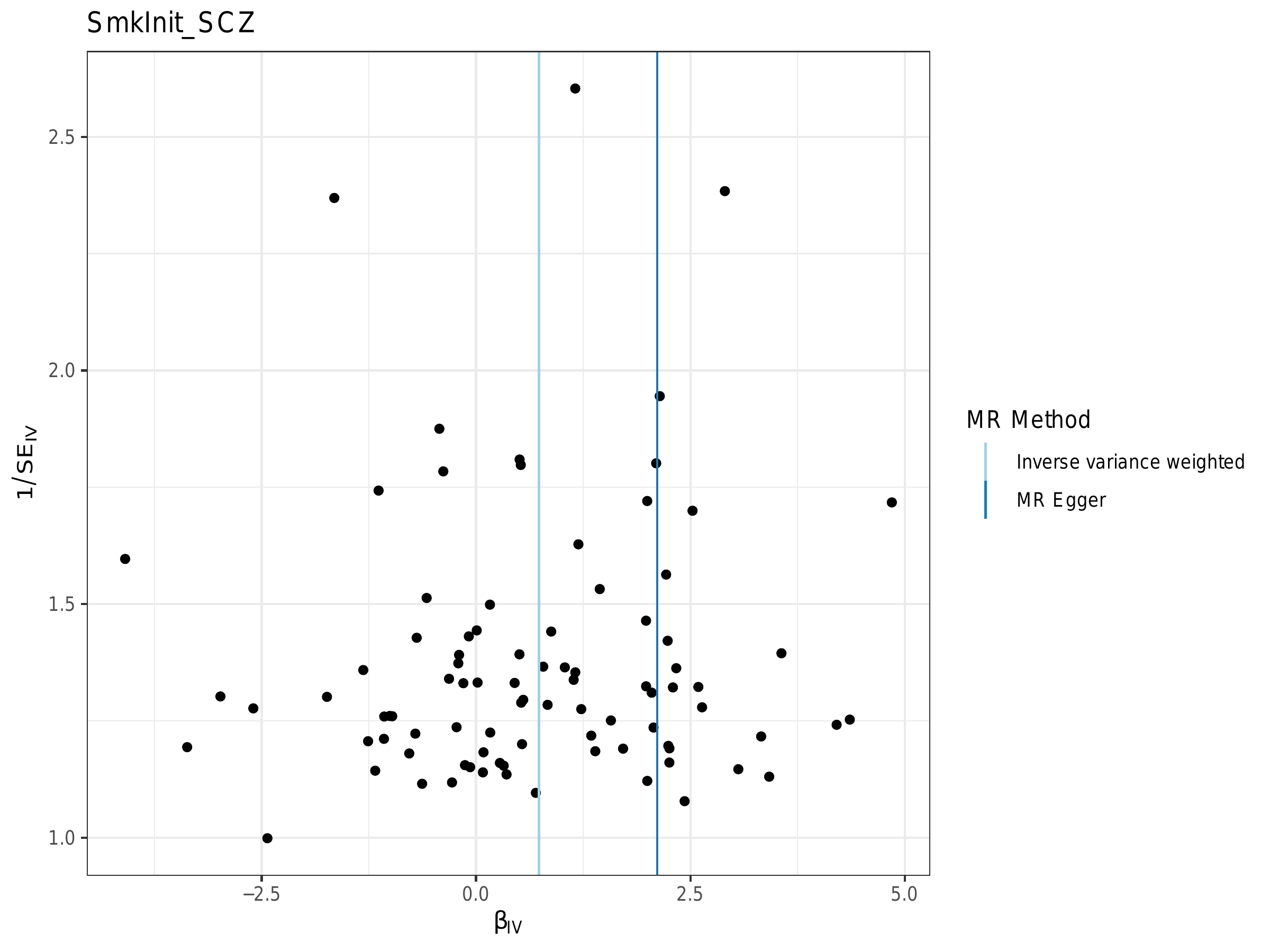

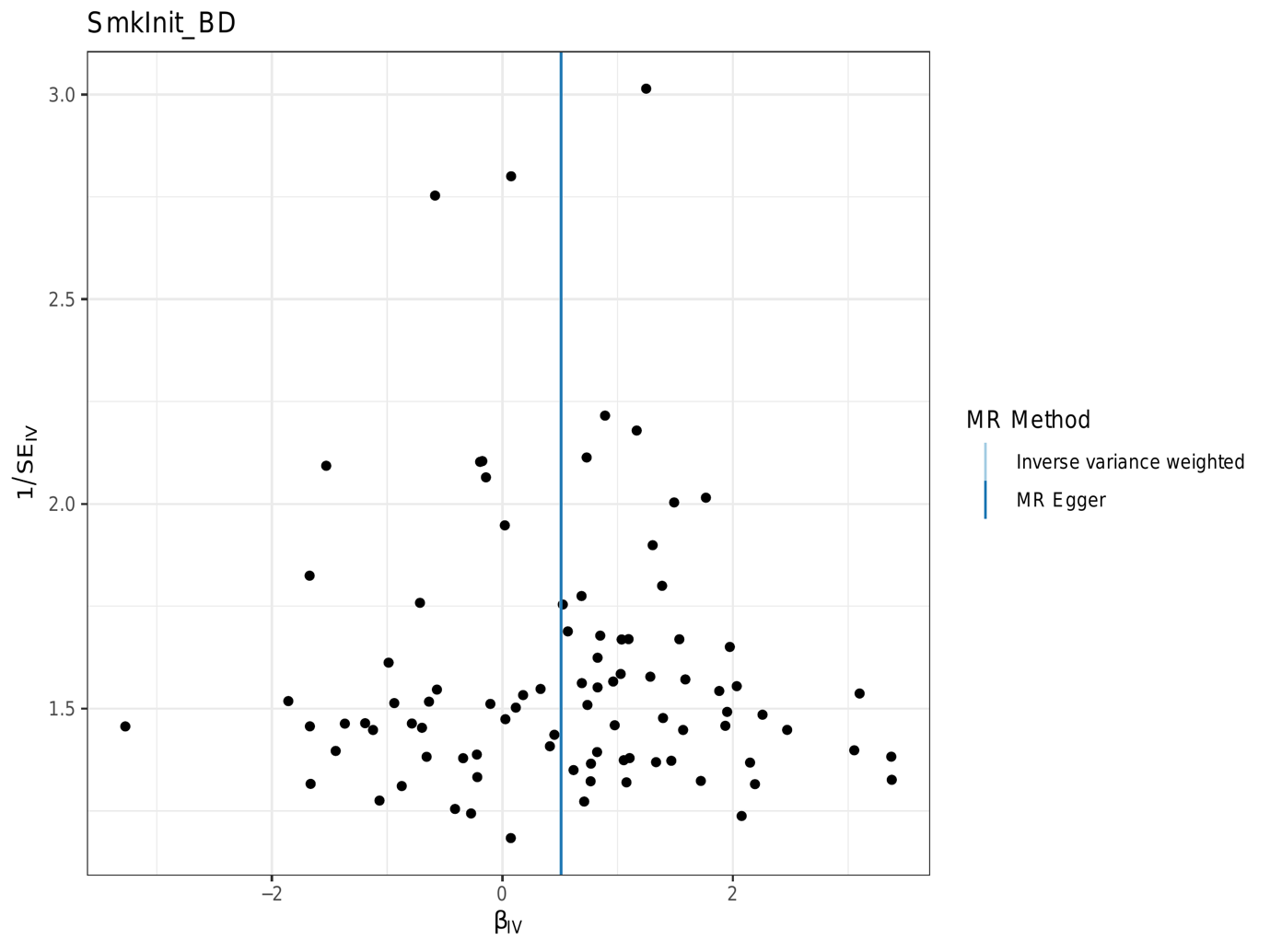

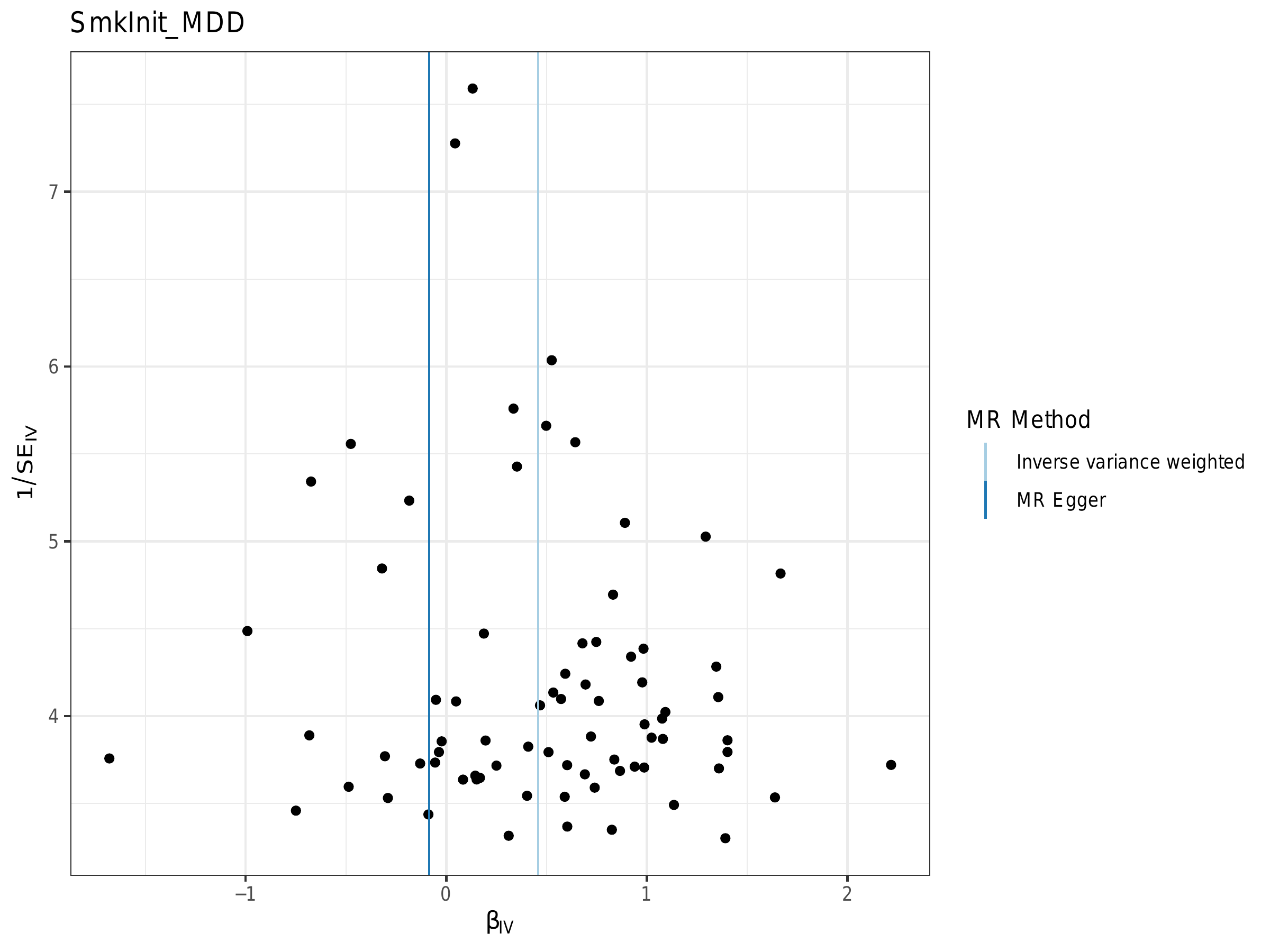

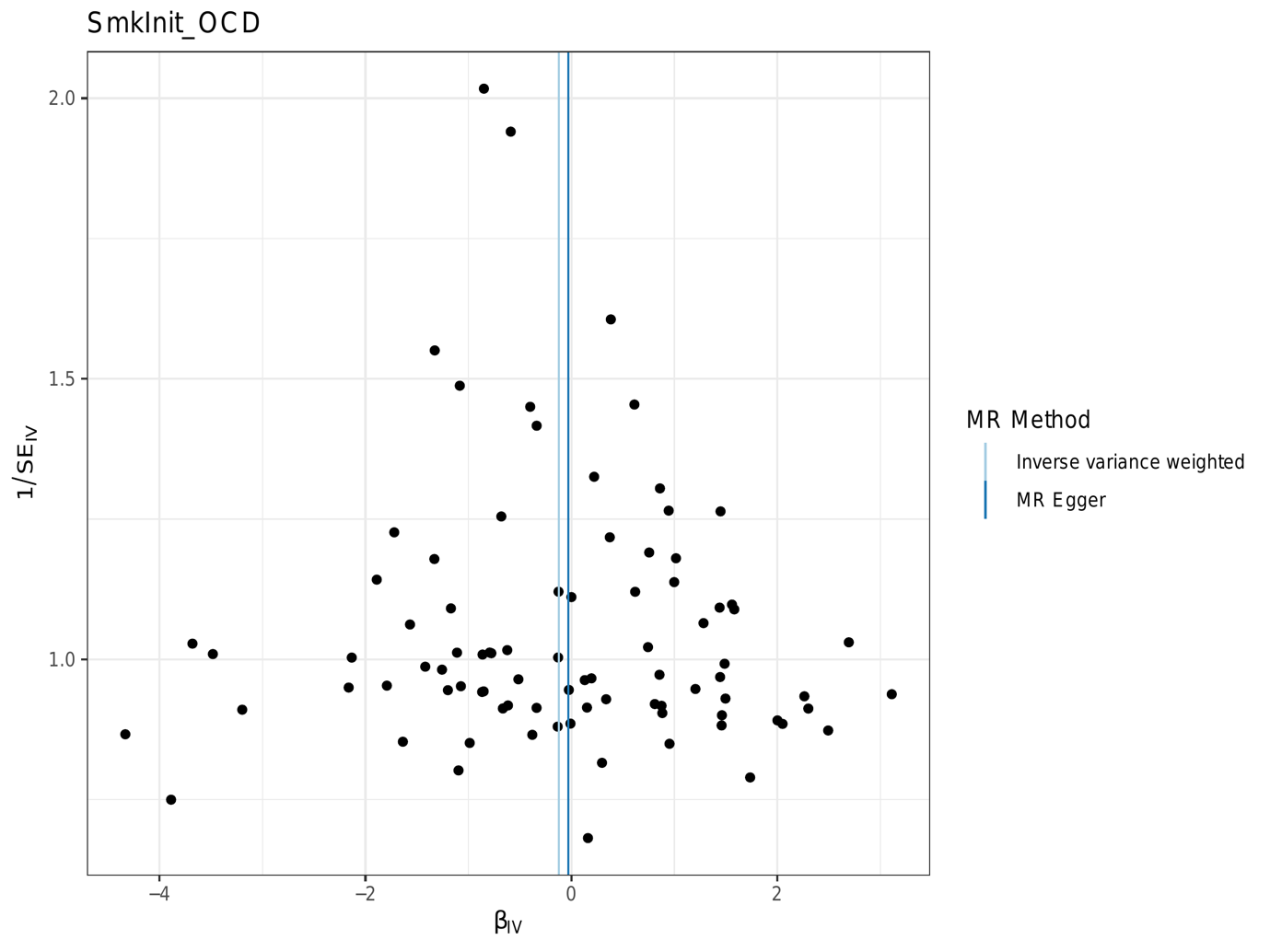

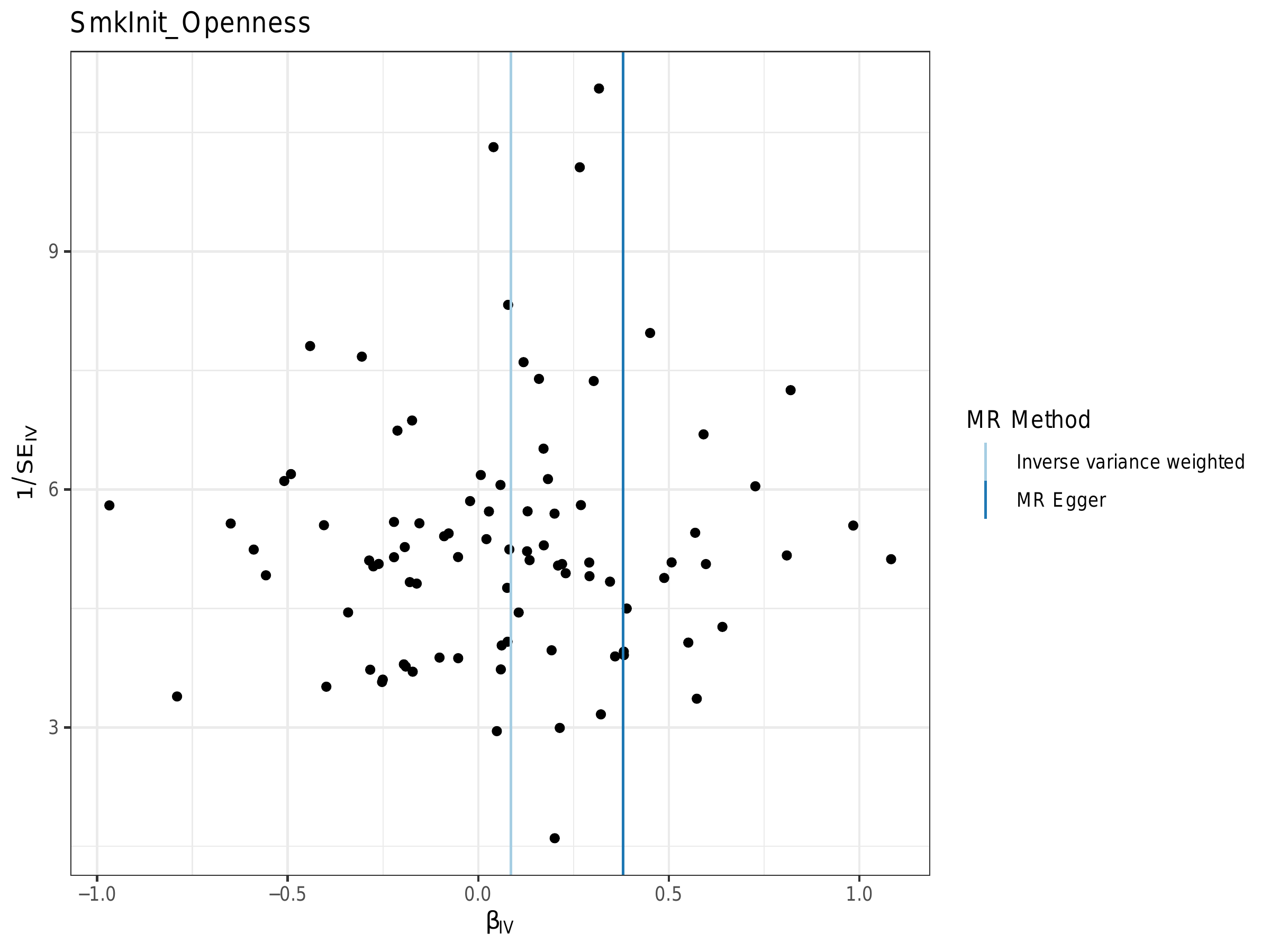

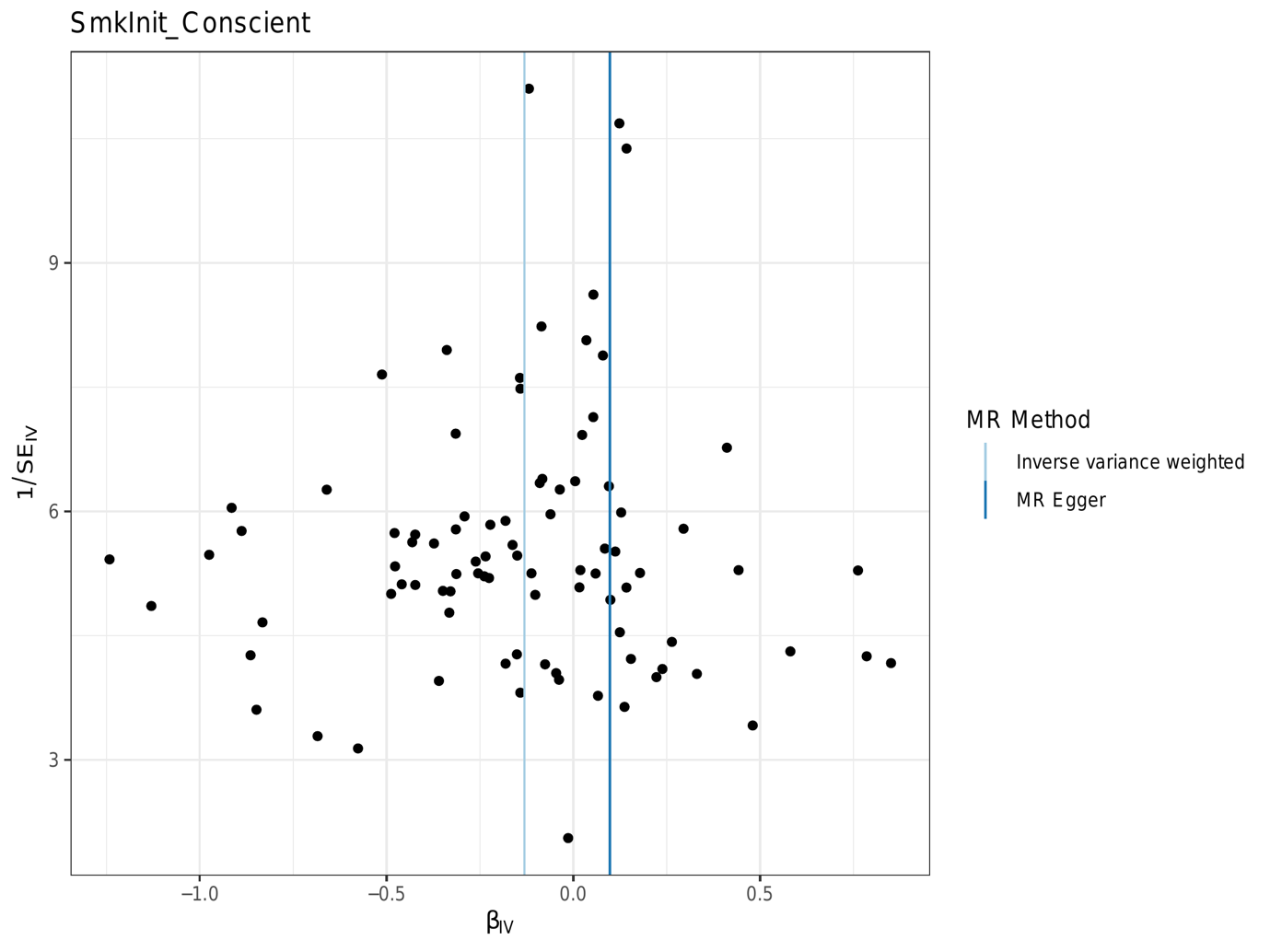

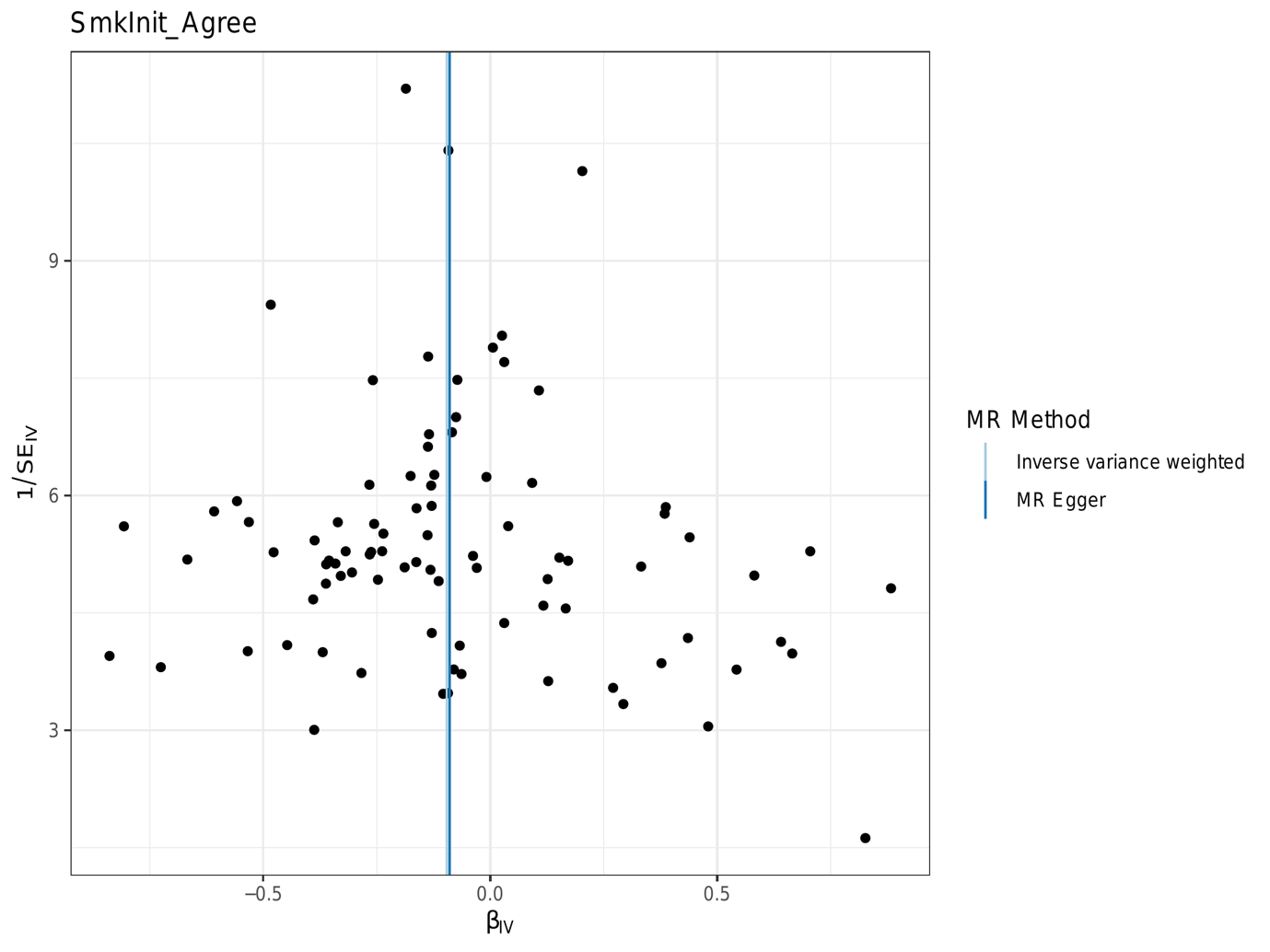

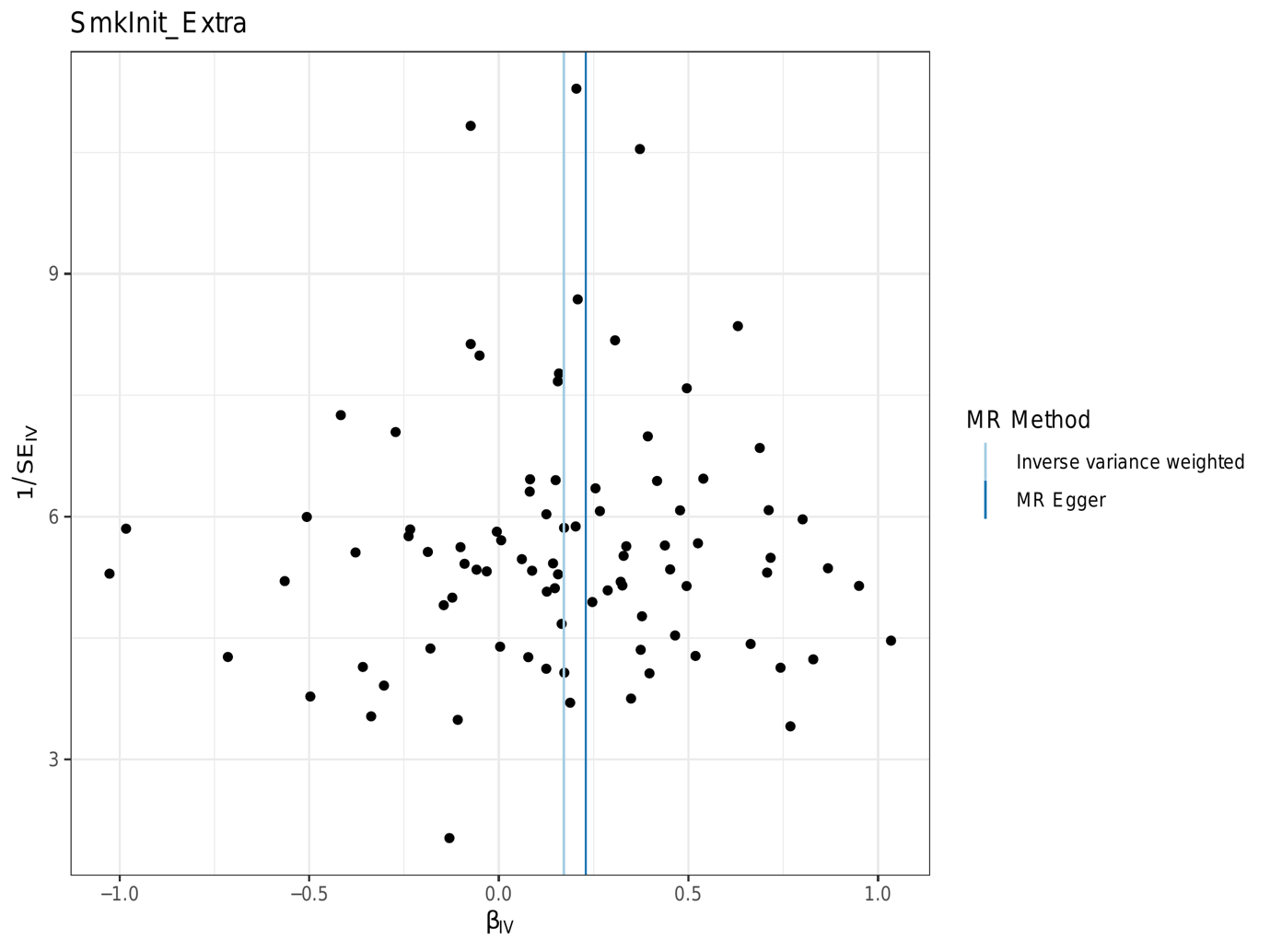

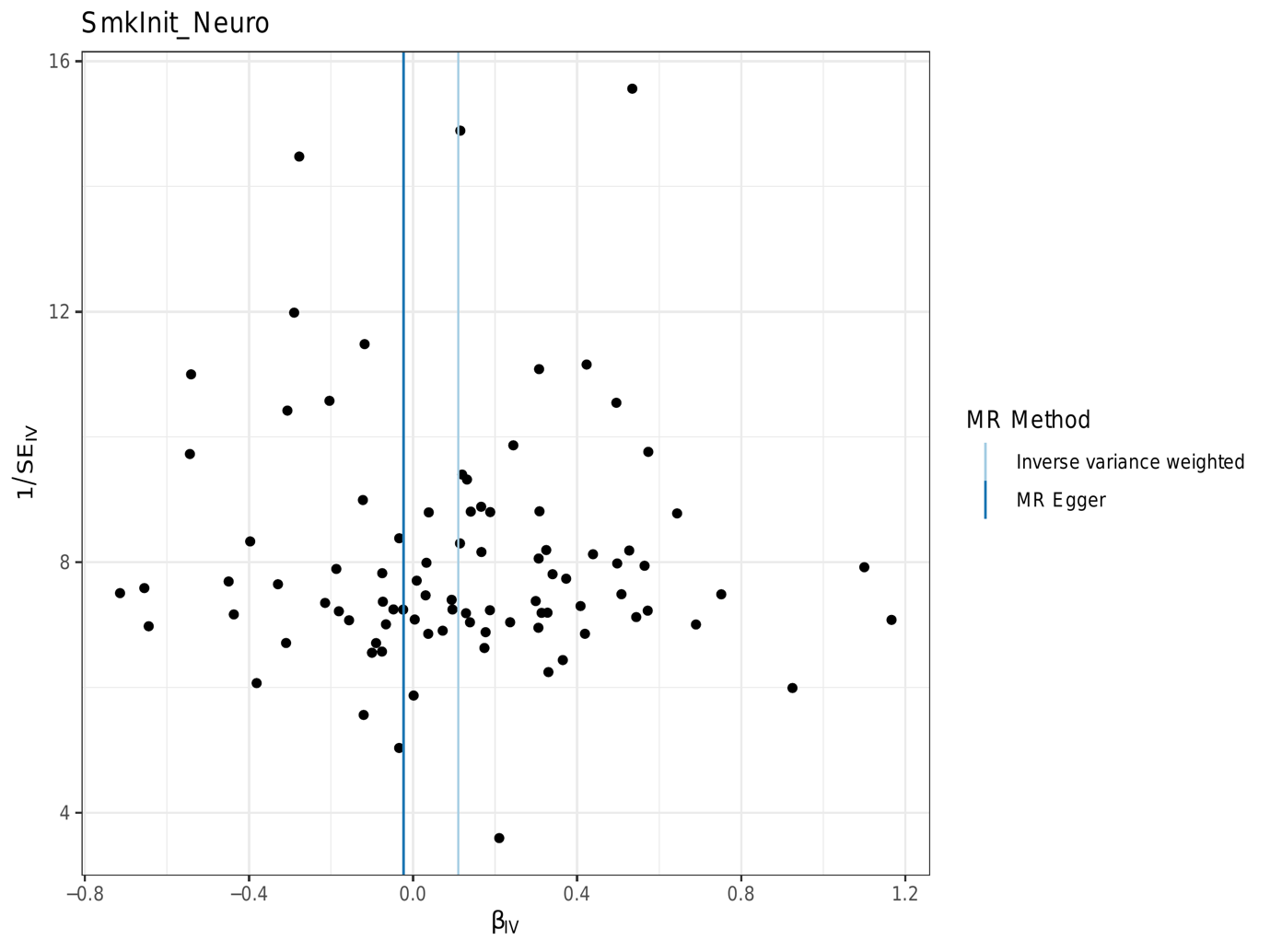

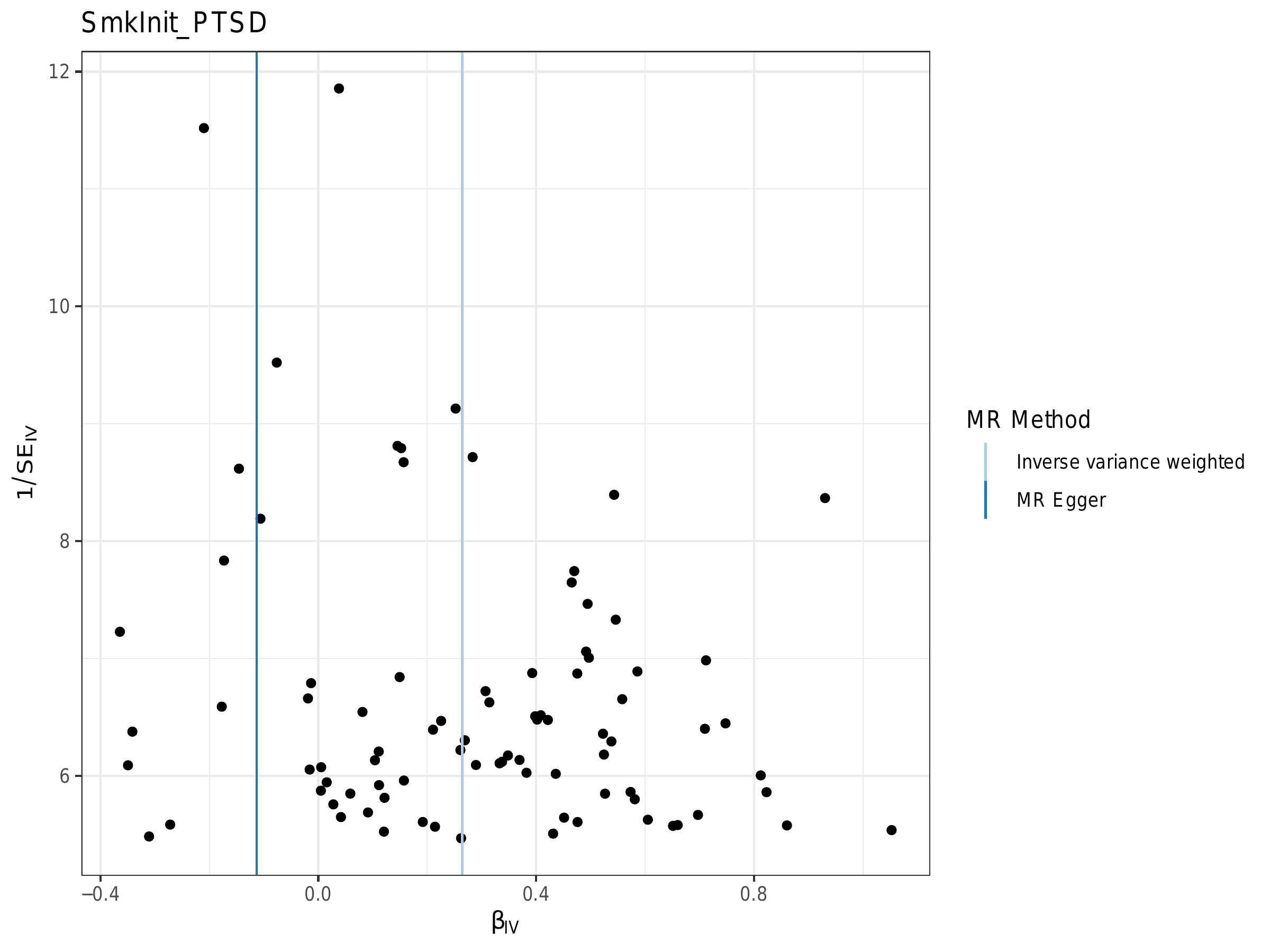

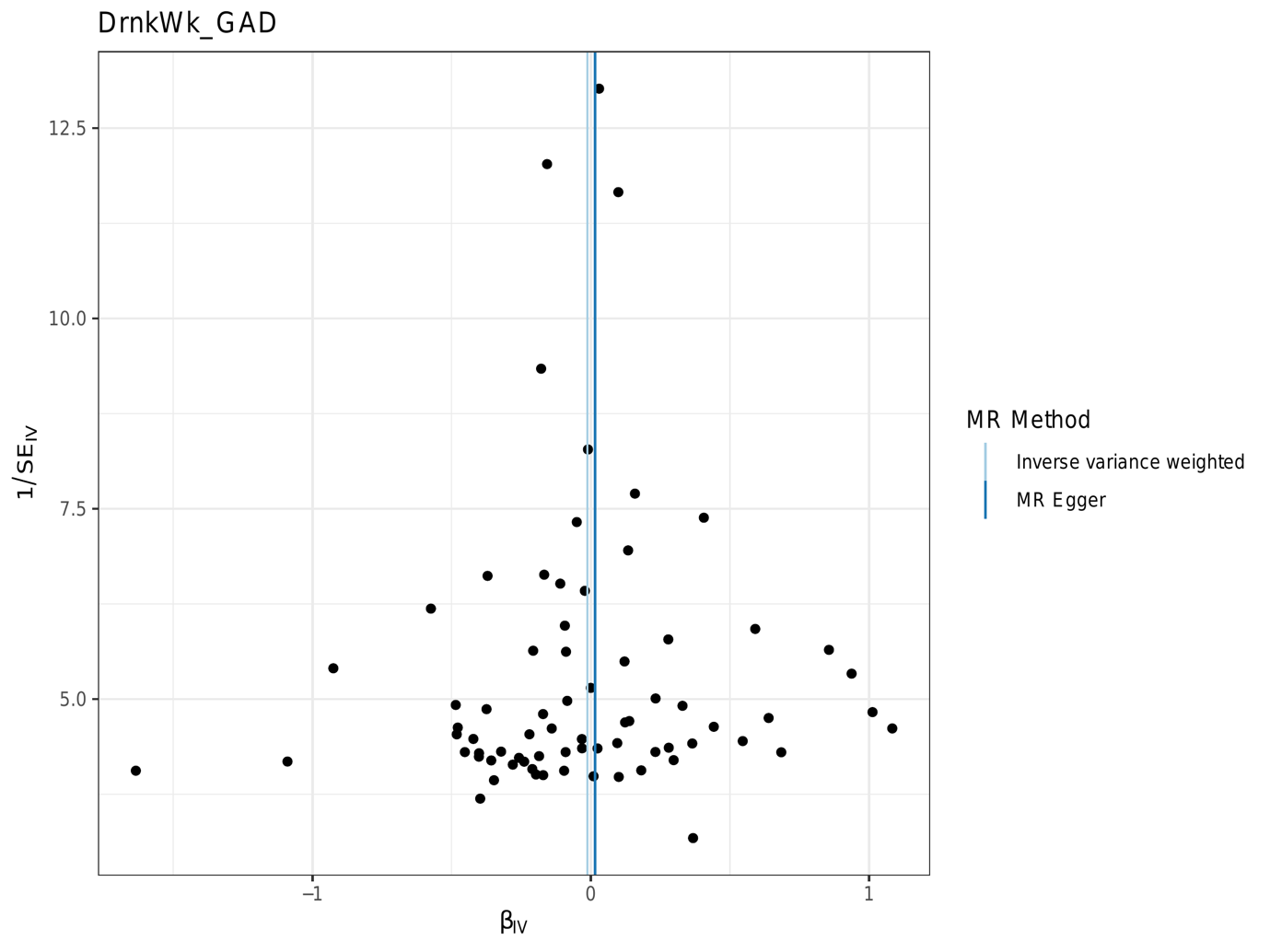

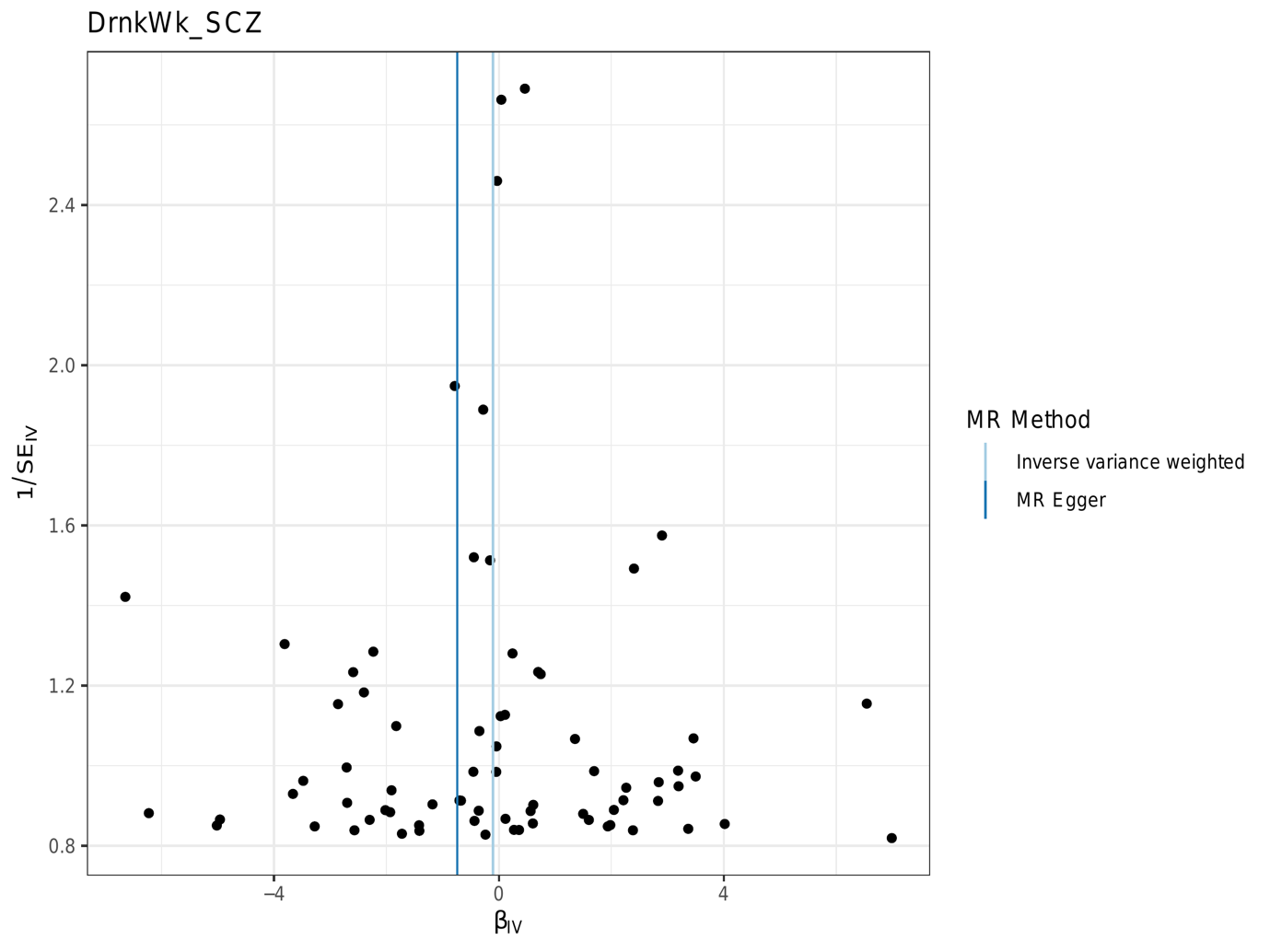

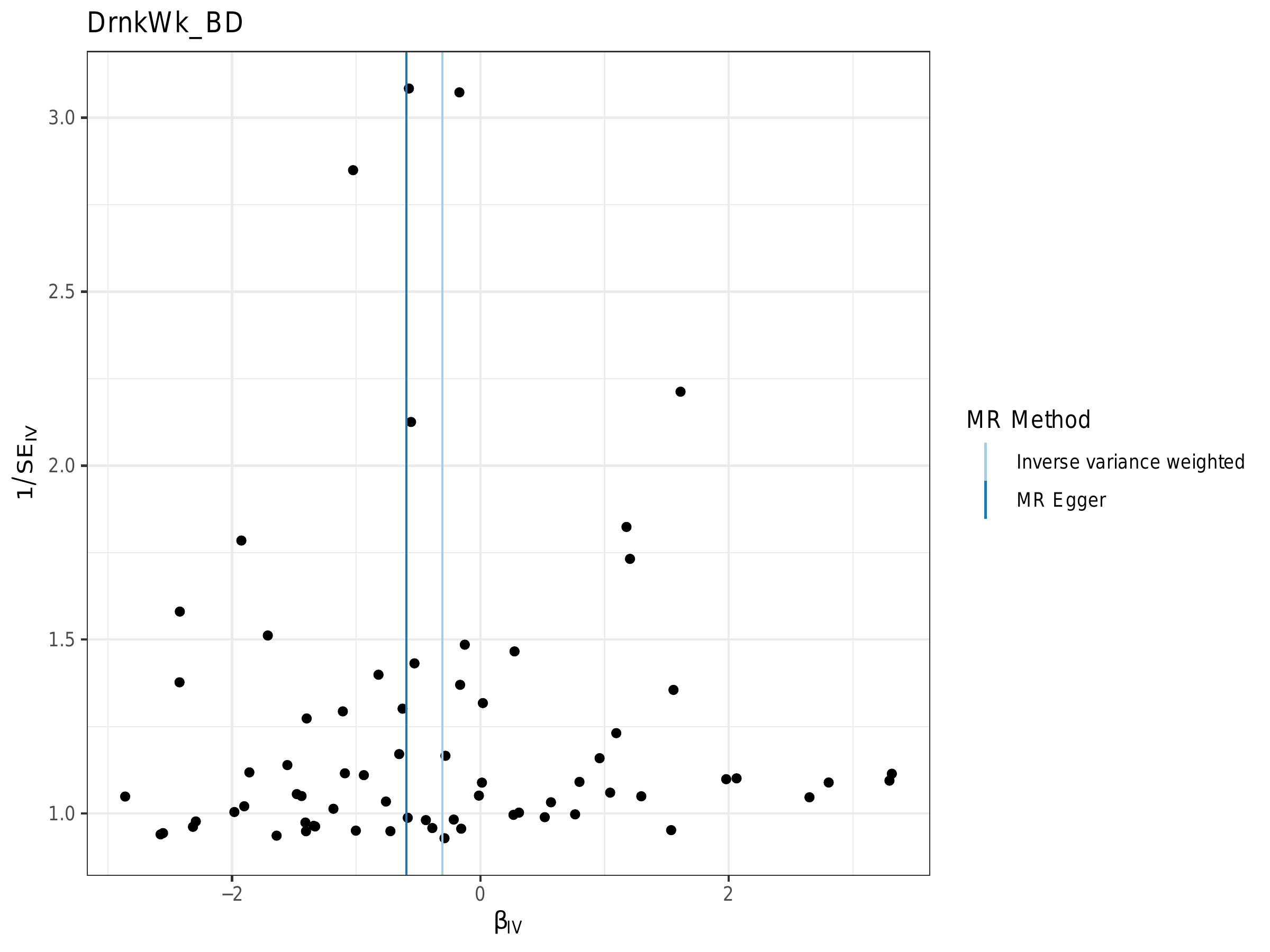

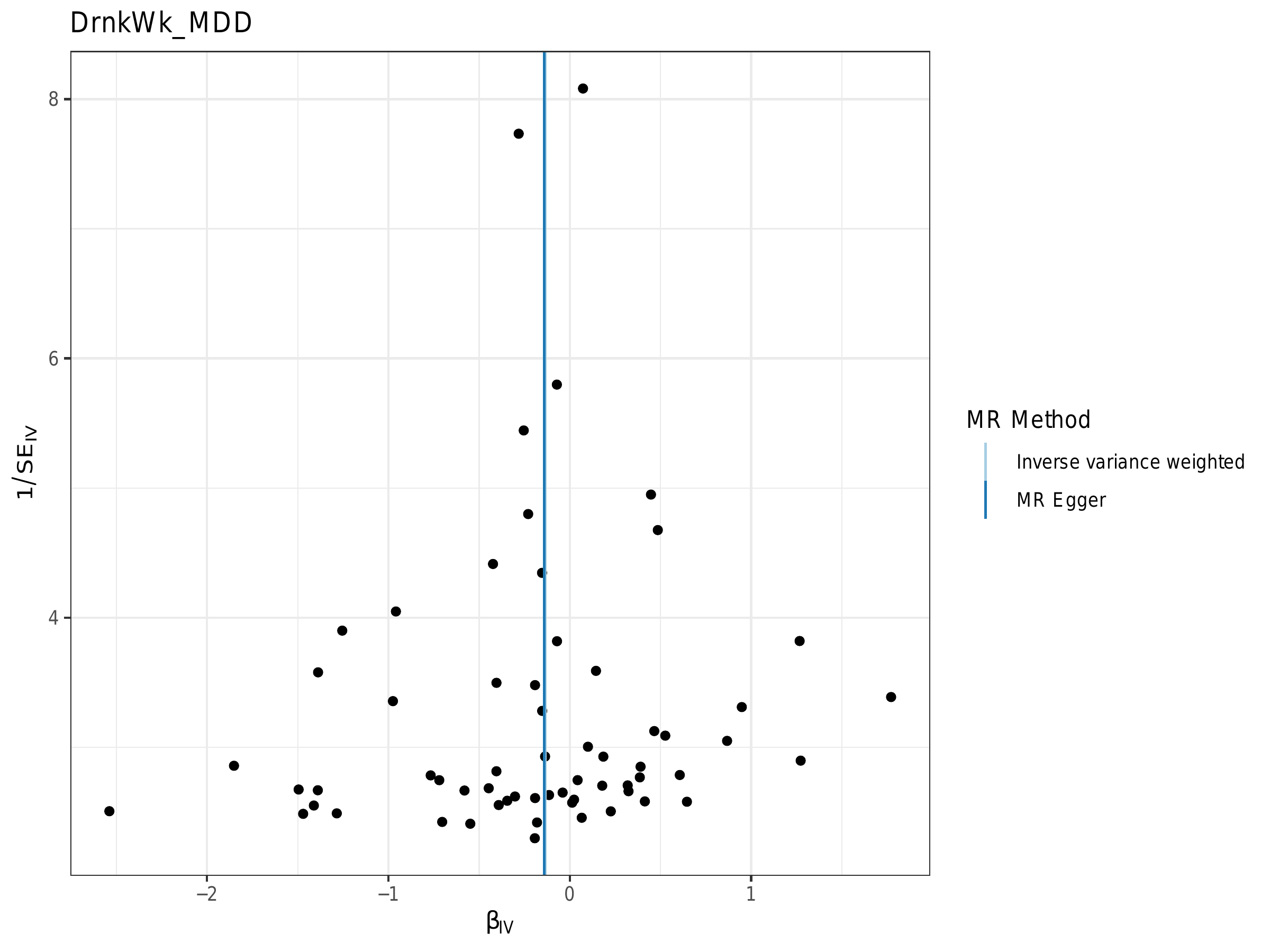

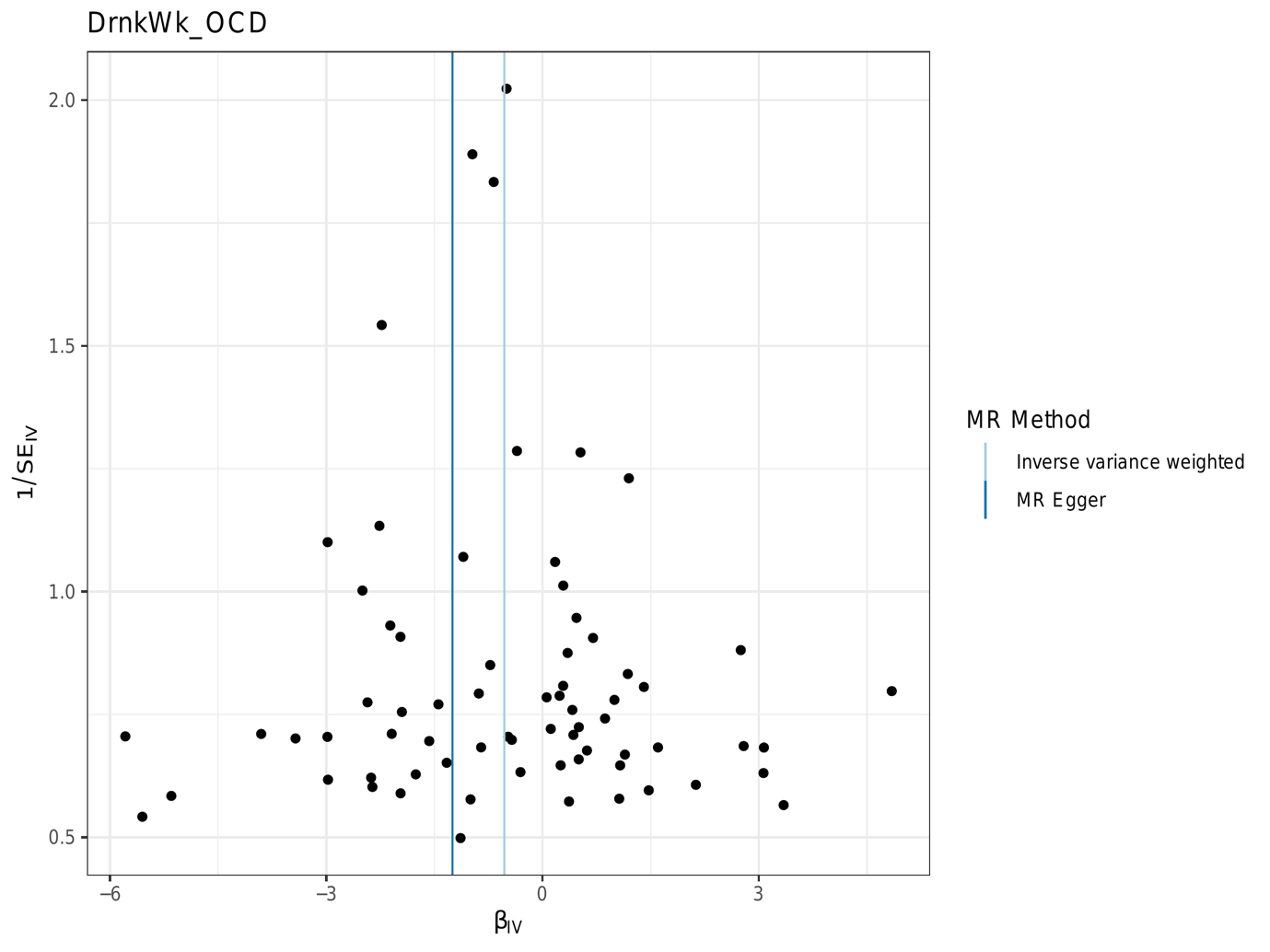

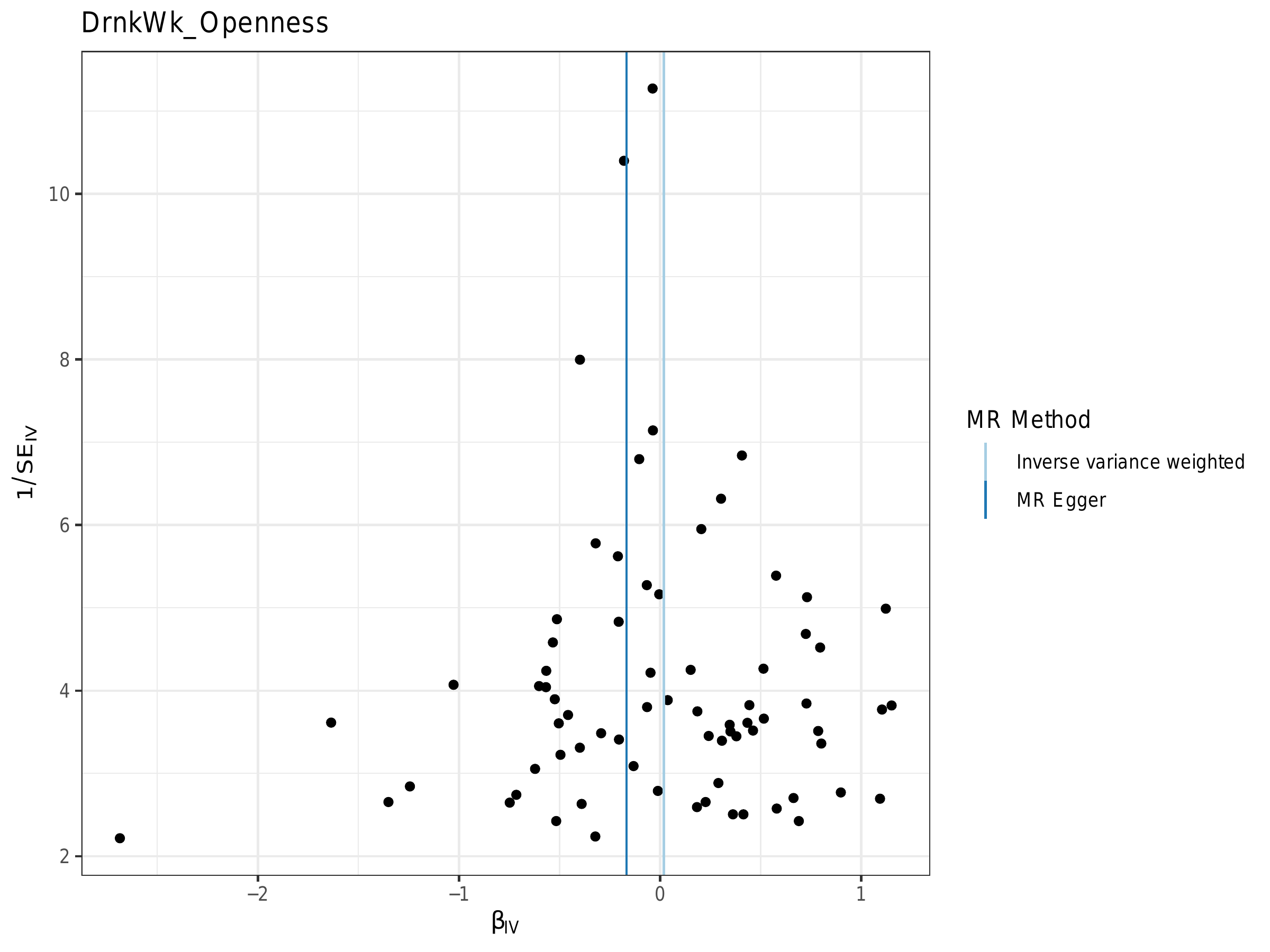

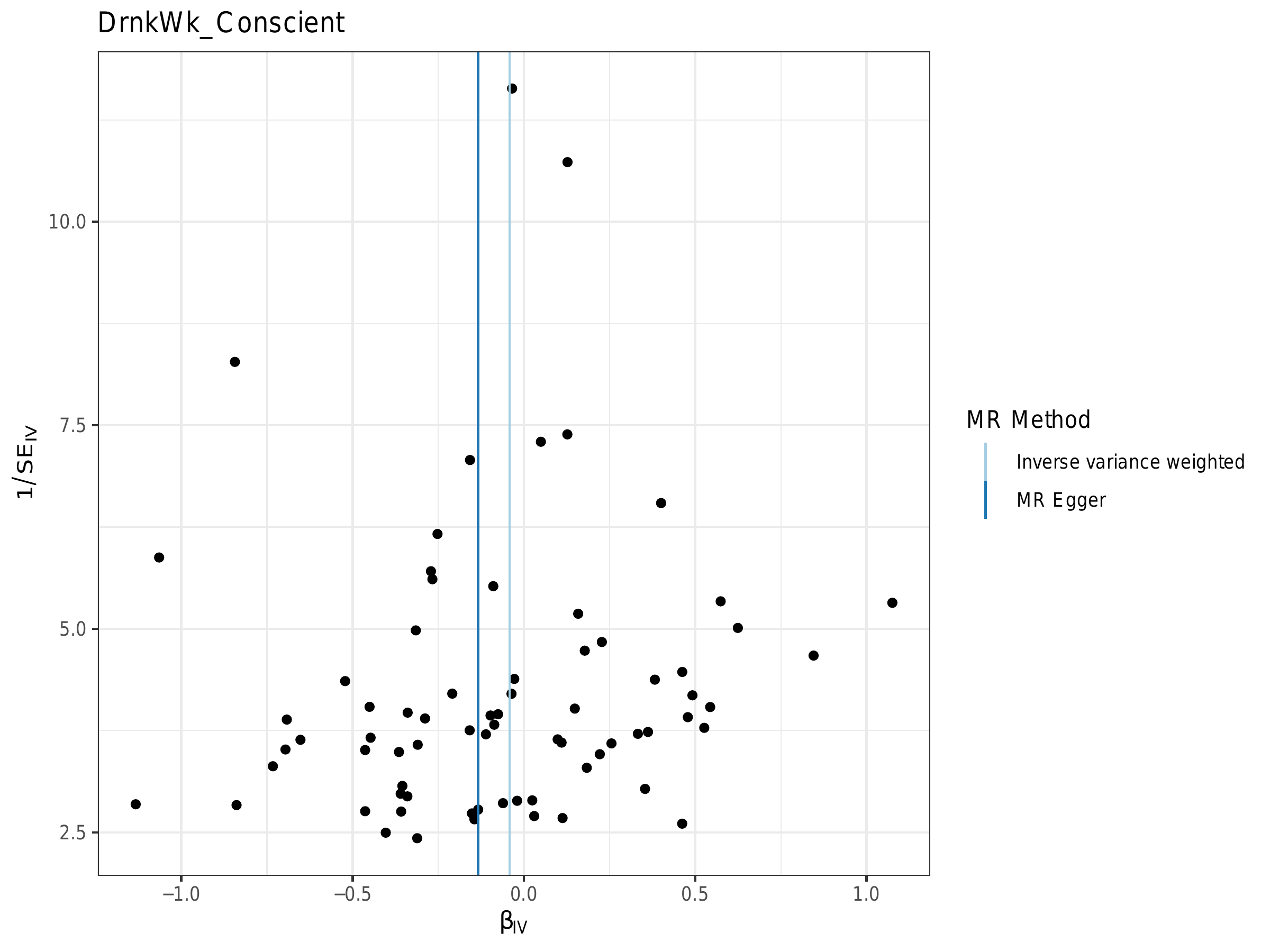

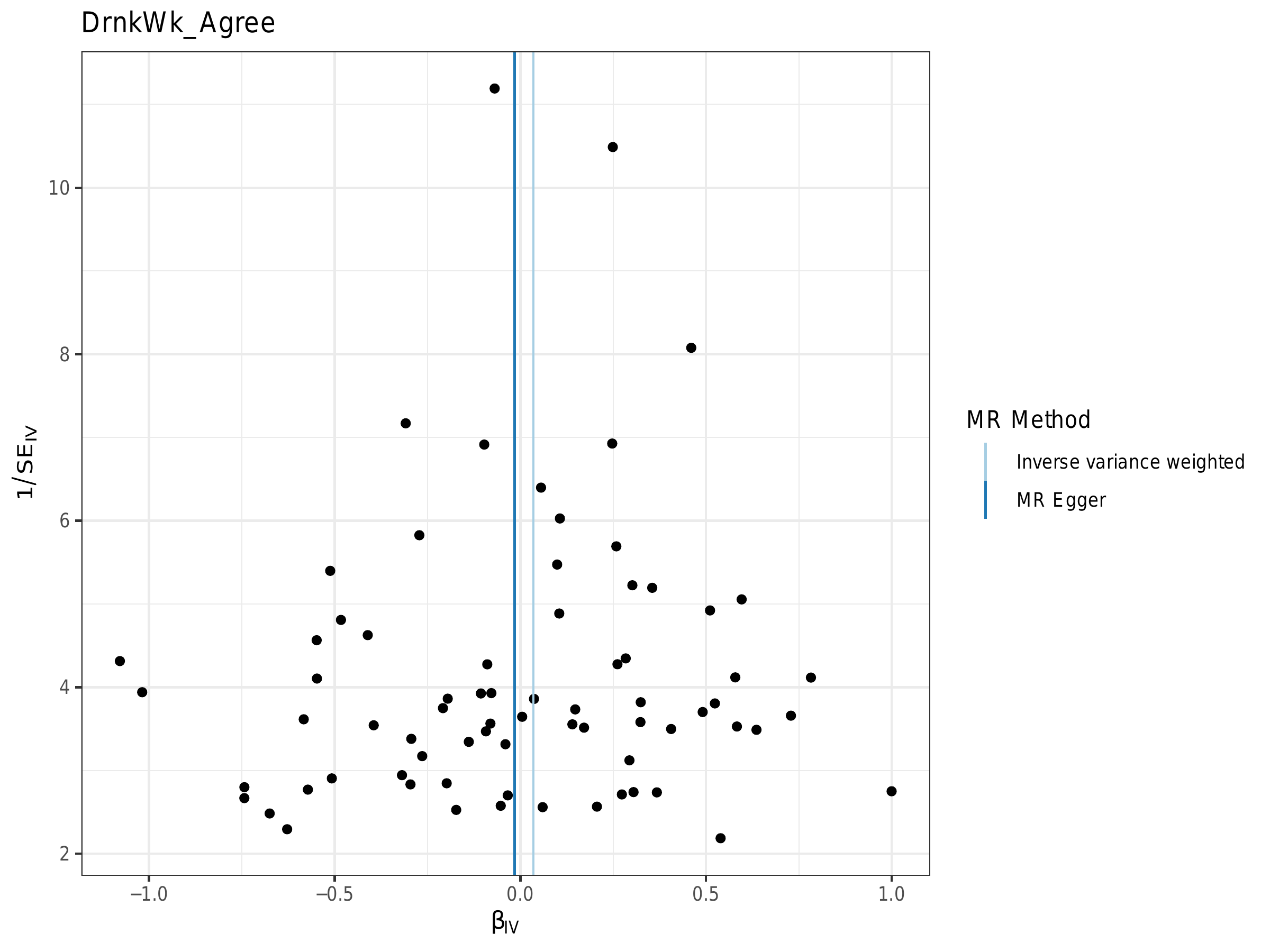

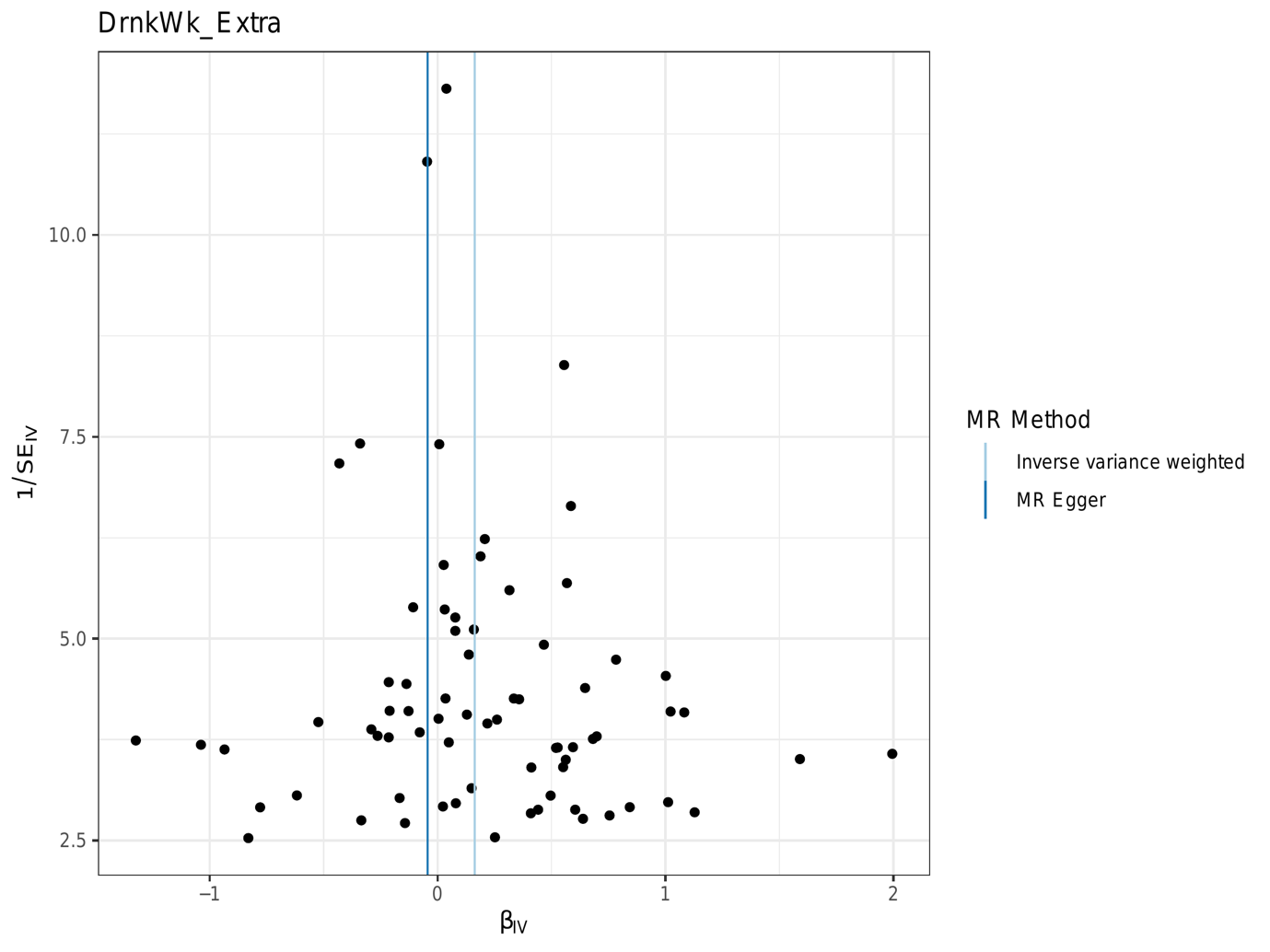

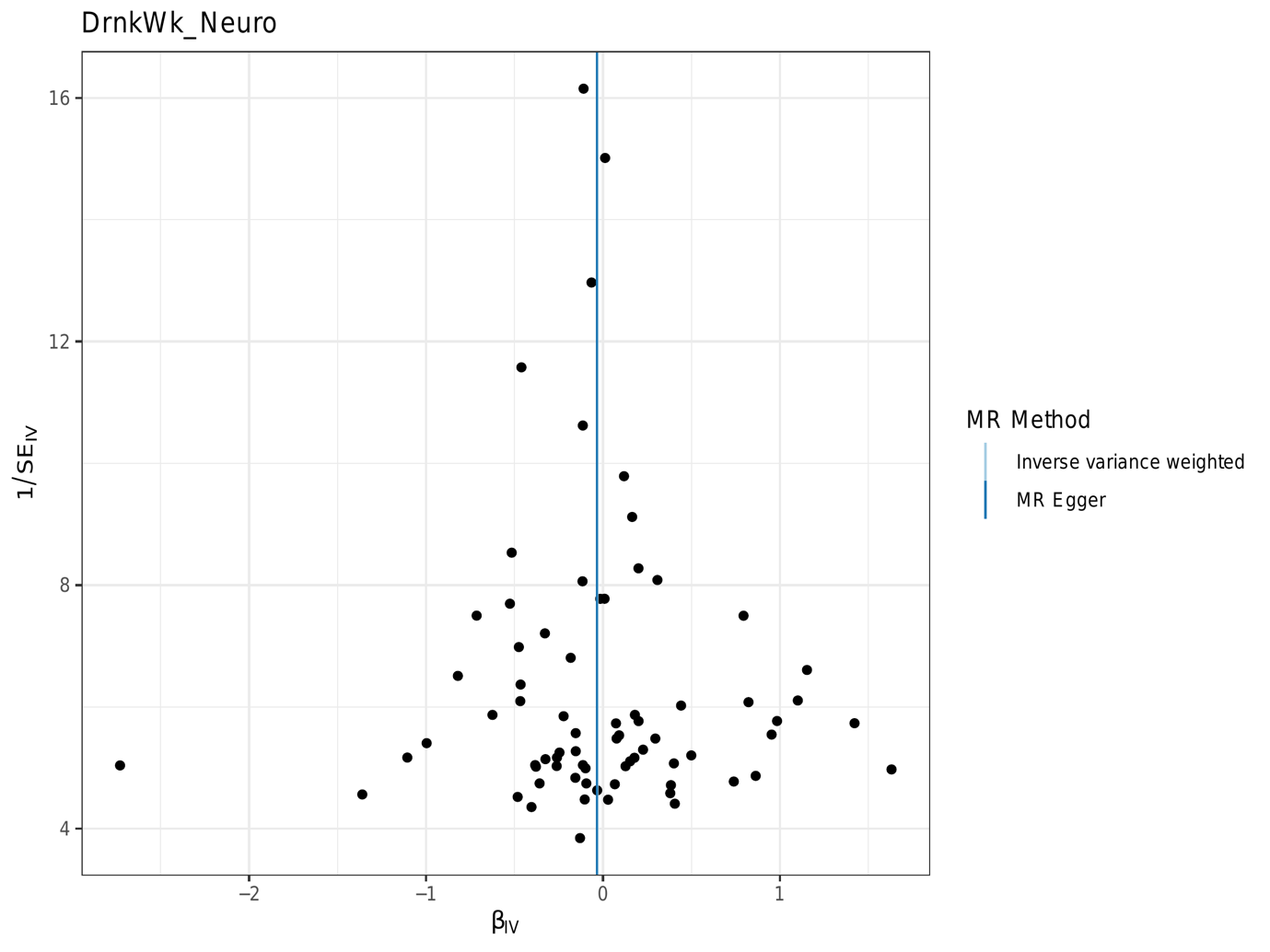

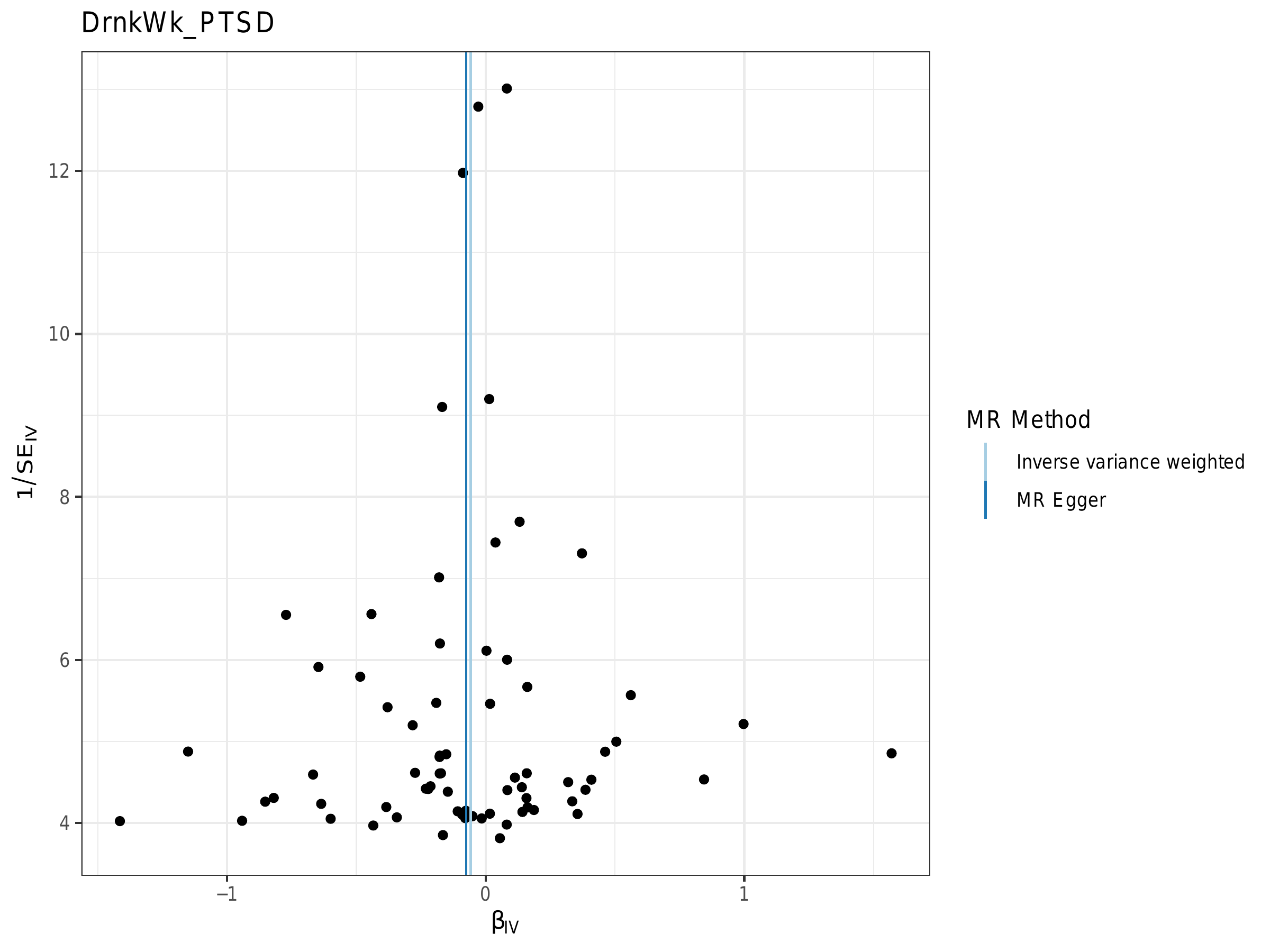

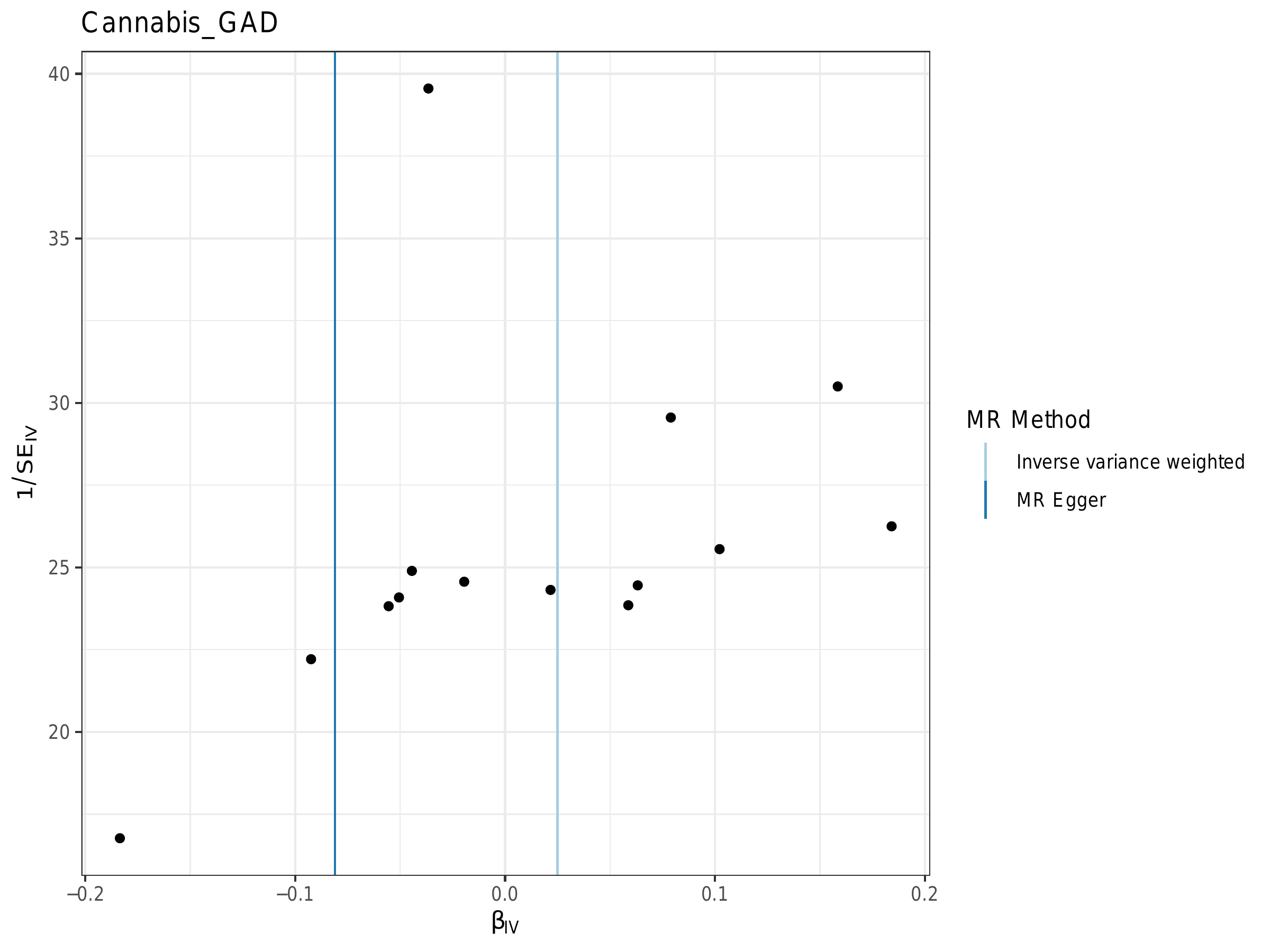

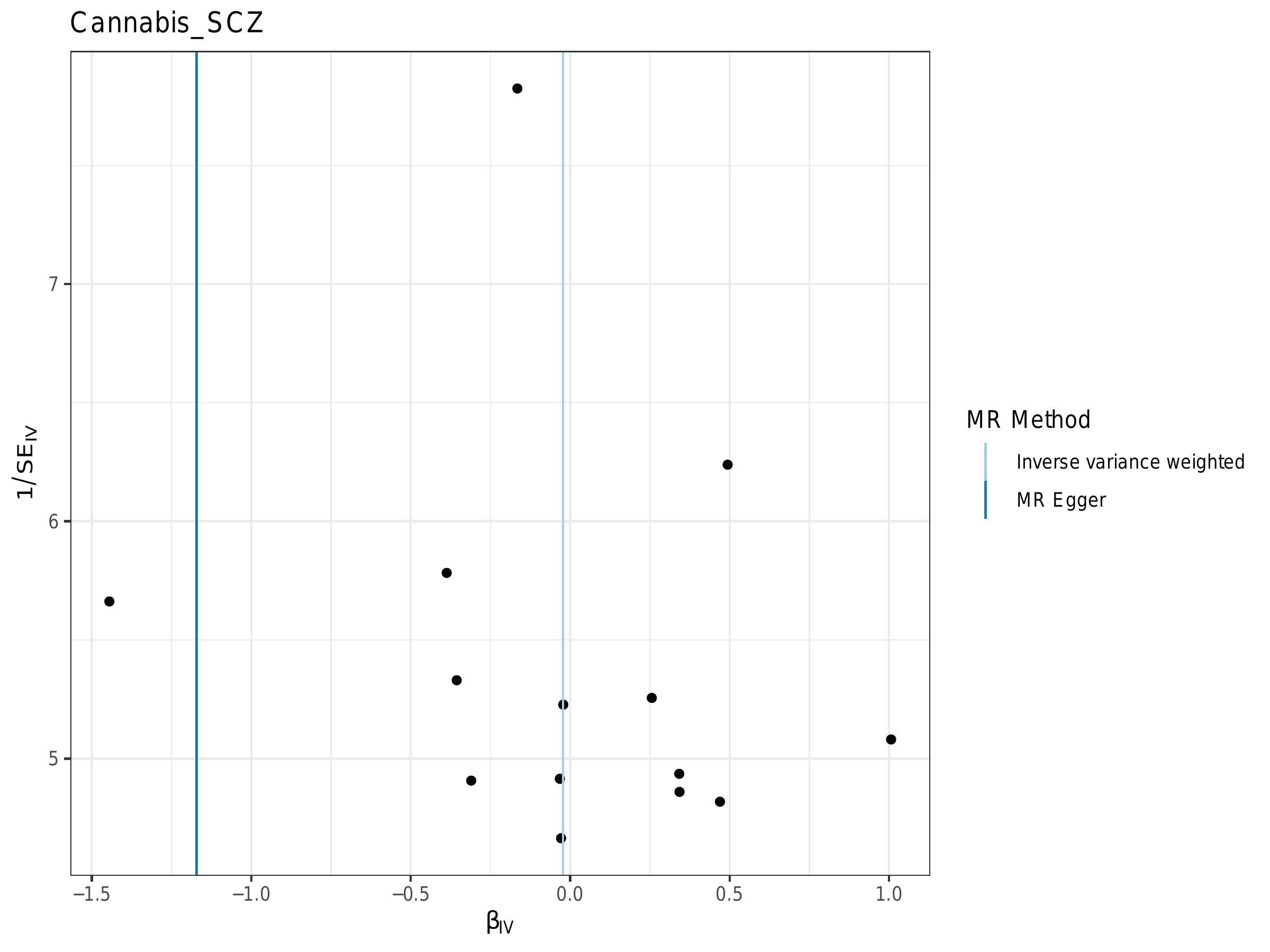

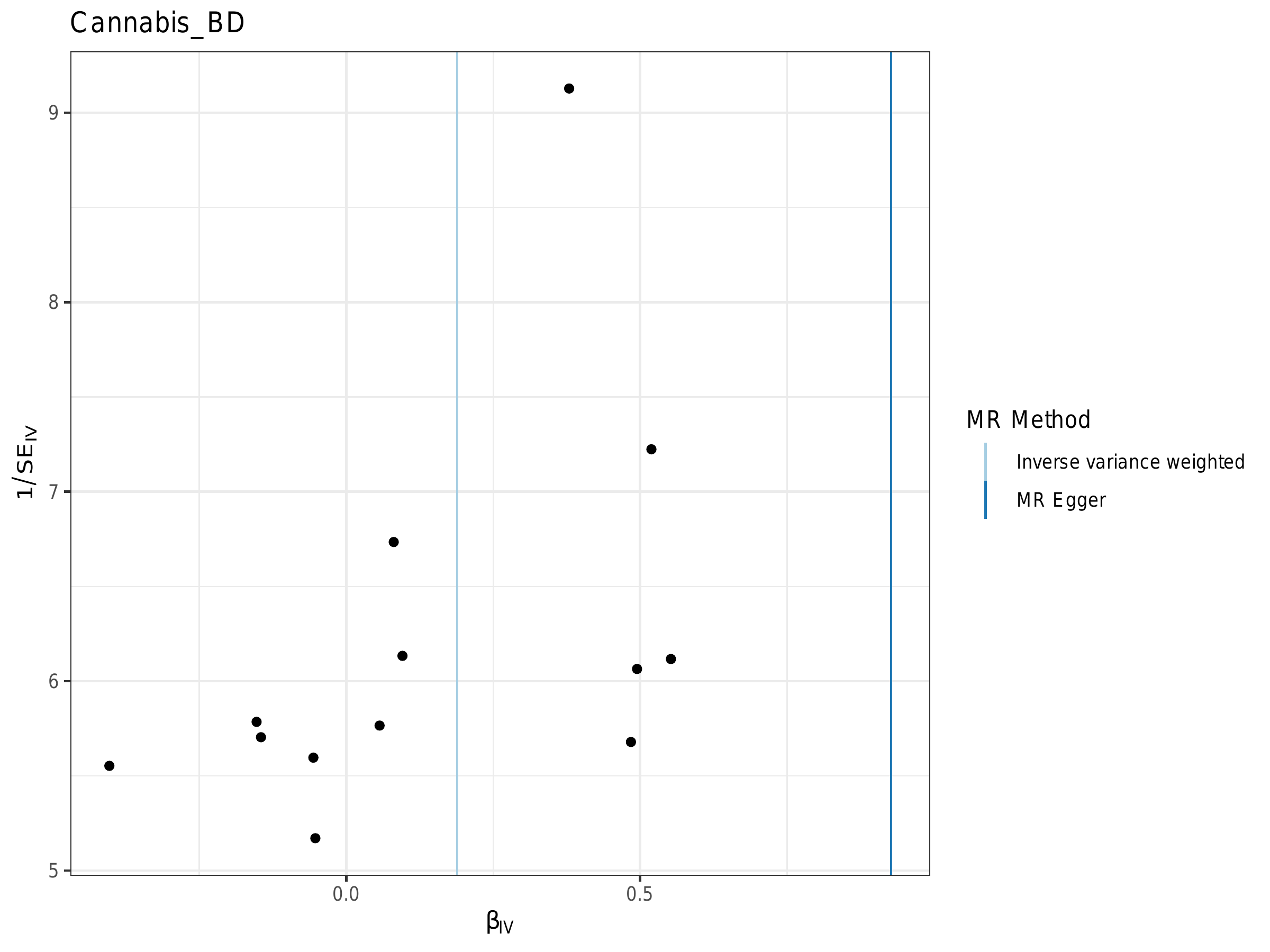

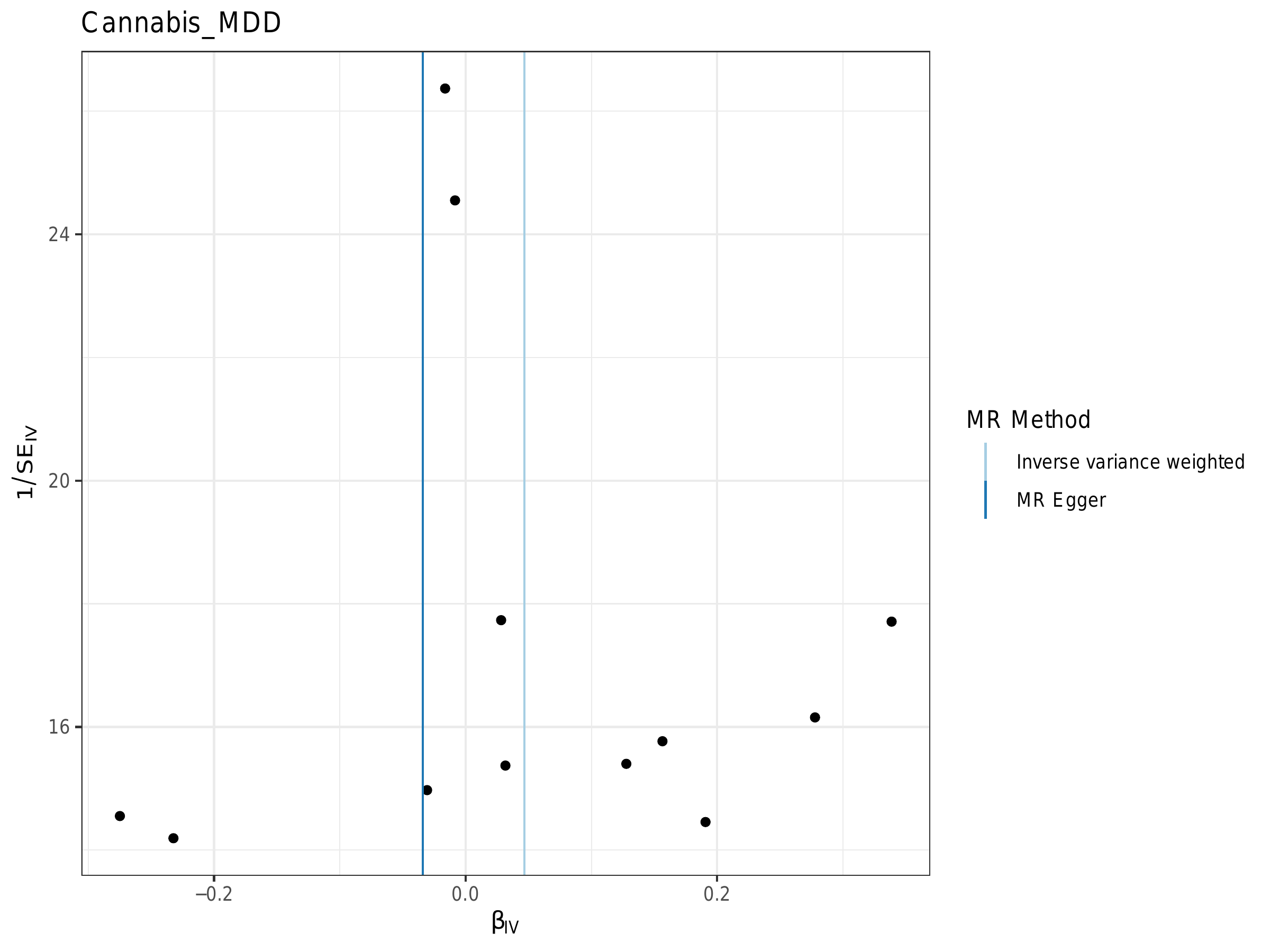

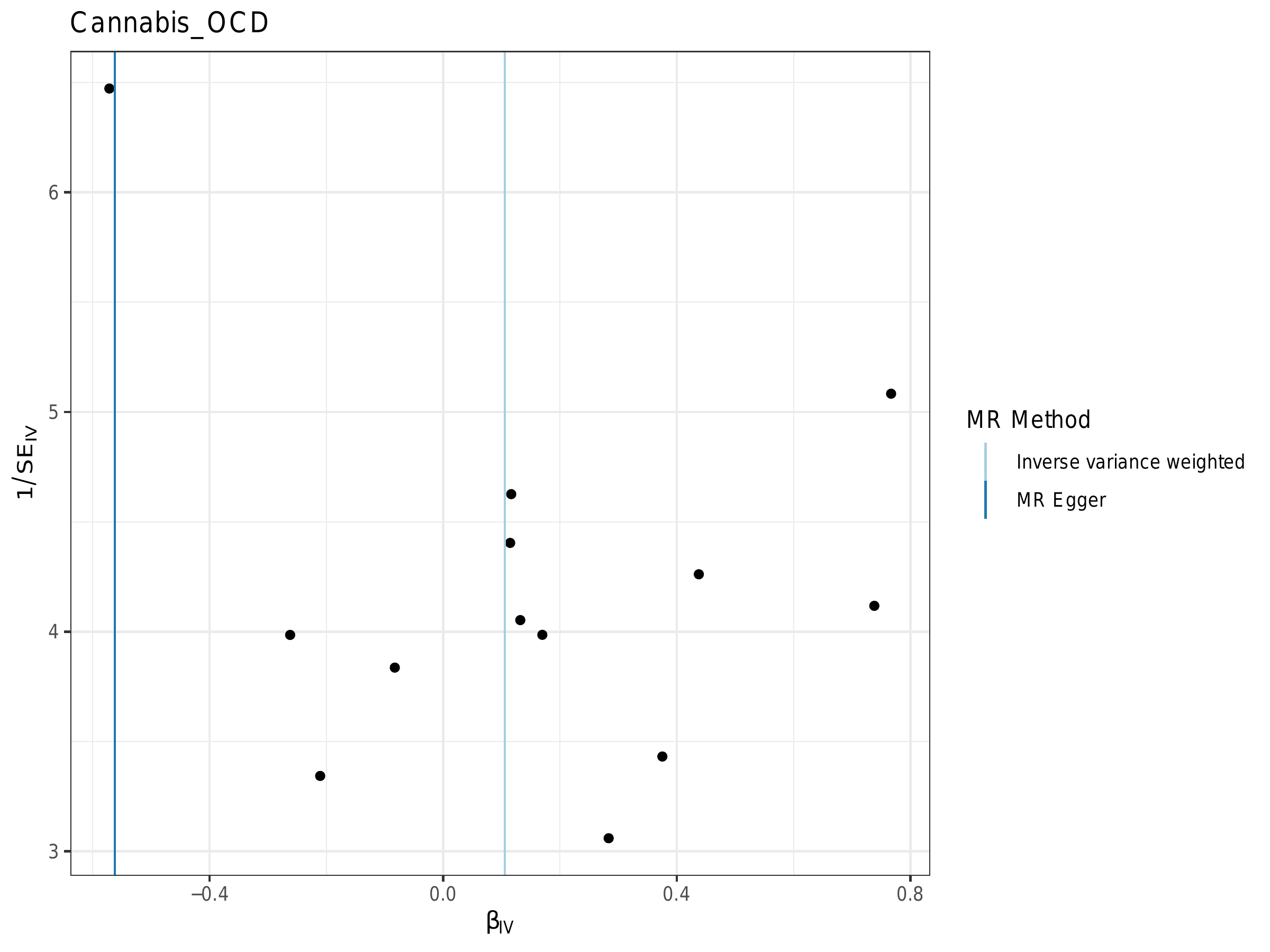

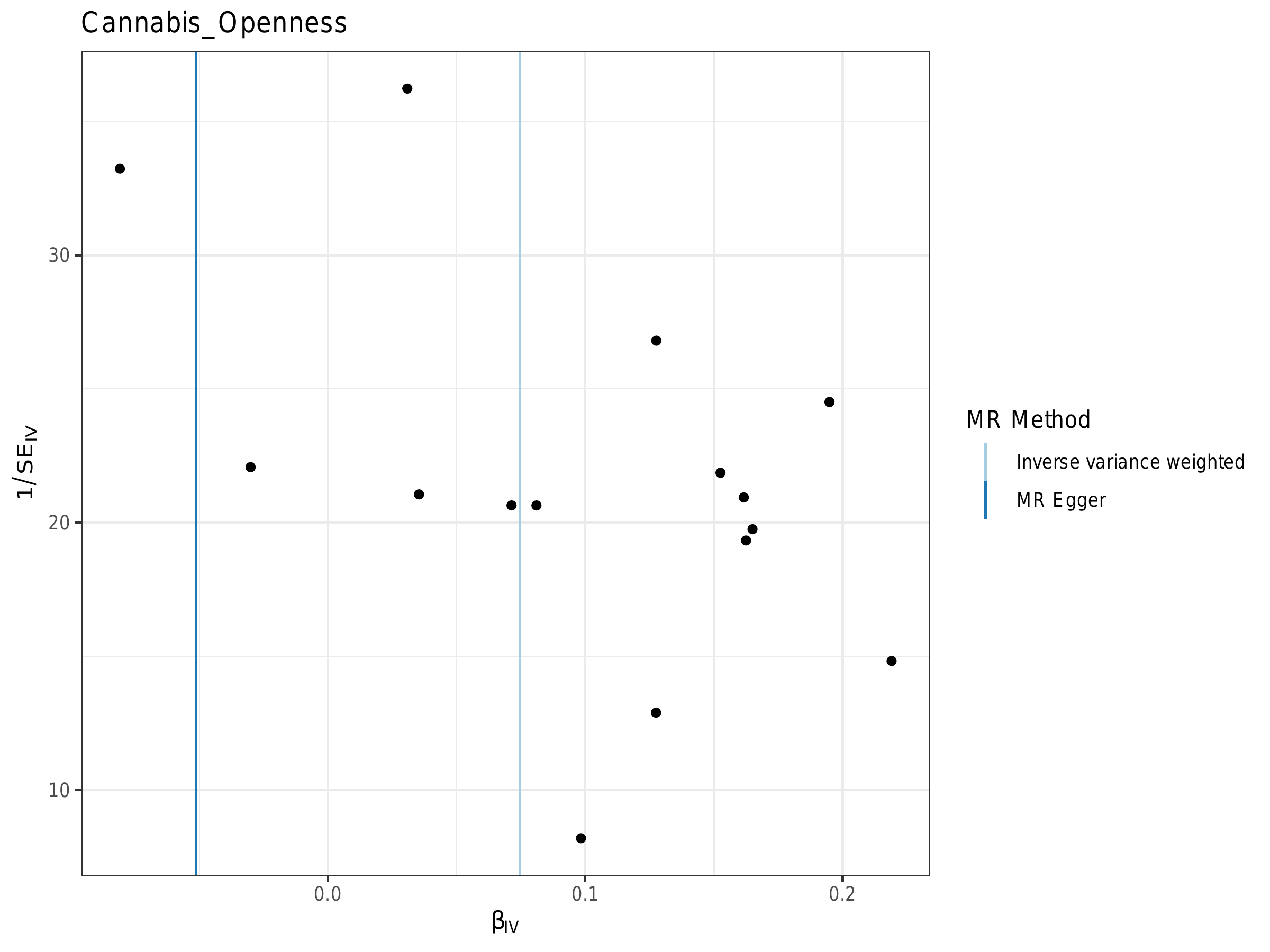

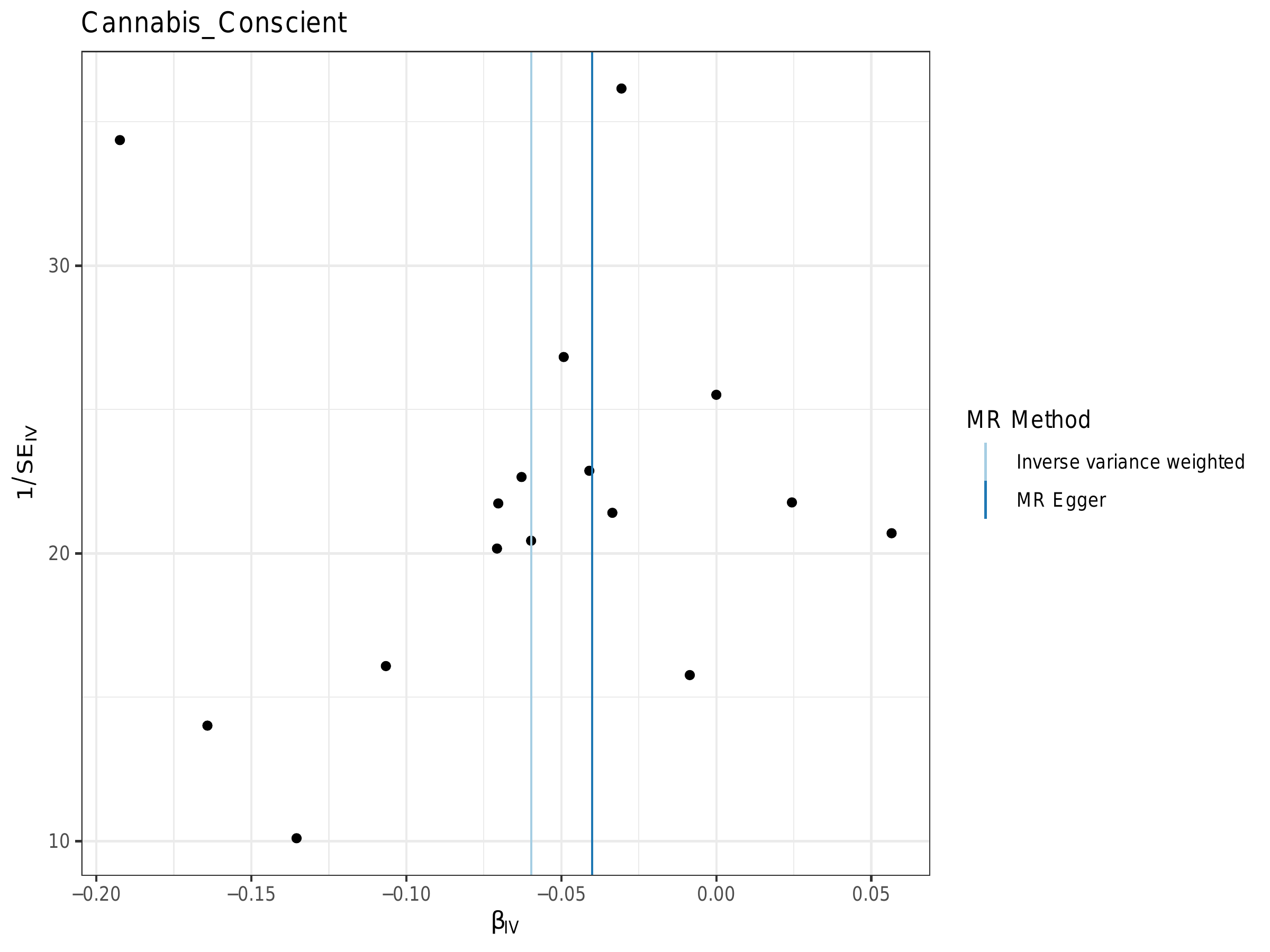

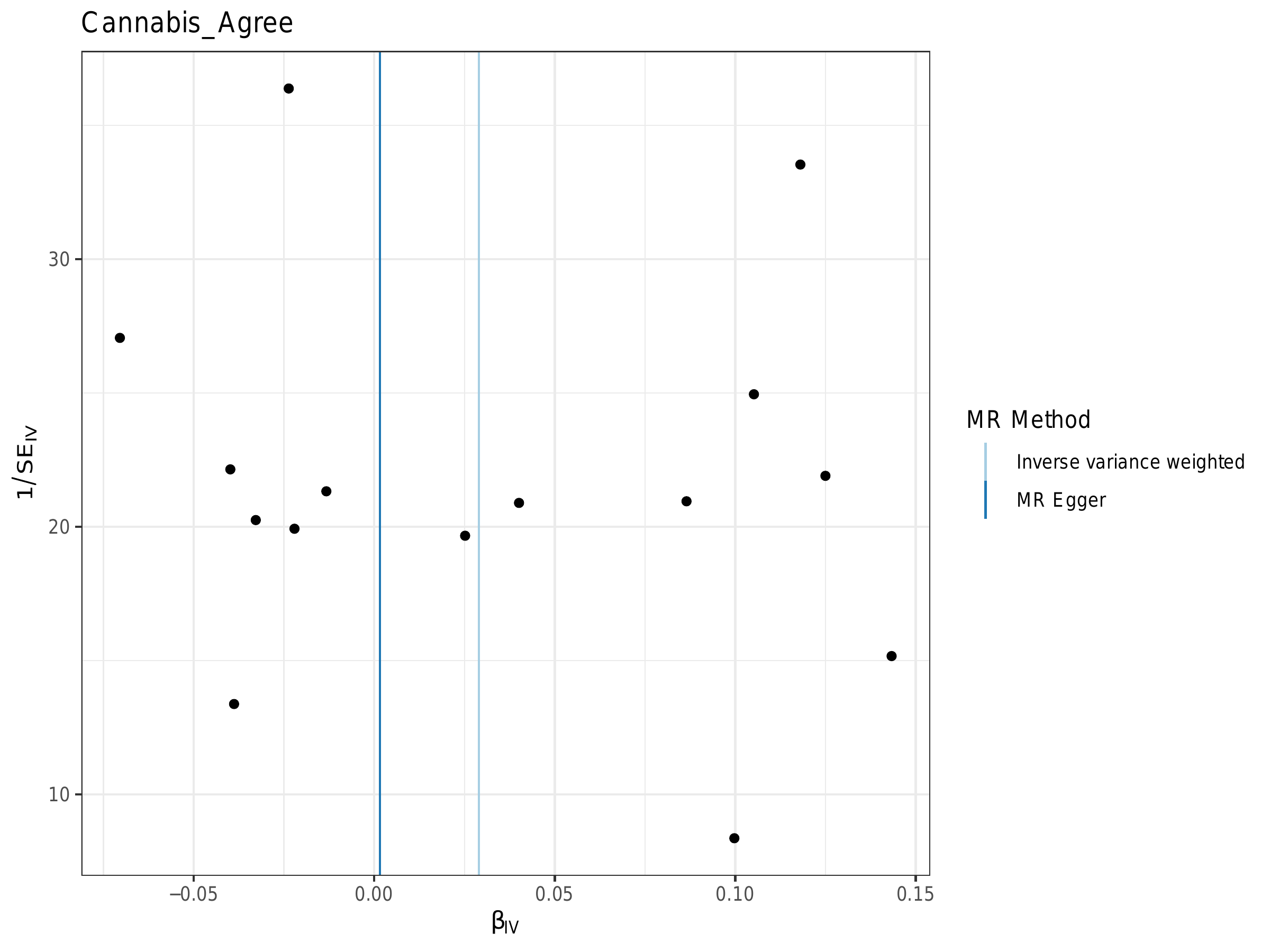

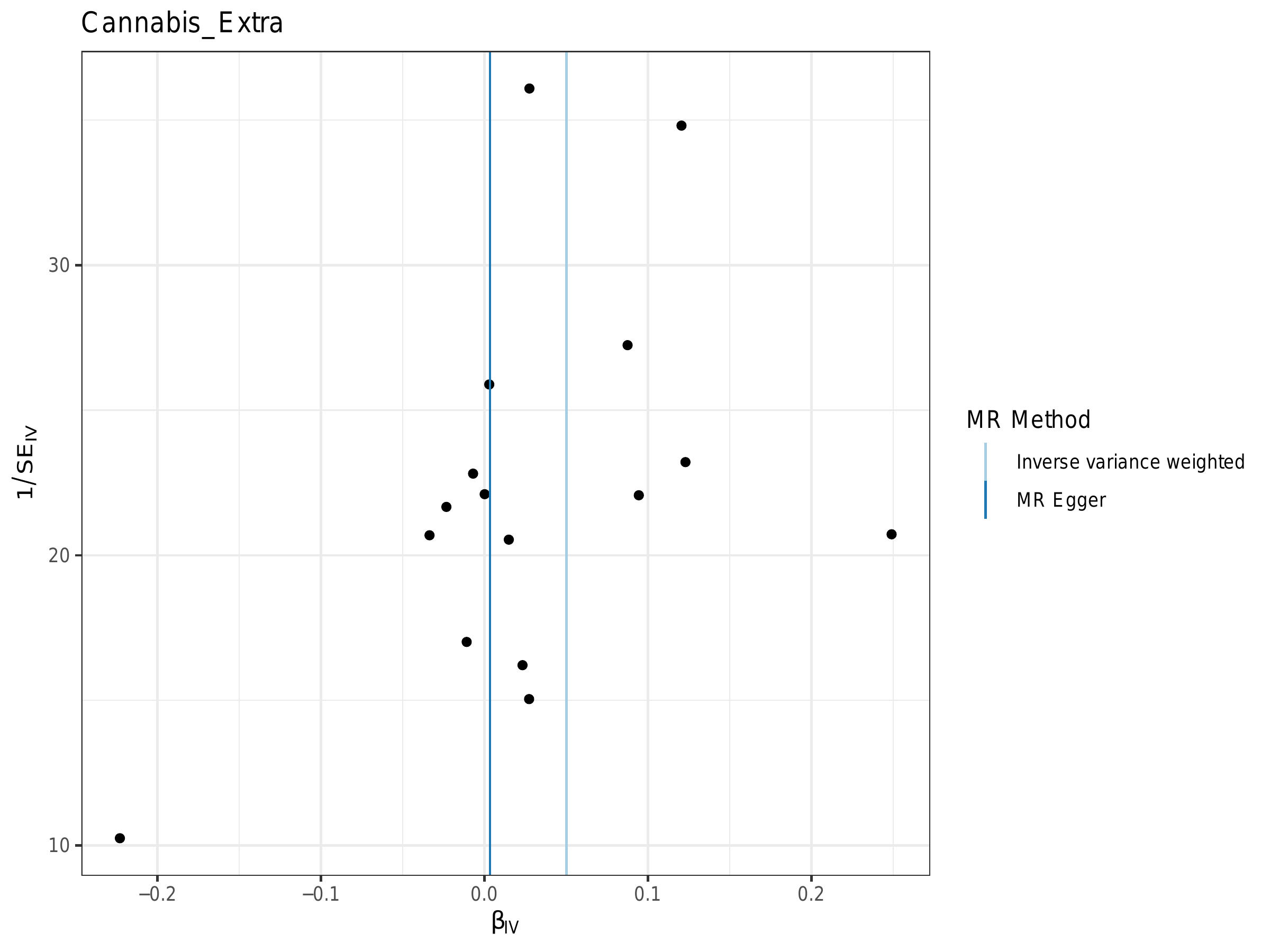

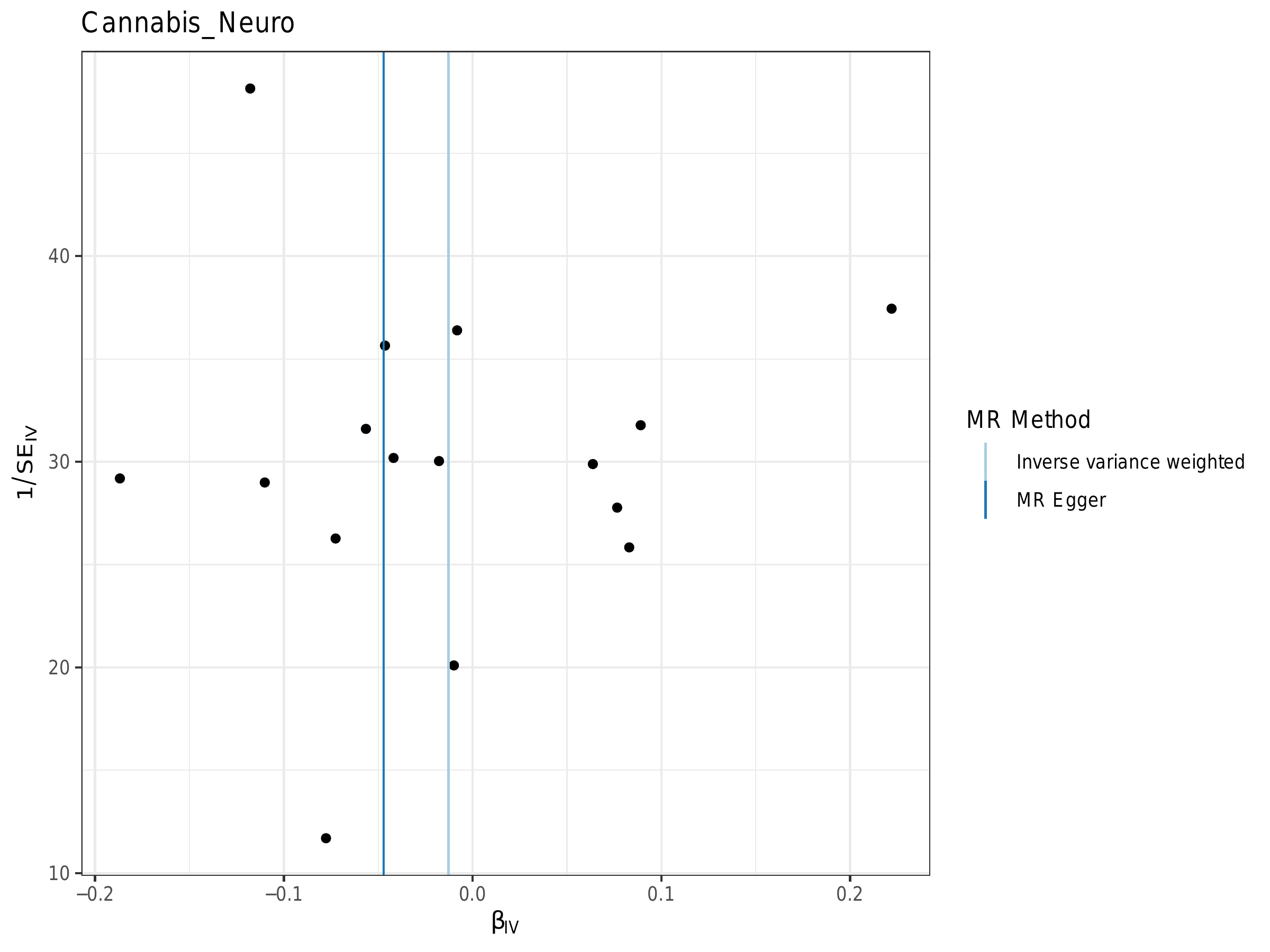

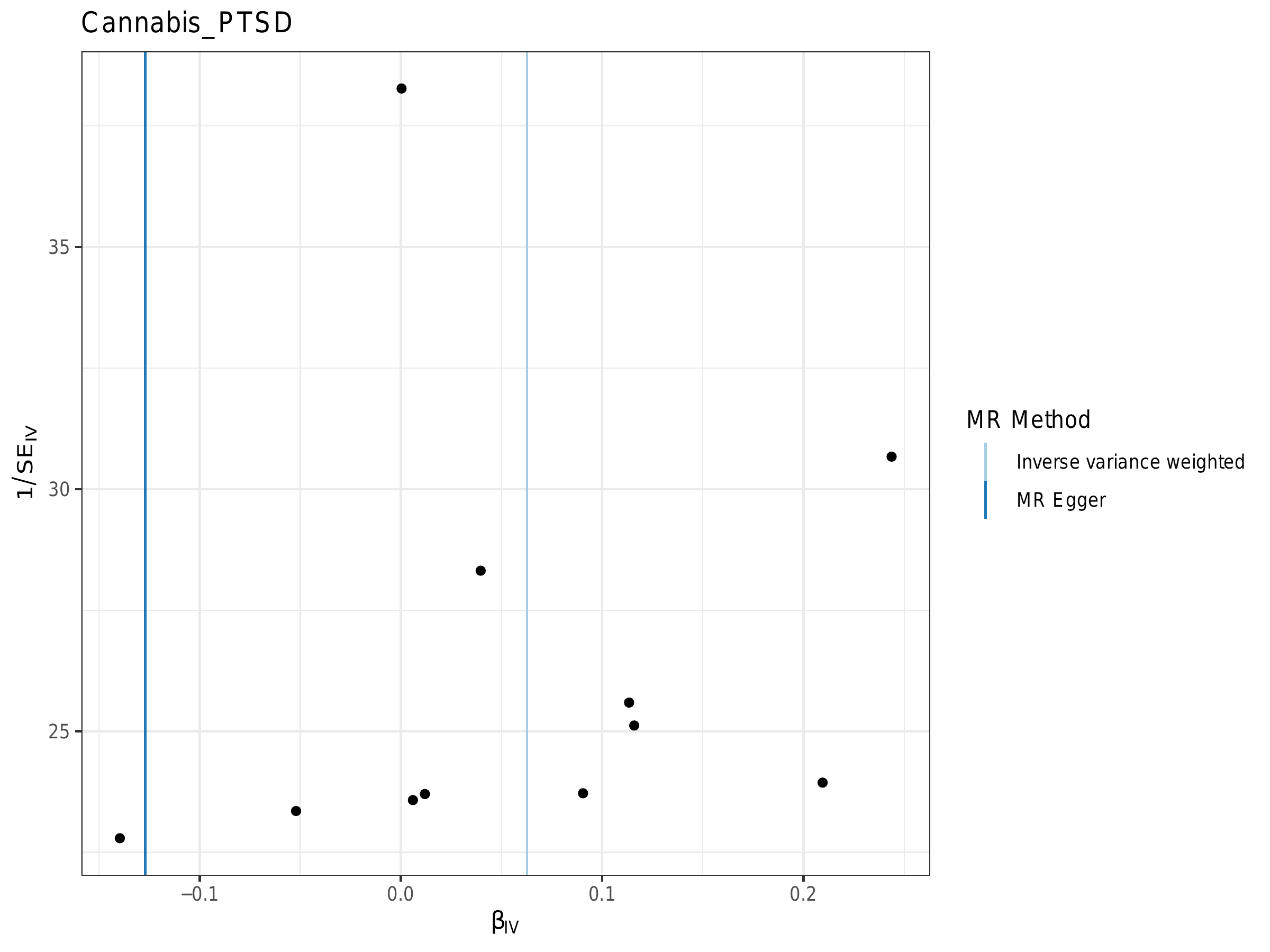

P2. Scatter plots
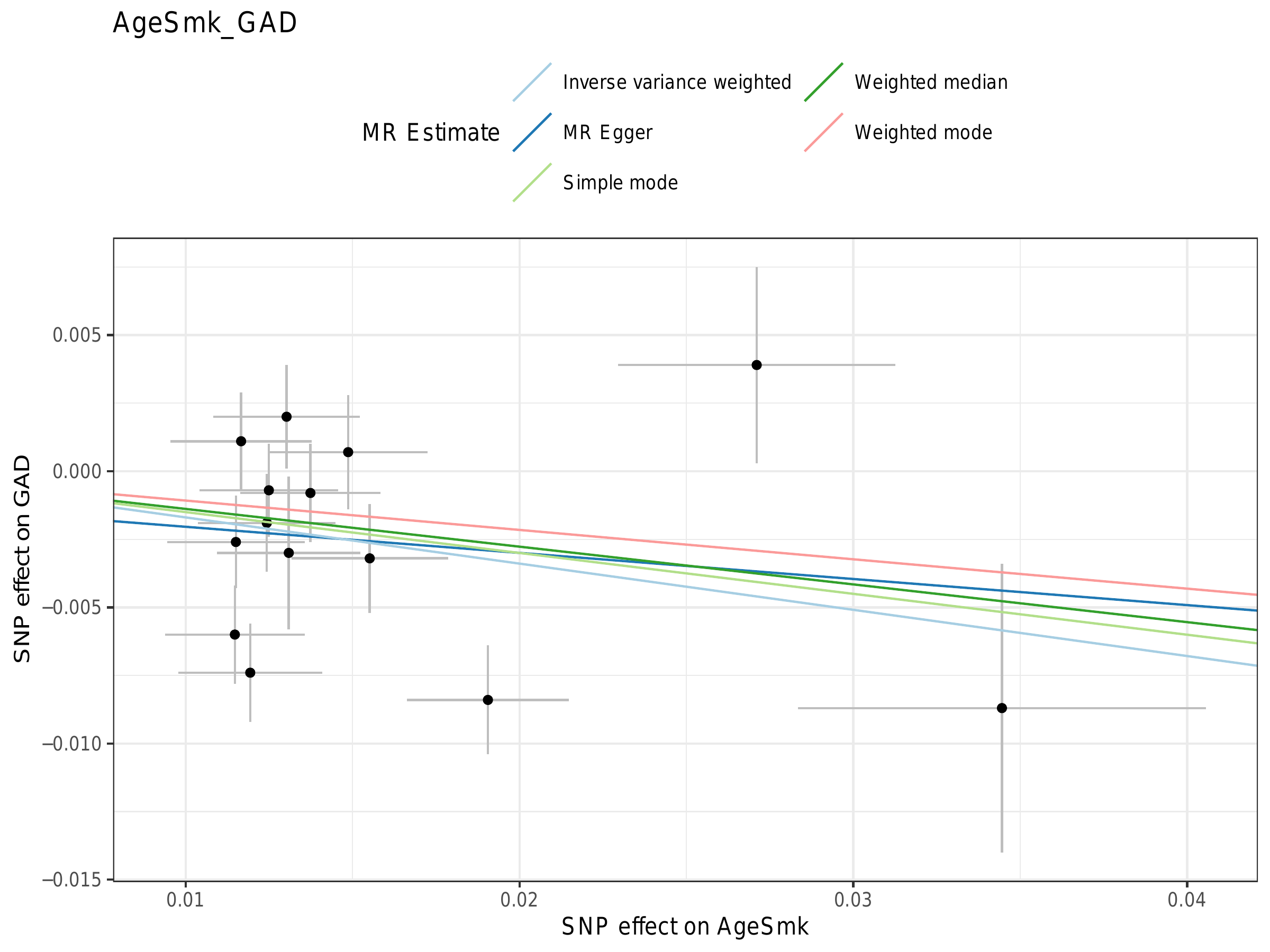

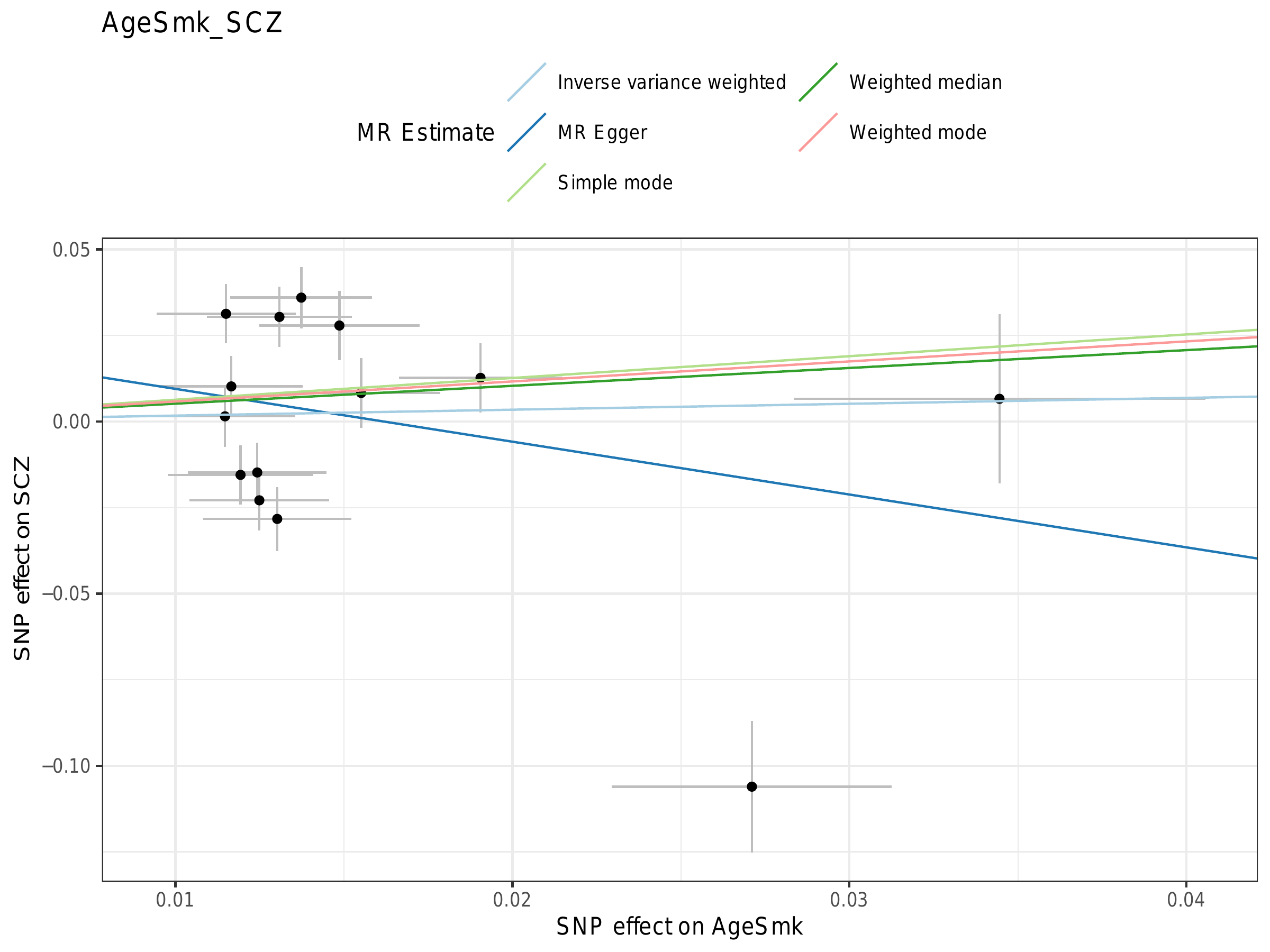

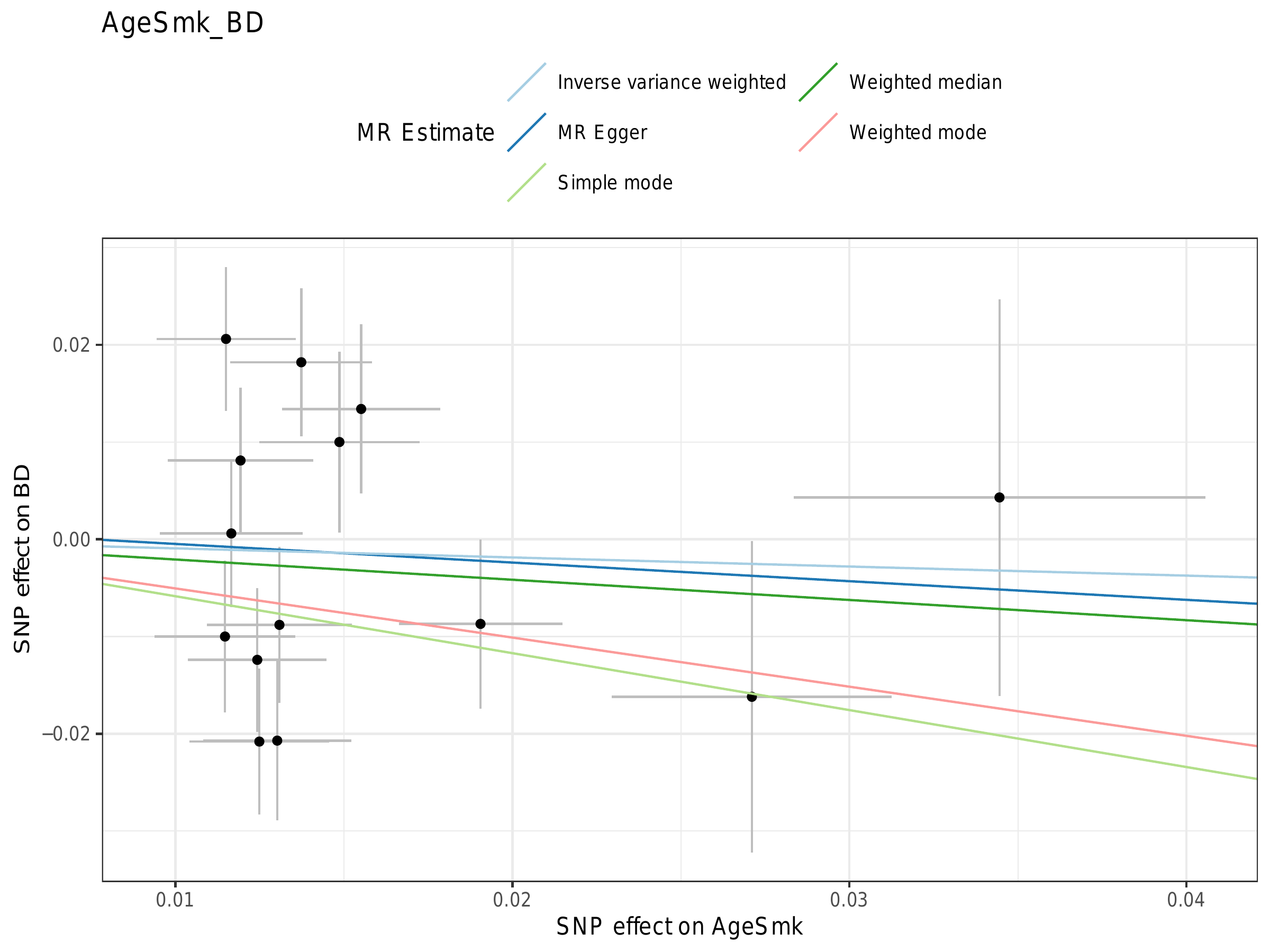

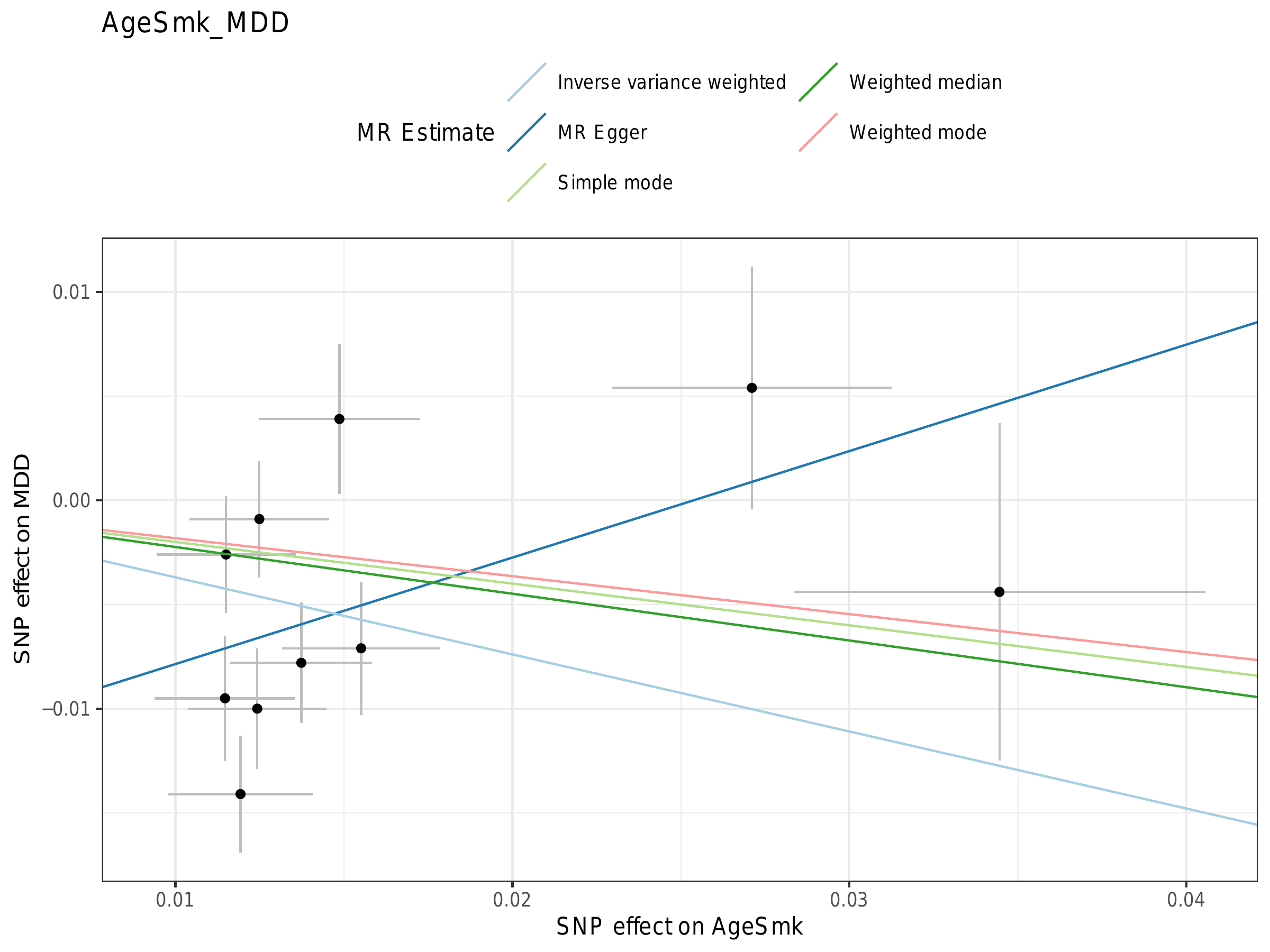

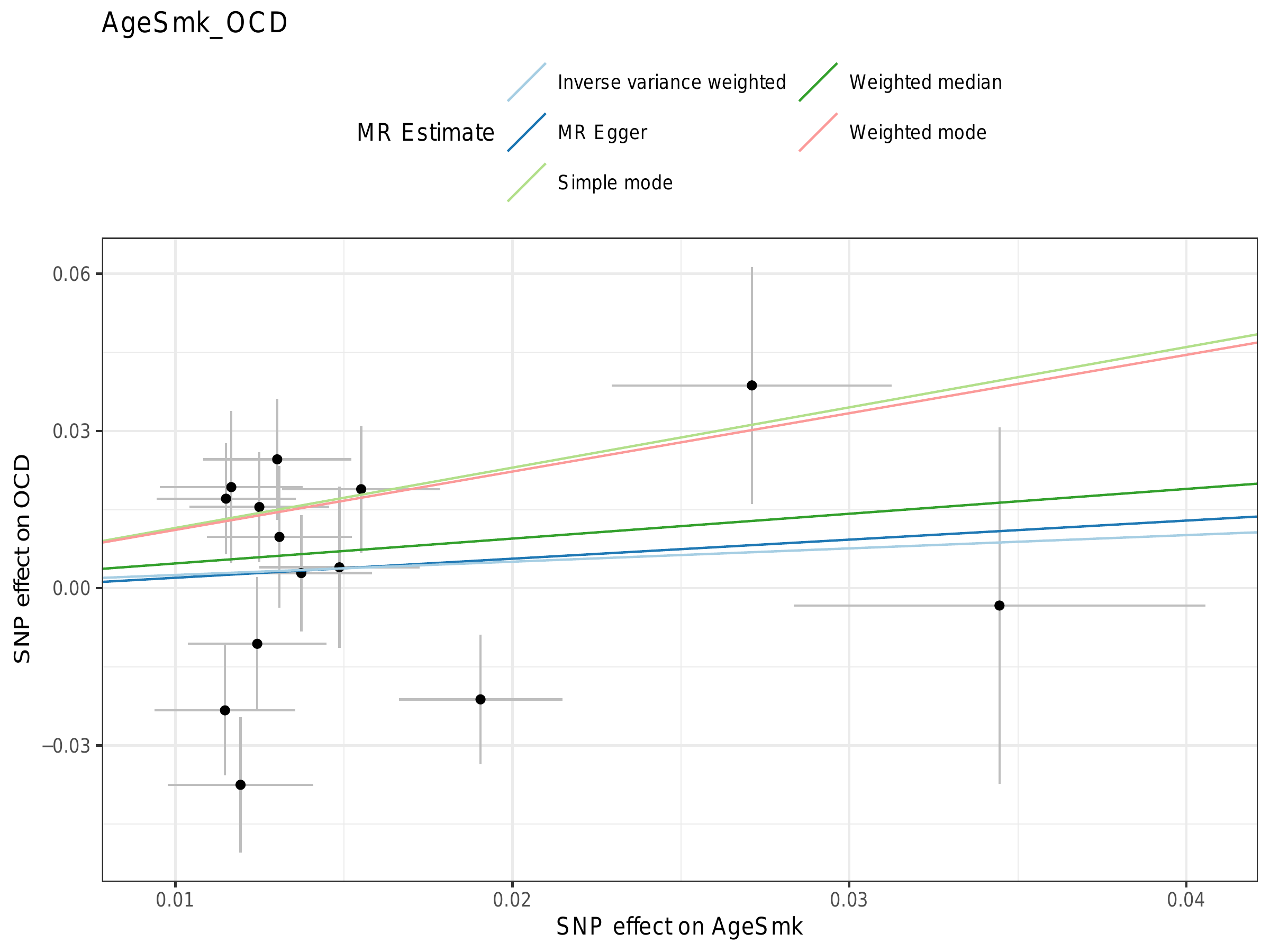

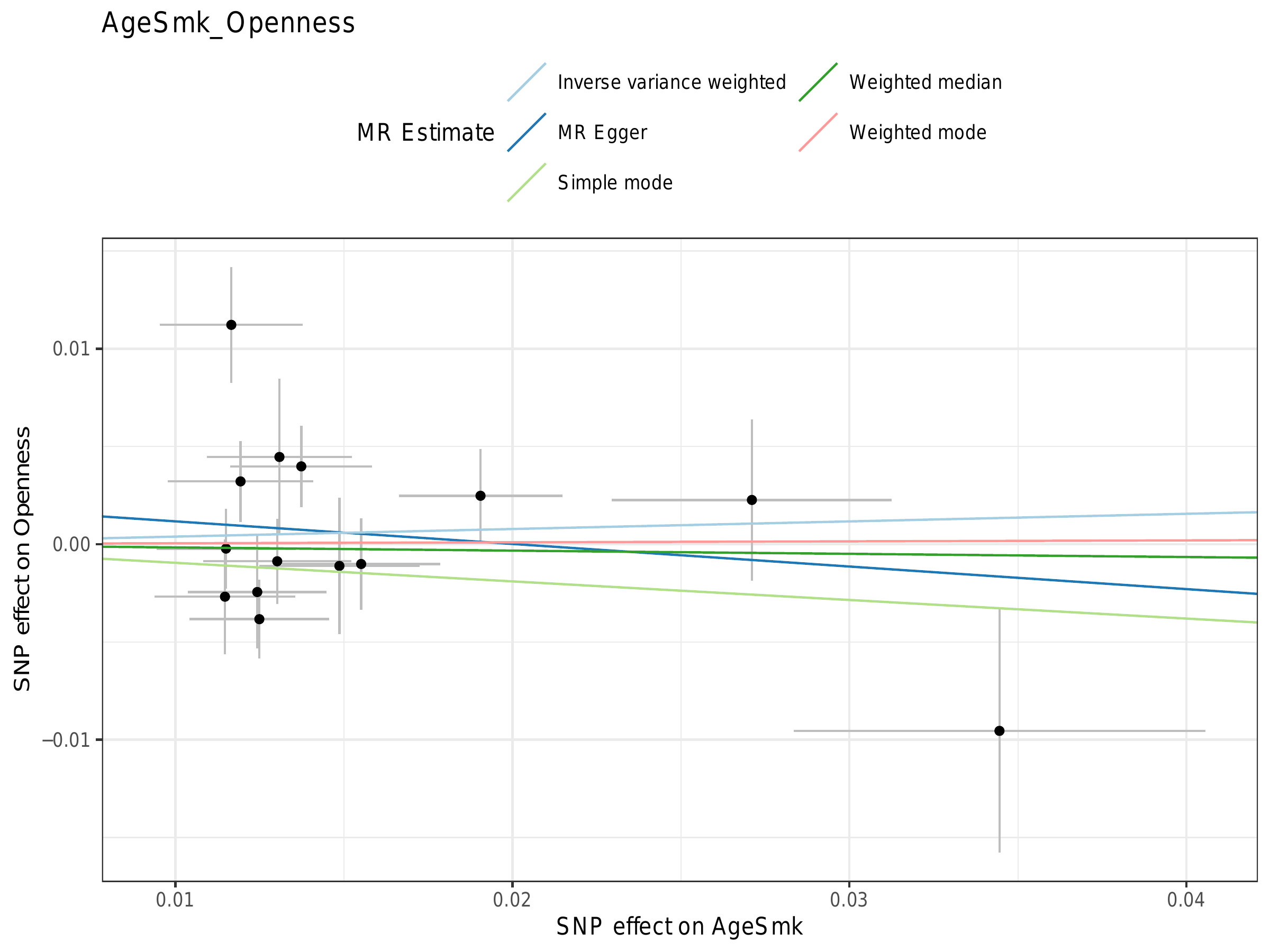

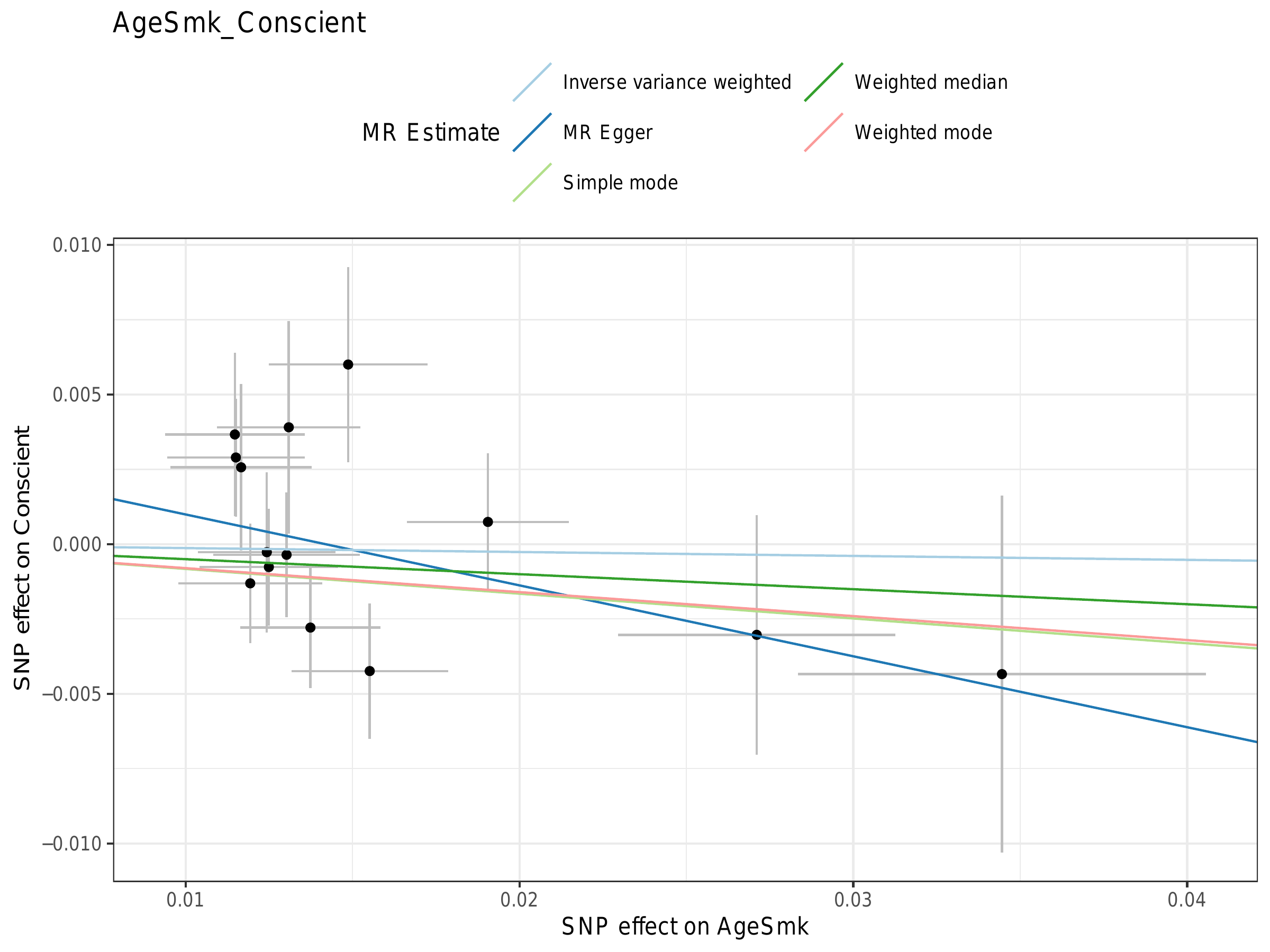

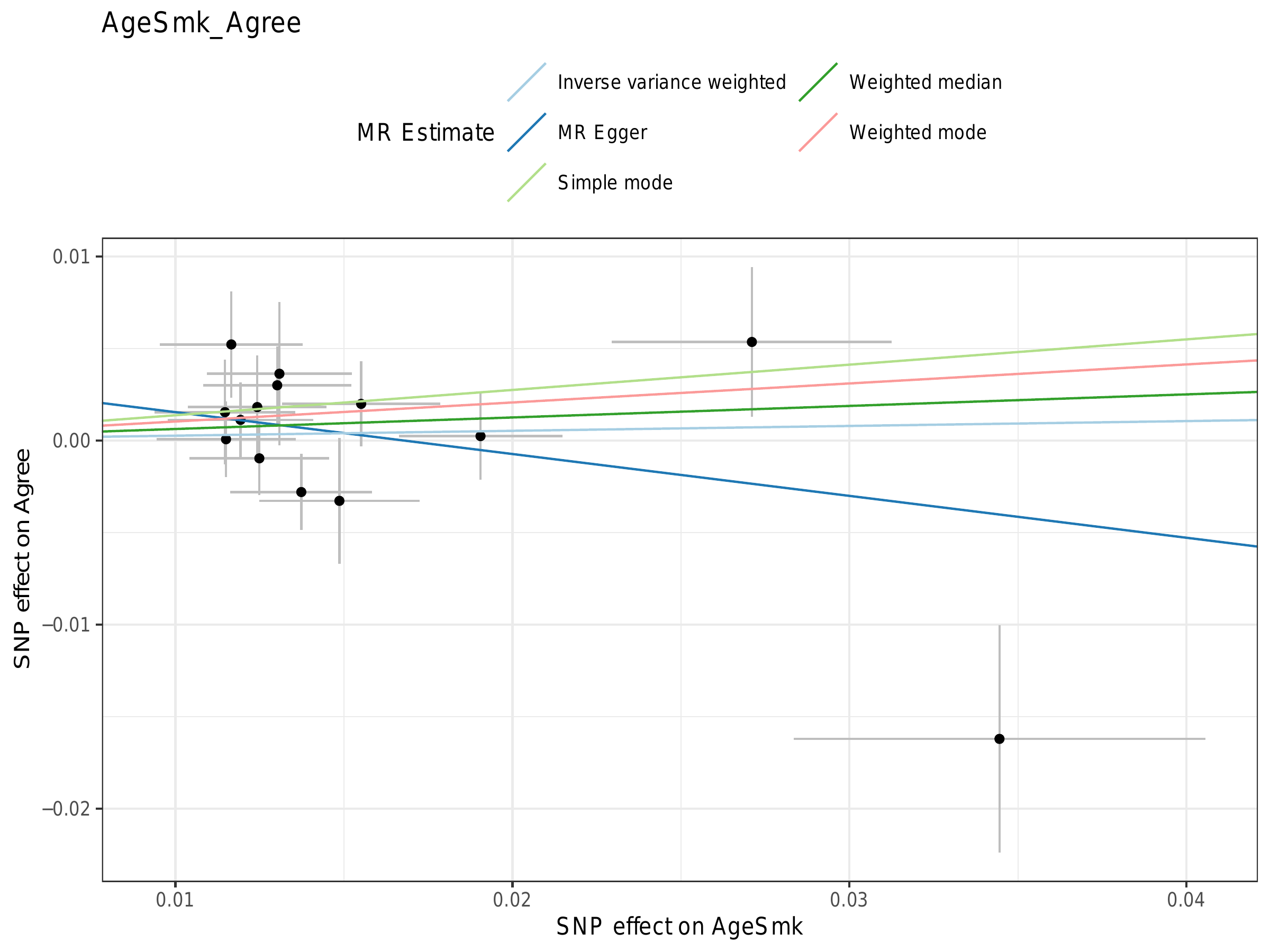

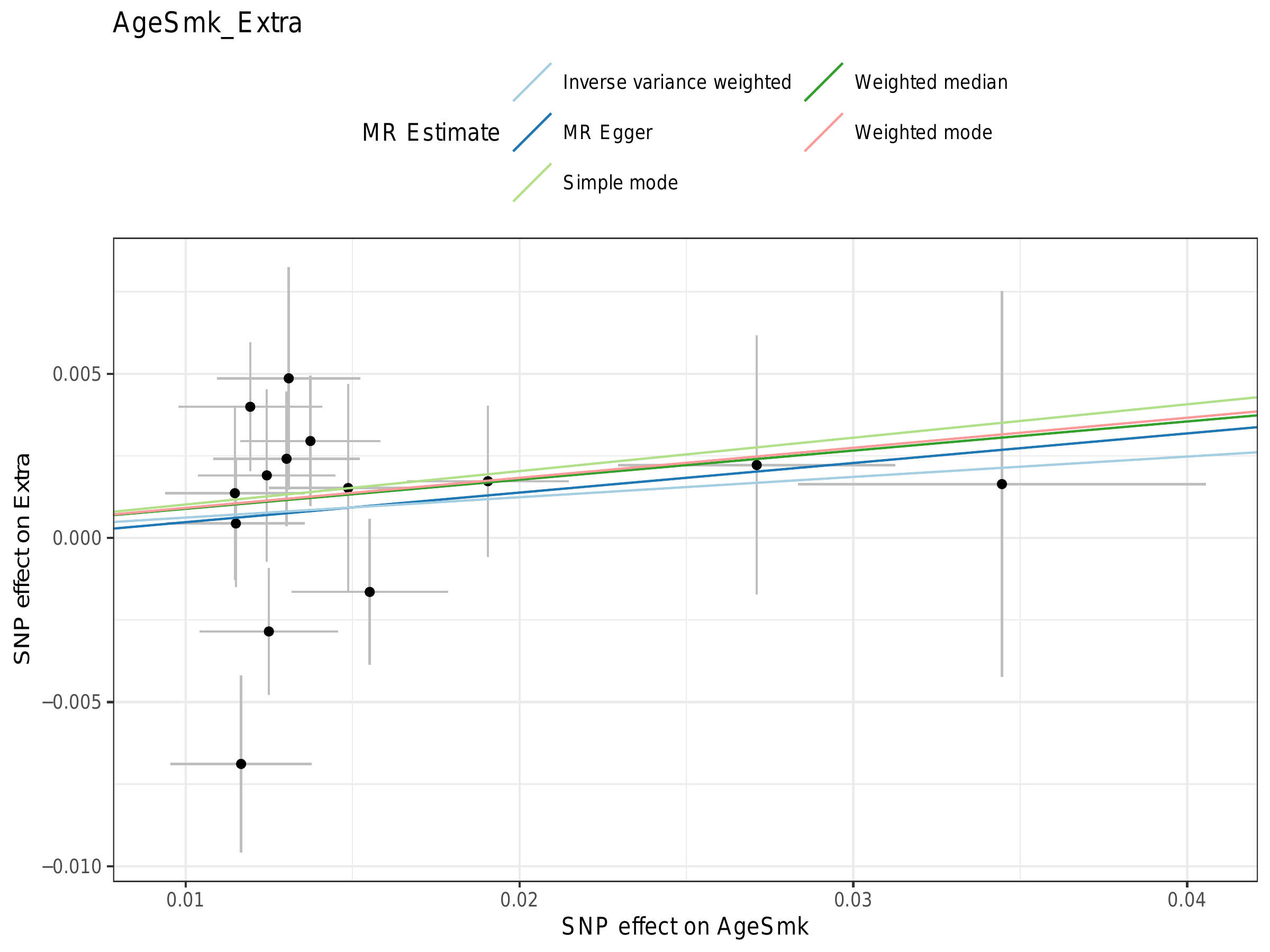

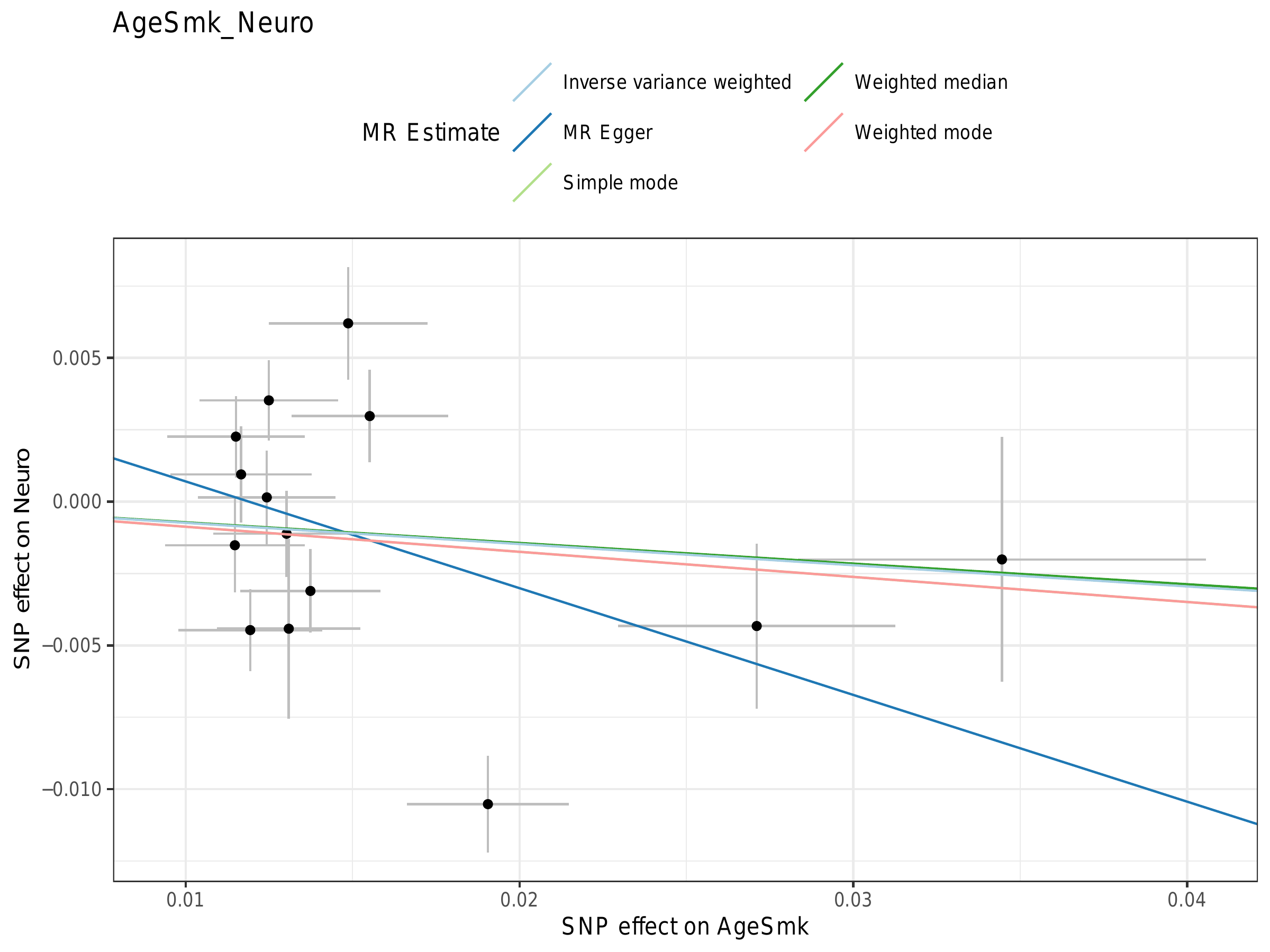

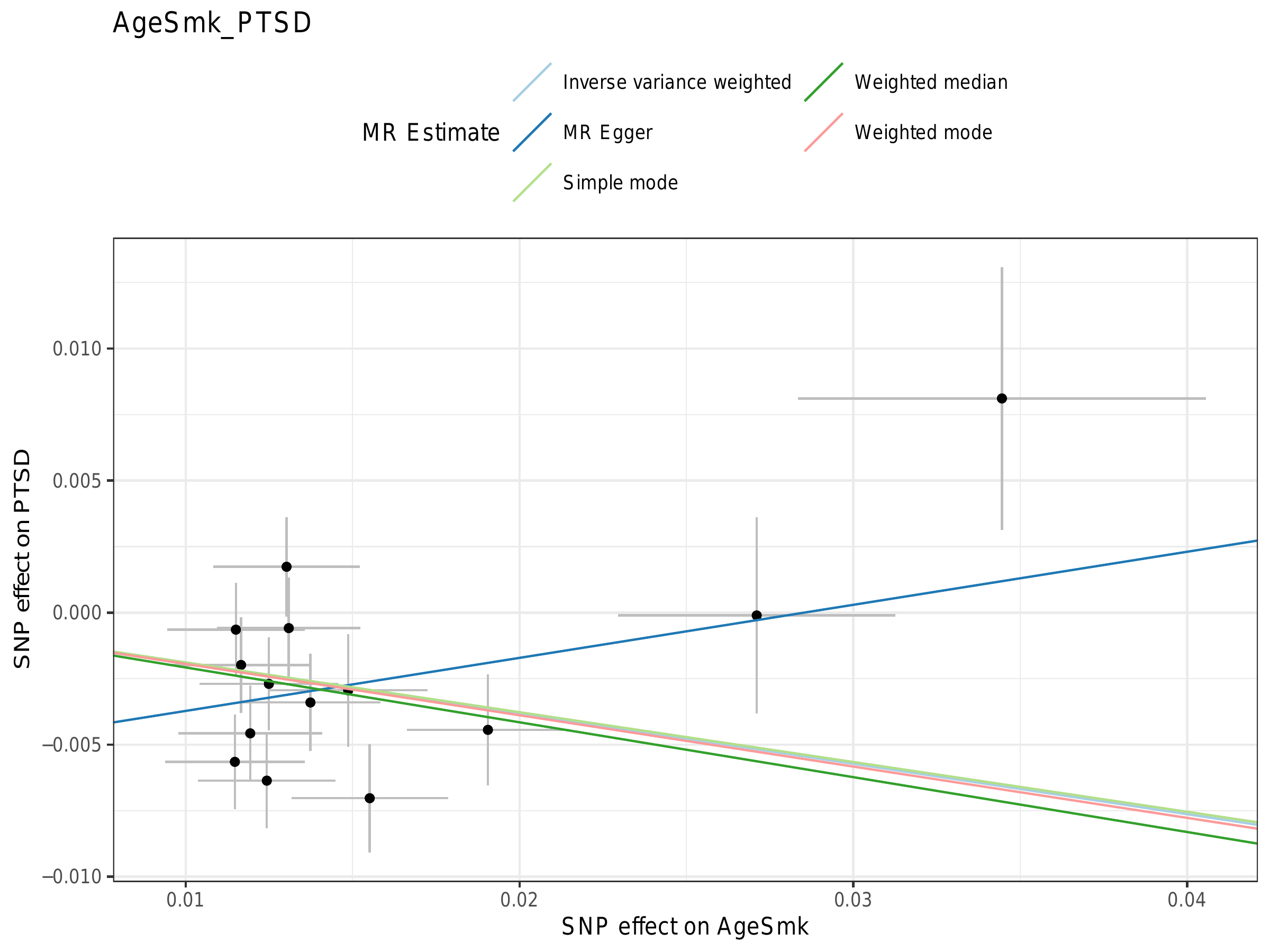

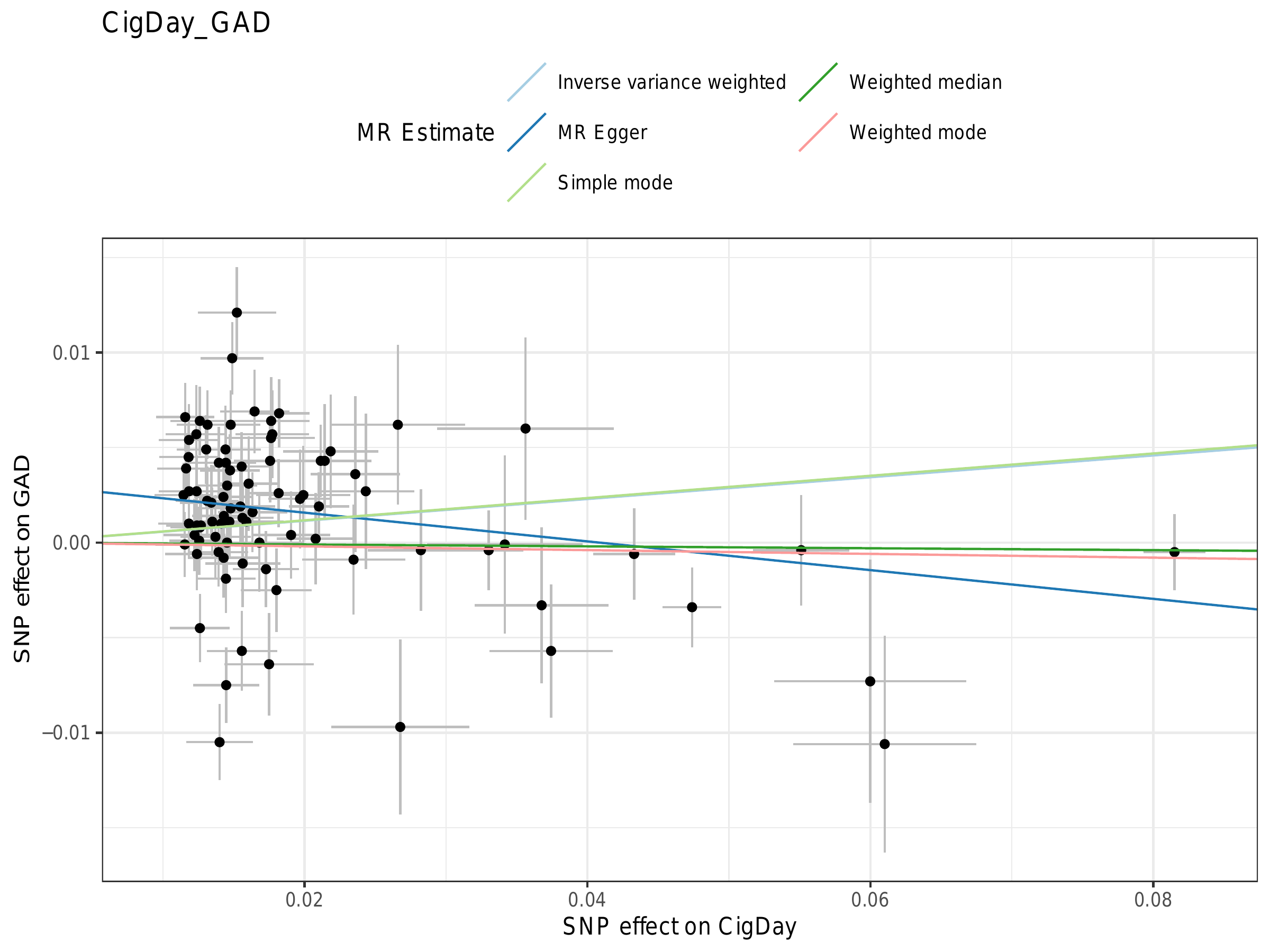

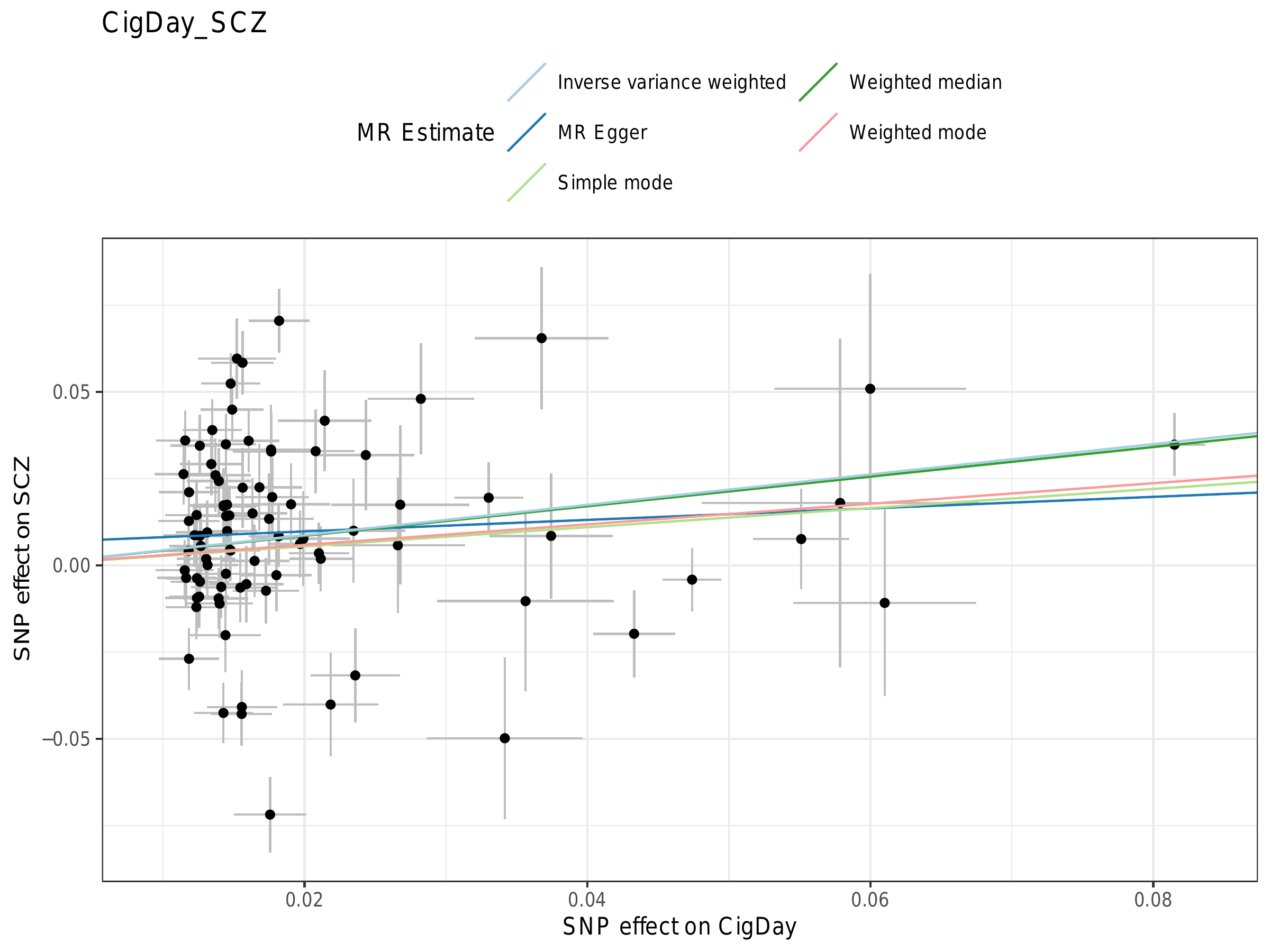

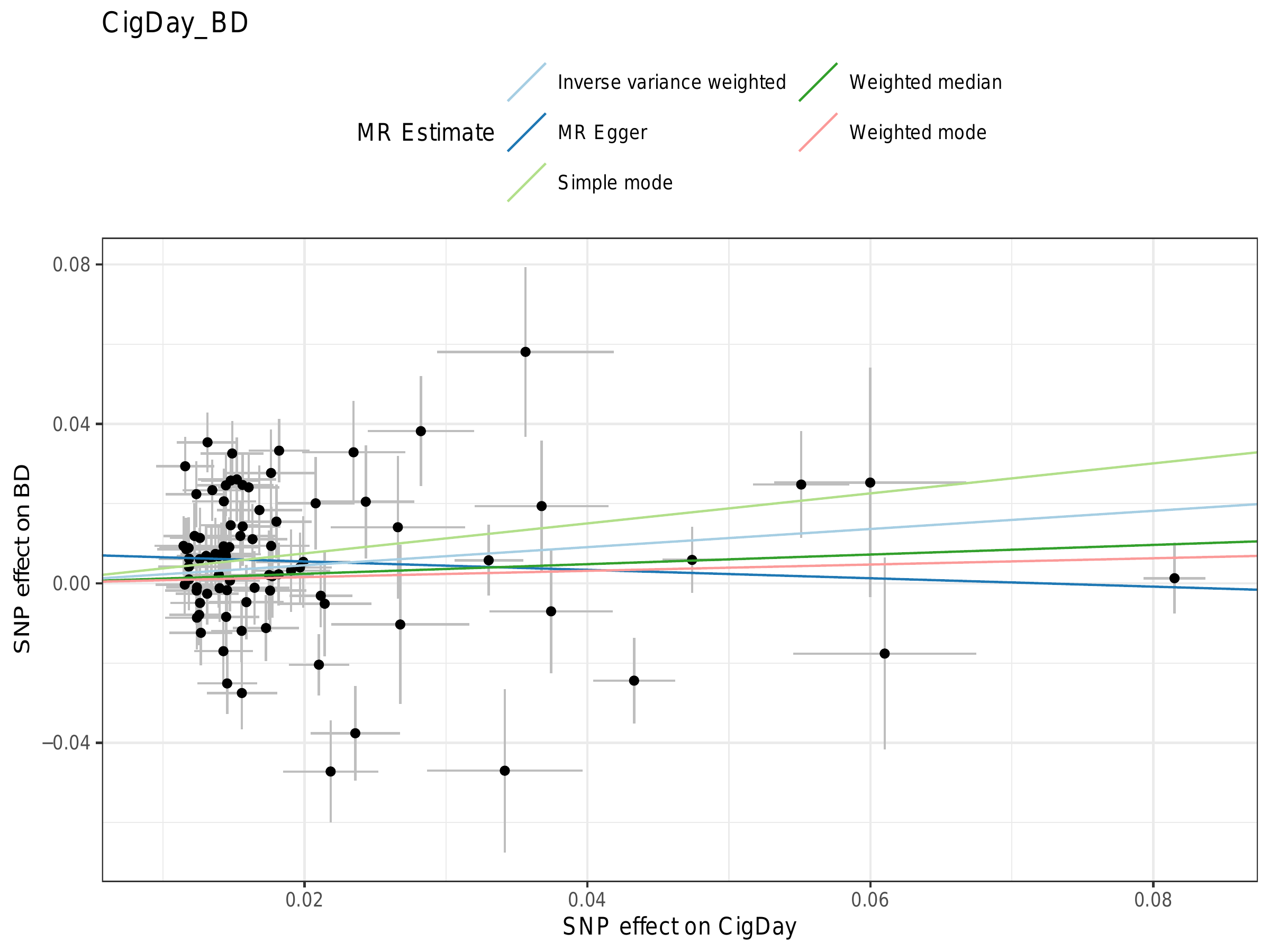

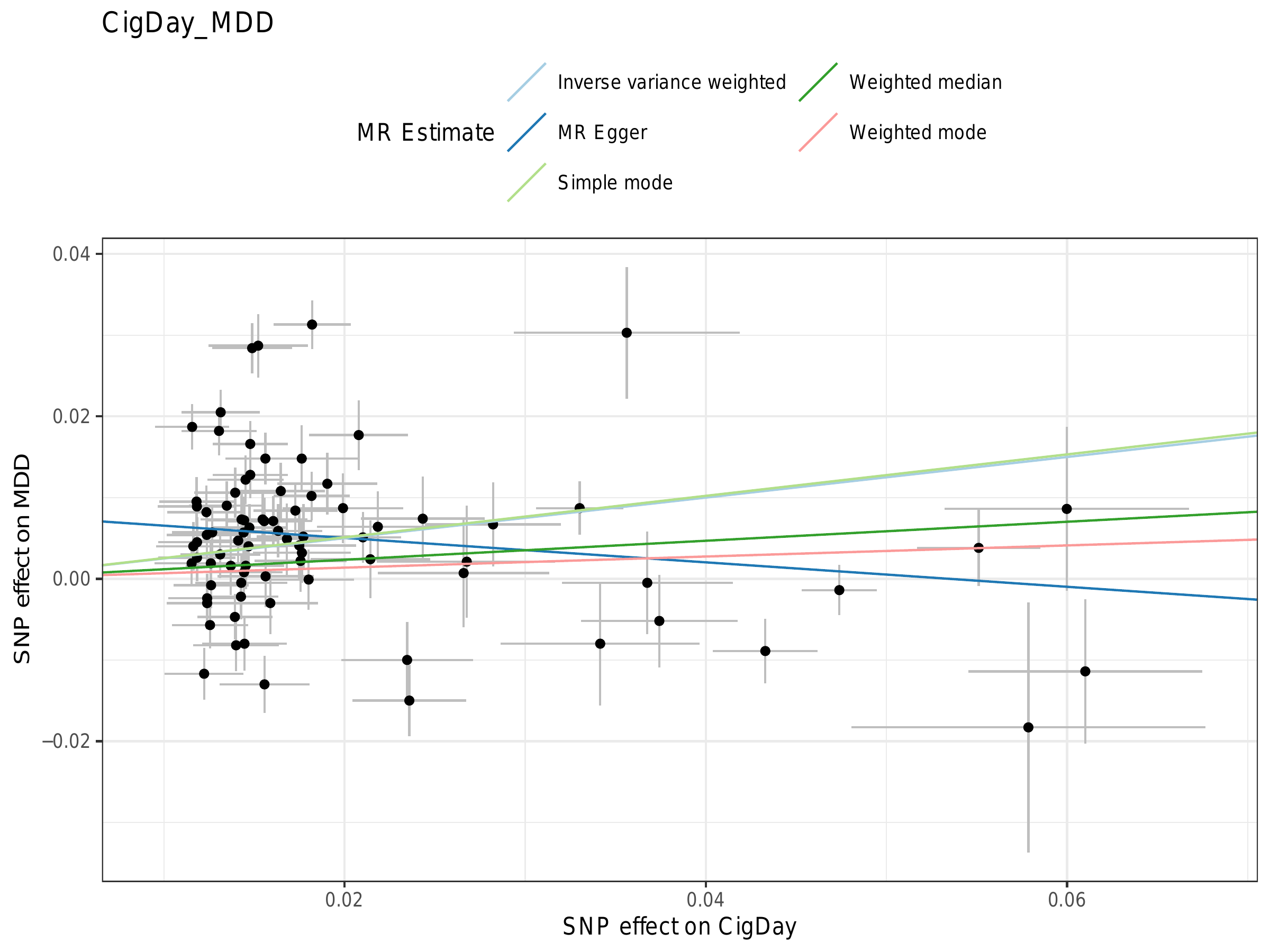

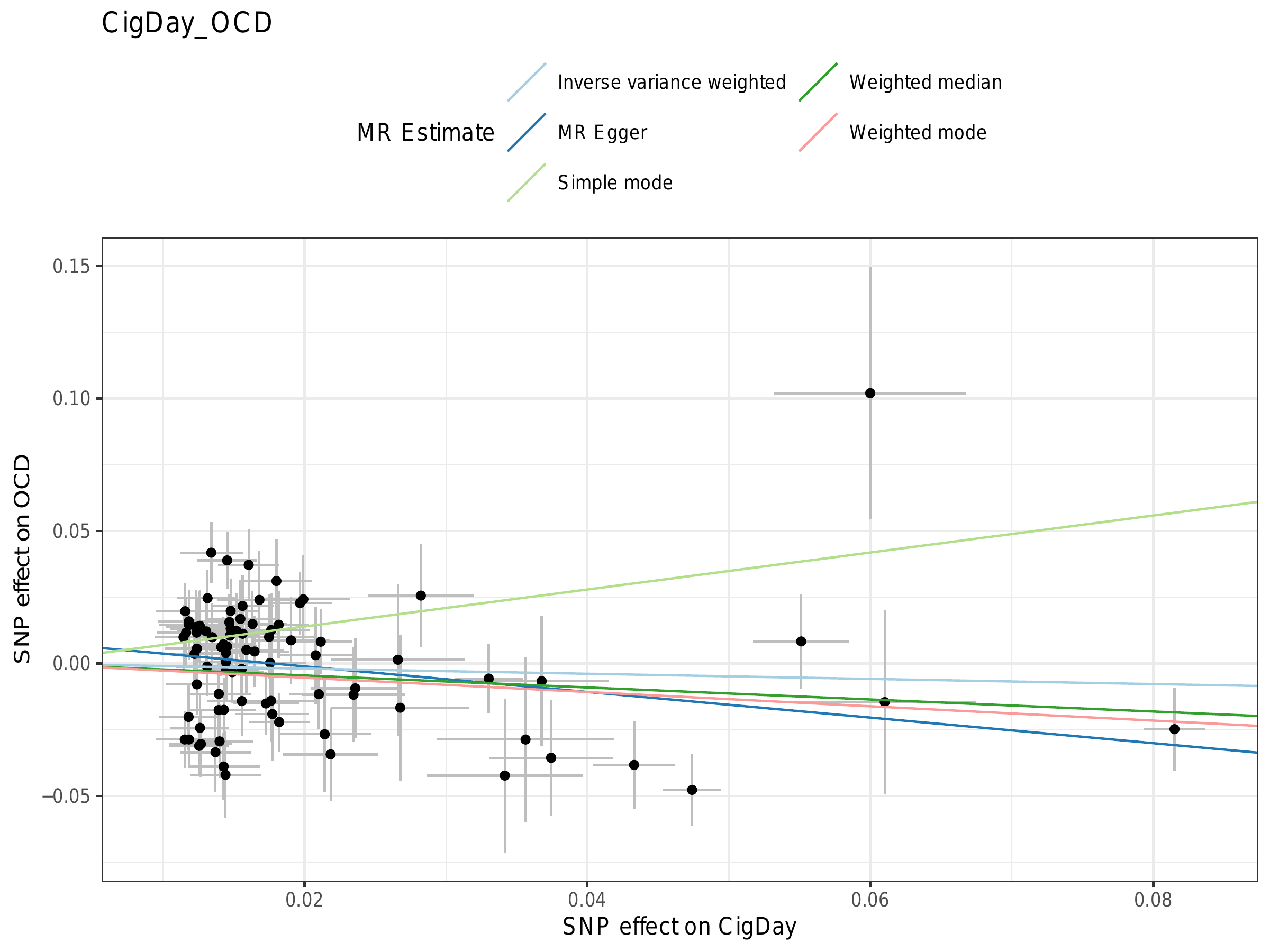

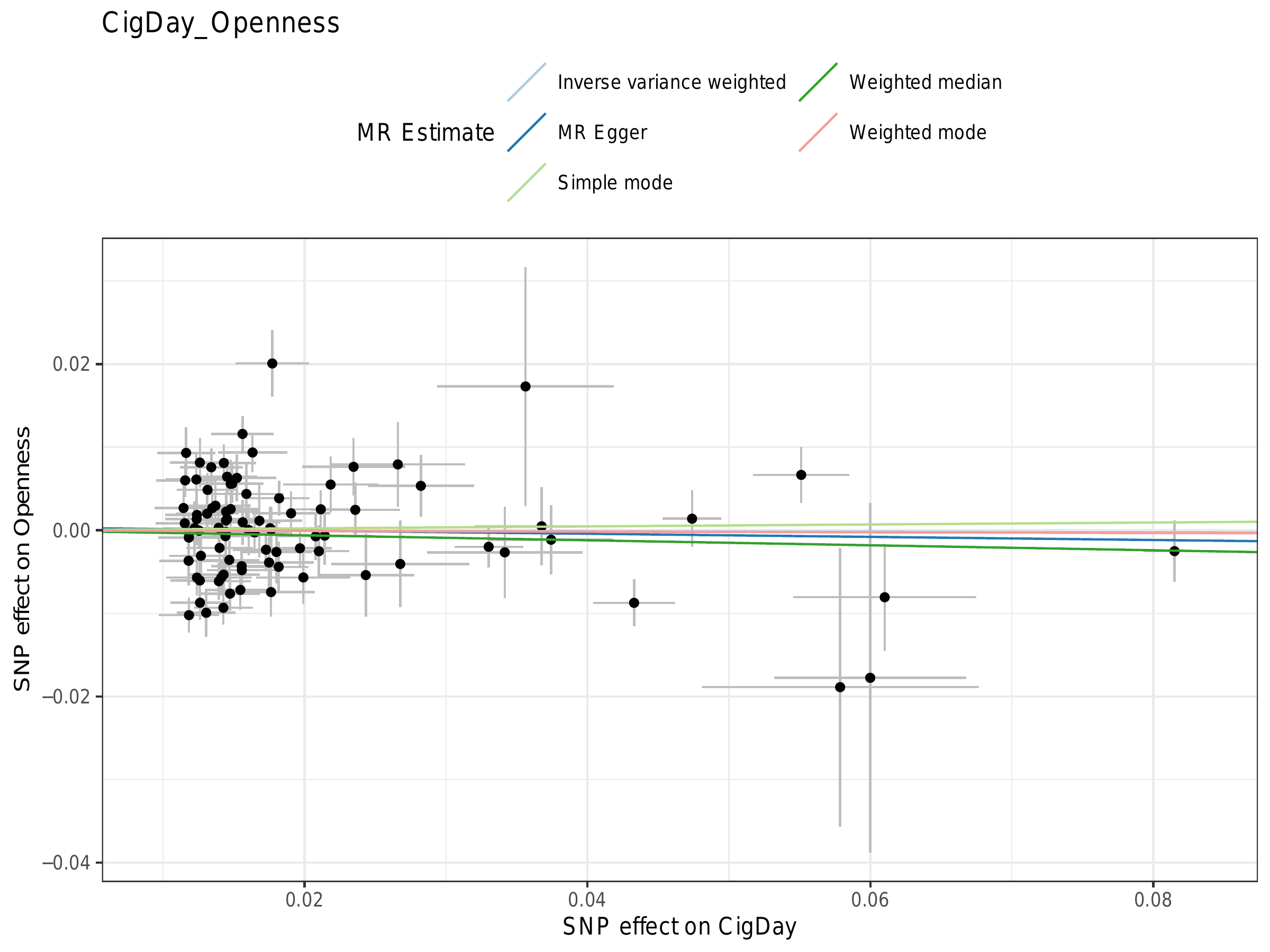

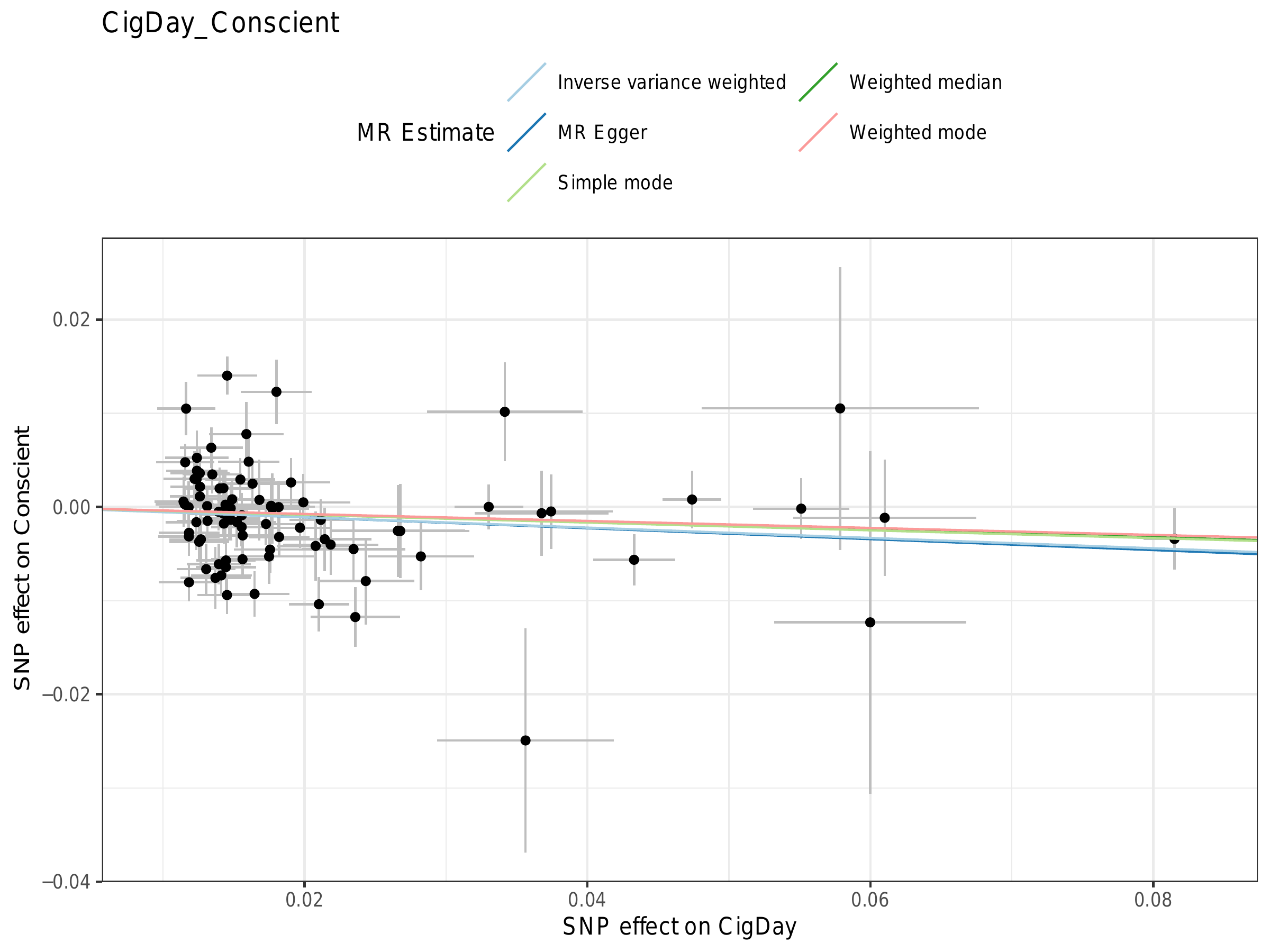

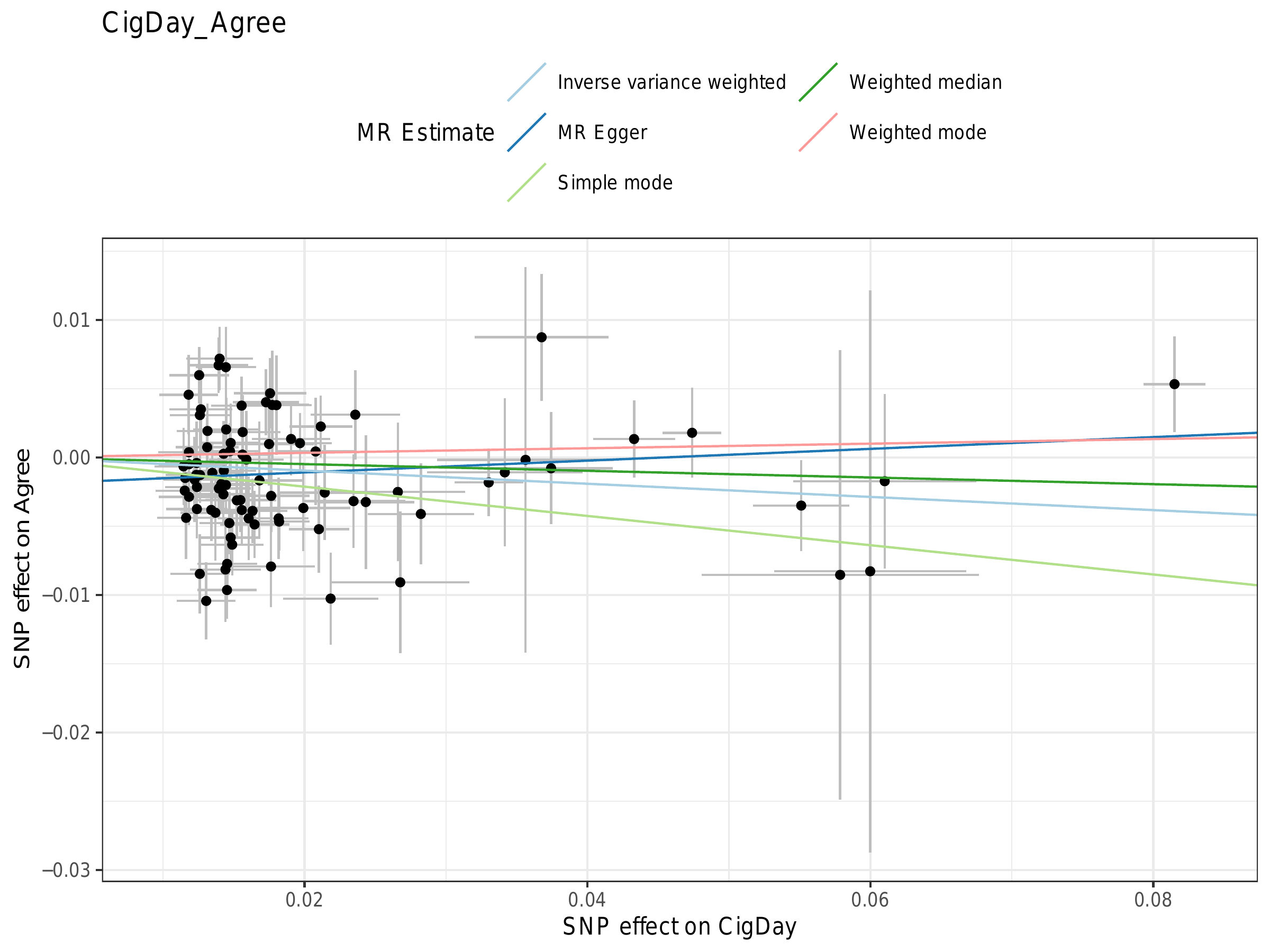

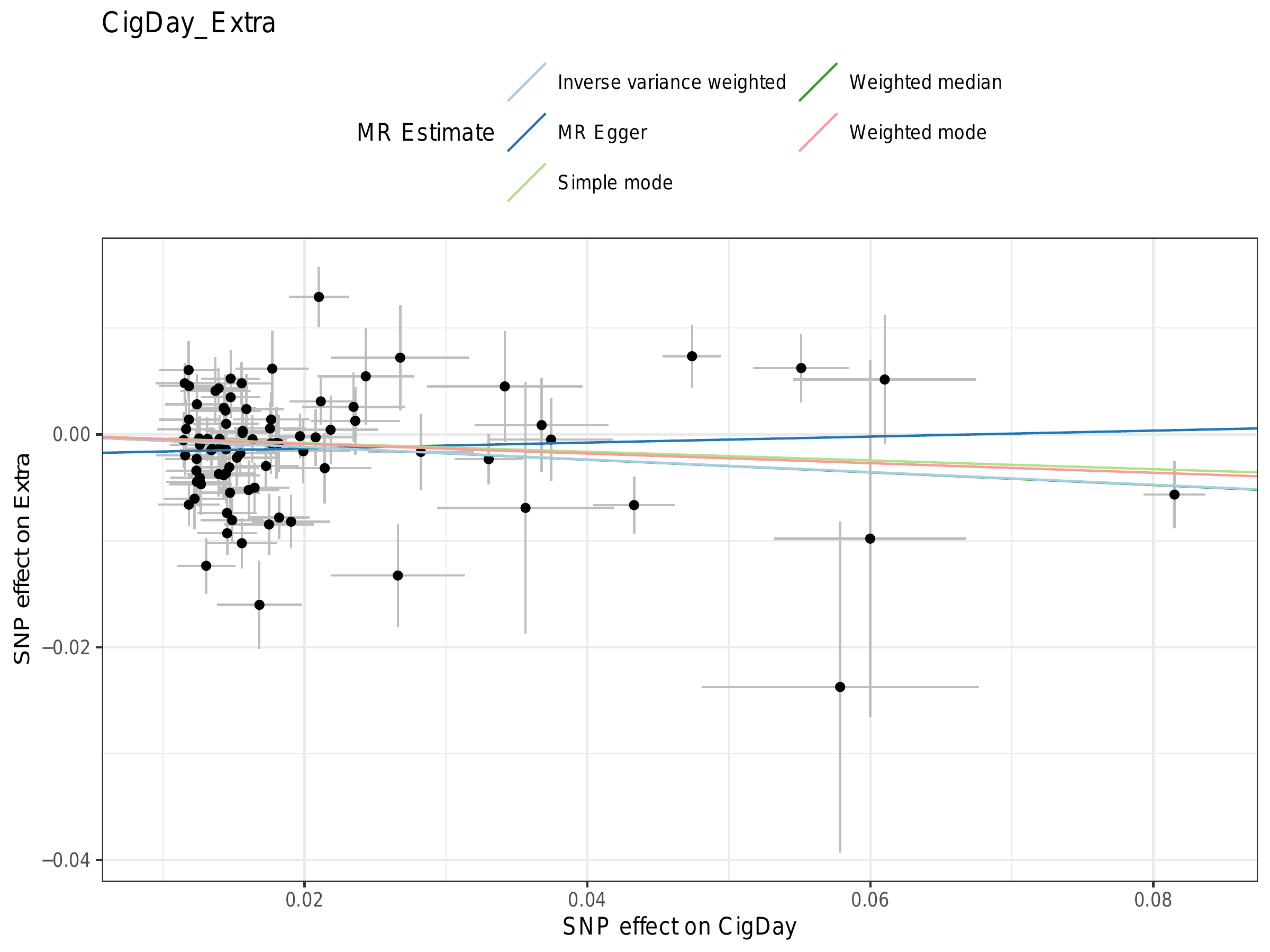

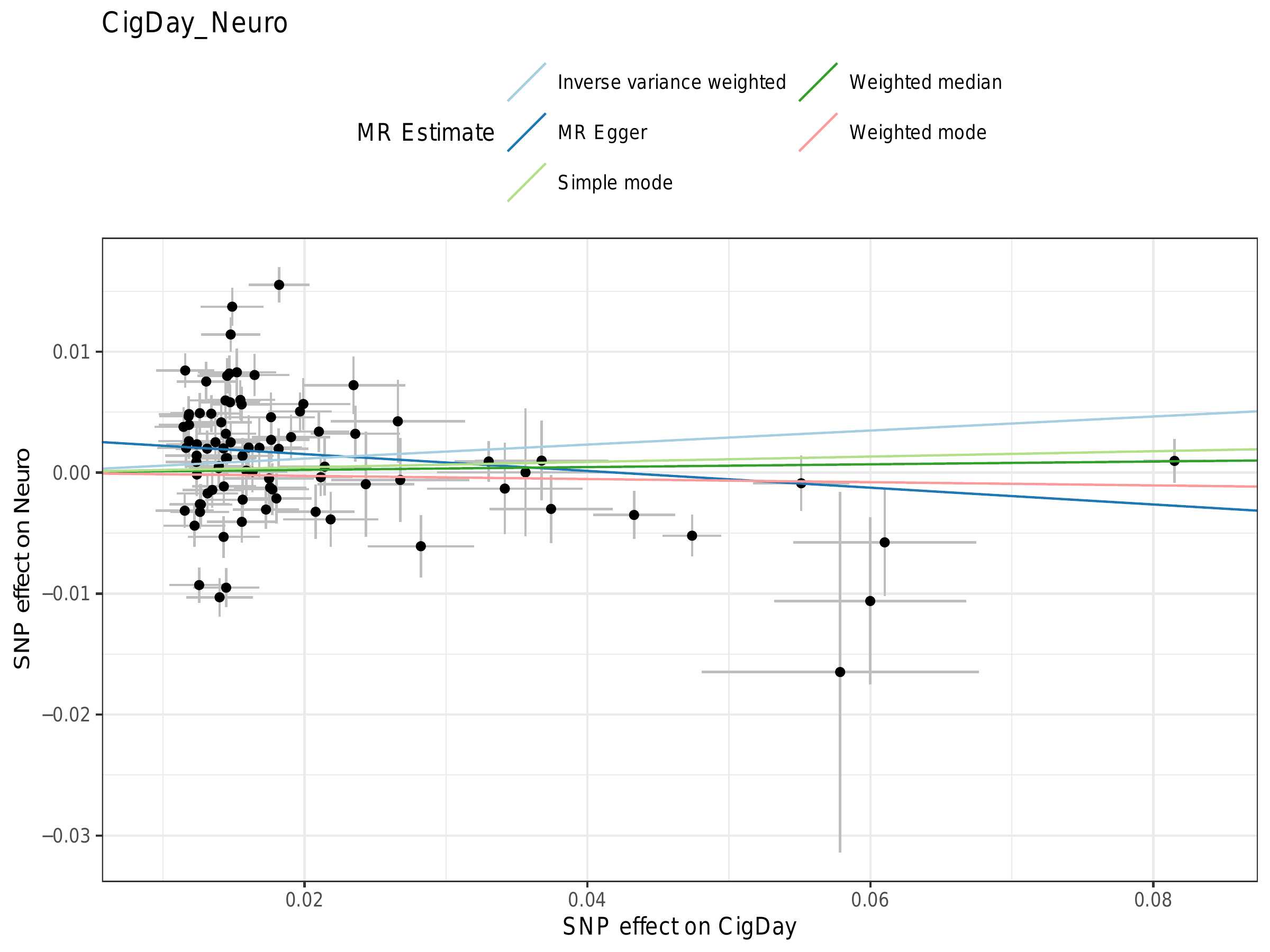

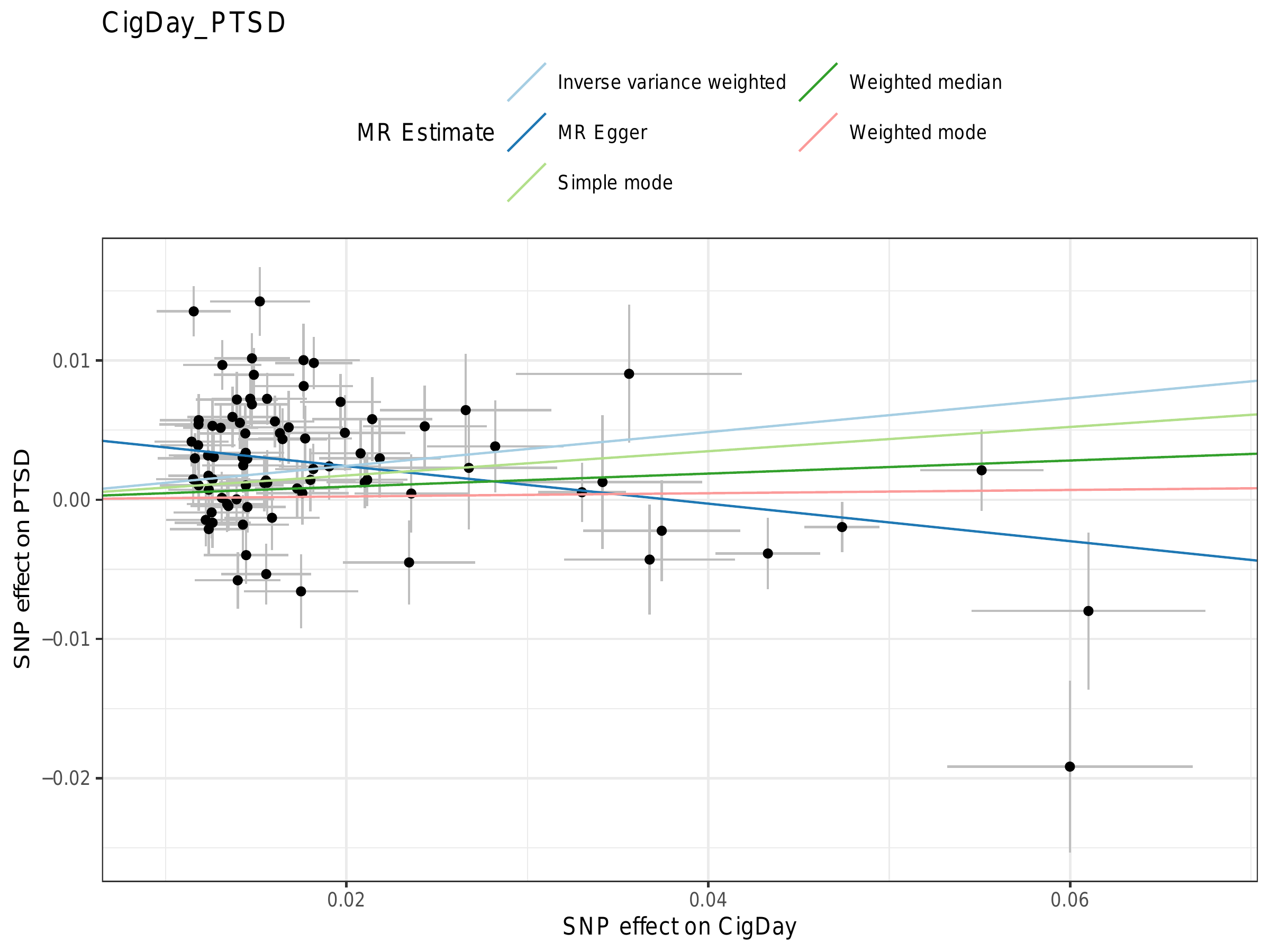

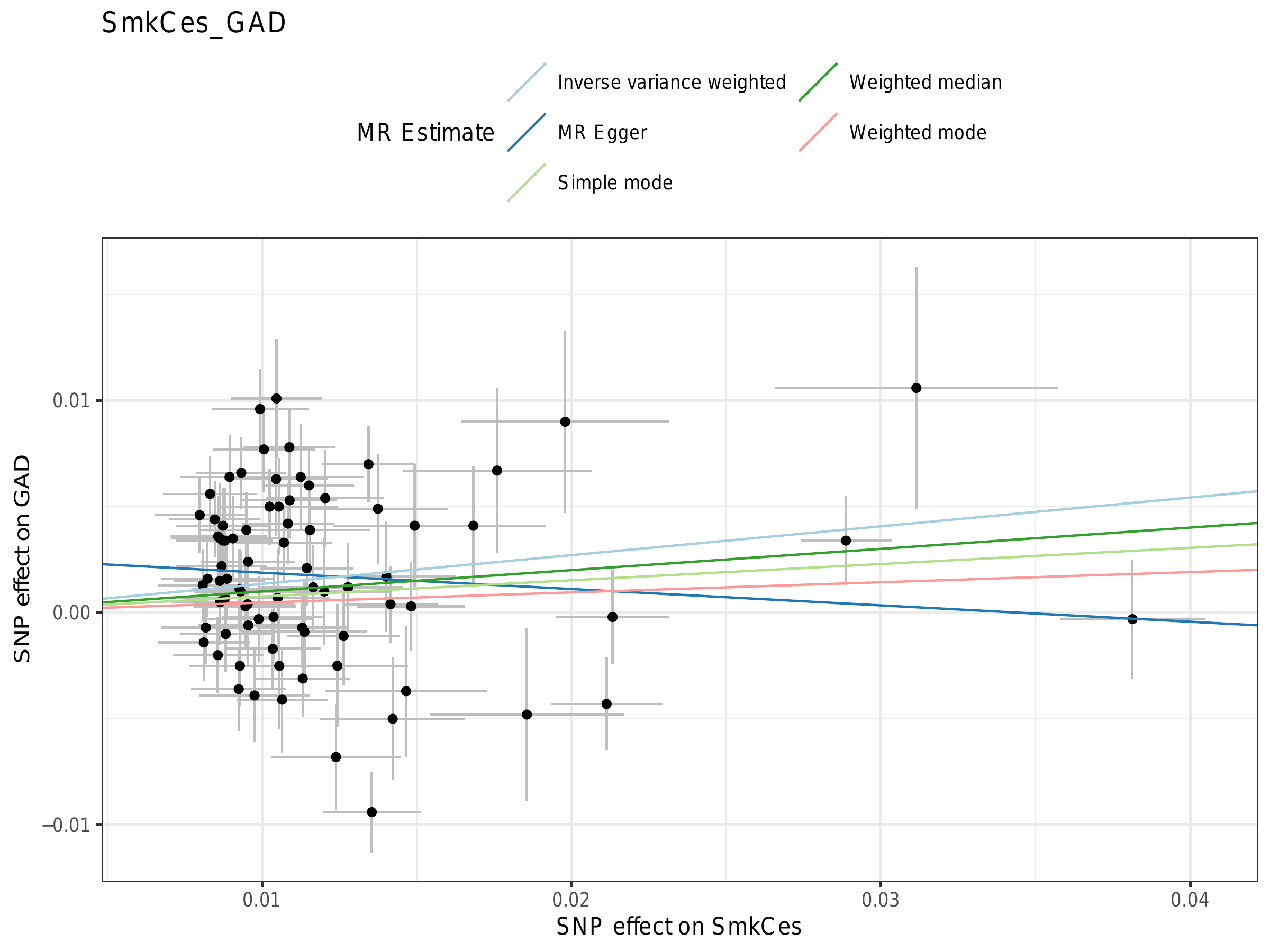

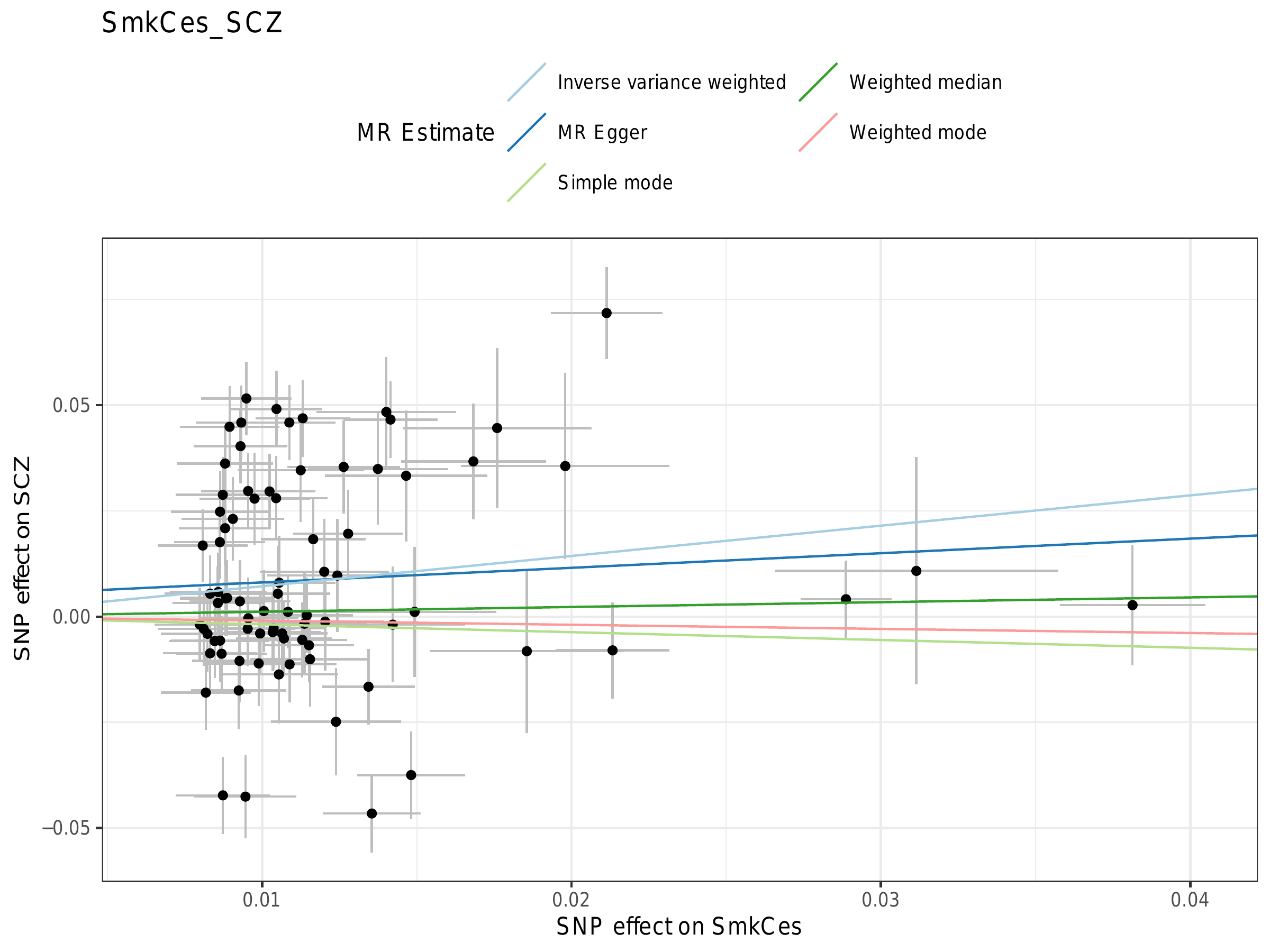

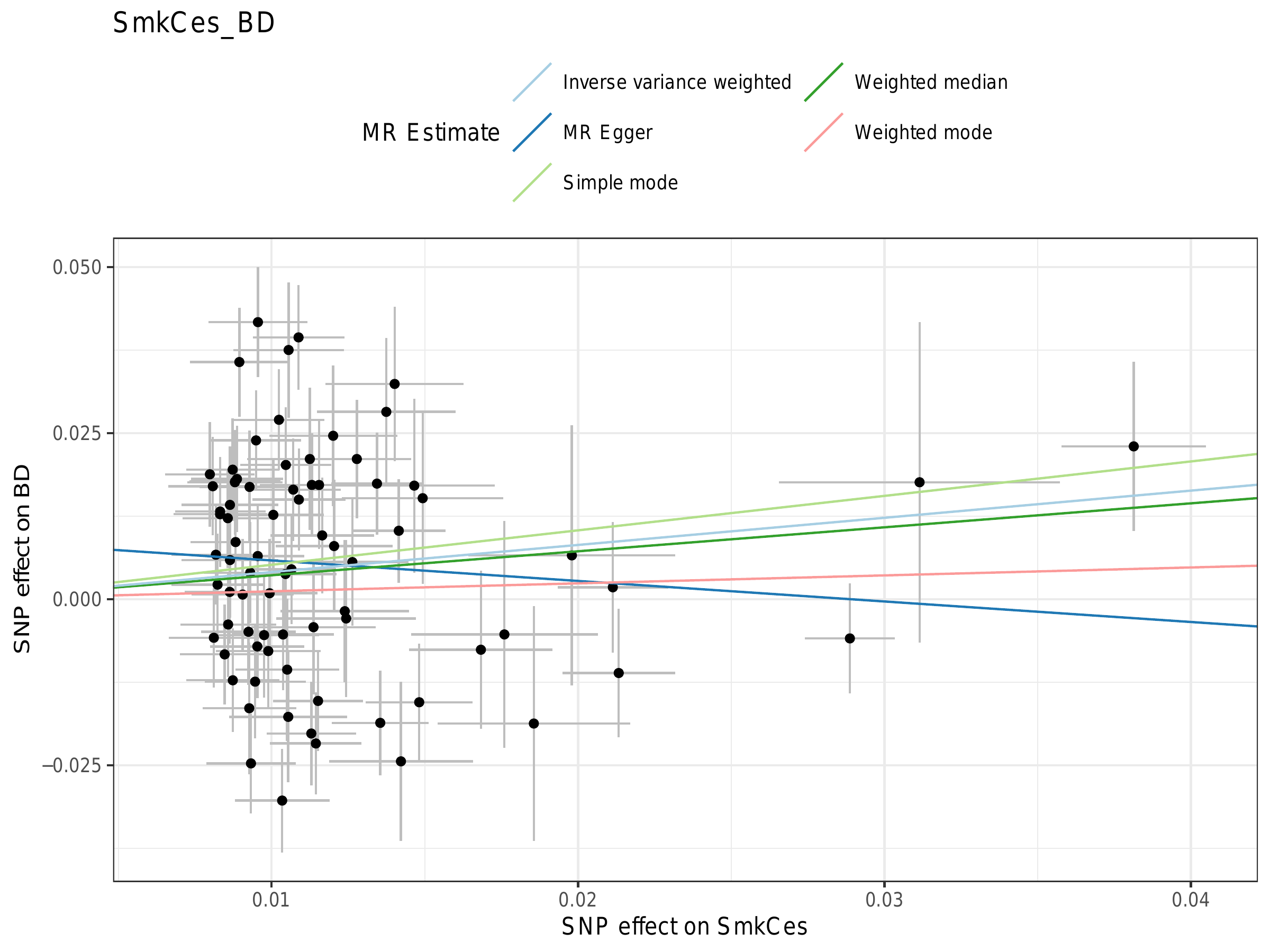

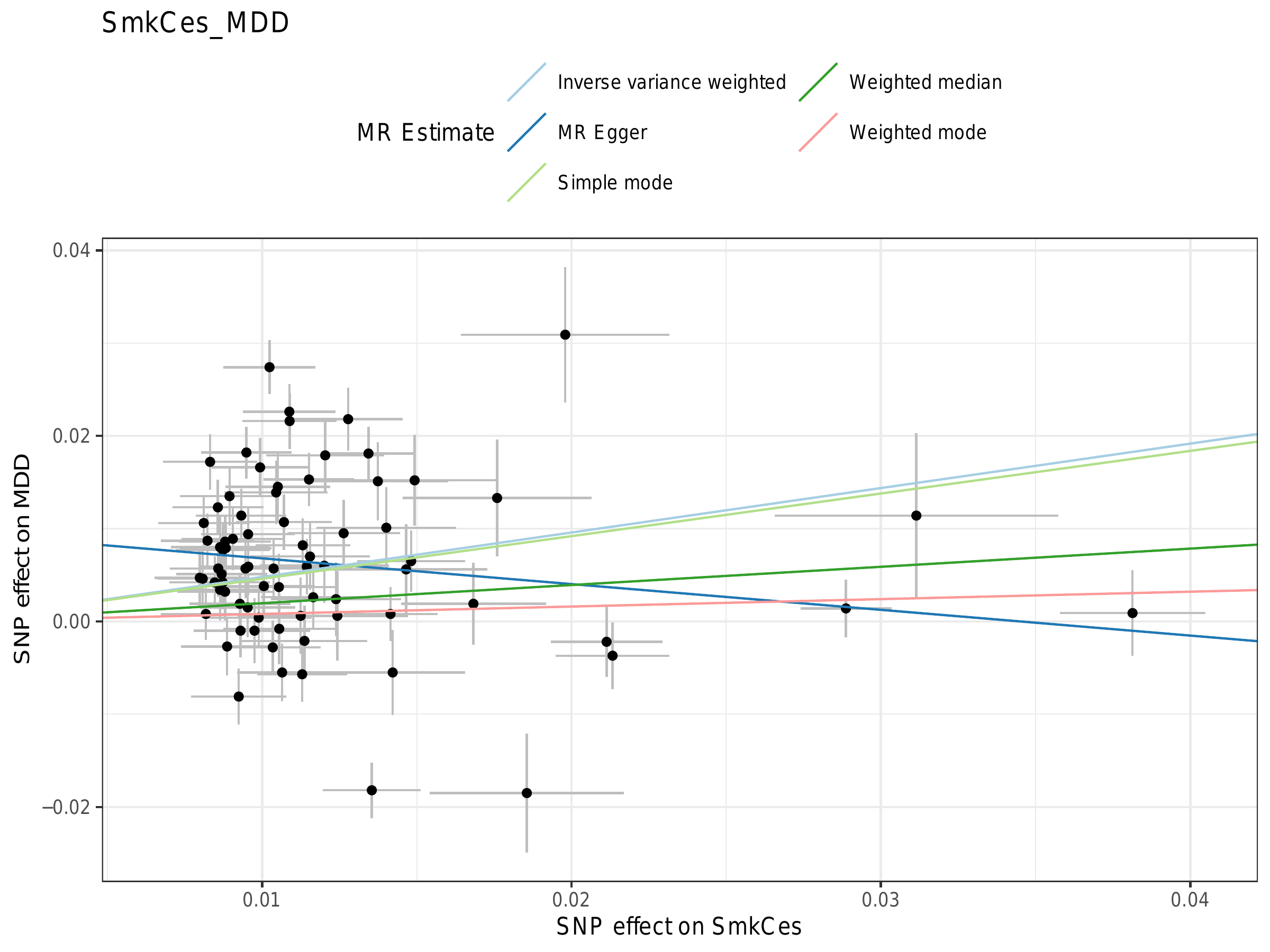

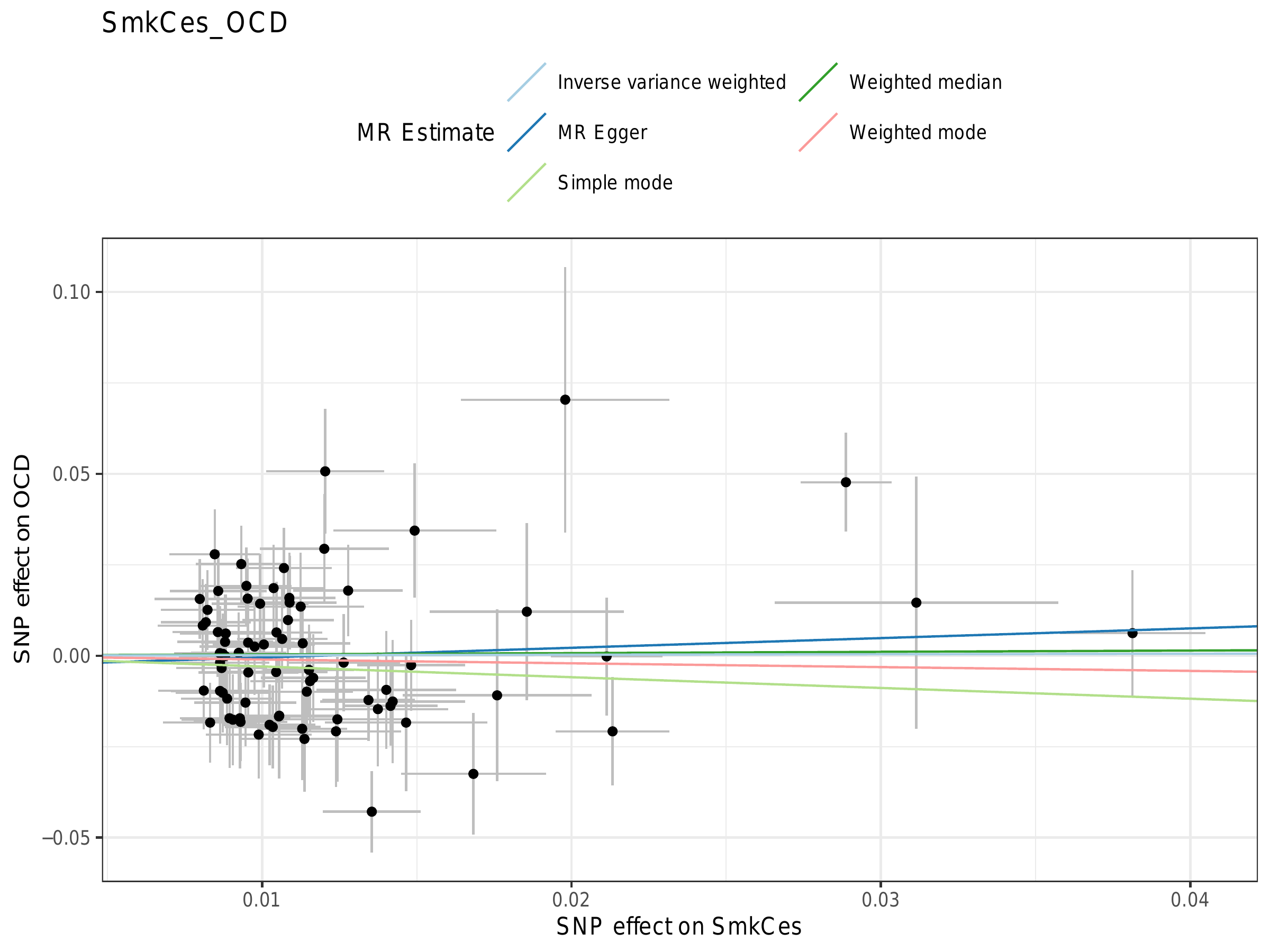

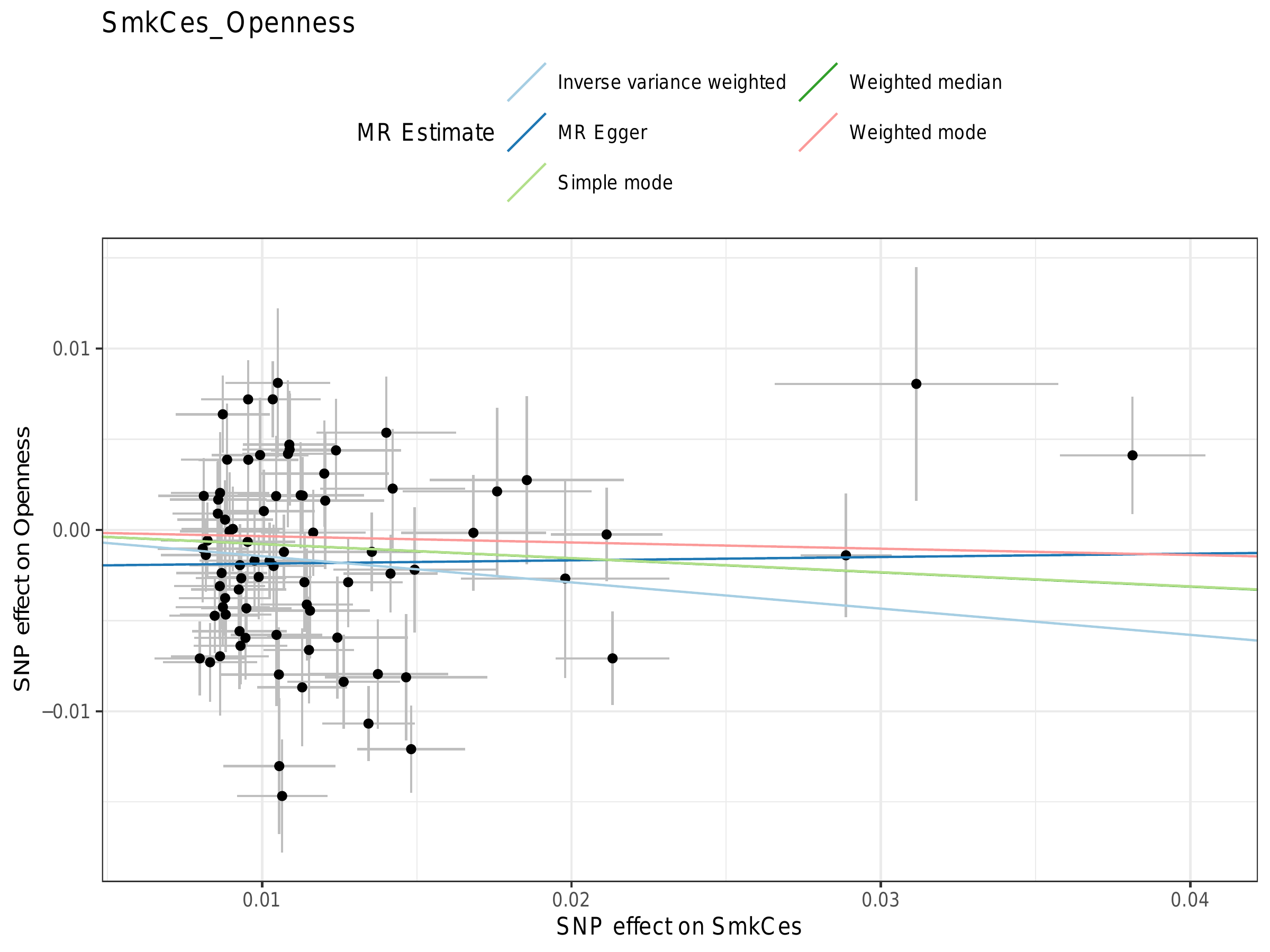

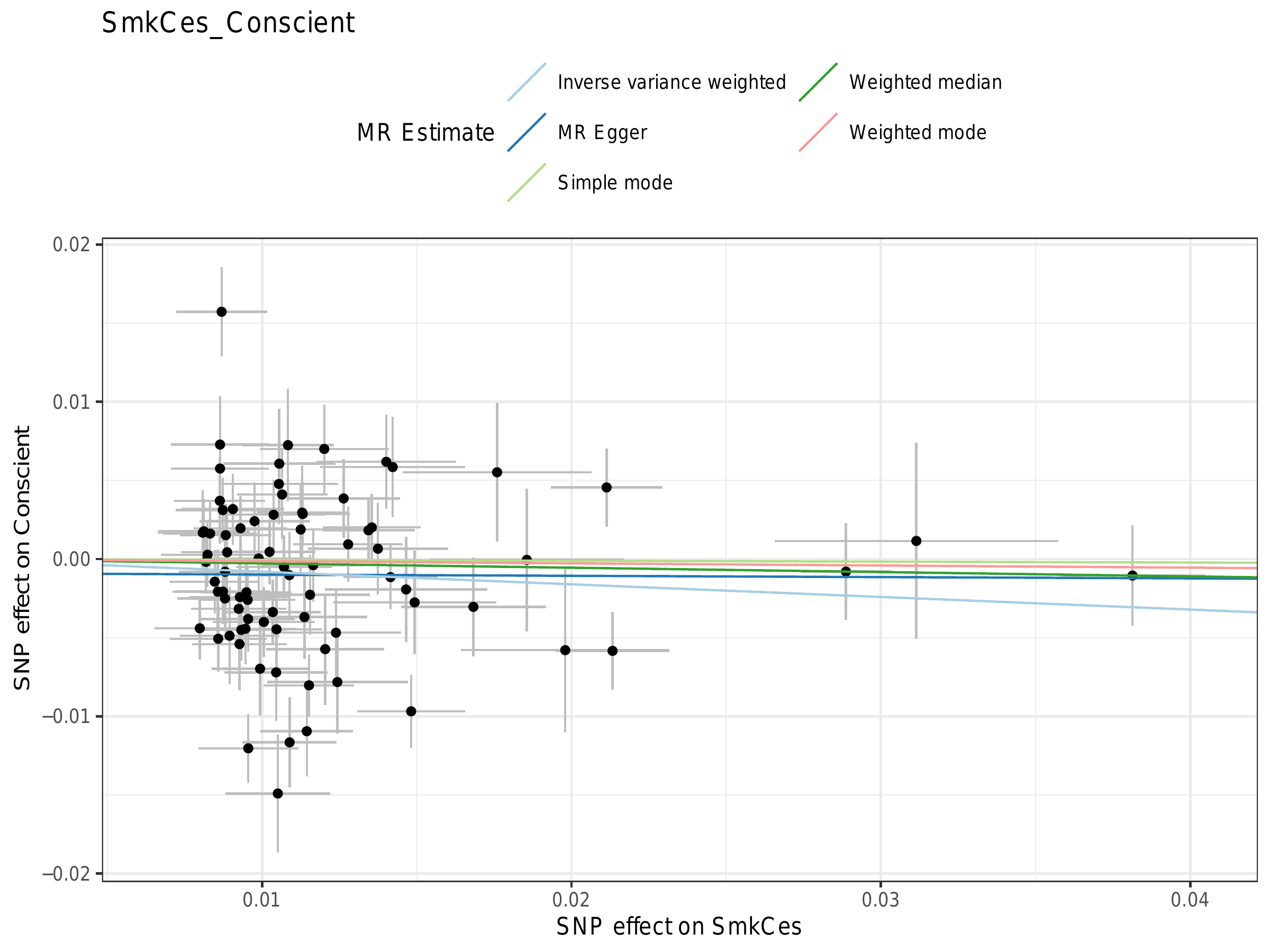

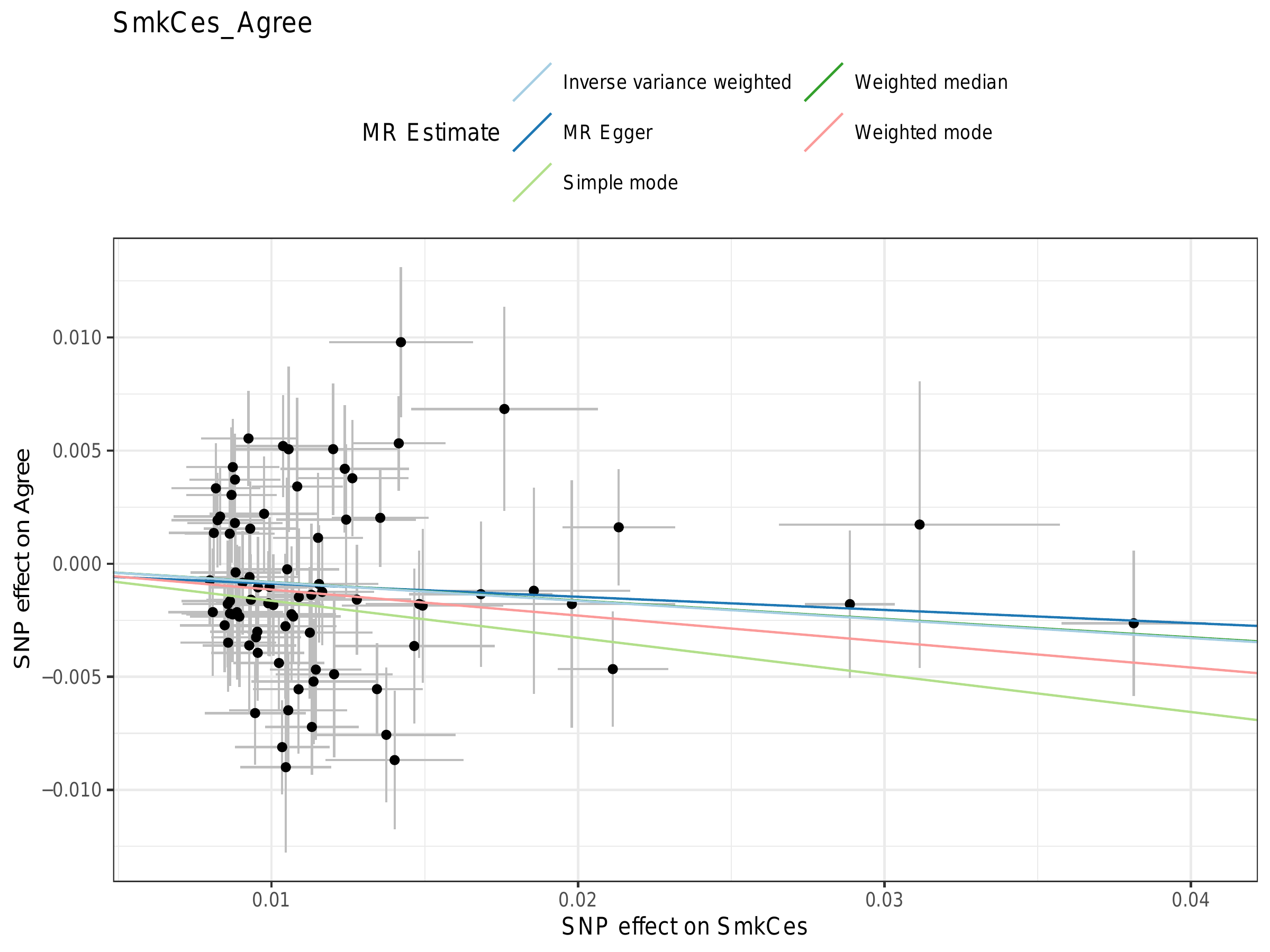

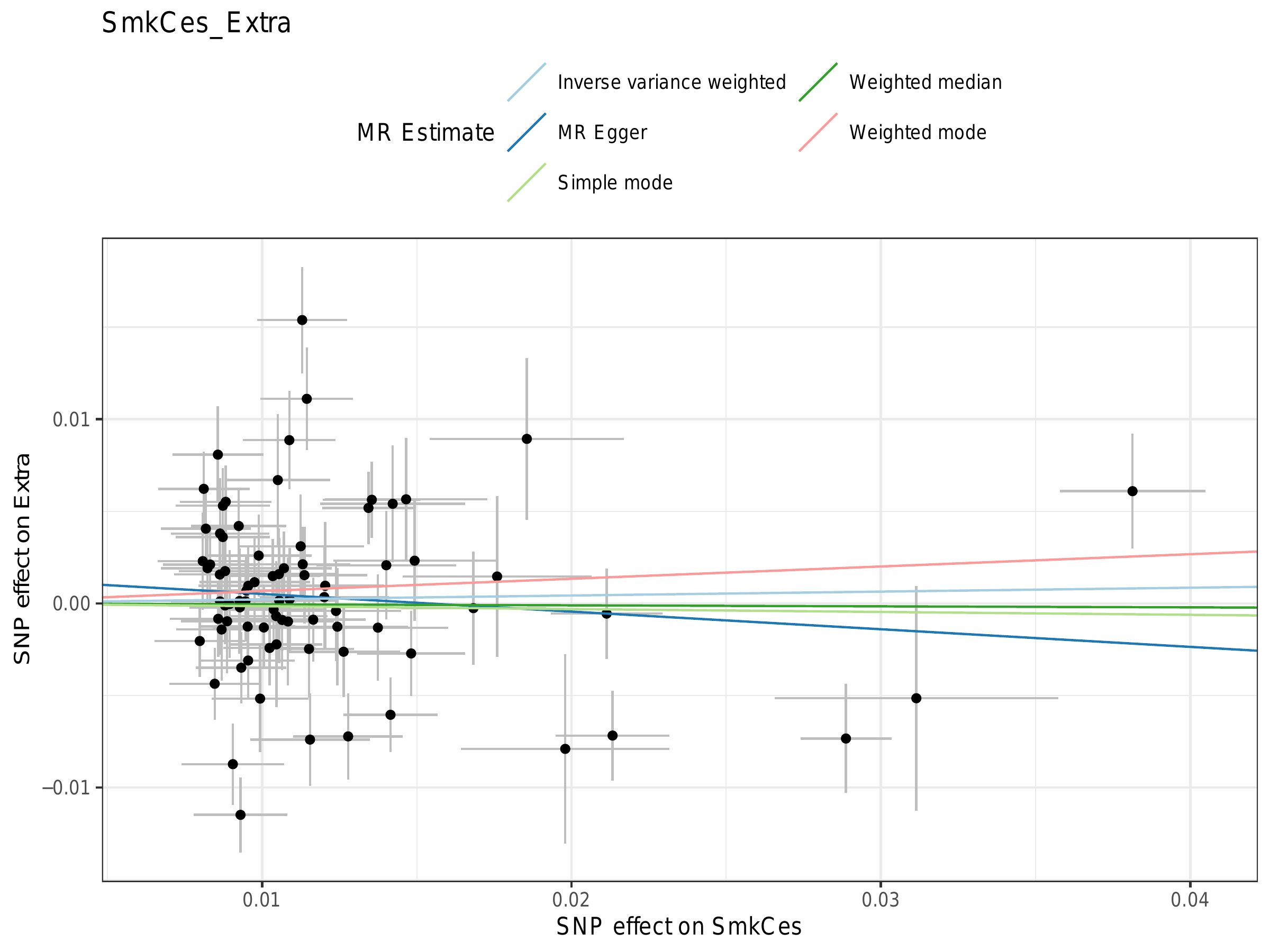

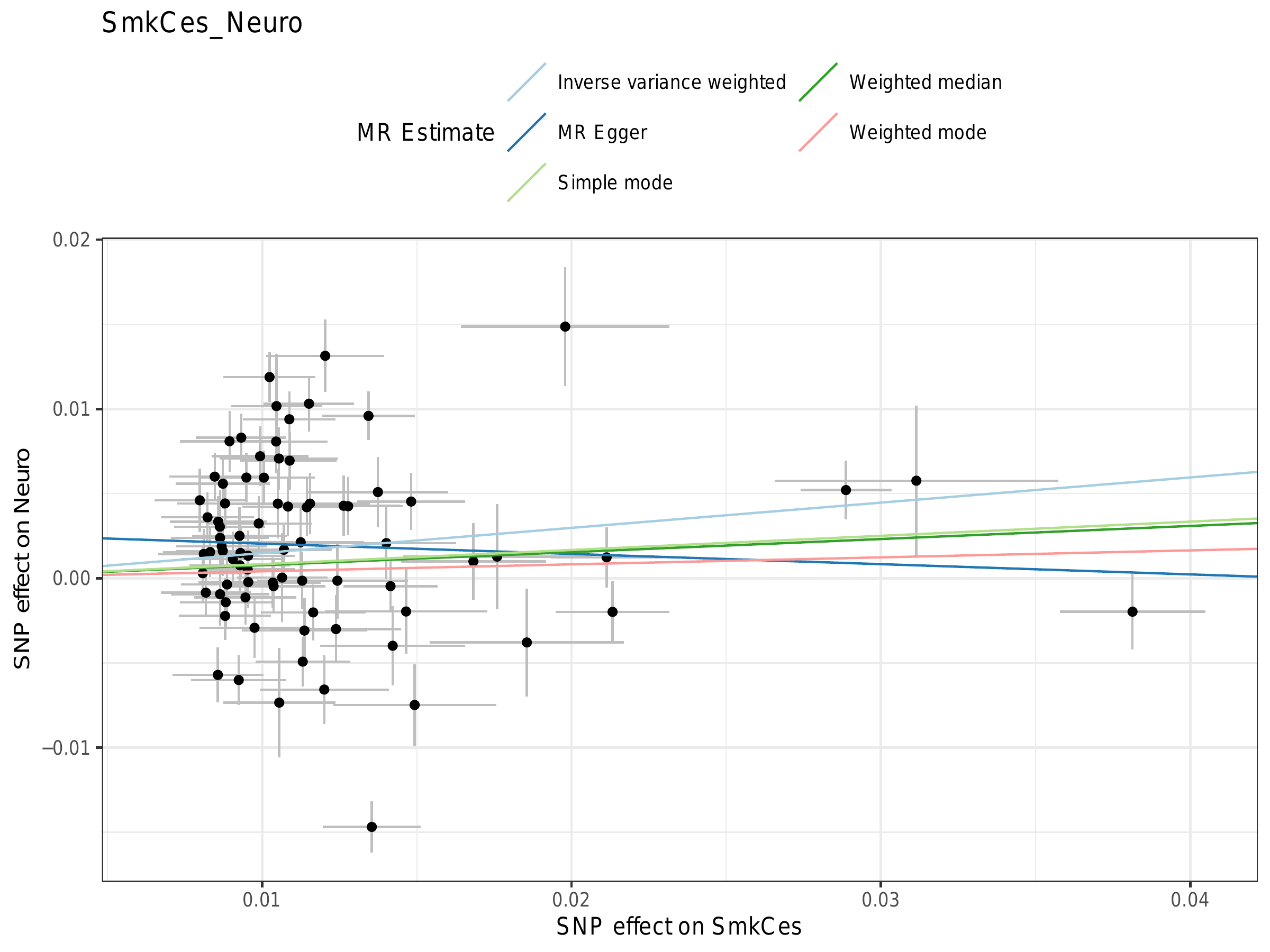

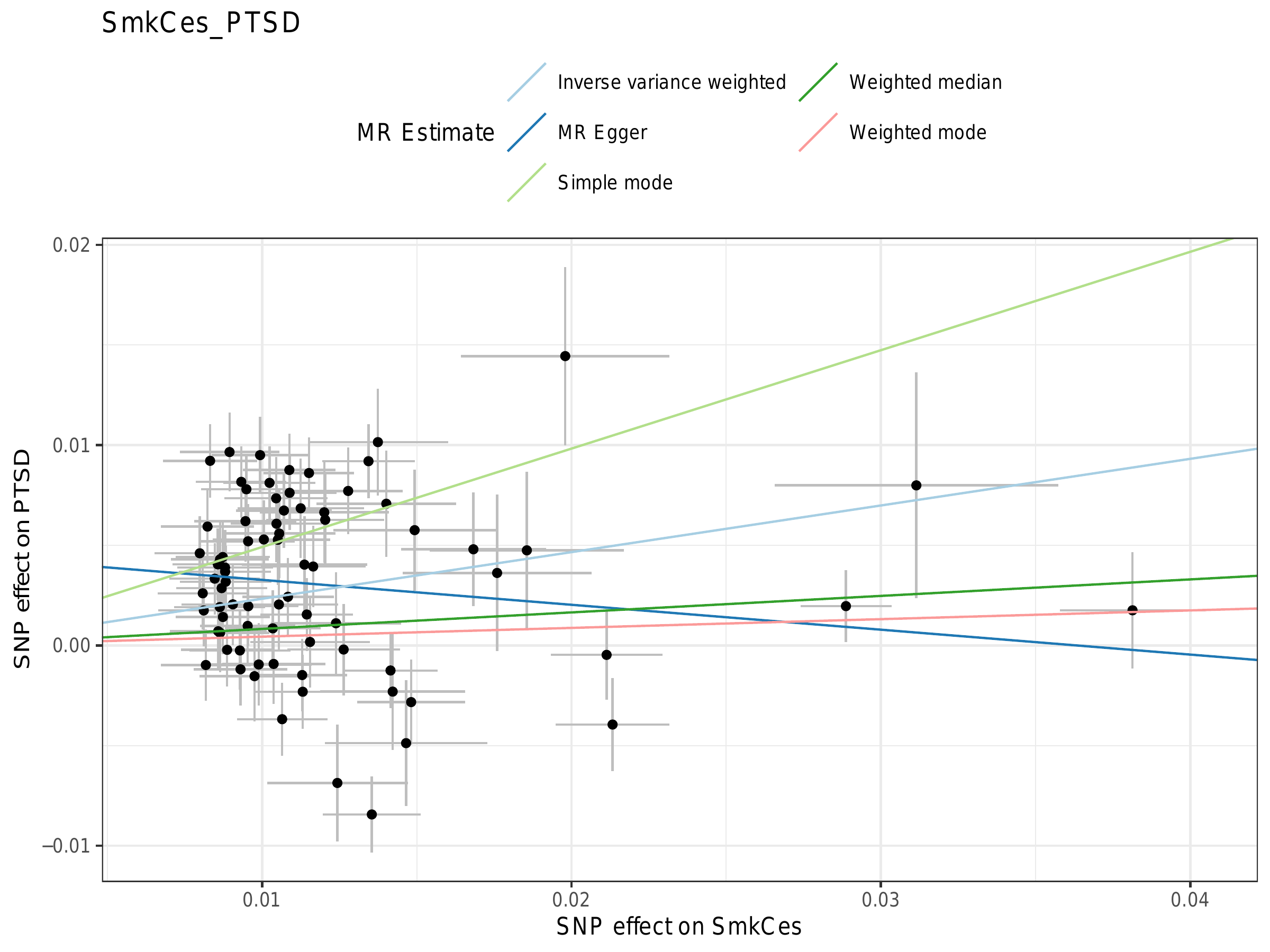

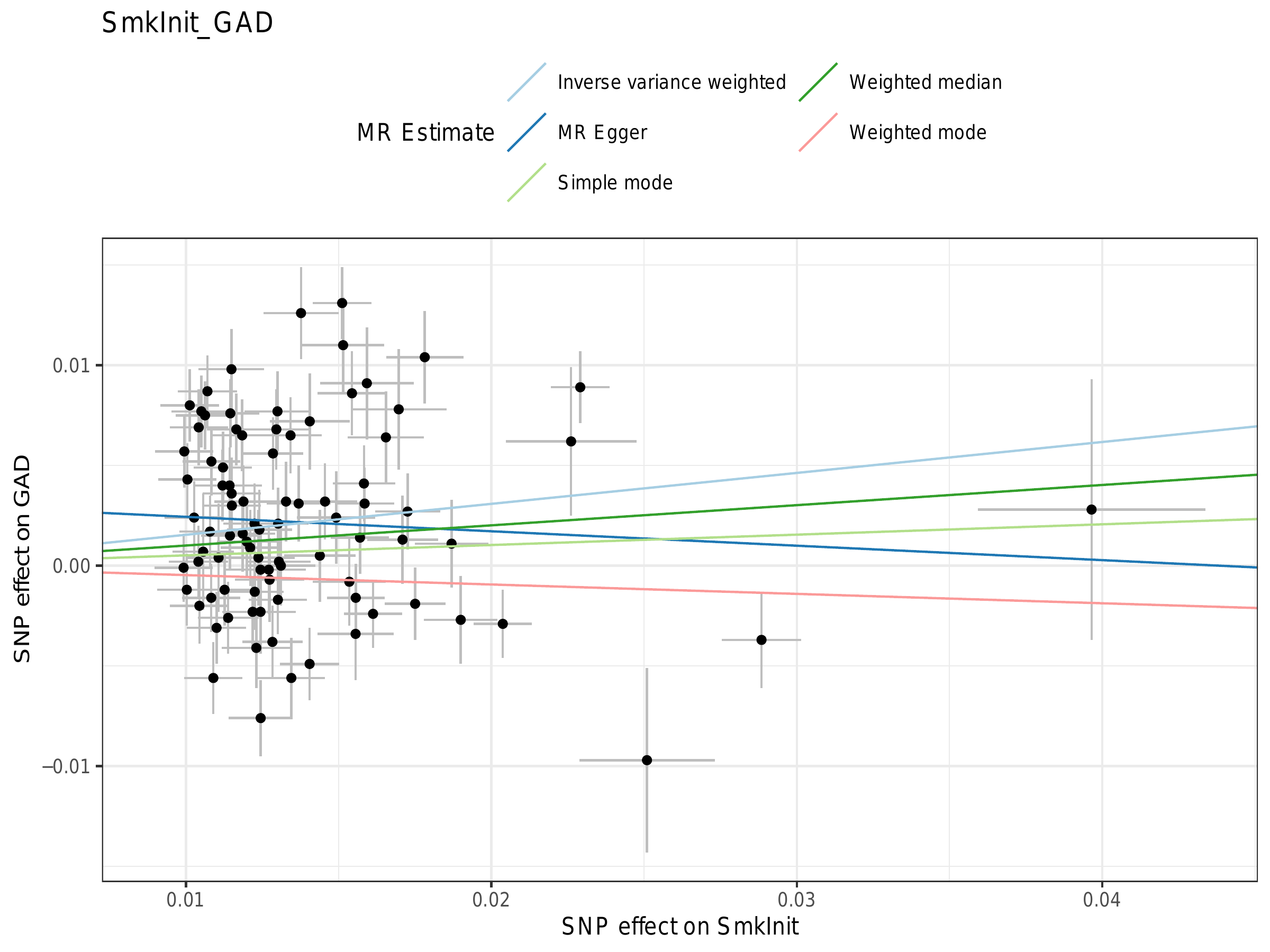

P3. Leave-one-out plots

**Reverse analyses**

P4. Funnel plots

P5. Scatter plots

P6. Leave-one-out plots
